# Deep RNA Sequencing of Intensive Care Unit Patients with COVID-19

**DOI:** 10.1101/2021.01.11.21249276

**Authors:** Sean F. Monaghan, Alger M. Fredericks, Maximilian S. Jentzsch, William G. Cioffi, Maya Cohen, William G. Fairbrother, Shivam J. Gandhi, Elizabeth O. Harrington, Gerard J. Nau, Jonathan S. Reichner, Corey E. Ventetuolo, Mitchell M. Levy, Alfred Ayala

**Affiliations:** Alpert Medical School of Brown University /Rhode Island Hospital, Department of Surgery, Division of Surgical Research; Alpert Medical School of Brown University /Rhode Island Hospital, Department of Medicine, Division of Pulmonary, Critical Care, and Sleep Medicine; Brown University; Alpert Medical School of Brown University /Rhode Island Hospital, Department of Medicine, Division of Infectious Disease

**Keywords:** COVID-19, RNA sequencing, RNA processing, pathogen identification, gene expression

## Abstract

**Purpose:** COVID-19 has impacted millions of patients across the world. Molecular testing occurring now identifies the presence of the virus at the sampling site: nasopharynx, nares, or oral cavity. RNA sequencing has the potential to establish both the presence of the virus and define the host’s response in COVID-19.

**Methods:** Single center, prospective study of patients with COVID-19 admitted to the intensive care unit where deep RNA sequencing (>100 million reads) of peripheral blood with computational biology analysis was done. All patients had positive SARS-CoV-2 PCR. Clinical data was prospectively collected.

**Results:** We enrolled fifteen patients at a single hospital. Patients were critically ill with a mortality of 47% and 67% were on a ventilator. All the patients had the SARS-CoV-2 RNA identified in the blood in addition to RNA from other viruses, bacteria, and archaea. The expression of many immune modulating genes, including PD-L1 and PD-L2, were significantly different in patients who died from COVID-19. Some proteins were influenced by alternative transcription and splicing events, as seen in HLA-C, HLA-E, NRP1 and NRP2. Entropy calculated from alternative RNA splicing and transcription start/end predicted mortality in these patients.

**Conclusions:** Current upper respiratory tract testing for COVID-19 only determines if the virus is present. Deep RNA sequencing with appropriate computational biology may provide important prognostic information and point to therapeutic foci to be precisely targeted in future studies.

**Take Home Message:** Deep RNA sequencing provides a novel diagnostic tool for critically ill patients. Among ICU patients with COVID-19, RNA sequencings can identify gene expression, pathogens (including SARS-CoV-2), and can predict mortality.

**Tweet:** Deep RNA sequencing is a novel technology that can assist in the care of critically ill COVID-19 patients & can be applied to other disease

## Introduction

Severe acute respiratory syndrome coronavirus 2 (SARS-CoV-2) causing coronavirus disease 2019 (COVID-19) has led to millions of cases worldwide.[1] Current testing is by polymerase chain reaction to detect viral RNA in the nares[2], but provides no insight into the host response. Patients with COVID-19 that require intensive care unit (ICU) care are sick and difficult to manage, thus, there is a need for other diagnostic tests during the hospital stay to assist the clinicians.

Deep RNA sequencing refers to a process of sequencing where (at least) 100 million reads of sequence are generated per sample. Deep sequencing allows for the study of low abundance RNA and biologic processes beyond gene expression. Typically, RNA sequencing data is aligned to the genome of interest, such as aligning to human genes when the sample comes from a human. Reads that do not align to the genome of interest are usually discarded. When the RNA sequencing is performed with this large number of reads, it could be used to identify the presence of specific pathogens in the blood by aligning the reads that would have been discarded to other genomes of interest. In COVID-19, sequencing reads of SARS-CoV-2 may provide insight into the biology of the virus during active illness. In addition, secondary infections could be identified, potentially allowing for better, pathogen-directed antibiotic treatment.

The host response to the virus is responsible for some of the morbidity and mortality observed.[3] Acute respiratory distress syndrome (ARDS) is the most common complication encountered with COVID-19.[3] Our laboratory has shown that there are significant changes in alternative RNA splicing and transcription start and end in ARDS as assessed by deep RNA sequencing.[4] These changes are thought to be due to the physiology of ARDS, e.g., hypoxia and acidosis, which are known to influence splicing. Whether this occurs in patients infected by COVID-19 is not known.

While RNA sequencing can be used to measure immune modulating gene expression, an alternative approach is the evaluation of global entropy, or disorder in the processing of RNA.[5] In this study, we propose that this entropy metric combined with Principal Component Analysis (PCA) can be leveraged to distinguish COVID-19 patients that develop life-threatening illness from those likely to recover.

Here we examine deep RNA sequencing data from patients in the ICU with COVID-19 to characterize both pathogens and host responses. We evaluate the sequences for the presence of the SARS-CoV-2 virus and other potential infectious agents. The host response to COVID-19 is also characterized. The long-term goal is to combine these measurements to better assist clinical decision-making.

## Methods

### Study design, Population and Setting

The study enrolled ICU participants at a single tertiary care hospital evidence of SARS-CoV-2 infection based on positive PCR from the nasopharynx documented during admission. All participants, or their appropriate surrogate, provided informed consent as approved by the Institutional Review Board (Approval #: 411616). Blood samples were collected on day 0 of ICU admission. Clinical data including COVID specific therapies was collected prospectively from the electronic medical record and participants were followed until hospital discharge or death. Ordinal scale was collected as previously described;[6] along with sepsis and associated SOFA score[7] and the diagnosis of ARDS.[8]

### RNA extraction and sequencing

Whole blood was collected in PAXgene tubes (Qiagen, Germantown, MD) and sent to Genewiz (South Plainfield, NJ) for RNA extraction, ribosomal RNA depletion and sequencing. Sequencing was done on Illumina HiSeq machines to provide 150 base pair, paired-end reads. Libraries were prepared to have three samples per lane. Each lane provided 350 million reads ensuring each sample had >100 million reads. Raw data was returned on password protected external hard drives to ensure the security of the genomic data.

### Computational Biology and Statistical Analysis

All computational analysis was done blinded to the clinical data. The data was assessed for quality control using FastQC.[9] RNA sequencing data was aligned to the human genome utilizing the STAR aligner.[10] Reads that aligned to the human genome were separated and are now referred to as ‘mapped’ reads. Reads that did not align to the human genome, which are typically discarded during standard RNA sequencing analysis, were kept and identified as ‘unmapped’ reads. The unmapped reads then aligned to the SARS-CoV-2 genome (NC_045512) and counted per sample using Magic-BLAST.[11] In addition, a coverage map of the SARS-CoV-2 genome was generated using all the subjects to identify the gene expression patterns of the virus in critically ill COVID-19 patients. The unmapped reads were further analyzed with Kraken2[12] using the PlusPFP index[13] to identify other bacterial, fungal, archaeal and viral pathogens.

Reads that aligned to the human genome, the mapped reads, also underwent analysis for gene expression, alternative RNA splicing, and alternative transcription start/end via Whippet.[5] When comparisons were made between groups (died vs. survived) differential gene expression was set with thresholds of both p<0.05 and +/- 1.5 log_2_ fold change. Alternative splicing was defined as core exon, alternative acceptor splice site, alternative donor splice site, retained intron, alternative first exon and alternative last exon. Alternative transcription start/end events were defined as tandem transcription start site and tandem alternative polyadenylation site. Alternative RNA splicing and alternative transcription start/end events were also compared between groups.[5] Significance was set at great than 2 log_2_ fold change as previously described.[4] Genes identified from the analysis of mapped reads were then evaluated by GO enrichment analysis (PANTHER Overrepresentation released 20200728).[14]

Whippet was also used to generate an entropy value for every identified alternative splicing and transcription event of each gene. These entropy values are created without the need for groups used in the gene expression analysis. In order to visualize this data a principal component analysis (PCA) was conducted to reduce the dimensionality of the dataset and to obtain an unsupervised overview of trends in entropy values among the samples. Raw entropy values from all samples were concatenated into one matrix and missing values were replaced with column means. Mortality was then overlaid onto the PCA plot to assess the ability of these raw entropy values to predict this outcome in this sample set. This analysis was done in R (version 3.6.3).

## Results

### Study Population, Participant Characteristics, and RNA sequencing

Fifteen participants were enrolled and had blood samples drawn on the first day of their ICU stay. Clinical and demographic data is reported in Table 1. The majority of participants were male (73%) and there were a diverse distribution in terms of race (60% not white) and ethnicity (60% Hispanic). The most common co-morbidity was hypertension and the median BMI was almost 30. Forty percent of participants had ARDS at the time the samples was drawn and the patients were distributed across the top of the ordinal scale[6] with a score of 5 as the most common in 53% of the patients. Most participants required a ventilator (67%) and 20% progressed to extracorporeal membrane oxygenation (ECMO); 27% required renal replacement. The median length of hospital stay was 22 days with a mortality rate of 47%.

**Table 1.** Demographics

| Table 1 - Demographics | All Patients<br>(n=15) | Alive (n=8) | Died (n=7) |
| --- | --- | --- | --- |
| Median age (IQR) | 66.2 (41.6-72.1) | 53.1 (34.5-72.8) | 66.2 (50.0-73.3) |
| Sex |  |  |  |
| Female - no. (%) | 4 (27) | 2 (25) | 2 (29) |
| Male - no. (%) | 11 (73) | 6 (75) | 5 (71) |
| Race |  |  |  |
| White - no. (%) | 6 (40) | 2 (25) | 4 (57) |
| Black or African American - no. (%) | 2 (13) | 1 (13) | 1 (14) |
| Other Race - no. (%) | 7 (47) | 5 (62) | 2 (29) |
| Ethnicity |  |  |  |
| Hispanic - no. (%) | 9 (60) | 4 (50) | 5 (71) |
| Not Hispanic - no. (%) | 6 (40) | 4 (50) | 2 (29) |
| Chronic Underlying Conditions - no. (%) |  |  |  |
| Hypertension | 9 (60) | 4 (50) | 5 (71) |
| Cardiovascular Disease | 2 (13) | 0 (0) | 2 (29) |
| Congestive Heart Failure | 2 (13) | 1 (13) | 1 (14) |
| Diabetes | 1 (7) | 0 (0) | 1 (14) |
| COPD | 1 (7) | 0 (0) | 1 (14) |
| Renal Failure | 1 (7) | 0 (0) | 1 (14) |
| Malignancy | 1 (7) | 1 (13) | 0 (0) |
| Liver Disease | 0 (0) | 0 (0) | 0 (0) |
| Median BMI (IQR) | 29. 6 (27.3-31.9) | 27.7 (24.8-31.9) | 30.3 (29.5-35) |
| At the time of sample |  |  |  |
| ARDS - no. (%) | 6 (40) | 3 (38) | 3 (43) |
| Vassopressor Support - no. (%) | 2 (13) | 1 (13) | 1 (14) |
| Median SOFA Score (IQR) | 6 (3-8) | 7.5 (4-8) | 3 (2-6) |
| Median APACHE II Score (IQR) | 18 (13.5-23) | 20.5 (17.3-24.5) | 12 (9-22) |
| Ordinal Scale 4 - no. (%) | 3 (20) | 3 (38) | 0(0) |
| Ordinal Scale 5 - no. (%) | 8 (53) | 1 (13) | 7 (100) |
| Ordinal Scale 6 - no. (%) | 1 (7) | 1 (13) | 0(0) |
| Ordinal Scale 7 - no. (%) | 3 (20) | 3 (38) | 0(0) |
| Hospital Course |  |  |  |
| Median Ventilator Days (IQR) | 6 (0-30) | 7.5 (0-45.75) | 2 (0-27) |
| Median ICU Length of Stay (IQR) | 15 (6.5-35.5) | 9 (3.5-36.25) | 18 (8-44) |
| Median Hospital Length of Stay (IQR) | 22 (13-49.5) | 25 (8.75-71.25) | 22 (15-45) |
| ECMO - no. (%) | 3 (20) | 2 (25) | 1 (14) |
| Acute Renal Replacement - no. (%) | 4 (27) | 2 (25) | 2 (29) |
| Thrombotic Event - no. (%) | 6 (40) | 3 (38) | 3 (43) |
| Death - no. (%) | 7 (47) |  |  |
| Discharge from hospital - no. (%) | 8 (53) |  |  |
| Median D Dimer (IQR) | 2980 (974-4312) | 2657 (885-4433) | 3370 (923-7180) |
| TEG hypercoagulable- no. (%) | 6 (40) | 3 (38) | 3 (43) |
| Remdesivir- no. (%) | 8 (53) | 4 (50) | 4 (57) |
| Plasma - no. (%) | 2 (13) | 1 (13) | 1 (14) |

All samples had sufficient RNA and RNA integrity numbers (RIN)[15] were adequate. The median of sequencing was 125,687,784 reads (95% CI 122,164,763 to 135,800,242) and greater than 90% of those reads were more than thirty bases. After using FastQC[9], all samples had mean quality scores over 30. The reads mapped to the human genome 62-66% of the time (Table E1).

### Identification of SARS-CoV-2 and other pathogens

Among the fifteen participant samples all participants had SARS-CoV-2 RNA detected. There were a total of 676 reads that align to the SARS-CoV-2 genome with each patient having between 18 and 98 reads. (Figure 1a) The majority of the reads corresponded to the RNA dependent RNA polymerase and N protein genes (Figure 1b). RNA from other pathogens including bacteria, viruses and archaea were identified in the blood of all patients (Table 2). Two participants had fungal RNA identified. Despite alignment to a robust database of organisms, each participant still had hundreds of thousands of unclassified reads. (Table 2) The taxonomy classification of “other sequences” (28384) align to elements of cellular organisms (bacterial, archaea, plant), but do not have enough specificity to identify a single species are listed (Table 2). The top ten bacterial reads by count at the species level for each patient is listed in Table E2. The top bacterial sequences from all patients were from either *Acinetobacter baumannii* or *Chryseobacterium gallinarum*. In patients who had the most counts of *C. gallinarum, A. baumannii* had significantly reduced counts compared to the counts in other patients (148.1 vs. 50905.3, p<0.05). Although sequences corresponding to *A. baumannii or C. gallinarum* were found in all patients, none of the patients had positive blood cultures drawn around the time of these samples. No counts of bacteria, virus, archaea, (Table 2) or specific bacteria (Table E2) correlated with mortality.

**Table 2.** Counts per patient from Kraken2

|  | Unclassified | Bacteria | Archaea | Other Sequences | Virus |
| --- | --- | --- | --- | --- | --- |
| Patient 1 | 166,039 | 252,424 | 160 | 388,924 | 317 |
| Patient 2 | 219,873 | 213,967 | 167 | 325,150 | 398 |
| Patient 3 | 216,633 | 258,383 | 286 | 463,595 | 621 |
| Patient 4 | 272,019 | 122,270 | 230 | 637,551 | 374 |
| Patient 5 | 266,733 | 119,180 | 224 | 552,125 | 363 |
| Patient 6 | 217,155 | 179,383 | 292 | 506,456 | 564 |
| Patient 7 | 144,690 | 111,078 | 153 | 321,752 | 302 |
| Patient 8 | 262,426 | 194,038 | 310 | 557,825 | 586 |
| Patient 9 | 294,609 | 325,558 | 1,055 | 475,941 | 768 |
| Patient 10 | 273,603 | 272,536 | 211 | 373,893 | 403 |
| Patient 11 | 222,280 | 175,230 | 169 | 333,625 | 368 |
| Patient 12 | 284,819 | 113,546 | 223 | 615,463 | 271 |
| Patient 13 | 308,700 | 103,060 | 238 | 469,993 | 298 |
| Patient 14 | 235,961 | 109,451 | 183 | 485,807 | 449 |
| Patient 15 | 179,668 | 130,323 | 188 | 419,667 | 353 |

**Figure 1:**
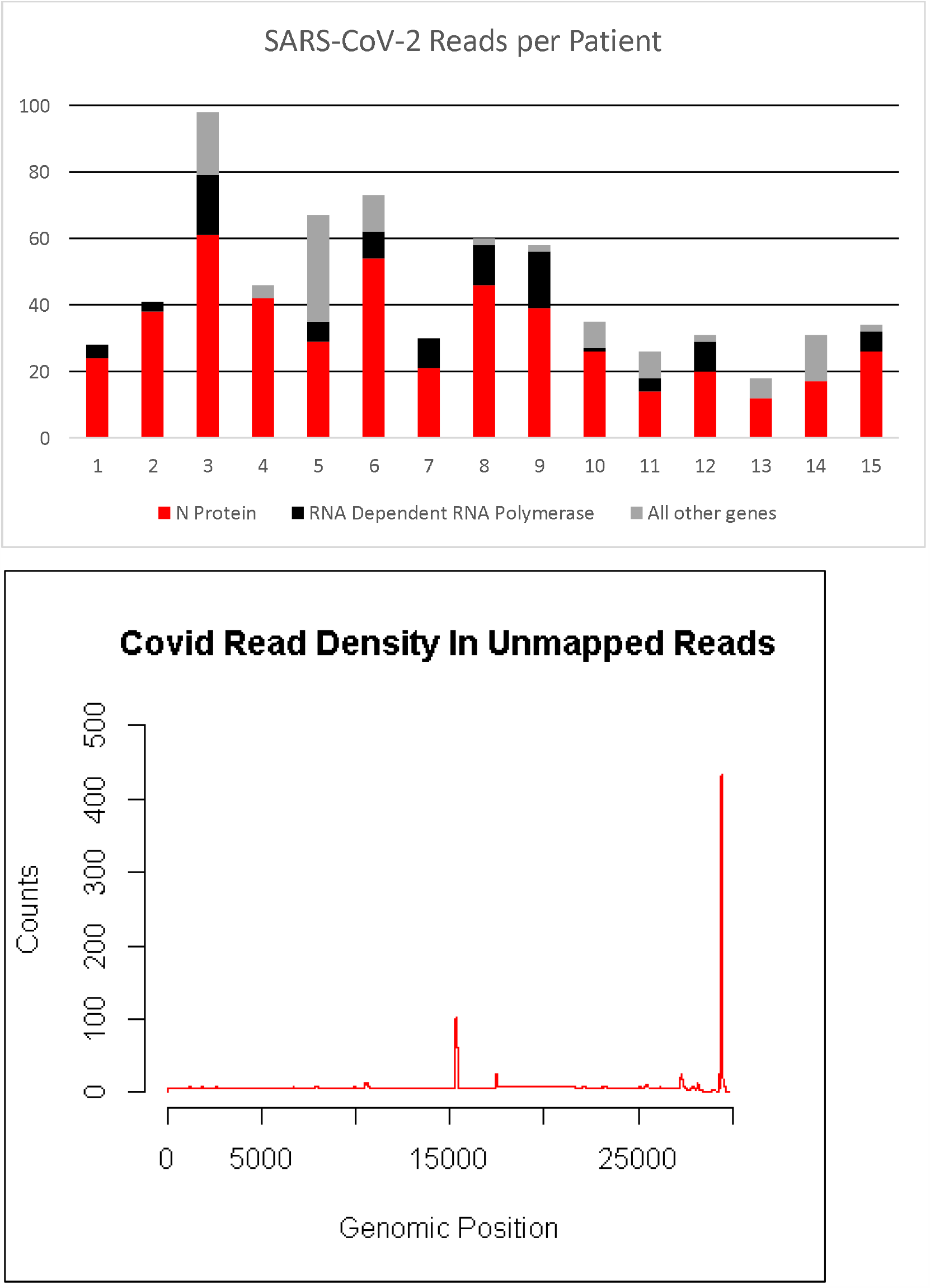
Top panel is the number of reads aligning to the SARS-CoV-2 genome from each patient. Most reads aligned to loci encoding the N protein (red bar) or the RNA dependent RNA polymerase (black bar). Bottom panel is the location where the cumulative reads from all the patients align to the SARS-CoV-2 genome. Genes encoding the RNA dependent RNA polymerase and the N protein are at positions ∼15,000 and ∼29,000, respectively.

### Genomic differences between participants who lived and those who died

Among participants who died there were 86 genes that increased in expression and 207 that decreased in expression (top results in Table 3, full list in Table E3, Figure E1). There were 88 significant alternative splicing events occurring in 84 unique genes (Top results Table 3, full list Table E4) and 2093 alternative transcription events occurring in 1769 unique genes (Top results Table 3, full list Table E5). ABCA13 was the only gene that had significant expression and alternative splicing events. Twenty-seven genes had significant expression and alternative transcription start/end differences. (Table 3) Eighteen genes had significantly different alternative splicing and alternative transcription start/end. (Table 3)

**Table 3.**
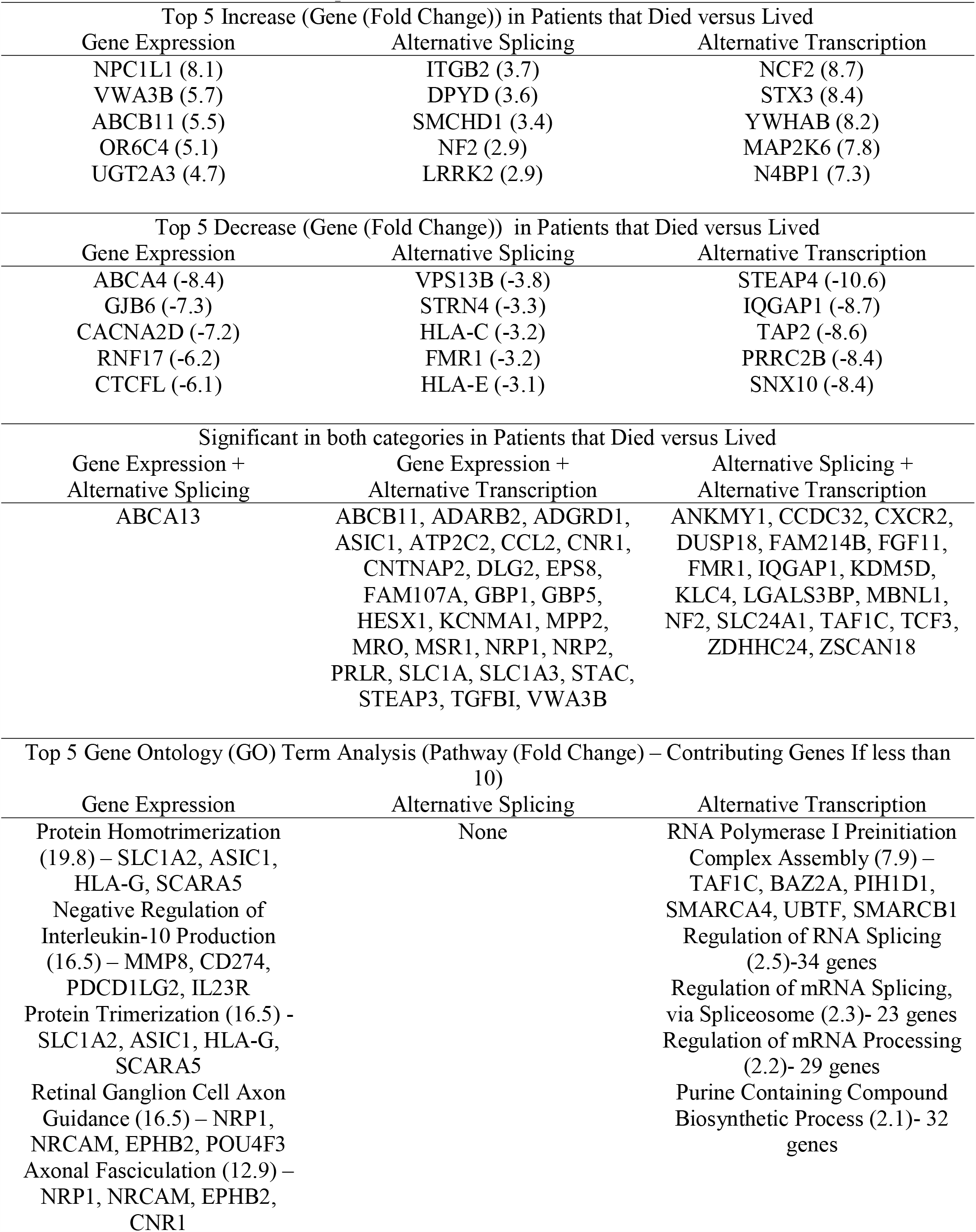
Gene difference between patients that died versus lived

The genes that were significant between groups then underwent GO term analysis to assess significant enrichment for a biological process. The top GO terms for gene expression and alternative transcription are listed in Table 3 (full list Tables E6 and E7). There were no significant GO terms for the genes impacted by alternative splicing.

### RNA entropy as a diagnostic tool

From the over 100 million RNA sequencing reads for each participant, computational analysis via Whippet assigns an entropy value for over 380,000 RNA splicing events and alternative transcription start/end events. Principal component analysis was then applied to these >380,000 entropy scores for each of the 15 participants and the first two principal components were plotted against each other (Figure 2). The sample points were then labeled based on their survival status. Survival status was not part of the principal component analysis itself. Participants whose PC2 value was above 0.00 had a mortality rate of 75% (6/8), up from the total group mortality of 46% (7/15) and significantly more than the 14% for those who land below that line (1/7, p=0.04).

**Figure 2:**
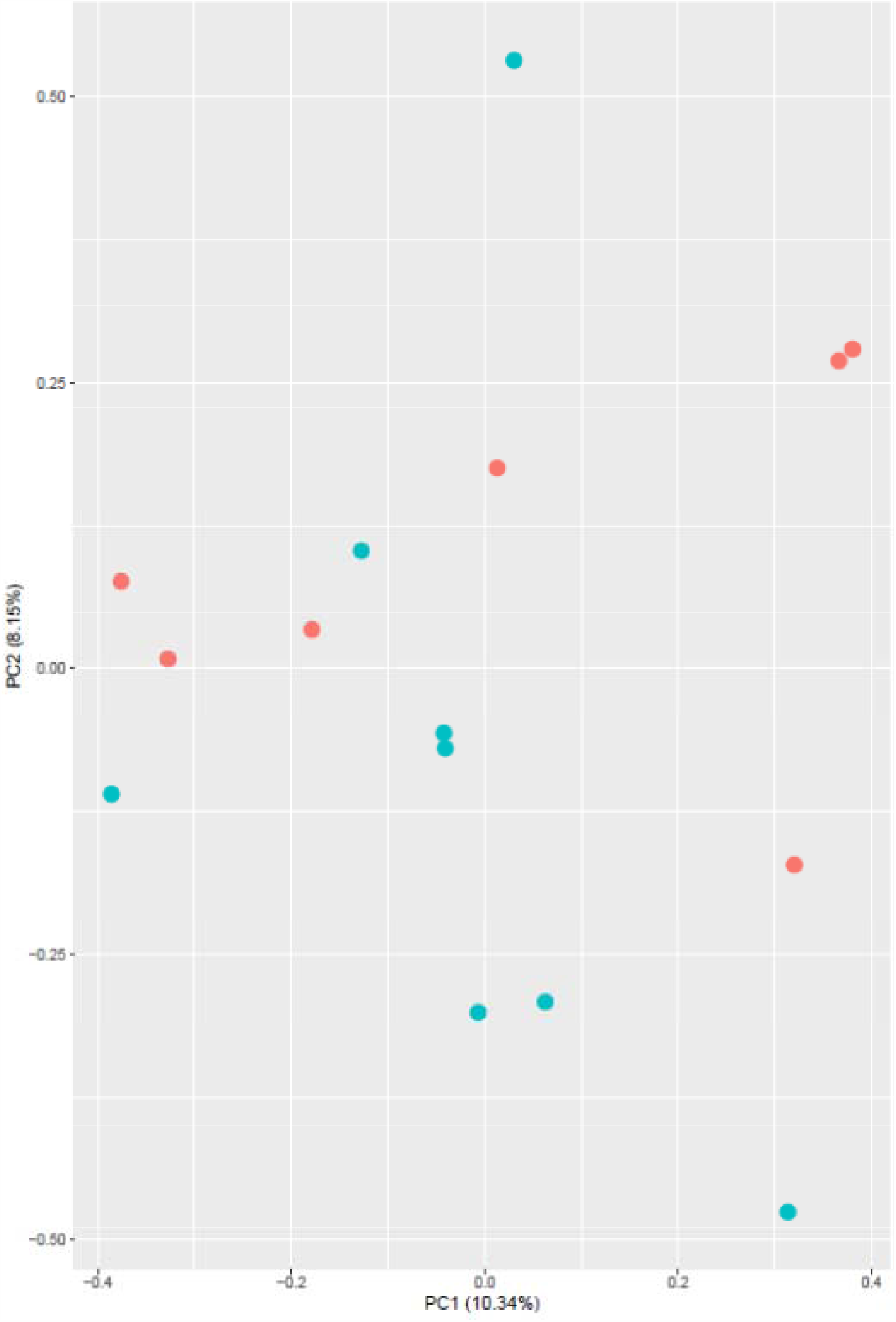
A graph created by the principal component analysis of the >380,000 entropy values related to alternative RNA splicing and alternative transcription start/end. Patients labeled in red died from COVID-19 and surviving patients are labeled with green dots. Mortality rate above PC2=0 is 75% and below is 14% (p=0.04)

## Discussion

This project used deep RNA sequencing of whole blood from participants in the ICU with COVID-19 as a novel diagnostic tool. The protocol extracted RNA from the whole blood, as opposed to fractionating the whole blood specimen. Analysis of whole blood increased the breadth of RNA being sequenced, both cell associated and cell-free, and its simplicity for clinical practice. Alternatively, more complicated techniques, such as single cell sequencing may speak more to pathogenesis but adds to the complexity of the protocol and analysis. Despite its isolation from whole blood, the RNA was of high quality (Supplement Table 1). A novel finding using RNA from whole blood from critically ill participants is that only 62-67% of the reads mapped to the human genome. This is less than the 85-97% of reads that typically map to the reference genome.[16] One major drawback is the timing needed for RNA sequencing and analysis. Currently, sequencing machines take ∼18 hours to generate data. The analysis can take additional time and is not yet clinically standardized. As technology advances and speed improves, however, this data will be increasingly accessible in the care of ICU patients.

SARS-CoV-2 RNA was identified in the unmapped reads in all patients (Figure 1a). This supports that detection of SARS-CoV-2 in the serum has been associated with clinical deterioration[17, 18] and RT-PCR identified the SARS-CoV-2 virus in the blood more often in the ICU patients than in the non-ICU patients.[19] The total number of reads in our dataset did not correlate with any outcomes, including mortality, ARDS, or coagulopathy. The low number of total reads, approximately 700 from nearly 2 billion from all the samples, explains the lack of success from other researchers identifying the virus in the blood. In early reports, RT-PCR directed at the N protein gene identified viral RNA in the plasma in 15% of patients.[20] Our data demonstrate the two most abundant genes in blood were the RNA Dependent RNA Polymerase and the N protein (Figure 1b). With this data we propose these locations (RNA dependent RNA polymerase or N protein) as potential therapeutic or diagnostic targets.[21]

Other authors have called for robust testing for potential co-infections with SARS-CoV-2.[22] With deep sequencing and computational analysis we have identified the RNA from multiple bacteria, viruses, and archaea in all of the specimens, as well as fungal RNA in two participants. This suggests deep RNA sequencing with computational analysis may be a novel tool for the identification of co-infections. More data is required with comparison to gold standards such as blood culture and pathogen-specific PCR. However, RNA sequencing has the benefit of being able to identify all pathogens with known genomes, including both RNA and DNA based organisms. Moreover, unclassified reads that do not align to any known organism (Table 2) or the other sequences that have cellular organism elements (Table 2) could provide evidence of novel pathogens before a genome is sequenced or the pathogen is cultured.

Critically ill COVID-19 patients provide a difficult clinical dilemma as it pertains to antibiotics. In severely ill patients, clinicians are more likely to prescribe antibiotics despite there not being an identified pathogen.[23] With identification of bacteria known to cause human disease from the RNA sequencing data, appropriate antibiotics could be prescribed to these patients. In this data set, we show that there were significantly more counts of *Acinetobacter baumannii* in a portion of patients (Table E2) and this bacterium has been associated with COVID-19.[24] Using a precision medicine approach with these data, patients with significantly elevated levels may potentially be treated with directed antibiotics, in the absence of more time-consuming positive culture data. While there was no difference in survival in participants with versus without identified bacteria in this study, antibiotic use was not standardized or prescribed prospectively based upon our results. In addition, analysis of the unmapped reads aligning to *Acinetobacter baumannii* (averaging over 50,000 among the six with increased reads) could provide insights into genes that are expressed in critical illness and provide novel diagnostic and therapeutic targets.

The immune response to SARS-CoV-2 has been the focus of much research since the pandemic started.[25] The successful use of corticosteroids in the critically ill with COVID-19 emphasizes the importance of the immune system in this disease.[26, 27] Because a significant proportion of COVID-19 patients do not respond to corticosteroids, there are still calls for a more precise approach.[28] PD-1 expression is increased in certain cell populations in patients with COVID-19[3, 29] but the uses of immune checkpoint inhibitors in cancer patients has been associated with more severe COVID-19.[30] Other authors suggest that immune checkpoint inhibitors may be useful in COVID-19.[31] Our data shows that patients who died had increased expression of PD-L1 and PD-L2 (Figure E1, CD274 and PDCD1Lg1, Table 3). This suggests that immune checkpoint inhibitors targeted against the PD-1 system might be considered in those patients identified to have increased expression of PD-L1 and PD-L2 because of their higher risk of death after ICU admission.

Numerous other immune targets are identified from these genomic changes. N4BP1 is induced by interferon and the interferon response has been implicated in COVID-19.[32, 33] Our data supports the role for interferons in COVID-19 as patients who died had 2.5-fold increase in expression of interferon 1 alpha (Table E3, IFNA1). Clinical features of COVID-19 also correlate with some of the genes identified. OR6C4 is an olfactory gene which we identified has exhibiting a 5 fold increased in expression in patients that died (Table 3, Table E3). This finding suggests that loss of smell may signify milder disease among patients in the ICU. Thrombotic complications are common in COVID-19 patients (9.5%) and patients admitted to the ICU have a higher incidence of venous thromboembolism.[34] Patients who died have significant decrease in gene expression and multiple changes in alternative transcription end (Table 3, Table E3 and E5) of both NRP1 and NRP2. Both these genes are associated with coagulation[35] and the COVID-19 spike protein binds both these receptors.[36] Previous work has shown that there is increased expression in both genes in the lungs of patients with COVID-19 when compared to controls.[37] In our study, the decrease NRP1 and NRP2 were seen in ICU patients who died compared to ICU patients who survived.

Many studies have attempted to utilize clinical data to predict mortality in COVID-19[38, 39] and some focus on cytokines.[40] For simplicity all these attempt to identify a few variables to predict mortality. Here we utilize over 380,000 variables with PCA to create a figure that improves mortality prediction based upon where the patient is on the graph (Figure 2, 75% versus 14%). A limitation to this form of analysis is that the PCA cannot identify a specific gene or event most responsible for outcomes; it uses all 380,000 data points. Accurate assessment of prognosis using sequencing technology might be valuable to inform end of life care discussions in the ICU.

Despite the limitations of this single-center study with a small patient number, we were still able to document that deep RNA sequencing and appropriate computational analysis yields valuable insight into the pathogenesis and host response of COVID-19 in critically ill patients. Novel drug targets were identified from SARS-CoV-2 RNA and the host response, including RNA dependent RNA polymerase, the N protein, and the PD-1 immune checkpoint pathway. The presence of pathogen RNA in the blood suggests co-infection should be reconsidered. Most importantly, PCA of the entropy of >380,000 events allowed use to group patients into those likely to die versus those likely to live, and this may be helpful in family discussions with critically ill patients. Translating these results to clinical practice will improve the diagnosis, assessment of prognosis, and therapy of COVID-19.

## Data Availability

Much of the data is supplied in the supplemental materials. Specific patient level data is not included due to privacy.

## Declarations

### Funding

This study was supported by funding from the US National Institutes of Health: P20 GM103652 (SFM, WGF, EOH, AA), T32 HL134625 (AMF, EOH), R01 GM 127472 (WGF), P20 GM121344 (GJN), R01 HL147525 (JSR), R01 HL141268 (CEV), R35 GM118097 (AA).

### Conflicts of Interest

not applicable

### Ethics approval

Institutional Review Board Approval # 411616

### Availability of data

See online supplement for publication of extensive data.

### Code availability

All code used is cited in the text

### Author Contributions

Drs. Monaghan and Fredericks had full access to all of the data in the study and take responsibility for the integrity of the data and the accuracy of the data analysis.

#### Concept and design

Monaghan, Fredericks, Jentzsch, Gandhi, Nau, Levy, Ayala

#### Acquisition, analysis, or interpretation of data

Monaghan, Fredericks, Jentzsch, Cohen, Gandhi, Nau, Ayala.

#### Drafting of the manuscript

Monaghan, Fredericks.

#### Critical revision of the manuscript for important intellectual content

Cioffi, Fairbrother, Harrington, Nau, Reichner, Ventetuolo, Levy, Ayala

#### Statistical analysis

Monaghan, Fredericks, Jentzsch, Gandhi

#### Obtained funding

Monaghan, Fredericks, Fairbrother, Harrington, Nau, Reichner, Ventetuolo, Ayala

#### Administrative, technical, or material support

Cioffi, Nau, Reichner, Ventetuolo, Levy, Ayala

#### Supervision

Monaghan, Cioffi, Levy, Ayala

#### Declaration of Interests

None

Figure E1: Volcano plot showing genes that have significant gene expression differences between patients who died from COVID-19 and those who survived. Red dots with labels have p<0.05 and greater than 1.5 log_2_ fold change between the groups.

## Notes

### Competing Interest Statement

The authors have declared no competing interest.

### Author Declarations

The Rhode Island Hospital Institutional Review Board approved this study (# 411616). Informed consent was given by all patients or surrogates.

