## Supplementary figures and images for "Deep RNA Sequencing of Intensive Care Unit Patients with COVID-19"

### Supplemental Figure 1

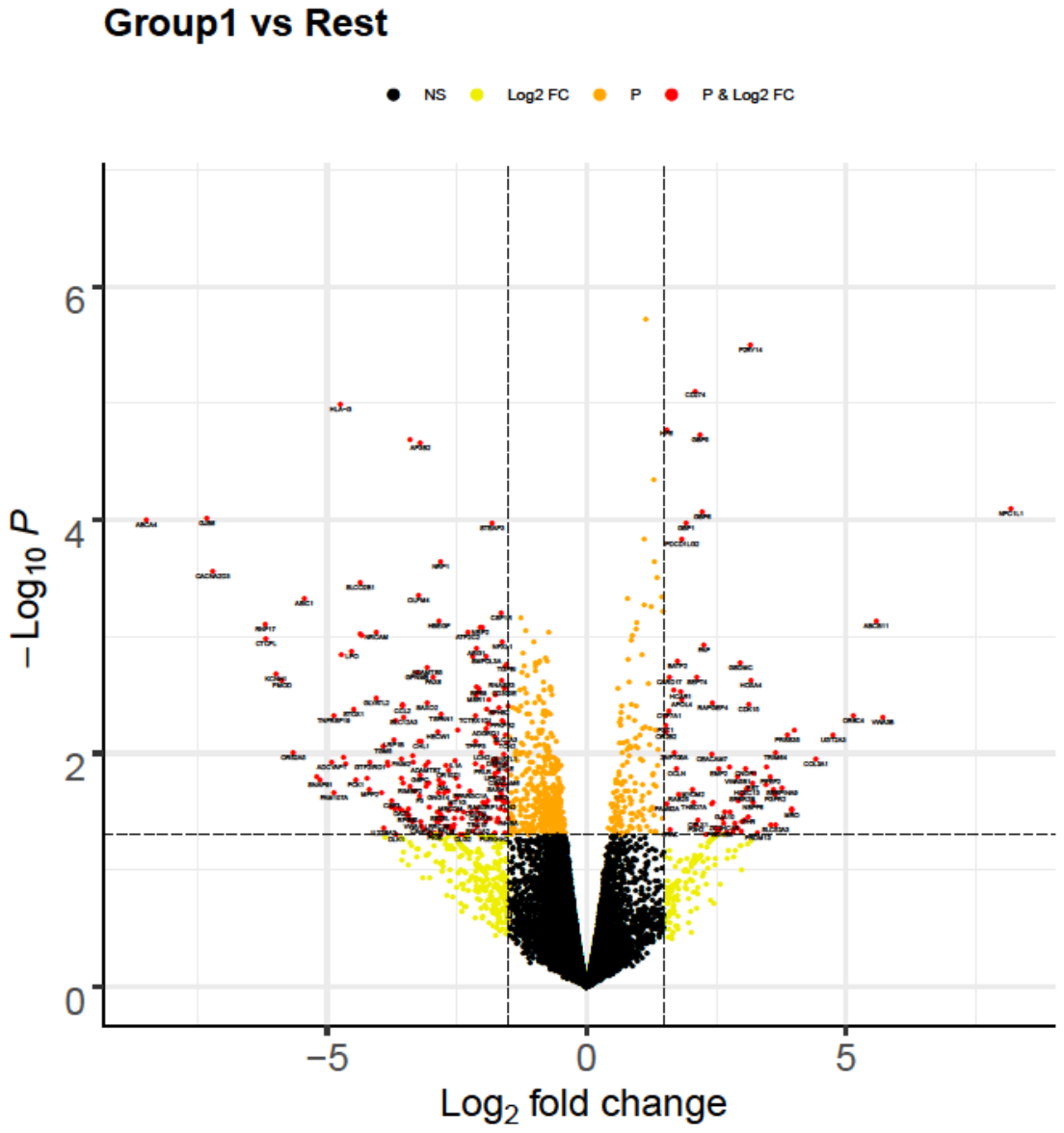
