## Supplementary material for "Deep RNA Sequencing of Intensive Care Unit Patients with COVID-19": File 1 of supplemental tables

Table S1 Sequencing Characteristics of each sample, quality assurance, and mapping

| Patient | RNA Integrity Number (RIN) | Concentration of RNA (ng/uL) | Number of Reads | Yield (Mbases) | Mean Quality Score (FastQC) | % Bases >= 30 | % Mapped | % Unmapped | %Multi-Mapped |
| --- | --- | --- | --- | --- | --- | --- | --- | --- | --- |
| 1 | 6.6 | 23.8 | 124,950,364 | 37485 | 35.69 | 92.71 | 64.84 | 30.96 | 4.19 |
| 2 | 6.8 | 38.5 | 126,204,089 | 37861 | 35.66 | 92.57 | 64.03 | 31.8 | 4.17 |
| 3 | 7.4 | 32.5 | 118,689,370 | 35607 | 35.53 | 91.99 | 62.86 | 33.34 | 3.81 |
| 4 | 8.2 | 10.9 | 155,014,322 | 46504 | 35.75 | 92.91 | 64.76 | 31.03 | 4.21 |
| 5 | 7.1 | 29.1 | 127,238,475 | 38172 | 35.69 | 92.63 | 63.93 | 32.04 | 4.04 |
| 6 | 7.8 | 49.1 | 119,389,888 | 35817 | 35.57 | 92.12 | 62.57 | 33.61 | 3.81 |
| 7 | 8.5 | 62.4 | 111,161,680 | 33349 | 35.77 | 93.08 | 65.25 | 30.68 | 4.06 |
| 8 | 7.1 | 108 | 142,740,108 | 42822 | 35.58 | 92.19 | 62.78 | 33.41 | 3.81 |
| 9 | 7.7 | 65.9 | 148,001,485 | 44400 | 35.62 | 92.28 | 65.48 | 30.27 | 4.25 |
| 10 | 8.5 | 37.6 | 140,672,037 | 42202 | 35.59 | 92.19 | 65.16 | 30.18 | 4.66 |
| 11 | 8.7 | 47.2 | 131,721,720 | 39517 | 35.64 | 92.4 | 66.75 | 28.77 | 4.47 |
| 12 | 9.1 | 13.2 | 125,687,784 | 37706 | 35.66 | 92.49 | 66.51 | 28.71 | 4.78 |
| 13 | 6.9 | 62.1 | 121,874,221 | 36562 | 35.65 | 92.42 | 65.33 | 30.73 | 3.94 |
| 14 | 7.5 | 92.3 | 122,677,639 | 36803 | 35.62 | 92.29 | 64.66 | 30.55 | 4.78 |
| 15 | 7.7 | 67.3 | 118,714,356 | 35614 | 35.66 | 92.53 | 65.34 | 30.38 | 4.29 |

Table S2: Bacteria with most reads per patient

| Patient 1 | Reads | Taxonomy | Name |
| --- | --- | --- | --- |
|  | 54445 | 470 | Acinetobacter baumannii |
|  | 14320 | 1324352 | Chryseobacterium gallinarum |
|  | 1609 | 1200984 | Streptomyces lividans 1326 |
|  | 1164 | 1747 | Cutibacterium acnes |
|  | 856 | 1658665 | Mitsuaria sp. 7 |
|  | 841 | 190721 | Ralstonia insidiosa |
|  | 790 | 1833 | Rhodococcus erythropolis |
|  | 665 | 1807790 | Rhodococcus sp. BH4 |
|  | 595 | 1045808 | Rhodococcus sp. YL-1 |
|  | 571 | 400667 | Acinetobacter baumannii ATCC 17978 |
| Patient 2 | Reads | Taxonomy | Name |
|  | 54309 | 470 | Acinetobacter baumannii |
|  | 16721 | 1324352 | Chryseobacterium gallinarum |
|  | 2916 | 1747 | Cutibacterium acnes |
|  | 1828 | 1200984 | Streptomyces lividans 1326 |
|  | 1128 | 1658665 | Mitsuaria sp. 7 |
|  | 1029 | 190721 | Ralstonia insidiosa |
|  | 722 | 1404 | Bacillus megaterium |
|  | 566 | 400667 | Acinetobacter baumannii ATCC 17978 |
|  | 549 | 1768242 | Paucibacter sp. KCTC 42545 |
|  | 543 | 398578 | Delftia acidovorans SPH-1 |
| Patient 3 | Reads | Taxonomy | Name |
|  | 45337 | 470 | Acinetobacter baumannii |
|  | 28595 | 1324352 | Chryseobacterium gallinarum |
|  | 2252 | 1200984 | Streptomyces lividans 1326 |
|  | 2238 | 1658665 | Mitsuaria sp. 7 |
|  | 2027 | 190721 | Ralstonia insidiosa |
|  | 1895 | 1747 | Cutibacterium acnes |
|  | 1274 | 76731 | Roseateles depolymerans |
|  | 1176 | 1768242 | Paucibacter sp. KCTC 42545 |
|  | 1089 | 80866 | Delftia acidovorans |
|  | 1074 | 983917 | Rubrivivax gelatinosus IL144 |
| Patient 4 | Reads | Taxonomy | Name |
|  | 12179 | 1324352 | Chryseobacterium gallinarum |
|  | 1277 | 1833 | Rhodococcus erythropolis |
|  | 1221 | 1200984 | Streptomyces lividans 1326 |
|  | 879 | 1807790 | Rhodococcus sp. BH4 |
|  | 874 | 1045808 | Rhodococcus sp. YL-1 |
|  | 814 | 1658665 | Mitsuaria sp. 7 |
|  | 791 | 1747 | Cutibacterium acnes |
|  | 759 | 190721 | Ralstonia insidiosa |
|  | 693 | 334542 | Rhodococcus qingshengii |
|  | 486 | 2490853 | Rhodococcus sp. NJ-530 |
| Patient 5 | Reads | Taxonomy | Name |
|  | 15287 | 1324352 | Chryseobacterium gallinarum |
|  | 1211 | 1747 | Cutibacterium acnes |
|  | 1138 | 1200984 | Streptomyces lividans 1326 |
|  | 1088 | 1404 | Bacillus megaterium |
|  | 1036 | 190721 | Ralstonia insidiosa |
|  | 1031 | 1658665 | Mitsuaria sp. 7 |
|  | 738 | 1833 | Rhodococcus erythropolis |
|  | 682 | 1807790 | Rhodococcus sp. BH4 |
|  | 599 | 1045808 | Rhodococcus sp. YL-1 |
|  | 594 | 1768242 | Paucibacter sp. KCTC 42545 |
| Patient 6 | Reads | Taxonomy | Name |
|  | 29265 | 1324352 | Chryseobacterium gallinarum |
|  | 2354 | 1747 | Cutibacterium acnes |
|  | 2284 | 1658665 | Mitsuaria sp. 7 |
|  | 2064 | 190721 | Ralstonia insidiosa |
|  | 1473 | 1200984 | Streptomyces lividans 1326 |
|  | 1167 | 1768242 | Paucibacter sp. KCTC 42545 |
|  | 1166 | 398578 | Delftia acidovorans SPH-1 |
|  | 1117 | 76731 | Roseateles depolymerans |
|  | 1043 | 1833 | Rhodococcus erythropolis |
|  | 1023 | 983917 | Rubrivivax gelatinosus IL144 |
| Patient 7 | Reads | Taxonomy | Name |
|  | 11938 | 1324352 | Chryseobacterium gallinarum |
|  | 1379 | 1200984 | Streptomyces lividans 1326 |
|  | 1294 | 1747 | Cutibacterium acnes |
|  | 982 | 1807790 | Rhodococcus sp. BH4 |
|  | 947 | 1833 | Rhodococcus erythropolis |
|  | 772 | 1045808 | Rhodococcus sp. YL-1 |
|  | 769 | 1658665 | Mitsuaria sp. 7 |
|  | 741 | 190721 | Ralstonia insidiosa |
|  | 664 | 573 | Klebsiella pneumoniae |
|  | 547 | 334542 | Rhodococcus qingshengii |
| Patient 8 | Reads | Taxonomy | Name |
|  | 30695 | 1324352 | Chryseobacterium gallinarum |
|  | 5470 | 1747 | Cutibacterium acnes |
|  | 1941 | 1200984 | Streptomyces lividans 1326 |
|  | 1888 | 1658665 | Mitsuaria sp. 7 |
|  | 1859 | 190721 | Ralstonia insidiosa |
|  | 1283 | 1833 | Rhodococcus erythropolis |
|  | 1073 | 1404 | Bacillus megaterium |
|  | 1061 | 76731 | Roseateles depolymerans |
|  | 1055 | 1768242 | Paucibacter sp. KCTC 42545 |
|  | 1004 | 1807790 | Rhodococcus sp. BH4 |
| Patient 9 | Reads | Taxonomy | Name |
|  | 53963 | 470 | Acinetobacter baumannii |
|  | 15935 | 1324352 | Chryseobacterium gallinarum |
|  | 6945 | 35841 | Bacillus thermoamylovorans |
|  | 2592 | 44008 | Enterococcus cecorum |
|  | 2457 | 1200984 | Streptomyces lividans 1326 |
|  | 2166 | 1422 | Geobacillus stearothermophilus |
|  | 1828 | 1747 | Cutibacterium acnes |
|  | 1625 | 743721 | Pseudoxanthomonas suwonensis 11-1 |
|  | 951 | 1658665 | Mitsuaria sp. 7 |
|  | 886 | 33934 | Anoxybacillus flavithermus |
| Patient 10 | Reads | Taxonomy | Name |
|  | 53269 | 470 | Acinetobacter baumannii |
|  | 14175 | 1324352 | Chryseobacterium gallinarum |
|  | 4703 | 571915 | Corynebacterium mustelae |
|  | 4408 | 1260 | Finegoldia magna |
|  | 3602 | 2081703 | Peptostreptococcaceae bacterium oral taxon 929 |
|  | 3413 | 54005 | Peptoniphilus harei |
|  | 2358 | 1200984 | Streptomyces lividans 1326 |
|  | 2326 | 28131 | Prevotella intermedia |
|  | 1663 | 2590900 | Tenuifilum thalassicum |
|  | 1660 | 36874 | Porphyromonas cangingivalis |
| Patient 11 | Reads | Taxonomy | Name |
|  | 44109 | 470 | Acinetobacter baumannii |
|  | 12541 | 1324352 | Chryseobacterium gallinarum |
|  | 1901 | 1200984 | Streptomyces lividans 1326 |
|  | 1296 | 1404 | Bacillus megaterium |
|  | 738 | 190721 | Ralstonia insidiosa |
|  | 733 | 1658665 | Mitsuaria sp. 7 |
|  | 710 | 1747 | Cutibacterium acnes |
|  | 472 | 400667 | Acinetobacter baumannii ATCC 17978 |
|  | 441 | 106654 | Acinetobacter nosocomialis |
|  | 426 | 1768242 | Paucibacter sp. KCTC 42545 |
| Patient 12 | Reads | Taxonomy | Name |
|  | 9881 | 1324352 | Chryseobacterium gallinarum |
|  | 1279 | 1200984 | Streptomyces lividans 1326 |
|  | 1246 | 1833 | Rhodococcus erythropolis |
|  | 979 | 1807790 | Rhodococcus sp. BH4 |
|  | 838 | 1045808 | Rhodococcus sp. YL-1 |
|  | 753 | 1404 | Bacillus megaterium |
|  | 651 | 1747 | Cutibacterium acnes |
|  | 629 | 1658665 | Mitsuaria sp. 7 |
|  | 614 | 334542 | Rhodococcus qingshengii |
|  | 575 | 190721 | Ralstonia insidiosa |
| Patient 13 | Reads | Taxonomy | Name |
|  | 11006 | 1324352 | Chryseobacterium gallinarum |
|  | 1068 | 1200984 | Streptomyces lividans 1326 |
|  | 1004 | 1404 | Bacillus megaterium |
|  | 888 | 1747 | Cutibacterium acnes |
|  | 710 | 1833 | Rhodococcus erythropolis |
|  | 670 | 1658665 | Mitsuaria sp. 7 |
|  | 669 | 1807790 | Rhodococcus sp. BH4 |
|  | 617 | 190721 | Ralstonia insidiosa |
|  | 606 | 1045808 | Rhodococcus sp. YL-1 |
|  | 491 | 334542 | Rhodococcus qingshengii |
| Patient 14 | Reads | Taxonomy | Name |
|  | 10791 | 1324352 | Chryseobacterium gallinarum |
|  | 1981 | 1200984 | Streptomyces lividans 1326 |
|  | 1108 | 1833 | Rhodococcus erythropolis |
|  | 925 | 1807790 | Rhodococcus sp. BH4 |
|  | 917 | 1045808 | Rhodococcus sp. YL-1 |
|  | 754 | 1747 | Cutibacterium acnes |
|  | 726 | 1658665 | Mitsuaria sp. 7 |
|  | 630 | 190721 | Ralstonia insidiosa |
|  | 544 | 334542 | Rhodococcus qingshengii |
|  | 542 | 2490853 | Rhodococcus sp. NJ-530 |
| Patient 15 | Reads | Taxonomy | Name |
|  | 17620 | 1324352 | Chryseobacterium gallinarum |
|  | 1535 | 1200984 | Streptomyces lividans 1326 |
|  | 1128 | 1747 | Cutibacterium acnes |
|  | 1093 | 1658665 | Mitsuaria sp. 7 |
|  | 964 | 1833 | Rhodococcus erythropolis |
|  | 933 | 1807790 | Rhodococcus sp. BH4 |
|  | 885 | 190721 | Ralstonia insidiosa |
|  | 842 | 1045808 | Rhodococcus sp. YL-1 |
|  | 752 | 573 | Klebsiella pneumoniae |
|  | 604 | 1768242 | Paucibacter sp. KCTC 42545 |
