## Supplementary material for "Deep RNA Sequencing of Intensive Care Unit Patients with COVID-19": File 2 of supplemental tables

Table S3. Genes with Significant Gene Expression Differences between Patients who died from COVID-19 and those who lived.

| gene | id | logFC | PValue |
| --- | --- | --- | --- |
| ABCA13 | ENSG00000179869 | -2.19182 | 0.00146 |
| ABCA4 | ENSG00000198691 | -8.48007 | 9.89E-05 |
| ABCB11 | ENSG00000073734 | 5.581795 | 0.00074 |
| AC009163.2 | ENSG00000260092 | -3.42695 | 0.034232 |
| AC009879.2 | ENSG00000285655 | -2.22409 | 0.032291 |
| AC087498.1 | ENSG00000180042 | -2.74587 | 0.021536 |
| AC109583.3 | ENSG00000284672 | 2.920314 | 0.025239 |
| ADAMTS3 | ENSG00000156140 | -2.05348 | 0.049721 |
| ADAMTS5 | ENSG00000154736 | -3.07599 | 0.001824 |
| ADAMTS7 | ENSG00000136378 | -3.10046 | 0.012815 |
| ADARB2 | ENSG00000185736 | -2.51876 | 0.016645 |
| ADCYAP1 | ENSG00000141433 | -4.91146 | 0.011941 |
| ADGRD1 | ENSG00000111452 | -1.94021 | 0.006206 |
| AL672142.1 | ENSG00000251184 | -3.343 | 0.012078 |
| AMPD1 | ENSG00000116748 | -3.46188 | 0.032031 |
| ANKRD2 | ENSG00000165887 | 3.550296 | 0.041721 |
| AP3B2 | ENSG00000103723 | -3.20494 | 2.19E-05 |
| APOD | ENSG00000189058 | -4.72501 | 0.001449 |
| APOL4 | ENSG00000100336 | 1.832949 | 0.003493 |
| ARC | ENSG00000198576 | -1.5738 | 0.048209 |
| ARG1 | ENSG00000118520 | -2.12305 | 0.001258 |
| ARHGAP20 | ENSG00000137727 | -3.01777 | 0.049347 |
| ARHGAP22 | ENSG00000128805 | -1.57161 | 0.031277 |
| ARPIN | ENSG00000242498 | -1.51562 | 0.004002 |
| ARSJ | ENSG00000180801 | 2.906552 | 0.044304 |
| ASIC1 | ENSG00000110881 | -5.44148 | 0.000479 |
| ASTL | ENSG00000188886 | -1.77099 | 0.048881 |
| ATOH8 | ENSG00000168874 | -2.00774 | 0.000837 |
| ATP1A4 | ENSG00000132681 | -4.33622 | 0.000967 |
| ATP2C2 | ENSG00000064270 | -2.2905 | 0.000927 |
| AZU1 | ENSG00000172232 | -1.80291 | 0.011494 |
| BATF2 | ENSG00000168062 | 1.749183 | 0.001651 |
| BEX1 | ENSG00000133169 | -1.69396 | 0.03829 |
| BOC | ENSG00000144857 | -2.36754 | 0.031813 |
| C11orf52 | ENSG00000149300 | 2.702639 | 0.04835 |
| C1orf127 | ENSG00000175262 | -1.70772 | 0.036783 |
| C1orf61 | ENSG00000125462 | -1.77133 | 0.015696 |
| C1QTNF9B | ENSG00000205863 | 3.466413 | 0.013079 |
| C8orf34 | ENSG00000165084 | -3.1741 | 0.04285 |
| CACNA2D3 | ENSG00000157445 | -7.2042 | 0.000273 |
| CALML6 | ENSG00000169885 | -2.43744 | 0.049571 |
| CARD17 | ENSG00000255221 | 1.593221 | 0.002263 |
| CBARP | ENSG00000099625 | -2.45934 | 0.019434 |
| CCL2 | ENSG00000108691 | -3.54842 | 0.00394 |
| CCL20 | ENSG00000115009 | -3.5965 | 0.029722 |
| CCL8 | ENSG00000108700 | -3.54394 | 0.003825 |
| CCNB3 | ENSG00000147082 | -3.94973 | 0.021887 |
| CD1E | ENSG00000158488 | -4.68331 | 0.010912 |
| CD24 | ENSG00000272398 | -1.75906 | 0.012575 |
| CD274 | ENSG00000120217 | 2.093773 | 7.83E-06 |
| CD300E | ENSG00000186407 | -1.58945 | 0.002761 |
| CDK15 | ENSG00000138395 | 3.12364 | 0.003794 |
| CEACAM6 | ENSG00000086548 | -1.59221 | 0.01655 |
| CEACAM7 | ENSG00000007306 | 2.409377 | 0.010295 |
| CEACAM8 | ENSG00000124469 | -1.70644 | 0.014435 |
| CELF4 | ENSG00000101489 | -4.64558 | 0.012382 |
| CFAP97D1 | ENSG00000231256 | 1.68038 | 0.002911 |
| CHL1 | ENSG00000134121 | -3.18509 | 0.008001 |
| CLDN16 | ENSG00000113946 | -2.61734 | 0.042432 |
| CLEC4F | ENSG00000152672 | -2.39808 | 0.036653 |
| CNDP1 | ENSG00000150656 | 3.065364 | 0.013742 |
| CNR1 | ENSG00000118432 | -1.61972 | 0.018722 |
| CNTNAP2 | ENSG00000174469 | -1.62698 | 0.022931 |
| COL3A1 | ENSG00000168542 | 4.421043 | 0.0111 |
| CPLX1 | ENSG00000168993 | 2.148074 | 0.036967 |
| CPNE7 | ENSG00000178773 | 3.0448 | 0.036932 |
| CRISP3 | ENSG00000096006 | -1.65978 | 0.025131 |
| CRYAB | ENSG00000109846 | -2.06123 | 0.033092 |
| CSF1R | ENSG00000182578 | -1.64517 | 0.000638 |
| CSF3 | ENSG00000108342 | -3.75706 | 0.025394 |
| CTCFL | ENSG00000124092 | -6.17853 | 0.001036 |
| CYP7A1 | ENSG00000167910 | 1.585266 | 0.004376 |
| DACH2 | ENSG00000126733 | 3.094649 | 0.020178 |
| DAZL | ENSG00000092345 | -3.56058 | 0.030485 |
| DDIT4L | ENSG00000145358 | -2.11969 | 0.029905 |
| DEFA1 | ENSG00000206047 | -1.92729 | 0.004211 |
| DEFA4 | ENSG00000164821 | -1.73896 | 0.01121 |
| DIRAS2 | ENSG00000165023 | -3.68252 | 0.0052 |
| DLC1 | ENSG00000164741 | -1.80369 | 0.004497 |
| DLG2 | ENSG00000150672 | -2.38187 | 0.049502 |
| DLK1 | ENSG00000185559 | -3.6699 | 0.049439 |
| DRP2 | ENSG00000102385 | -3.768 | 0.029502 |
| DZIP1L | ENSG00000158163 | -1.59263 | 0.025503 |
| EIF5AL1 | ENSG00000253626 | -2.1238 | 0.002706 |
| EMP2 | ENSG00000213853 | 2.548333 | 0.013729 |
| EPHB2 | ENSG00000133216 | -1.69089 | 0.004093 |
| EPS8 | ENSG00000151491 | -2.08322 | 0.002807 |
| ERG | ENSG00000157554 | -1.65272 | 0.022094 |
| ERICH2 | ENSG00000204334 | -2.81104 | 0.03728 |
| ESRRB | ENSG00000119715 | -2.57513 | 0.044101 |
| F3 | ENSG00000117525 | -3.21227 | 0.023016 |
| FAIM2 | ENSG00000135472 | -3.57054 | 0.011249 |
| FAM107A | ENSG00000168309 | -4.86483 | 0.021915 |
| FAM47E-STBD1 | ENSG00000272414 | -1.73965 | 0.008276 |
| FAM92A | ENSG00000188343 | 1.548593 | 0.027174 |
| FAP | ENSG00000078098 | 2.260431 | 0.001188 |
| FGF12 | ENSG00000114279 | -3.02745 | 0.028637 |
| FGFR2 | ENSG00000066468 | 3.631718 | 0.022573 |
| FIGN | ENSG00000182263 | -1.95065 | 0.038785 |
| FMOD | ENSG00000122176 | -5.86736 | 0.002394 |
| FOXQ1 | ENSG00000164379 | -1.66482 | 0.030377 |
| FPR3 | ENSG00000187474 | -1.87987 | 0.003486 |
| FST | ENSG00000134363 | 3.197903 | 0.018276 |
| FUT1 | ENSG00000174951 | -1.85312 | 0.036512 |
| G6PC | ENSG00000131482 | -3.20513 | 0.015232 |
| GAL | ENSG00000069482 | -2.76045 | 0.018115 |
| GALNT9 | ENSG00000182870 | -3.13264 | 0.045021 |
| GBP1 | ENSG00000117228 | 1.917507 | 0.000106 |
| GBP5 | ENSG00000154451 | 2.189412 | 1.85E-05 |
| GBP6 | ENSG00000183347 | 2.229525 | 8.46E-05 |
| GHR | ENSG00000112964 | 3.116139 | 0.035661 |
| GJA10 | ENSG00000135355 | 2.656173 | 0.031582 |
| GJB6 | ENSG00000121742 | -7.31728 | 9.66E-05 |
| GLYATL1 | ENSG00000166840 | -1.59199 | 0.010213 |
| GLYATL2 | ENSG00000156689 | -4.05262 | 0.003405 |
| GNG14 | ENSG00000283980 | -2.85982 | 0.022098 |
| GPNMB | ENSG00000136235 | -3.27011 | 0.002022 |
| GPR162 | ENSG00000250510 | -1.60714 | 0.005379 |
| GPR33 | ENSG00000214943 | -2.68453 | 0.042065 |
| GPR84 | ENSG00000139572 | -1.52939 | 0.013872 |
| GRAMD2A | ENSG00000175318 | -3.22955 | 0.007859 |
| GSDMC | ENSG00000147697 | 2.962965 | 0.001677 |
| GTF2IRD1 | ENSG00000006704 | -4.17342 | 0.011838 |
| GUCY1A2 | ENSG00000152402 | -2.54384 | 0.036016 |
| HAS2 | ENSG00000170961 | -2.5994 | 0.046168 |
| HBEGF | ENSG00000113070 | -2.84653 | 0.000727 |
| HCAR1 | ENSG00000196917 | 1.814637 | 0.002998 |
| HECW1 | ENSG00000002746 | -2.86236 | 0.006519 |
| HESX1 | ENSG00000163666 | -2.05249 | 0.049178 |
| HFE | ENSG00000010704 | 1.541127 | 1.67E-05 |
| HLA-G | ENSG00000204632 | -4.74041 | 1.04E-05 |
| HOXA4 | ENSG00000197576 | 3.16283 | 0.002352 |
| HOXC13 | ENSG00000123364 | 3.08613 | 0.020187 |
| HS3ST6 | ENSG00000162040 | 2.75156 | 0.013038 |
| HTR2A | ENSG00000102468 | -2.82188 | 0.018475 |
| IFNA1 | ENSG00000197919 | 2.55039 | 0.045365 |
| IL1A | ENSG00000115008 | -2.53042 | 0.011772 |
| IL22RA2 | ENSG00000164485 | -3.90858 | 0.044352 |
| IL23R | ENSG00000162594 | 2.434776 | 0.026008 |
| INHBA | ENSG00000122641 | -1.51252 | 0.036002 |
| IP6K3 | ENSG00000161896 | -3.04833 | 0.011784 |
| KCNH1 | ENSG00000143473 | -5.98993 | 0.002118 |
| KCNJ8 | ENSG00000121361 | -3.07368 | 0.018289 |
| KCNMA1 | ENSG00000156113 | -2.18965 | 0.047068 |
| KCTD19 | ENSG00000168676 | -2.71566 | 0.021017 |
| KLF14 | ENSG00000266265 | -3.40564 | 2.09E-05 |
| KRT81 | ENSG00000205426 | 2.304019 | 0.049556 |
| LCN2 | ENSG00000148346 | -2.02681 | 0.009782 |
| LGR4 | ENSG00000205213 | -1.7732 | 0.007234 |
| LPO | ENSG00000167419 | -4.52429 | 0.001331 |
| LRP1B | ENSG00000168702 | -3.71689 | 0.007657 |
| LRRC36 | ENSG00000159708 | -2.883 | 0.039068 |
| LRRTM3 | ENSG00000198739 | -2.49244 | 0.031192 |
| LY6G6D | ENSG00000244355 | 2.488845 | 0.043702 |
| LYNX1 | ENSG00000180155 | -1.94791 | 0.027297 |
| MECOM | ENSG00000085276 | -2.629 | 0.027555 |
| MMP27 | ENSG00000137675 | -3.34805 | 0.010705 |
| MMP8 | ENSG00000118113 | -2.1464 | 0.012327 |
| MPP2 | ENSG00000108852 | -4.18349 | 0.020787 |
| MRO | ENSG00000134042 | 3.952571 | 0.031128 |
| MS4A3 | ENSG00000149516 | -1.58659 | 0.018736 |
| MSR1 | ENSG00000038945 | -2.13024 | 0.003188 |
| MT1E | ENSG00000169715 | -1.57291 | 0.012458 |
| MT1G | ENSG00000125144 | -2.49088 | 0.024385 |
| MT1M | ENSG00000205364 | -2.80341 | 0.04976 |
| MYOM2 | ENSG00000036448 | 2.045115 | 0.020264 |
| NBPF6 | ENSG00000186086 | 3.203384 | 0.026739 |
| NEU4 | ENSG00000204099 | -2.57551 | 0.043161 |
| NFXL1 | ENSG00000170448 | -1.62626 | 0.001129 |
| NLGN1 | ENSG00000169760 | -3.56353 | 0.016392 |
| NNMT | ENSG00000166741 | -3.43386 | 0.030174 |
| NPAS4 | ENSG00000174576 | -2.87979 | 0.032172 |
| NPC1L1 | ENSG00000015520 | 8.17472 | 8.09E-05 |
| NRCAM | ENSG00000091129 | -4.0523 | 0.000913 |
| NRP1 | ENSG00000099250 | -2.81239 | 0.000229 |
| NRP2 | ENSG00000118257 | -2.05228 | 0.00084 |
| OCLN | ENSG00000197822 | 1.732905 | 0.013629 |
| OLFM4 | ENSG00000102837 | -3.24031 | 0.000442 |
| OLR1 | ENSG00000173391 | -1.72426 | 0.024423 |
| OR10Z1 | ENSG00000198967 | -2.66396 | 0.014213 |
| OR1C1 | ENSG00000221888 | -2.60619 | 0.042725 |
| OR1E1 | ENSG00000180016 | 2.988241 | 0.038349 |
| OR1N2 | ENSG00000171501 | -3.54002 | 0.018116 |
| OR2AJ1 | ENSG00000177275 | -3.04233 | 0.017813 |
| OR2B2 | ENSG00000168131 | 1.529864 | 0.006643 |
| OR2M3 | ENSG00000228198 | -2.55727 | 0.041684 |
| OR2T5 | ENSG00000203661 | -3.83719 | 0.011831 |
| OR51E1 | ENSG00000180785 | 2.518595 | 0.048885 |
| OR52A5 | ENSG00000171944 | -5.6532 | 0.010053 |
| OR56A1 | ENSG00000180934 | -2.48102 | 0.006305 |
| OR6C4 | ENSG00000179626 | 5.140443 | 0.004808 |
| OR7D2 | ENSG00000188000 | -5.19908 | 0.016102 |
| P2RY14 | ENSG00000174944 | 3.155904 | 3.22E-06 |
| P3H2 | ENSG00000090530 | 2.102961 | 0.041323 |
| PALM | ENSG00000099864 | -2.70801 | 0.042856 |
| PAX6 | ENSG00000007372 | -2.9587 | 0.002218 |
| PCDHB5 | ENSG00000113209 | 2.561043 | 0.045806 |
| PCDHGB1 | ENSG00000254221 | -3.02383 | 0.04876 |
| PCK1 | ENSG00000124253 | -4.44992 | 0.017065 |
| PDCD1LG2 | ENSG00000197646 | 1.836185 | 0.000145 |
| PFKFB2 | ENSG00000123836 | -1.62643 | 0.005304 |
| PID1 | ENSG00000153823 | -1.89416 | 0.005518 |
| PIGR | ENSG00000162896 | -2.67017 | 0.020939 |
| PKIB | ENSG00000135549 | -2.93182 | 0.047701 |
| PLEKHG5 | ENSG00000171680 | 2.983761 | 0.047327 |
| PLEKHH2 | ENSG00000152527 | -1.78756 | 0.049118 |
| PLPP4 | ENSG00000203805 | 4.00358 | 0.006391 |
| PLXNB2 | ENSG00000196576 | -1.5672 | 0.001809 |
| POU4F3 | ENSG00000091010 | 3.596459 | 0.020681 |
| PPARGC1A | ENSG00000109819 | -2.24331 | 0.021179 |
| PPEF2 | ENSG00000156194 | 3.541861 | 0.01609 |
| PRDM13 | ENSG00000112238 | 3.295741 | 0.048674 |
| PRDM16 | ENSG00000142611 | 2.859383 | 0.039918 |
| PRLR | ENSG00000113494 | -2.00329 | 0.013033 |
| PRSS35 | ENSG00000146250 | 3.866753 | 0.006992 |
| PTGES | ENSG00000148344 | -1.58997 | 0.026215 |
| PXT1 | ENSG00000179165 | 1.521497 | 0.005861 |
| RAB25 | ENSG00000132698 | 1.768133 | 0.022636 |
| RAPGEF4 | ENSG00000091428 | 2.42815 | 0.003736 |
| RASGRF1 | ENSG00000058335 | -2.00068 | 0.026368 |
| RIMBP2 | ENSG00000060709 | -3.40103 | 0.019112 |
| RIMS4 | ENSG00000101098 | -2.65224 | 0.044379 |
| RNASE3 | ENSG00000169397 | -1.64007 | 0.002403 |
| RNF150 | ENSG00000170153 | 3.950052 | 0.030305 |
| RNF17 | ENSG00000132972 | -6.19309 | 0.00079 |
| RNF182 | ENSG00000180537 | -4.36506 | 0.000955 |
| RPE65 | ENSG00000116745 | -3.45096 | 0.033593 |
| SASH1 | ENSG00000111961 | -1.71882 | 0.018405 |
| SAXO2 | ENSG00000188659 | -3.07421 | 0.003664 |
| SCARA5 | ENSG00000168079 | -3.6946 | 0.029632 |
| SEMA3D | ENSG00000153993 | -2.7122 | 0.012758 |
| Sept4 | ENSG00000108387 | 2.127643 | 0.002205 |
| SERPINA9 | ENSG00000170054 | 3.760873 | 0.019672 |
| SERPINB10 | ENSG00000242550 | -2.07097 | 0.002972 |
| SEZ6L | ENSG00000100095 | -2.82931 | 0.033172 |
| SFTPC | ENSG00000168484 | -2.84498 | 0.016882 |
| SHISA2 | ENSG00000180730 | -1.72961 | 0.039198 |
| SLC12A3 | ENSG00000070915 | -3.52729 | 0.004889 |
| SLC16A9 | ENSG00000165449 | -2.86745 | 0.038943 |
| SLC1A2 | ENSG00000110436 | -2.10957 | 0.042008 |
| SLC1A3 | ENSG00000079215 | -1.56283 | 0.007011 |
| SLC4A10 | ENSG00000144290 | 2.768269 | 0.031642 |
| SLC52A3 | ENSG00000101276 | 3.639265 | 0.041132 |
| SLCO2B1 | ENSG00000137491 | -4.36317 | 0.000344 |
| SMIM2 | ENSG00000139656 | -2.59766 | 0.045626 |
| SMIM22 | ENSG00000267795 | -1.90924 | 0.025627 |
| SMPDL3A | ENSG00000172594 | -1.93655 | 0.001492 |
| SNAP25 | ENSG00000132639 | 2.813854 | 0.04758 |
| SNAP91 | ENSG00000065609 | -5.13606 | 0.016724 |
| SOX17 | ENSG00000164736 | 2.619591 | 0.036523 |
| SPACA1 | ENSG00000118434 | -2.78306 | 0.035128 |
| SPATA31A1 | ENSG00000204849 | 2.698442 | 0.048549 |
| SPATA31E1 | ENSG00000177992 | -3.09363 | 0.019298 |
| SPRR2B | ENSG00000196805 | 2.990849 | 0.02288 |
| STAC | ENSG00000144681 | 1.608918 | 0.04493 |
| STBD1 | ENSG00000118804 | -1.76004 | 0.003161 |
| STEAP3 | ENSG00000115107 | -1.81895 | 0.000108 |
| STOX1 | ENSG00000165730 | -4.49045 | 0.004251 |
| SYT3 | ENSG00000213023 | 2.823687 | 0.044938 |
| SYT6 | ENSG00000134207 | -3.8293 | 0.012582 |
| TBX15 | ENSG00000092607 | -2.11221 | 0.037664 |
| TCN2 | ENSG00000185339 | -1.52927 | 0.008057 |
| TCTEX1D1 | ENSG00000152760 | -2.14543 | 0.004746 |
| TENM4 | ENSG00000149256 | -3.30645 | 0.021035 |
| TGFBI | ENSG00000120708 | -1.54407 | 0.001747 |
| TGM5 | ENSG00000104055 | -3.92204 | 0.008686 |
| THSD7A | ENSG00000005108 | 2.065831 | 0.026499 |
| TLN2 | ENSG00000171914 | -1.51846 | 0.026403 |
| TMEM119 | ENSG00000183160 | 1.528066 | 0.004774 |
| TMEM125 | ENSG00000179178 | 2.518866 | 0.048939 |
| TMEM196 | ENSG00000173452 | -2.86873 | 0.036113 |
| TMEM37 | ENSG00000171227 | -3.20028 | 0.038312 |
| TNFRSF19 | ENSG00000127863 | -4.87455 | 0.004785 |
| TPPP3 | ENSG00000159713 | -2.14289 | 0.007872 |
| TRIM64 | ENSG00000204450 | 3.637029 | 0.009772 |
| TRIM64B | ENSG00000189253 | 3.220045 | 0.014921 |
| TSPAN1 | ENSG00000117472 | -2.80213 | 0.004568 |
| TUBB8P12 | ENSG00000173213 | -2.85541 | 0.032129 |
| UGT1A9 | ENSG00000241119 | -3.40778 | 0.037705 |
| UGT2A3 | ENSG00000135220 | 4.749973 | 0.007091 |
| UGT3A2 | ENSG00000168671 | -4.22709 | 0.01655 |
| UPK3A | ENSG00000100373 | -1.7745 | 0.015043 |
| USP43 | ENSG00000154914 | 3.451056 | 0.018766 |
| UTF1 | ENSG00000171794 | -1.98632 | 0.034041 |
| VWA1 | ENSG00000179403 | -3.34925 | 0.038297 |
| VWA3B | ENSG00000168658 | 5.706282 | 0.005006 |
| VWA5B1 | ENSG00000158816 | 2.909241 | 0.016048 |
| WNT3A | ENSG00000154342 | -2.6569 | 0.031016 |
| ZC2HC1B | ENSG00000118491 | 2.644247 | 0.039815 |
| ZFP57 | ENSG00000204644 | 2.41721 | 0.027614 |
| ZNF705A | ENSG00000196946 | 1.68455 | 0.0098 |

TableS 4 Alternative Splicing Differences between Patients who died from COVID-19 and those who lived. (Coord – location of the event, Type CE=core exon, AA=alternative acceptor splice site, AD=alternative donor splice site, RI=retained intron, AF=alternative first exon, AL=alternative last exon, PSI=percent spliced in, DeltaPSI is the change)

| GENE | ID | Coord | Strand | Type | Psi_Died | Psi_Lived | DeltaPsi |
| --- | --- | --- | --- | --- | --- | --- | --- |
| ABCA13 | ENSG00000179869 | 7:48367794-48367908 | + | CE | 0.87448 | 0.12431 | 0.75017 |
| ABHD14A-ACY1 | ENSG00000114786 | 3:51986505-51986604 | + | RI | 0.66701 | 0.11521 | 0.5518 |
| ANKMY1 | ENSG00000144504 | 2:240509348-240509455 | - | CE | 0.17247 | 0.69743 | -0.52495 |
| ASCC1 | ENSG00000138303 | 10:72102850-72102967 | - | CE | 0.49658 | 0.10866 | 0.38791 |
| C17orf53 | ENSG00000125319 | 17:44154553-44154555 | + | AA | 0.080264 | 0.3502 | -0.26994 |
| CACNA2D4 | ENSG00000151062 | 12:1811662-1811723 | - | CE | 0.60897 | 0.13663 | 0.47234 |
| CATSPER1 | ENSG00000175294 | 11:66020317-66020389 | - | CE | 0.12402 | 0.81171 | -0.68769 |
| CATSPERG | ENSG00000099338 | 19:38344120-38344295 | + | RI | 0.11789 | 0.53877 | -0.42088 |
| CCDC144A | ENSG00000170160 | 17:16737538-16737675 | + | CE | 0.058281 | 0.30288 | -0.2446 |
| CCDC32 | ENSG00000128891 | 15:40564791-40564975 | - | AD | 0.37671 | 0.077779 | 0.29893 |
| CD200 | ENSG00000091972 | 3:112347558-112347562 | + | AA | 0.14924 | 0.69828 | -0.54905 |
| CDC14A | ENSG00000079335 | 1:100485826-100485901 | + | CE | 0.08911 | 0.015364 | 0.073746 |
| CXCL2 | ENSG00000081041 | 4:74098923-74099020 | - | RI | 0.42269 | 0.076886 | 0.3458 |
| CXCR2 | ENSG00000180871 | 2:218130850-218131002 | + | AF | 0.001056 | 0.005115 | -0.00406 |
| DERL3 | ENSG00000099958 | 22:23838138-23838351 | - | AD | 0.83299 | 0.16918 | 0.6638 |
| DET1 | ENSG00000140543 | 15:88536312-88536381 | - | CE | 0.075632 | 0.35264 | -0.27701 |
| DGLUCY | ENSG00000133943 | 14:91223679-91223795 | + | CE | 0.18836 | 0.82648 | -0.63812 |
| DHDDS | ENSG00000117682 | 1:26442708-26442730 | + | AA | 0.10283 | 0.50581 | -0.40298 |
| DNAJA3 | ENSG00000103423 | 16:4454811-4454927 | + | CE | 0.10073 | 0.72284 | -0.62212 |
| DNAJB14 | ENSG00000164031 | 4:99924760-99924815 | - | CE | 0.076563 | 0.41183 | -0.33527 |
| DNAJC1 | ENSG00000136770 | 10:21928506-21928552 | - | CE | 0.90193 | 0.22184 | 0.68009 |
| DOCK9 | ENSG00000088387 | 13:98846520-98846556 | - | CE | 0.59374 | 0.12702 | 0.46672 |
| DPYD | ENSG00000188641 | 1:97719758-97720926 | - | AL | 0.057497 | 0.004676 | 0.052821 |
| DUSP18 | ENSG00000167065 | 22:30664955-30665121 | - | CE | 0.21104 | 0.036503 | 0.17454 |
| ECM1 | ENSG00000143369 | 1:150509835-150509921 | + | AA | 0.12673 | 0.55115 | -0.42442 |
| FAM214B | ENSG00000005238 | 9:35108738-35111268 | - | RI | 0.39404 | 0.079324 | 0.31472 |
| FGF11 | ENSG00000161958 | 17:7442594-7442792 | + | CE | 0.14538 | 0.77019 | -0.6248 |
| FMR1 | ENSG00000102081 | X:147930873-147930964 | + | CE | 0.099738 | 0.92817 | -0.82843 |
| HLA-A | ENSG00000206503 | 6:29944617-29944634 | + | AD | 0.006735 | 0.040571 | -0.03384 |
| HLA-B | ENSG00000234745 | 6:31354527-31354632 | - | RI | 0.003587 | 0.00062 | 0.002967 |
| HLA-C | ENSG00000204525 | 6:31269526-31269543 | - | AA | 0.006439 | 0.060605 | -0.05417 |
| HLA-E | ENSG00000204592 | 6:30489596-30489725 | + | RI | 0.000763 | 0.006458 | -0.0057 |
| IQGAP1 | ENSG00000140575 | 15:90475689-90475754 | + | AL | 0.014793 | 0.00228 | 0.012513 |
| ITGB2 | ENSG00000160255 | 21:44907987-44908182 | - | AL | 0.038378 | 0.002955 | 0.035422 |
| KCTD13 | ENSG00000174943 | 16:29911661-29911814 | - | AD | 0.1156 | 0.54503 | -0.42944 |
| KDM5D | ENSG00000012817 | Y:19715584-19715614 | - | AD | 0.4753 | 0.11799 | 0.35731 |
| KIF9 | ENSG00000088727 | 3:47282140-47282385 | - | CE | 0.75064 | 0.1069 | 0.64374 |
| KLC4 | ENSG00000137171 | 6:43060183-43060238 | + | AD | 0.11397 | 0.63575 | -0.52177 |
| LGALS3BP | ENSG00000108679 | 17:78977001-78977139 | - | RI | 0.10031 | 0.022913 | 0.077393 |
| LRRK2 | ENSG00000188906 | 12:40300813-40301064 | + | CE | 0.066544 | 0.008885 | 0.057659 |
| MBNL1 | ENSG00000152601 | 3:152396167-152396224 | + | CE | 0.001873 | 0.01195 | -0.01008 |
| NCKIPSD | ENSG00000213672 | 3:48682136-48682156 | - | AA | 0.58529 | 0.12479 | 0.4605 |
| NF2 | ENSG00000186575 | 22:29683065-29683079 | + | AD | 0.71689 | 0.090081 | 0.62681 |
| NOS1AP | ENSG00000198929 | 1:162300633-162300647 | + | AA | 0.6301 | 0.12506 | 0.50504 |
| ORC3 | ENSG00000135336 | 6:87656903-87656905 | + | AA | 0.5488 | 0.10473 | 0.44407 |
| PATJ | ENSG00000132849 | 1:62153358-62153481 | + | CE | 0.66332 | 0.14015 | 0.52317 |
| PET100 | ENSG00000229833 | 19:7630660-7630822 | + | RI | 0.86094 | 0.12444 | 0.7365 |
| PKM | ENSG00000067225 | 15:72221168-72221284 | - | CE | 0.001686 | 0.010144 | -0.00846 |
| PLEKHJ1 | ENSG00000104886 | 19:2233906-2233997 | - | RI | 0.11423 | 0.50147 | -0.38724 |
| POGK | ENSG00000143157 | 1:166839605-166839769 | + | AD | 0.8574 | 0.12904 | 0.72835 |
| PRKAA1 | ENSG00000132356 | 5:40774928-40774972 | - | CE | 0.48463 | 0.10106 | 0.38357 |
| RC3H1 | ENSG00000135870 | 1:173938872-173938874 | - | AA | 0.022543 | 0.13881 | -0.11627 |
| RGS2 | ENSG00000116741 | 1:192810268-192810369 | + | RI | 0.043143 | 0.009204 | 0.03394 |
| RHAG | ENSG00000112077 | 6:49606729-49606847 | - | AD | 0.073033 | 0.29339 | -0.22036 |
| RIOK1 | ENSG00000124784 | 6:7395021-7395052 | + | AA | 0.50194 | 0.1235 | 0.37844 |
| SH2D3C | ENSG00000095370 | 9:127766901-127767271 | - | CE | 0.27347 | 0.050322 | 0.22315 |
| SHPRH | ENSG00000146414 | 6:145945638-145945656 | - | AA | 0.26482 | 0.059454 | 0.20536 |
| SLC24A1 | ENSG00000074621 | 15:65638128-65638181 | + | CE | 0.58234 | 0.10983 | 0.47251 |
| SMCHD1 | ENSG00000101596 | 18:2784845-2784987 | + | CE | 0.044217 | 0.004057 | 0.04016 |
| ST7L | ENSG00000007341 | 1:112607241-112607597 | - | AL | 0.15503 | 0.66754 | -0.51251 |
| STRN4 | ENSG00000090372 | 19:46727547-46727567 | - | AA | 0.048641 | 0.47937 | -0.43073 |
| TAF1C | ENSG00000103168 | 16:84180345-84180867 | - | RI | 0.12362 | 0.74059 | -0.61698 |
| TBC1D22A | ENSG00000054611 | 22:46792668-46792804 | + | CE | 0.098992 | 0.467 | -0.368 |
| TCF3 | ENSG00000071564 | 19:1612431-1612433 | - | AA | 0.017846 | 0.092239 | -0.07439 |
| TMC5 | ENSG00000103534 | 16:19417696-19418092 | + | AF | 0.1712 | 0.86162 | -0.69042 |
| TMEM154 | ENSG00000170006 | 4:152646558-152646992 | - | AL | 0.009644 | 0.046208 | -0.03656 |
| TPD52 | ENSG00000076554 | 8:80050445-80050471 | - | CE | 0.058884 | 0.28214 | -0.22326 |
| TREML4 | ENSG00000188056 | 6:41232363-41232504 | + | CE | 0.042848 | 0.35552 | -0.31268 |
| TRIP10 | ENSG00000125733 | 19:6750432-6750511 | + | RI | 0.12277 | 0.77724 | -0.65447 |
| TTC21A | ENSG00000168026 | 3:39131521-39131646 | + | AF | 0.24061 | 0.048599 | 0.19201 |
| TTYH3 | ENSG00000136295 | 7:2659940-2660040 | + | CE | 0.73898 | 0.13377 | 0.60521 |
| TXNDC16 | ENSG00000087301 | 14:52488363-52488486 | - | CE | 0.17447 | 0.91711 | -0.74264 |
| TXNDC16 | ENSG00000087301 | 14:52490391-52490451 | - | CE | 0.17307 | 0.90062 | -0.72755 |
| TXNDC16 | ENSG00000087301 | 14:52457090-52457174 | - | CE | 0.14821 | 0.85911 | -0.71091 |
| TYMP | ENSG00000025708 | 22:50529732-50529754 | - | AA | 0.49139 | 0.087863 | 0.40352 |
| TYMP | ENSG00000025708 | 22:50529720-50529731 | - | AA | 0.52365 | 0.10242 | 0.42123 |
| TYMP | ENSG00000025708 | 22:50529755-50529903 | - | RI | 0.47171 | 0.08006 | 0.39165 |
| UBB | ENSG00000170315 | 17:16381463-16381515 | + | AD | 0.008911 | 0.001867 | 0.007044 |
| USP21 | ENSG00000143258 | 1:161163982-161164163 | + | RI | 0.15082 | 0.8656 | -0.71479 |
| VPS13B | ENSG00000132549 | 8:99209592-99209787 | + | AL | 0.001084 | 0.015372 | -0.01429 |
| VPS13D | ENSG00000048707 | 1:12253722-12253826 | + | CE | 0.87586 | 0.1689 | 0.70697 |
| XYLB | ENSG00000093217 | 3:38374462-38374502 | + | CE | 0.91057 | 0.12671 | 0.78386 |
| ZBTB21 | ENSG00000173276 | 21:42002897-42002961 | - | CE | 0.66967 | 0.1578 | 0.51187 |
| ZDHHC24 | ENSG00000174165 | 11:66529312-66529488 | - | CE | 0.73858 | 0.11964 | 0.61895 |
| ZNF516 | ENSG00000101493 | 18:76490681-76490779 | - | AF | 0.071187 | 0.31533 | -0.24414 |
| ZNF623 | ENSG00000183309 | 8:143636162-143636415 | + | AF | 0.62551 | 0.12997 | 0.49553 |
| ZNF718 | ENSG00000250312 | 4:124674-125296 | + | AD | 0.45759 | 0.099792 | 0.3578 |
| ZSCAN18 | ENSG00000121413 | 19:58097930-58098114 | - | AD | 0.37628 | 0.081224 | 0.29506 |

Table S5 Alternative Transcription Differences between Patients who died from COVID-19 and those who lived. (Coord – location of the event, Type TS=tandem transcription start site, TE=tandem alternative polyadenylation site, PSI=percent spliced in, DeltaPSI is the change)

| Gene | Gene | Coord | Strand | Type | Psi_died | Psi_lived | DeltaPsi |
| --- | --- | --- | --- | --- | --- | --- | --- |
| AADACL2 | ENSG00000197953 | 3:151756992-151757766 | + | TE | 0.12278 | 0.62539 | -0.50261 |
| ABCA5 | ENSG00000154265 | 17:69315361-69315425 | - | TS | 0.008016 | 0.10832 | -0.1003 |
| ABCB1 | ENSG00000085563 | 7:87503859-87503863 | - | TE | 0.095812 | 0.88774 | -0.79193 |
| ABCB1 | ENSG00000085563 | 7:87503633-87503858 | - | TE | 0.67462 | 0.11388 | 0.56074 |
| ABCB11 | ENSG00000073734 | 2:168922982-168923382 | - | TE | 0.06782 | 0.69927 | -0.63145 |
| ABCB4 | ENSG00000005471 | 7:87402059-87402302 | - | TE | 0.11249 | 0.47967 | -0.36718 |
| ABCB7 | ENSG00000131269 | X:75051554-75053279 | - | TE | 0.37066 | 0.033258 | 0.3374 |
| ABCB7 | ENSG00000131269 | X:75053280-75053281 | - | TE | 0.090284 | 0.021849 | 0.068435 |
| ABCC3 | ENSG00000108846 | 17:50691092-50691244 | + | TE | 0.026119 | 0.12761 | -0.10149 |
| ABCD3 | ENSG00000117528 | 1:94518497-94518663 | + | TE | 0.17017 | 0.035033 | 0.13514 |
| ABHD14B | ENSG00000114779 | 3:51968530-51968552 | - | TE | 0.007836 | 0.052145 | -0.04431 |
| ABHD16A | ENSG00000204427 | 6:31702612-31702984 | - | TS | 0.007345 | 0.085321 | -0.07798 |
| ABHD17A | ENSG00000129968 | 19:1876979-1877425 | - | TE | 0.016063 | 0.13396 | -0.1179 |
| ABHD17C | ENSG00000136379 | 15:80755620-80755621 | + | TE | 0.090339 | 0.017776 | 0.072562 |
| ABHD5 | ENSG00000011198 | 3:43718798-43719013 | + | TE | 0.00766 | 0.000403 | 0.007257 |
| ABI2 | ENSG00000138443 | 2:203328391-203328394 | + | TS | 0.12381 | 0.59184 | -0.46803 |
| ABI2 | ENSG00000138443 | 2:203427270-203427528 | + | TE | 0.001711 | 0.047083 | -0.04537 |
| ABL2 | ENSG00000143322 | 1:179107718-179108898 | - | TE | 0.015825 | 0.096193 | -0.08037 |
| ABLIM3 | ENSG00000173210 | 5:149258291-149258413 | + | TE | 0.10103 | 0.46664 | -0.36561 |
| AC244197.3 | ENSG00000241489 | X:149532995-149533935 | - | TS | 0.77103 | 0.11344 | 0.65758 |
| ACADM | ENSG00000117054 | 1:75762692-75762809 | + | TE | 0.57783 | 0.096143 | 0.48168 |
| ACADSB | ENSG00000196177 | 10:123053695-123053878 | + | TE | 0.005154 | 0.087524 | -0.08237 |
| ACE | ENSG00000159640 | 17:63497154-63497438 | + | TE | 0.11803 | 0.48977 | -0.37174 |
| ACLY | ENSG00000131473 | 17:41866908-41866916 | - | TE | 0.041534 | 0.004657 | 0.036877 |
| ACLY | ENSG00000131473 | 17:41867574-41867904 | - | TE | 0.038696 | 0.004687 | 0.034009 |
| ACOT1 | ENSG00000184227 | 14:73543711-73543794 | + | TE | 0.092897 | 0.52687 | -0.43397 |
| ACOT7 | ENSG00000097021 | 1:6264269-6264272 | - | TE | 0.018862 | 0.38481 | -0.36595 |
| ACOT8 | ENSG00000101473 | 20:45857398-45857406 | - | TS | 0.072229 | 0.31109 | -0.23886 |
| ACOX3 | ENSG00000087008 | 4:8366961-8367080 | - | TE | 0.12973 | 0.01743 | 0.1123 |
| ACP1 | ENSG00000143727 | 2:272558-272630 | + | TE | 0.10581 | 0.021769 | 0.084044 |
| ACSL6 | ENSG00000164398 | 5:131953898-131953903 | - | TE | 0.09787 | 0.00831 | 0.08956 |
| ACTL6A | ENSG00000136518 | 3:179588242-179588398 | + | TE | 0.16363 | 0.036114 | 0.12751 |
| ACTN1 | ENSG00000072110 | 14:68874318-68874322 | - | TE | 0.003769 | 0.00062 | 0.003149 |
| ACVR1 | ENSG00000115170 | 2:157736444-157736448 | - | TE | 0.089804 | 0.006743 | 0.083061 |
| ACVR2A | ENSG00000121989 | 2:147927505-147927929 | + | TE | 0.042611 | 0.004758 | 0.037854 |
| ADA | ENSG00000196839 | 20:44619810-44619847 | - | TE | 0.068033 | 0.014288 | 0.053746 |
| ADAM17 | ENSG00000151694 | 2:9555793-9555830 | - | TS | 0.23215 | 0.054246 | 0.17791 |
| ADAM17 | ENSG00000151694 | 2:9489811-9489856 | - | TE | 0.009558 | 0.002022 | 0.007536 |
| ADAM8 | ENSG00000151651 | 10:133276851-133276863 | - | TS | 0.011548 | 0.071882 | -0.06033 |
| ADAM8 | ENSG00000151651 | 10:133262403-133262419 | - | TE | 0.011994 | 0.049379 | -0.03739 |
| ADAM9 | ENSG00000168615 | 8:39105002-39105137 | + | TE | 0.002513 | 0.015088 | -0.01258 |
| ADARB2 | ENSG00000185736 | 10:1183151-1183369 | - | TE | 0.072582 | 0.01463 | 0.057951 |
| ADARB2 | ENSG00000185736 | 10:1183043-1183150 | - | TE | 0.23651 | 0.015301 | 0.22121 |
| ADD1 | ENSG00000087274 | 4:2899436-2899777 | + | TE | 0.042284 | 0.009777 | 0.032507 |
| ADGRD1 | ENSG00000111452 | 12:131139289-131141247 | + | TE | 0.11452 | 0.52477 | -0.41025 |
| ADGRD1 | ENSG00000111452 | 12:131141248-131141460 | + | TE | 0.11711 | 0.47746 | -0.36035 |
| ADGRD1 | ENSG00000111452 | 12:131139264-131139288 | + | TE | 0.89291 | 0.09425 | 0.79866 |
| ADH5 | ENSG00000197894 | 4:99072186-99072441 | - | TE | 0.006467 | 0.032605 | -0.02614 |
| ADHFE1 | ENSG00000147576 | 8:66468808-66468809 | + | TE | 0.033138 | 0.38823 | -0.3551 |
| ADORA2B | ENSG00000170425 | 17:15974679-15975737 | + | TE | 0.1509 | 0.021327 | 0.12957 |
| ADPRH | ENSG00000144843 | 3:119579676-119579851 | + | TS | 0.35129 | 0.07691 | 0.27438 |
| ADSL | ENSG00000239900 | 22:40366615-40366996 | + | TE | 0.1643 | 0.039874 | 0.12442 |
| AES | ENSG00000104964 | 19:3053212-3053548 | - | TE | 0.039651 | 0.005677 | 0.033974 |
| AFTPH | ENSG00000119844 | 2:64592036-64592132 | + | TE | 0.21207 | 0.021191 | 0.19088 |
| AGAP6 | ENSG00000204149 | 10:50010426-50010497 | + | TE | 0.058847 | 0.014275 | 0.044572 |
| AGK | ENSG00000006530 | 7:141654098-141654226 | + | TE | 0.022021 | 0.002106 | 0.019915 |
| AGK | ENSG00000006530 | 7:141654227-141654231 | + | TE | 0.010616 | 0.002085 | 0.008532 |
| AGK | ENSG00000006530 | 7:141653066-141653831 | + | TE | 0.001923 | 0.010835 | -0.00891 |
| AGK | ENSG00000006530 | 7:141652889-141653065 | + | TE | 0.003208 | 0.14273 | -0.13952 |
| AHCYL1 | ENSG00000168710 | 1:110023736-110023741 | + | TE | 0.009375 | 0.15221 | -0.14284 |
| AHCYL2 | ENSG00000158467 | 7:129225050-129225439 | + | TS | 0.17526 | 0.041692 | 0.13357 |
| AHCYL2 | ENSG00000158467 | 7:129427176-129427211 | + | TE | 0.084645 | 0.003531 | 0.081114 |
| AIFM1 | ENSG00000156709 | X:130129365-130129369 | - | TE | 0.11586 | 0.47718 | -0.36132 |
| AIP | ENSG00000110711 | 11:67483041-67483048 | + | TS | 0.092818 | 0.50058 | -0.40776 |
| AK5 | ENSG00000154027 | 1:77558602-77558731 | + | TE | 0.032944 | 0.15145 | -0.11851 |
| AK5 | ENSG00000154027 | 1:77559963-77559969 | + | TE | 0.16682 | 0.03745 | 0.12937 |
| AKAP7 | ENSG00000118507 | 6:131281944-131283532 | + | TE | 0.24028 | 0.020683 | 0.21959 |
| AL590764.2 | ENSG00000285171 | X:71111530-71111571 | - | TS | 0.018165 | 0.081873 | -0.06371 |
| ALCAM | ENSG00000170017 | 3:105575915-105576861 | + | TE | 0.18682 | 0.002971 | 0.18385 |
| ALDH16A1 | ENSG00000161618 | 19:49470589-49470589 | + | TE | 0.041083 | 0.19303 | -0.15194 |
| ALDH3B1 | ENSG00000006534 | 11:68028695-68028696 | + | TE | 0.006576 | 0.074901 | -0.06833 |
| ALG1 | ENSG00000033011 | 16:5085391-5085532 | + | TE | 0.016787 | 0.072242 | -0.05546 |
| ALG2 | ENSG00000119523 | 9:99217367-99217371 | - | TE | 0.005207 | 0.13781 | -0.1326 |
| ALG3 | ENSG00000214160 | 3:184242459-184242676 | - | TE | 0.32674 | 0.05138 | 0.27536 |
| ALKBH4 | ENSG00000160993 | 7:102464864-102464876 | - | TS | 0.096461 | 0.49763 | -0.40117 |
| ALOX5 | ENSG00000012779 | 10:45445508-45445687 | + | TE | 0.02821 | 0.000312 | 0.027898 |
| ALPK1 | ENSG00000073331 | 4:112441203-112441371 | + | TE | 0.026819 | 0.17288 | -0.14607 |
| AMBRA1 | ENSG00000110497 | 11:46591365-46591423 | - | TS | 0.58397 | 0.11029 | 0.47368 |
| AMFR | ENSG00000159461 | 16:56362002-56362022 | - | TE | 0.002529 | 0.01758 | -0.01505 |
| AMIGO2 | ENSG00000139211 | 12:47076498-47076599 | - | TE | 0.003433 | 0.067309 | -0.06388 |
| AMMECR1 | ENSG00000101935 | X:110196372-110196409 | - | TE | 0.069944 | 0.016299 | 0.053646 |
| AMPH | ENSG00000078053 | 7:38384818-38384925 | - | TE | 0.15798 | 0.016218 | 0.14176 |
| AMT | ENSG00000145020 | 3:49416826-49416869 | - | TE | 0.10985 | 0.014881 | 0.094969 |
| ANGPT1 | ENSG00000154188 | 8:107497262-107498010 | - | TS | 0.070122 | 0.28287 | -0.21275 |
| ANGPTL2 | ENSG00000136859 | 9:127122635-127122883 | - | TS | 0.51817 | 0.11097 | 0.4072 |
| ANGPTL2 | ENSG00000136859 | 9:127087349-127089138 | - | TE | 0.30326 | 0.057362 | 0.2459 |
| ANKHD1 | ENSG00000131503 | 5:140539850-140539856 | + | TE | 0.049142 | 0.36808 | -0.31894 |
| ANKMY1 | ENSG00000144504 | 2:240561092-240561110 | - | TS | 0.050693 | 0.32941 | -0.27872 |
| ANKRD11 | ENSG00000167522 | 16:89490245-89490274 | - | TS | 0.15019 | 0.010523 | 0.13967 |
| ANKRD17 | ENSG00000132466 | 4:73075686-73076290 | - | TE | 0.004589 | 0.036466 | -0.03188 |
| ANKRD28 | ENSG00000206560 | 3:15667239-15668305 | - | TE | 0.28981 | 0.023874 | 0.26593 |
| ANKRD36 | ENSG00000135976 | 2:97113552-97113936 | + | TS | 0.047349 | 0.25191 | -0.20456 |
| ANKRD37 | ENSG00000186352 | 4:185400017-185400241 | + | TE | 0.26288 | 0.055355 | 0.20752 |
| ANKRD49 | ENSG00000168876 | 11:94498071-94498263 | + | TE | 0.0036 | 0.030246 | -0.02665 |
| ANKRD6 | ENSG00000135299 | 6:89631337-89631477 | + | TE | 0.083299 | 0.006595 | 0.076704 |
| ANKRD6 | ENSG00000135299 | 6:89630433-89631336 | + | TE | 0.005939 | 0.12616 | -0.12022 |
| ANKS1A | ENSG00000064999 | 6:35088606-35088963 | + | TE | 0.003195 | 0.03971 | -0.03652 |
| ANKS3 | ENSG00000168096 | 16:4700171-4700303 | - | TS | 0.41934 | 0.097608 | 0.32173 |
| ANXA2 | ENSG00000182718 | 15:60347327-60347331 | - | TE | 0.001517 | 0.012859 | -0.01134 |
| ANXA4 | ENSG00000196975 | 2:69825782-69825783 | + | TE | 0.057027 | 0.006191 | 0.050836 |
| AP1AR | ENSG00000138660 | 4:112231889-112232174 | + | TS | 0.53784 | 0.11085 | 0.42699 |
| AP3D1 | ENSG00000065000 | 19:2100989-2100993 | - | TE | 0.11794 | 0.004879 | 0.11306 |
| AP3D1 | ENSG00000065000 | 19:2100988-2100988 | - | TE | 0.033277 | 0.004779 | 0.028498 |
| AP3S2 | ENSG00000157823 | 15:89893881-89893977 | - | TS | 0.27239 | 0.057141 | 0.21525 |
| AP3S2 | ENSG00000157823 | 15:89893978-89894028 | - | TS | 0.22063 | 0.037078 | 0.18356 |
| AP4E1 | ENSG00000081014 | 15:51005895-51005900 | + | TE | 0.039601 | 0.25848 | -0.21888 |
| AP4E1 | ENSG00000081014 | 15:51002502-51003217 | + | TE | 0.005501 | 0.027946 | -0.02244 |
| AP4M1 | ENSG00000221838 | 7:100107149-100107179 | + | TE | 0.069144 | 0.015388 | 0.053756 |
| AP5M1 | ENSG00000053770 | 14:57288802-57288925 | + | TE | 0.11339 | 0.52687 | -0.41348 |
| AP5Z1 | ENSG00000242802 | 7:4791115-4791518 | + | TE | 0.007921 | 0.23491 | -0.22699 |
| APBB2 | ENSG00000163697 | 4:40814607-40815323 | - | TE | 0.007082 | 0.070452 | -0.06337 |
| APBB3 | ENSG00000113108 | 5:140564372-140564374 | - | TS | 0.10271 | 0.50818 | -0.40547 |
| APC2 | ENSG00000115266 | 19:1465410-1465418 | + | TE | 0.003969 | 0.11103 | -0.10706 |
| APH1A | ENSG00000117362 | 1:150265404-150265405 | - | TE | 0.014845 | 0.13655 | -0.12171 |
| API5 | ENSG00000166181 | 11:43342433-43342664 | + | TE | 0.001153 | 0.035699 | -0.03455 |
| APOL2 | ENSG00000128335 | 22:36226210-36227341 | - | TE | 0.012291 | 0.090117 | -0.07783 |
| APOPT1 | ENSG00000256053 | 14:103590181-103590504 | + | TE | 0.033444 | 0.25798 | -0.22454 |
| APPL2 | ENSG00000136044 | 12:105173935-105174016 | - | TE | 0.034781 | 0.001086 | 0.033695 |
| APTX | ENSG00000137074 | 9:32972749-32972762 | - | TE | 0.14893 | 0.02315 | 0.12578 |
| AQP3 | ENSG00000165272 | 9:33447597-33447633 | - | TS | 0.49597 | 0.047658 | 0.44832 |
| ARGLU1 | ENSG00000134884 | 13:106567573-106567623 | - | TS | 0.00665 | 0.03533 | -0.02868 |
| ARHGAP11A | ENSG00000198826 | 15:32637904-32638110 | + | TE | 0.11295 | 0.013199 | 0.099753 |
| ARHGAP9 | ENSG00000123329 | 12:57488727-57488814 | - | TS | 0.014039 | 0.082105 | -0.06807 |
| ARHGEF2 | ENSG00000116584 | 1:155946854-155947941 | - | TE | 0.06404 | 0.005512 | 0.058528 |
| ARHGEF40 | ENSG00000165801 | 14:21089014-21089414 | + | TE | 0.001717 | 0.062586 | -0.06087 |
| ARID1B | ENSG00000049618 | 6:157206167-157208361 | + | TE | 0.000873 | 0.005402 | -0.00453 |
| ARID3B | ENSG00000179361 | 15:74595611-74598129 | + | TE | 0.012223 | 0.002676 | 0.009547 |
| ARID4B | ENSG00000054267 | 1:235167884-235168543 | - | TE | 0.002848 | 0.028559 | -0.02571 |
| ARL4A | ENSG00000122644 | 7:12688169-12688406 | + | TE | 0.031589 | 0.18429 | -0.1527 |
| ARMC6 | ENSG00000105676 | 19:19033578-19033612 | + | TS | 0.35261 | 0.083951 | 0.26866 |
| ARMC8 | ENSG00000114098 | 3:138298374-138298384 | + | TE | 0.003969 | 0.35267 | -0.3487 |
| ARMC8 | ENSG00000114098 | 3:138295934-138296254 | + | TE | 0.002993 | 0.01375 | -0.01076 |
| ARMCX3 | ENSG00000102401 | X:101626485-101627841 | + | TE | 0.002371 | 0.04838 | -0.04601 |
| ARMCX5 | ENSG00000125962 | X:102604154-102604159 | + | TE | 0.30334 | 0.025027 | 0.27831 |
| ARMCX6 | ENSG00000198960 | X:101615118-101615119 | - | TE | 0.29171 | 0.018204 | 0.27351 |
| ARMCX6 | ENSG00000198960 | X:101615120-101615125 | - | TE | 0.018249 | 0.2585 | -0.24025 |
| ARNT | ENSG00000143437 | 1:150810859-150811680 | - | TE | 0.001046 | 0.023174 | -0.02213 |
| ARNT | ENSG00000143437 | 1:150811961-150812110 | - | TE | 0.001083 | 0.055071 | -0.05399 |
| ARRDC2 | ENSG00000105643 | 19:18012913-18014100 | + | TE | 0.012199 | 0.35616 | -0.34396 |
| ASCC2 | ENSG00000100325 | 22:29788609-29788610 | - | TE | 0.00991 | 0.1277 | -0.11779 |
| ASIC1 | ENSG00000110881 | 12:50083278-50083611 | + | TE | 0.14247 | 0.62831 | -0.48584 |
| ASMTL | ENSG00000169093 | X:1403139-1403140 | - | TE | 0.19183 | 0.039148 | 0.15268 |
| ASPH | ENSG00000198363 | 8:61624539-61624602 | - | TE | 0.0504 | 0.009539 | 0.040861 |
| ATF3 | ENSG00000162772 | 1:212619367-212619535 | + | TE | 0.034174 | 0.35879 | -0.32462 |
| ATF7IP2 | ENSG00000166669 | 16:10387378-10387737 | + | TE | 0.052099 | 0.50052 | -0.44842 |
| ATG16L1 | ENSG00000085978 | 2:233251682-233251684 | + | TS | 0.48983 | 0.07544 | 0.41439 |
| ATG16L1 | ENSG00000085978 | 2:233251701-233251705 | + | TS | 0.11399 | 0.48725 | -0.37326 |
| ATG16L1 | ENSG00000085978 | 2:233251673-233251681 | + | TS | 0.31643 | 0.077809 | 0.23862 |
| ATG4B | ENSG00000168397 | 2:241673850-241673854 | + | TE | 0.13939 | 0.010902 | 0.12849 |
| ATL1 | ENSG00000198513 | 14:50633050-50633068 | + | TE | 0.070869 | 0.36479 | -0.29392 |
| ATP2C2 | ENSG00000064270 | 16:84463614-84464000 | + | TE | 0.12273 | 0.62215 | -0.49941 |
| ATP2C2 | ENSG00000064270 | 16:84464180-84464187 | + | TE | 0.61138 | 0.1341 | 0.47728 |
| ATP5F1D | ENSG00000099624 | 19:1241768-1241769 | + | TS | 0.11541 | 0.006969 | 0.10845 |
| ATP5MC3 | ENSG00000154518 | 2:175178186-175178402 | - | TE | 0.085097 | 0.011645 | 0.073451 |
| ATP5MC3 | ENSG00000154518 | 2:175181664-175181695 | - | TS | 0.079274 | 0.4785 | -0.39922 |
| ATP6AP2 | ENSG00000182220 | X:40592471-40592502 | + | TE | 0.023688 | 0.17756 | -0.15387 |
| ATP6V1A | ENSG00000114573 | 3:113809335-113809697 | + | TE | 0.000434 | 0.011868 | -0.01143 |
| ATP8B3 | ENSG00000130270 | 19:1798761-1798763 | - | TE | 0.067949 | 0.28721 | -0.21926 |
| ATPAF1 | ENSG00000123472 | 1:46635041-46635044 | - | TE | 0.016798 | 0.002786 | 0.014013 |
| ATXN1 | ENSG00000124788 | 6:16614975-16614978 | - | TE | 0.12176 | 0.003448 | 0.11831 |
| ATXN2L | ENSG00000168488 | 16:28836751-28836919 | + | TE | 0.028128 | 0.00601 | 0.022118 |
| ATXN2L | ENSG00000168488 | 16:28837216-28837221 | + | TE | 0.2816 | 0.029325 | 0.25228 |
| ATXN7 | ENSG00000285258 | 3:63864557-63864559 | + | TS | 0.29478 | 0.068112 | 0.22667 |
| AUTS2 | ENSG00000158321 | 7:70789748-70790013 | + | TE | 0.087334 | 0.017973 | 0.069361 |
| AXIN2 | ENSG00000168646 | 17:65558046-65558729 | - | TE | 0.15947 | 0.032009 | 0.12747 |
| B3GALT4 | ENSG00000235863 | 6:33277132-33278687 | + | TS | 0.13934 | 0.006269 | 0.13307 |
| B4GALNT3 | ENSG00000139044 | 12:563479-563509 | + | TE | 0.087738 | 0.015611 | 0.072127 |
| B4GALT3 | ENSG00000158850 | 1:161177423-161177468 | - | TS | 0.084083 | 0.64765 | -0.56356 |
| BABAM2 | ENSG00000158019 | 2:27890792-27890792 | + | TS | 0.50054 | 0.07448 | 0.42606 |
| BACE1 | ENSG00000186318 | 11:117289535-117289807 | - | TE | 0.004788 | 0.029735 | -0.02495 |
| BAG1 | ENSG00000107262 | 9:33255049-33255218 | - | TE | 0.10526 | 0.011574 | 0.093687 |
| BAIAP3 | ENSG00000007516 | 16:1348641-1348755 | + | TE | 0.076004 | 0.34852 | -0.27252 |
| BARD1 | ENSG00000138376 | 2:214809684-214809704 | - | TS | 0.60613 | 0.11693 | 0.4892 |
| BAZ2A | ENSG00000076108 | 12:56598359-56598368 | - | TE | 0.001213 | 0.006688 | -0.00547 |
| BBS1 | ENSG00000174483 | 11:66533614-66533627 | + | TE | 0.011376 | 0.082932 | -0.07156 |
| BBS10 | ENSG00000179941 | 12:76344474-76344505 | - | TE | 0.096141 | 0.001845 | 0.094296 |
| BCAS4 | ENSG00000124243 | 20:50876573-50876723 | + | TE | 0.00732 | 0.088142 | -0.08082 |
| BCL11A | ENSG00000119866 | 2:60450520-60451145 | - | TE | 0.35161 | 0.054592 | 0.29702 |
| BCL2L11 | ENSG00000153094 | 2:111164133-111164231 | + | TE | 0.009976 | 0.19845 | -0.18848 |
| BCL6 | ENSG00000113916 | 3:187722398-187722431 | - | TE | 0.010492 | 0.13439 | -0.12389 |
| BCL7B | ENSG00000106635 | 7:73557610-73557671 | - | TS | 0.13754 | 0.83973 | -0.70219 |
| BCL7B | ENSG00000106635 | 7:73557487-73557609 | - | TS | 0.86239 | 0.16171 | 0.70068 |
| BCO2 | ENSG00000197580 | 11:112218142-112218157 | + | TE | 0.87879 | 0.1431 | 0.73569 |
| BCO2 | ENSG00000197580 | 11:112217991-112218117 | + | TE | 0.092479 | 0.74459 | -0.65212 |
| BCOR | ENSG00000183337 | X:40051248-40051254 | - | TE | 0.14349 | 0.013055 | 0.13043 |
| BECN1 | ENSG00000126581 | 17:42810780-42810928 | - | TE | 0.12951 | 0.008981 | 0.12053 |
| BET1 | ENSG00000105829 | 7:93993242-93993259 | - | TE | 0.017404 | 0.07803 | -0.06063 |
| BEX2 | ENSG00000133134 | X:103309346-103309353 | - | TE | 0.529 | 0.10349 | 0.42551 |
| BEX3 | ENSG00000166681 | X:103378061-103378071 | + | TE | 0.25326 | 0.03168 | 0.22158 |
| BICD1 | ENSG00000151746 | 12:32377878-32377923 | + | TE | 0.12488 | 0.023975 | 0.10091 |
| BIN1 | ENSG00000136717 | 2:127107279-127107288 | - | TS | 0.069733 | 0.40509 | -0.33536 |
| BIRC2 | ENSG00000110330 | 11:102377990-102378236 | + | TE | 0.002742 | 0.023244 | -0.0205 |
| BLOC1S6 | ENSG00000104164 | 15:45606395-45606585 | + | TE | 0.001168 | 0.035492 | -0.03432 |
| BLOC1S6 | ENSG00000104164 | 15:45606586-45606607 | + | TE | 0.01917 | 0.078008 | -0.05884 |
| BNIPL | ENSG00000163141 | 1:151036321-151036569 | + | TS | 0.031511 | 0.15146 | -0.11995 |
| BOLA2B | ENSG00000169627 | 16:30193698-30194306 | - | TS | 0.090221 | 0.47044 | -0.38022 |
| BOLA2-SMG1P6 | ENSG00000261740 | 16:29443230-29443248 | - | TE | 0.066296 | 0.012791 | 0.053505 |
| BPHL | ENSG00000137274 | 6:3153196-3153578 | + | TE | 0.090649 | 0.39881 | -0.30816 |
| BRAF | ENSG00000157764 | 7:140732563-140734313 | - | TE | 0.36858 | 0.070311 | 0.29826 |
| BRAF | ENSG00000157764 | 7:140730665-140732562 | - | TE | 0.095206 | 0.018593 | 0.076613 |
| BRAT1 | ENSG00000106009 | 7:2537881-2538764 | - | TE | 0.28832 | 0.011402 | 0.27692 |
| BRD8 | ENSG00000112983 | 5:138156882-138156914 | - | TE | 0.35198 | 0.077635 | 0.27434 |
| BRF1 | ENSG00000185024 | 14:105247194-105247894 | - | TS | 0.038131 | 0.37839 | -0.34026 |
| BRF1 | ENSG00000185024 | 14:105210367-105210588 | - | TE | 0.10065 | 0.5546 | -0.45395 |
| BRPF1 | ENSG00000156983 | 3:9748002-9748015 | + | TE | 0.01615 | 0.075067 | -0.05892 |
| BRWD1 | ENSG00000185658 | 21:39187246-39187417 | - | TE | 0.011506 | 0.002279 | 0.009227 |
| BSG | ENSG00000172270 | 19:583488-583492 | + | TE | 0.040665 | 0.000895 | 0.03977 |
| BSG | ENSG00000172270 | 19:583493-583493 | + | TE | 0.004403 | 0.000905 | 0.003498 |
| BST2 | ENSG00000130303 | 19:17403241-17403326 | - | TE | 0.032579 | 0.1396 | -0.10702 |
| BTLA | ENSG00000186265 | 3:112466108-112466383 | - | TE | 0.023738 | 0.14326 | -0.11952 |
| BTNL8 | ENSG00000113303 | 5:180950662-180950904 | + | TE | 0.002439 | 0.025579 | -0.02314 |
| BUB1 | ENSG00000169679 | 2:110637832-110637833 | - | TE | 0.11251 | 0.54516 | -0.43266 |
| C11orf21 | ENSG00000110665 | 11:2297173-2297921 | - | TE | 0.000958 | 0.023584 | -0.02263 |
| C11orf58 | ENSG00000110696 | 11:16748158-16748196 | + | TE | 0.4989 | 0.12131 | 0.37759 |
| C12orf10 | ENSG00000139637 | 12:53299718-53299749 | + | TS | 0.17114 | 0.036644 | 0.13449 |
| C12orf29 | ENSG00000133641 | 12:88048234-88048423 | + | TE | 0.28685 | 0.057698 | 0.22916 |
| C15orf61 | ENSG00000189227 | 15:67526418-67526541 | + | TE | 0.26288 | 0.019621 | 0.24326 |
| C16orf58 | ENSG00000140688 | 16:31489494-31489497 | - | TE | 0.15246 | 0.012055 | 0.14041 |
| C17orf58 | ENSG00000186665 | 17:67991336-67992103 | - | TE | 0.023737 | 0.14631 | -0.12258 |
| C17orf80 | ENSG00000141219 | 17:73247528-73247530 | + | TE | 0.005207 | 0.035955 | -0.03075 |
| C19orf44 | ENSG00000105072 | 19:16520094-16521350 | + | TE | 0.15837 | 0.037803 | 0.12056 |
| C19orf48 | ENSG00000167747 | 19:50798516-50798910 | - | TE | 0.092495 | 0.016997 | 0.075498 |
| C1orf112 | ENSG00000000460 | 1:169852790-169853037 | + | TE | 0.021605 | 0.005274 | 0.016331 |
| C1orf122 | ENSG00000197982 | 1:37809450-37809452 | + | TE | 0.11452 | 0.010648 | 0.10387 |
| C1orf43 | ENSG00000143612 | 1:154206706-154206719 | - | TE | 0.00943 | 0.001495 | 0.007934 |
| C1QTNF6 | ENSG00000133466 | 22:37180167-37180170 | - | TE | 0.3057 | 0.031823 | 0.27388 |
| C2 | ENSG00000166278 | 6:31945462-31945521 | + | TE | 0.081387 | 0.01807 | 0.063316 |
| C2 | ENSG00000166278 | 6:31945672-31945672 | + | TE | 0.079425 | 0.017583 | 0.061842 |
| C20orf204 | ENSG00000196421 | 20:64039779-64039962 | + | TE | 0.5752 | 0.12483 | 0.45038 |
| C22orf23 | ENSG00000128346 | 22:37943526-37944246 | - | TE | 0.032692 | 0.25155 | -0.21886 |
| C22orf39 | ENSG00000242259 | 22:19440908-19441694 | - | TE | 0.1307 | 0.008946 | 0.12175 |
| C8orf82 | ENSG00000213563 | 8:144526219-144527323 | - | TE | 0.010983 | 0.04453 | -0.03355 |
| C9orf72 | ENSG00000147894 | 9:27547108-27548422 | - | TE | 0.0358 | 0.005477 | 0.030324 |
| CADM1 | ENSG00000182985 | 11:115174307-115175340 | - | TE | 0.21243 | 0.040319 | 0.17211 |
| CALM1 | ENSG00000198668 | 14:90405073-90405107 | + | TE | 0.003639 | 0.019154 | -0.01552 |
| CALM3 | ENSG00000160014 | 19:46609306-46609770 | + | TE | 0.00477 | 0.000594 | 0.004176 |
| CALM3 | ENSG00000160014 | 19:46609125-46609234 | + | TE | 0.000525 | 0.069568 | -0.06904 |
| CALML4 | ENSG00000129007 | 15:68205299-68205488 | - | TS | 0.014333 | 0.1068 | -0.09247 |
| CAMK2B | ENSG00000058404 | 7:44217150-44217155 | - | TE | 0.90955 | 0.13883 | 0.77072 |
| CAMK2B | ENSG00000058404 | 7:44217156-44217558 | - | TE | 0.091524 | 0.85189 | -0.76036 |
| CAMTA1 | ENSG00000171735 | 1:6785454-6785463 | + | TS | 0.07316 | 0.3128 | -0.23964 |
| CAMTA1 | ENSG00000171735 | 1:6785464-6785479 | + | TS | 0.071475 | 0.31812 | -0.24665 |
| CAMTA2 | ENSG00000108509 | 17:4987113-4987370 | - | TS | 0.031318 | 0.13103 | -0.09971 |
| CAPN1 | ENSG00000014216 | 11:65208478-65209143 | + | TS | 0.004521 | 0.040104 | -0.03558 |
| CAPN12 | ENSG00000182472 | 19:38730534-38730558 | - | TE | 0.035145 | 0.22333 | -0.18819 |
| CAPN5 | ENSG00000149260 | 11:77123688-77124054 | + | TE | 0.16091 | 0.022847 | 0.13806 |
| CAPRIN1 | ENSG00000135387 | 11:34099515-34100464 | + | TE | 0.000415 | 0.00816 | -0.00775 |
| CAPRIN2 | ENSG00000110888 | 12:30710340-30710470 | - | TE | 0.018196 | 0.12481 | -0.10662 |
| CARD8 | ENSG00000105483 | 19:48211377-48211616 | - | TE | 0.000173 | 0.024473 | -0.0243 |
| CASK | ENSG00000147044 | X:41519211-41519220 | - | TE | 0.031082 | 0.005191 | 0.025891 |
| CASTOR3 | ENSG00000239521 | 7:100200653-100200659 | - | TE | 0.018848 | 0.13562 | -0.11678 |
| CASTOR3 | ENSG00000239521 | 7:100202241-100202603 | - | TE | 0.094102 | 0.010674 | 0.083427 |
| CAT | ENSG00000121691 | 11:34471548-34472058 | + | TE | 0.094323 | 0.41533 | -0.321 |
| CBWD1 | ENSG00000172785 | 9:121409-121409 | - | TE | 0.081296 | 0.88595 | -0.80466 |
| CBWD1 | ENSG00000172785 | 9:121038-121040 | - | TE | 0.91575 | 0.083007 | 0.83274 |
| CCDC148 | ENSG00000153237 | 2:158171974-158172259 | - | TE | 0.1322 | 0.73693 | -0.60473 |
| CCDC24 | ENSG00000159214 | 1:43996525-43996528 | + | TE | 0.059815 | 0.29059 | -0.23078 |
| CCDC25 | ENSG00000147419 | 8:27733311-27733317 | - | TE | 0.011414 | 0.055523 | -0.04411 |
| CCDC28A | ENSG00000024862 | 6:138793313-138793317 | + | TE | 0.018833 | 0.26164 | -0.24281 |
| CCDC30 | ENSG00000186409 | 1:42473418-42473560 | + | TE | 0.5072 | 0.12305 | 0.38415 |
| CCDC32 | ENSG00000128891 | 15:40553099-40553103 | - | TE | 0.11704 | 0.00959 | 0.10745 |
| CCDC88A | ENSG00000115355 | 2:55419316-55419363 | - | TS | 0.086562 | 0.014056 | 0.072507 |
| CCDC88A | ENSG00000115355 | 2:55419644-55419661 | - | TS | 0.16817 | 0.039792 | 0.12838 |
| CCDC91 | ENSG00000123106 | 12:28549192-28549194 | + | TE | 0.012407 | 0.070299 | -0.05789 |
| CCL2 | ENSG00000108691 | 17:34255285-34255425 | + | TS | 0.13119 | 0.027914 | 0.10327 |
| CCM2 | ENSG00000136280 | 7:45027026-45027633 | + | TS | 0.52301 | 0.12648 | 0.39653 |
| CCND3 | ENSG00000112576 | 6:41934985-41936107 | - | TE | 0.038676 | 0.005591 | 0.033085 |
| CCNG1 | ENSG00000113328 | 5:163445015-163445016 | + | TE | 0.52428 | 0.1061 | 0.41818 |
| CCNJL | ENSG00000135083 | 5:160312578-160312600 | - | TS | 0.60811 | 0.1292 | 0.47891 |
| CCNL1 | ENSG00000163660 | 3:157160214-157160760 | - | TS | 0.27549 | 0.040324 | 0.23517 |
| CCNL1 | ENSG00000163660 | 3:157160111-157160137 | - | TS | 0.018667 | 0.077012 | -0.05835 |
| CCNL2 | ENSG00000221978 | 1:1399313-1399317 | - | TS | 0.032666 | 0.15141 | -0.11874 |
| CCNL2 | ENSG00000221978 | 1:1386482-1386494 | - | TE | 0.012344 | 0.062552 | -0.05021 |
| CCNL2 | ENSG00000221978 | 1:1386708-1386712 | - | TE | 0.012301 | 0.089349 | -0.07705 |
| CCNT1 | ENSG00000129315 | 12:48693930-48694436 | - | TE | 0.000595 | 0.005079 | -0.00448 |
| CCNT2 | ENSG00000082258 | 2:134954800-134956843 | + | TE | 0.000793 | 0.041645 | -0.04085 |
| CCP110 | ENSG00000103540 | 16:19551196-19551477 | + | TE | 0.075912 | 0.002697 | 0.073216 |
| CCR3 | ENSG00000183625 | 3:46266621-46266706 | + | TE | 0.024377 | 0.16166 | -0.13728 |
| CCSER2 | ENSG00000107771 | 10:84513449-84513870 | + | TE | 0.009926 | 0.000805 | 0.009121 |
| CCT5 | ENSG00000150753 | 5:10264656-10264902 | + | TE | 0.18298 | 0.01783 | 0.16515 |
| CCT6A | ENSG00000146731 | 7:56063013-56063178 | + | TE | 0.051892 | 0.006888 | 0.045003 |
| CD14 | ENSG00000170458 | 5:140633652-140633701 | - | TS | 0.023975 | 0.20709 | -0.18311 |
| CD163 | ENSG00000177575 | 12:7471176-7471179 | - | TE | 0.074535 | 0.40561 | -0.33108 |
| CD22 | ENSG00000012124 | 19:35346566-35346766 | + | TE | 0.079863 | 0.005567 | 0.074296 |
| CD226 | ENSG00000150637 | 18:69864314-69864439 | - | TE | 0.025295 | 0.29419 | -0.2689 |
| CD33 | ENSG00000105383 | 19:51235823-51235966 | + | TE | 0.029948 | 0.12383 | -0.09388 |
| CD36 | ENSG00000135218 | 7:80674148-80674409 | + | TE | 0.35599 | 0.043816 | 0.31217 |
| CD3G | ENSG00000160654 | 11:118353119-118353837 | + | TE | 0.009394 | 0.001239 | 0.008155 |
| CD86 | ENSG00000114013 | 3:122119438-122119820 | + | TE | 0.042733 | 0.009642 | 0.033091 |
| CD9 | ENSG00000010278 | 12:6238135-6238139 | + | TE | 0.2997 | 0.021497 | 0.27821 |
| CD99L2 | ENSG00000102181 | X:150768729-150769101 | - | TE | 0.025316 | 0.002361 | 0.022956 |
| CD99L2 | ENSG00000102181 | X:150768367-150768728 | - | TE | 0.020387 | 0.00234 | 0.018047 |
| CDC123 | ENSG00000151465 | 10:12195966-12196195 | + | TS | 0.5169 | 0.11011 | 0.4068 |
| CDCA3 | ENSG00000111665 | 12:6844799-6844858 | - | TE | 0.093592 | 0.53553 | -0.44194 |
| CDK10 | ENSG00000185324 | 16:89696355-89696364 | + | TE | 0.28663 | 0.02745 | 0.25918 |
| CDK12 | ENSG00000167258 | 17:39531845-39532477 | + | TE | 0.000574 | 0.002605 | -0.00203 |
| CDK13 | ENSG00000065883 | 7:40095463-40095482 | + | TE | 0.001835 | 0.064392 | -0.06256 |
| CDK16 | ENSG00000102225 | X:47223009-47223017 | + | TS | 0.5149 | 0.10951 | 0.40539 |
| CDK2AP2 | ENSG00000167797 | 11:67508001-67508076 | - | TS | 0.1627 | 0.025546 | 0.13716 |
| CDK2AP2 | ENSG00000167797 | 11:67508077-67508091 | - | TS | 0.15988 | 0.029126 | 0.13075 |
| CDK5RAP2 | ENSG00000136861 | 9:120389196-120389198 | - | TE | 0.025356 | 0.20673 | -0.18138 |
| CDK5RAP2 | ENSG00000136861 | 9:120389199-120389200 | - | TE | 0.025099 | 0.14968 | -0.12458 |
| CDK5RAP2 | ENSG00000136861 | 9:120388876-120389194 | - | TE | 0.025404 | 0.14186 | -0.11646 |
| CDK6 | ENSG00000105810 | 7:92614714-92615139 | - | TE | 0.022051 | 0.003495 | 0.018556 |
| CDK8 | ENSG00000132964 | 13:26254139-26254769 | + | TS | 0.16098 | 0.033803 | 0.12717 |
| CDKN2C | ENSG00000123080 | 1:50973893-50974558 | + | TE | 0.011048 | 0.152 | -0.14095 |
| CDYL | ENSG00000153046 | 6:4955548-4955551 | + | TE | 0.30147 | 0.024677 | 0.27679 |
| CEBPZ | ENSG00000115816 | 2:37201784-37201903 | - | TE | 0.076019 | 0.36965 | -0.29363 |
| CEP164 | ENSG00000110274 | 11:117351790-117351942 | + | TE | 0.091108 | 0.6279 | -0.53679 |
| CEP41 | ENSG00000106477 | 7:130398842-130399039 | - | TE | 0.058336 | 0.010938 | 0.047397 |
| CEP78 | ENSG00000148019 | 9:78236110-78236206 | + | TS | 0.044231 | 0.29227 | -0.24804 |
| CEP95 | ENSG00000258890 | 17:64537947-64537948 | + | TE | 0.040253 | 0.27759 | -0.23734 |
| CEPT1 | ENSG00000134255 | 1:111184809-111185100 | + | TE | 0.001762 | 0.008634 | -0.00687 |
| CERS1 | ENSG00000223802 | 19:18868552-18869390 | - | TE | 0.34493 | 0.07667 | 0.26826 |
| CERS3 | ENSG00000154227 | 15:100402511-100402865 | - | TE | 0.080537 | 0.38351 | -0.30297 |
| CES2 | ENSG00000172831 | 16:66943971-66944306 | + | TE | 0.03938 | 0.006756 | 0.032624 |
| CES4A | ENSG00000172824 | 16:67009736-67009752 | + | TE | 0.06114 | 0.27095 | -0.20981 |
| CFAP410 | ENSG00000160226 | 21:44339118-44339393 | - | TS | 0.12138 | 0.50934 | -0.38796 |
| CFAP77 | ENSG00000188523 | 9:132573250-132573317 | + | TE | 0.14261 | 0.032744 | 0.10986 |
| CFL2 | ENSG00000165410 | 14:34712008-34712036 | - | TE | 0.10036 | 0.006373 | 0.09399 |
| CFLAR | ENSG00000003402 | 2:201163974-201164110 | + | TE | 0.000611 | 4.06E-05 | 0.00057 |
| CHCHD1 | ENSG00000172586 | 10:73783074-73783371 | + | TE | 0.15686 | 0.035716 | 0.12115 |
| CHCHD3 | ENSG00000106554 | 7:132785525-132785660 | - | TE | 0.11888 | 0.008053 | 0.11082 |
| CHCHD4 | ENSG00000163528 | 3:14112077-14112092 | - | TE | 0.022994 | 0.45277 | -0.42978 |
| CHCHD7 | ENSG00000170791 | 8:56217331-56217465 | + | TE | 0.16874 | 0.00956 | 0.15918 |
| CHD4 | ENSG00000111642 | 12:6570082-6570086 | - | TE | 0.22305 | 0.033592 | 0.18945 |
| CHD4 | ENSG00000111642 | 12:6570580-6570689 | - | TE | 0.005354 | 0.18295 | -0.1776 |
| CHD4 | ENSG00000111642 | 12:6606710-6606993 | - | TS | 0.059156 | 0.26816 | -0.209 |
| CHD4 | ENSG00000111642 | 12:6570158-6570170 | - | TE | 0.058537 | 0.008111 | 0.050426 |
| CHD4 | ENSG00000111642 | 12:6570184-6570185 | - | TE | 0.005743 | 0.048495 | -0.04275 |
| CHD7 | ENSG00000171316 | 8:60865016-60865125 | + | TE | 0.007703 | 0.06461 | -0.05691 |
| CHFR | ENSG00000072609 | 12:132840975-132841596 | - | TE | 0.009263 | 0.000877 | 0.008386 |
| CHFR | ENSG00000072609 | 12:132840910-132840974 | - | TE | 0.016076 | 0.000859 | 0.015217 |
| CHKA | ENSG00000110721 | 11:68053512-68053819 | - | TE | 0.027246 | 0.005629 | 0.021617 |
| CHMP7 | ENSG00000147457 | 8:23260588-23261992 | + | TE | 0.000925 | 0.031255 | -0.03033 |
| CHRNA7 | ENSG00000175344 | 15:32172991-32173018 | + | TE | 0.11452 | 0.022252 | 0.092273 |
| CHSY1 | ENSG00000131873 | 15:101177101-101178980 | - | TE | 0.000396 | 0.009253 | -0.00886 |
| CIAPIN1 | ENSG00000005194 | 16:57428169-57428186 | - | TE | 0.002847 | 0.020519 | -0.01767 |
| CIART | ENSG00000159208 | 1:150286738-150286776 | + | TE | 0.087523 | 0.91775 | -0.83023 |
| CIART | ENSG00000159208 | 1:150286430-150286737 | + | TE | 0.91893 | 0.085814 | 0.83312 |
| CITED2 | ENSG00000164442 | 6:139373074-139373297 | - | TE | 0.033288 | 0.00271 | 0.030578 |
| CKLF | ENSG00000217555 | 16:66552573-66552793 | + | TS | 0.009236 | 0.002043 | 0.007193 |
| CLDN11 | ENSG00000013297 | 3:170434093-170434099 | + | TE | 0.44461 | 0.09555 | 0.34906 |
| CLDN12 | ENSG00000157224 | 7:90412644-90413411 | + | TE | 0.10752 | 0.016979 | 0.090543 |
| CLEC12B | ENSG00000256660 | 12:10010632-10010758 | + | TS | 0.36811 | 0.069183 | 0.29893 |
| CLEC1B | ENSG00000165682 | 12:9993065-9993287 | - | TE | 0.18561 | 0.004046 | 0.18156 |
| CLEC2D | ENSG00000069493 | 12:9669728-9669734 | + | TS | 0.014097 | 0.19588 | -0.18178 |
| CLEC2D | ENSG00000069493 | 12:9669717-9669727 | + | TS | 0.013921 | 0.13483 | -0.12091 |
| CLK1 | ENSG00000013441 | 2:200853247-200853331 | - | TE | 0.051311 | 0.20545 | -0.15414 |
| CLK1 | ENSG00000013441 | 2:200853332-200853449 | - | TE | 0.56704 | 0.046264 | 0.52078 |
| CLK3 | ENSG00000179335 | 15:74629707-74630116 | + | TE | 0.018355 | 0.14385 | -0.1255 |
| CLK4 | ENSG00000113240 | 5:178621812-178622392 | - | TE | 0.14442 | 0.006406 | 0.13801 |
| CLK4 | ENSG00000113240 | 5:178603589-178603757 | - | TE | 0.004947 | 0.069673 | -0.06473 |
| CLMN | ENSG00000165959 | 14:95191527-95191529 | - | TE | 0.042259 | 0.000441 | 0.041818 |
| CLN5 | ENSG00000102805 | 13:77000458-77002429 | + | TE | 0.019917 | 0.002029 | 0.017888 |
| CLN6 | ENSG00000128973 | 15:68208222-68208410 | - | TE | 0.0946 | 0.021123 | 0.073477 |
| CLN6 | ENSG00000128973 | 15:68229704-68229716 | - | TS | 0.70649 | 0.12969 | 0.5768 |
| CLN6 | ENSG00000128973 | 15:68257107-68257211 | - | TS | 0.328 | 0.070374 | 0.25762 |
| CLOCK | ENSG00000134852 | 4:55434823-55435021 | - | TE | 0.000379 | 0.002257 | -0.00188 |
| CLOCK | ENSG00000134852 | 4:55435022-55435594 | - | TE | 0.000385 | 0.046696 | -0.04631 |
| CLTA | ENSG00000122705 | 9:36212057-36212058 | + | TE | 0.003773 | 0.044464 | -0.04069 |
| CLTRN | ENSG00000147003 | X:15627439-15628127 | - | TE | 0.086577 | 0.85379 | -0.76721 |
| CLTRN | ENSG00000147003 | X:15627318-15627438 | - | TE | 0.90764 | 0.14718 | 0.76046 |
| CMC1 | ENSG00000187118 | 3:28319774-28319776 | + | TE | 0.001954 | 0.028424 | -0.02647 |
| CMTM3 | ENSG00000140931 | 16:66604310-66604449 | + | TS | 0.8579 | 0.1809 | 0.67699 |
| CMTM5 | ENSG00000166091 | 14:23377047-23377062 | + | TS | 0.23383 | 0.050923 | 0.18291 |
| CNEP1R1 | ENSG00000205423 | 16:50035417-50036630 | + | TE | 0.000527 | 0.008488 | -0.00796 |
| CNNM2 | ENSG00000148842 | 10:102918317-102918356 | + | TS | 0.025282 | 0.33372 | -0.30844 |
| CNNM3 | ENSG00000168763 | 2:96816250-96817502 | + | TS | 0.17918 | 0.73357 | -0.55439 |
| CNOT1 | ENSG00000125107 | 16:58520684-58520694 | - | TE | 0.002135 | 0.011737 | -0.0096 |
| CNR1 | ENSG00000118432 | 6:88143856-88145004 | - | TE | 0.018009 | 0.080323 | -0.06231 |
| CNTN4 | ENSG00000144619 | 3:3056356-3057953 | + | TE | 0.93895 | 0.20737 | 0.73158 |
| CNTN4 | ENSG00000144619 | 3:3056120-3056355 | + | TE | 0.060198 | 0.79914 | -0.73894 |
| CNTNAP2 | ENSG00000174469 | 7:148415780-148415940 | + | TE | 0.11958 | 0.016615 | 0.10296 |
| CNTRL | ENSG00000119397 | 9:121177163-121177608 | + | TE | 0.29313 | 0.019311 | 0.27382 |
| COA4 | ENSG00000181924 | 11:73872669-73872996 | - | TE | 0.17548 | 0.74131 | -0.56583 |
| COASY | ENSG00000068120 | 17:42565906-42566037 | + | TE | 0.06162 | 0.46977 | -0.40815 |
| COBLL1 | ENSG00000082438 | 2:164684816-164684824 | - | TE | 0.028705 | 0.006725 | 0.02198 |
| COBLL1 | ENSG00000082438 | 2:164684825-164685820 | - | TE | 0.042963 | 0.006752 | 0.036212 |
| COBLL1 | ENSG00000082438 | 2:164685821-164685903 | - | TE | 0.043173 | 0.010145 | 0.033028 |
| COG5 | ENSG00000164597 | 7:107202564-107203630 | - | TE | 0.000516 | 0.019749 | -0.01923 |
| COL18A1 | ENSG00000182871 | 21:45512188-45512567 | + | TE | 0.02832 | 0.0034 | 0.024921 |
| COL4A3BP | ENSG00000113163 | 5:75373363-75374096 | - | TE | 0.000494 | 0.02388 | -0.02339 |
| COL9A1 | ENSG00000112280 | 6:70216684-70216742 | - | TE | 0.039059 | 0.17002 | -0.13096 |
| COMMD7 | ENSG00000149600 | 20:32702691-32702702 | - | TE | 0.11759 | 0.009139 | 0.10845 |
| COPG1 | ENSG00000181789 | 3:129277294-129277771 | + | TE | 0.009187 | 0.13207 | -0.12288 |
| COQ10A | ENSG00000135469 | 12:56270459-56270964 | + | TE | 0.30729 | 0.053071 | 0.25422 |
| COQ4 | ENSG00000167113 | 9:128334069-128334072 | + | TE | 0.031751 | 0.2672 | -0.23544 |
| COQ6 | ENSG00000119723 | 14:73963109-73963111 | + | TE | 0.049731 | 0.20997 | -0.16024 |
| COX6A1 | ENSG00000111775 | 12:120440738-120440742 | + | TE | 0.022776 | 0.43094 | -0.40816 |
| CPD | ENSG00000108582 | 17:30378905-30379615 | + | TS | 0.087151 | 0.001377 | 0.085774 |
| CPEB4 | ENSG00000113742 | 5:173955910-173956235 | + | TE | 0.004513 | 0.087198 | -0.08269 |
| CPTP | ENSG00000224051 | 1:1327241-1327471 | + | TE | 0.00426 | 0.04854 | -0.04428 |
| CRELD2 | ENSG00000184164 | 22:49918635-49918695 | + | TS | 0.396 | 0.089095 | 0.30691 |
| CREM | ENSG00000095794 | 10:35127117-35127161 | + | TS | 0.078703 | 0.36198 | -0.28328 |
| CRLS1 | ENSG00000088766 | 20:6040044-6040053 | + | TE | 0.014073 | 0.31667 | -0.3026 |
| CRMP1 | ENSG00000072832 | 4:5820784-5821242 | - | TE | 0.54905 | 0.11003 | 0.43902 |
| CRYGN | ENSG00000127377 | 7:151429970-151430000 | - | TE | 0.107 | 0.7382 | -0.63121 |
| CRYZ | ENSG00000116791 | 1:74732956-74733018 | - | TS | 0.091131 | 0.48692 | -0.39579 |
| CRYZL1 | ENSG00000205758 | 21:33589467-33589467 | - | TE | 0.009098 | 0.095351 | -0.08625 |
| CRYZL1 | ENSG00000205758 | 21:33589461-33589466 | - | TE | 0.048288 | 0.007789 | 0.040499 |
| CSAD | ENSG00000139631 | 12:53158139-53158510 | - | TE | 0.002257 | 0.055098 | -0.05284 |
| CSF2RA | ENSG00000198223 | X:1309925-1309934 | + | TE | 0.055472 | 0.00397 | 0.051502 |
| CSF2RA | ENSG00000198223 | X:1309935-1309935 | + | TE | 0.27829 | 0.004005 | 0.27428 |
| CSGALNACT1 | ENSG00000147408 | 8:19405780-19406069 | - | TE | 0.001836 | 0.037325 | -0.03549 |
| CSNK1G3 | ENSG00000151292 | 5:123614456-123614560 | + | TE | 0.003073 | 0.030192 | -0.02712 |
| CSNK2A1 | ENSG00000101266 | 20:482857-482864 | - | TE | 0.026013 | 0.004157 | 0.021856 |
| CSNK2A1 | ENSG00000101266 | 20:498166-498196 | - | TS | 0.11846 | 0.69493 | -0.57647 |
| CSNK2A1 | ENSG00000101266 | 20:472498-472507 | - | TE | 0.18399 | 0.03781 | 0.14618 |
| CTCF | ENSG00000102974 | 16:67638967-67639039 | + | TE | 0.013808 | 0.002515 | 0.011293 |
| CTDSPL2 | ENSG00000137770 | 15:44524544-44525947 | + | TE | 0.008261 | 0.000737 | 0.007524 |
| CTDSPL2 | ENSG00000137770 | 15:44524109-44524543 | + | TE | 0.019103 | 0.002039 | 0.017064 |
| CTNNA1 | ENSG00000044115 | 5:138934832-138935028 | + | TE | 0.002051 | 0.078676 | -0.07663 |
| CTNNB1 | ENSG00000168036 | 3:41240444-41240445 | + | TE | 0.15445 | 0.027515 | 0.12694 |
| CTNNBIP1 | ENSG00000178585 | 1:9910264-9910336 | - | TS | 0.11852 | 0.72927 | -0.61074 |
| CTPS1 | ENSG00000171793 | 1:41011658-41011779 | + | TE | 0.053956 | 0.25663 | -0.20268 |
| CTSC | ENSG00000109861 | 11:88326086-88326116 | - | TE | 0.021606 | 0.001141 | 0.020466 |
| CUL4A | ENSG00000139842 | 13:113263487-113263582 | + | TE | 0.004807 | 0.1109 | -0.10609 |
| CXCR2 | ENSG00000180871 | 2:218134777-218134846 | + | TE | 3.68E-05 | 0.000152 | -0.00011 |
| CXCR2 | ENSG00000180871 | 2:218134847-218135208 | + | TE | 3.73E-05 | 0.000152 | -0.00012 |
| CXCR2 | ENSG00000180871 | 2:218135209-218135318 | + | TE | 3.66E-05 | 0.00927 | -0.00923 |
| CXCR2 | ENSG00000180871 | 2:218135319-218135403 | + | TE | 3.65E-05 | 0.000149 | -0.00011 |
| CXorf38 | ENSG00000185753 | X:40629033-40629936 | - | TE | 0.011902 | 0.050442 | -0.03854 |
| CYB5A | ENSG00000166347 | 18:74253295-74253295 | - | TE | 0.17204 | 0.038988 | 0.13305 |
| CYB5B | ENSG00000103018 | 16:69462430-69466264 | + | TE | 0.002771 | 0.083005 | -0.08023 |
| CYB5R1 | ENSG00000159348 | 1:202961869-202961875 | - | TE | 0.15392 | 0.014183 | 0.13974 |
| CYFIP2 | ENSG00000055163 | 5:157266128-157266135 | + | TS | 0.066221 | 0.27742 | -0.2112 |
| CYFIP2 | ENSG00000055163 | 5:157266123-157266127 | + | TS | 0.065765 | 0.26635 | -0.20058 |
| CYP1A1 | ENSG00000140465 | 15:74720156-74720429 | - | TE | 0.72004 | 0.12482 | 0.59522 |
| CYP20A1 | ENSG00000119004 | 2:203297015-203297226 | + | TE | 0.028615 | 0.15378 | -0.12516 |
| CYP2R1 | ENSG00000186104 | 11:14878009-14878011 | - | TE | 0.019805 | 0.004544 | 0.015261 |
| CYSLTR2 | ENSG00000152207 | 13:48706862-48708176 | + | TE | 0.02066 | 0.10916 | -0.0885 |
| CYYR1 | ENSG00000166265 | 21:26573177-26573284 | - | TS | 0.1086 | 0.45794 | -0.34933 |
| DAB2 | ENSG00000153071 | 5:39373021-39373425 | - | TE | 0.009601 | 0.040892 | -0.03129 |
| DACT1 | ENSG00000165617 | 14:58645473-58647170 | + | TE | 0.039072 | 0.37053 | -0.33146 |
| DAGLB | ENSG00000164535 | 7:6447935-6448012 | - | TS | 0.1468 | 0.85993 | -0.71313 |
| DAGLB | ENSG00000164535 | 7:6409600-6410035 | - | TE | 0.052828 | 0.31599 | -0.26316 |
| DAGLB | ENSG00000164535 | 7:6447933-6447934 | - | TS | 0.76979 | 0.1485 | 0.62129 |
| DAPK3 | ENSG00000167657 | 19:3958458-3959637 | - | TE | 0.022134 | 0.25893 | -0.2368 |
| DARS2 | ENSG00000117593 | 1:173824847-173824959 | + | TS | 0.15706 | 0.02212 | 0.13494 |
| DAZAP2 | ENSG00000183283 | 12:51238826-51238836 | + | TS | 0.056021 | 0.0124 | 0.043621 |
| DBF4B | ENSG00000161692 | 17:44731016-44731218 | + | TE | 0.12224 | 0.55282 | -0.43058 |
| DBP | ENSG00000105516 | 19:48630693-48630730 | - | TE | 0.11719 | 0.023089 | 0.094101 |
| DCAF10 | ENSG00000122741 | 9:37861140-37863325 | + | TE | 0.006314 | 0.027525 | -0.02121 |
| DCAF17 | ENSG00000115827 | 2:171480974-171481132 | + | TE | 0.054248 | 0.003266 | 0.050982 |
| DCHS2 | ENSG00000197410 | 4:154231742-154232036 | - | TE | 0.089672 | 0.88605 | -0.79637 |
| DCHS2 | ENSG00000197410 | 4:154232037-154237159 | - | TE | 0.91152 | 0.10814 | 0.80338 |
| DCP2 | ENSG00000172795 | 5:113013850-113020123 | + | TE | 0.005666 | 0.064124 | -0.05846 |
| DCUN1D4 | ENSG00000109184 | 4:51913529-51914036 | + | TE | 0.004758 | 0.024715 | -0.01996 |
| DDAH2 | ENSG00000213722 | 6:31730246-31730617 | - | TS | 0.005916 | 0.034106 | -0.02819 |
| DDOST | ENSG00000244038 | 1:20651767-20651776 | - | TE | 0.015438 | 0.12864 | -0.1132 |
| DDT | ENSG00000099977 | 22:23971370-23971623 | - | TE | 0.037823 | 0.20591 | -0.16808 |
| DDX10 | ENSG00000178105 | 11:108665069-108665081 | + | TS | 0.090993 | 0.37706 | -0.28606 |
| DDX10 | ENSG00000178105 | 11:108940919-108940925 | + | TE | 0.035501 | 0.14838 | -0.11288 |
| DDX19A | ENSG00000168872 | 16:70372090-70372297 | + | TE | 0.06505 | 0.005522 | 0.059528 |
| DDX24 | ENSG00000089737 | 14:94048291-94050919 | - | TE | 0.29085 | 0.024508 | 0.26634 |
| DDX24 | ENSG00000089737 | 14:94050938-94051019 | - | TE | 0.019484 | 0.095913 | -0.07643 |
| DDX3X | ENSG00000215301 | X:41348584-41349694 | + | TE | 0.001937 | 0.046948 | -0.04501 |
| DDX42 | ENSG00000198231 | 17:63819210-63819247 | + | TE | 0.002731 | 0.012106 | -0.00937 |
| DDX5 | ENSG00000108654 | 17:64499914-64500109 | - | TE | 0.000918 | 0.013414 | -0.0125 |
| DDX59 | ENSG00000118197 | 1:200669908-200669969 | - | TS | 0.47536 | 0.11031 | 0.36505 |
| DELE1 | ENSG00000081791 | 5:141938521-141938948 | + | TE | 0.001756 | 0.042806 | -0.04105 |
| DENND1C | ENSG00000205744 | 19:6467210-6467497 | - | TE | 0.022457 | 0.1555 | -0.13304 |
| DENND4C | ENSG00000137145 | 9:19372037-19372594 | + | TE | 0.023782 | 0.004588 | 0.019194 |
| DEPDC1B | ENSG00000035499 | 5:60596912-60597628 | - | TE | 0.67524 | 0.16365 | 0.51159 |
| DERA | ENSG00000023697 | 12:16036814-16037280 | + | TE | 0.14673 | 0.007647 | 0.13909 |
| DERA | ENSG00000023697 | 12:16036690-16036813 | + | TE | 0.080973 | 0.007526 | 0.073448 |
| DFFB | ENSG00000169598 | 1:3857267-3857280 | + | TS | 0.32352 | 0.058915 | 0.26461 |
| DGKZ | ENSG00000149091 | 11:46379831-46379947 | + | TE | 0.011909 | 0.099564 | -0.08766 |
| DGUOK | ENSG00000114956 | 2:73958960-73958961 | + | TE | 0.065738 | 0.008454 | 0.057284 |
| DHFR | ENSG00000228716 | 5:80628017-80628123 | - | TE | 0.021875 | 0.003763 | 0.018112 |
| DHRS13 | ENSG00000167536 | 17:28897781-28897783 | - | TE | 0.41119 | 0.091383 | 0.31981 |
| DHX33 | ENSG00000005100 | 17:5468749-5468829 | - | TS | 0.35678 | 0.08116 | 0.27562 |
| DICER1 | ENSG00000100697 | 14:95090492-95090663 | - | TE | 0.000285 | 0.021677 | -0.02139 |
| DIO1 | ENSG00000211452 | 1:53910108-53910109 | + | TE | 0.45509 | 0.066892 | 0.38819 |
| DKC1 | ENSG00000130826 | X:154776799-154776911 | + | TE | 0.13614 | 0.01086 | 0.12528 |
| DLG2 | ENSG00000150672 | 11:83459609-83459788 | - | TE | 0.80995 | 0.142 | 0.66795 |
| DLG2 | ENSG00000150672 | 11:83457919-83459065 | - | TE | 0.11763 | 0.53939 | -0.42176 |
| DLG4 | ENSG00000132535 | 17:7189895-7190285 | - | TE | 0.23873 | 0.051198 | 0.18753 |
| DLGAP4 | ENSG00000080845 | 20:36527232-36527239 | + | TE | 0.035169 | 0.004367 | 0.030802 |
| DMAC2 | ENSG00000105341 | 19:41431339-41431755 | - | TE | 0.020814 | 0.085065 | -0.06425 |
| DMTF1 | ENSG00000135164 | 7:87152426-87152467 | + | TS | 0.077962 | 0.32663 | -0.24867 |
| DMXL2 | ENSG00000104093 | 15:51447788-51447790 | - | TE | 0.028478 | 0.12707 | -0.0986 |
| DNAJB1 | ENSG00000132002 | 19:14515649-14516170 | - | TE | 0.017828 | 0.002149 | 0.015679 |
| DNAJB11 | ENSG00000090520 | 3:186585794-186585799 | + | TE | 0.033614 | 0.14857 | -0.11496 |
| DNAJC13 | ENSG00000138246 | 3:132539022-132539032 | + | TE | 0.21637 | 0.003685 | 0.21268 |
| DNAJC14 | ENSG00000135392 | 12:55821829-55822110 | - | TE | 0.011508 | 0.090818 | -0.07931 |
| DNAJC19 | ENSG00000205981 | 3:180989715-180989727 | - | TS | 0.16924 | 0.040587 | 0.12865 |
| DNAJC21 | ENSG00000168724 | 5:34955201-34955867 | + | TE | 0.050131 | 0.01139 | 0.038742 |
| DNAJC21 | ENSG00000168724 | 5:34955169-34955200 | + | TE | 0.084906 | 0.012206 | 0.0727 |
| DNAJC27 | ENSG00000115137 | 2:24947359-24947748 | - | TE | 0.005082 | 0.038468 | -0.03339 |
| DNAJC28 | ENSG00000177692 | 21:33488208-33489424 | - | TE | 0.50495 | 0.11616 | 0.38879 |
| DNAJC4 | ENSG00000110011 | 11:64230567-64230757 | + | TS | 0.084547 | 0.008727 | 0.07582 |
| DNAJC4 | ENSG00000110011 | 11:64234282-64234286 | + | TE | 0.22173 | 0.034157 | 0.18757 |
| DNAJC5B | ENSG00000147570 | 8:66099937-66100134 | + | TE | 0.077587 | 0.41206 | -0.33448 |
| DNASE1L2 | ENSG00000167968 | 16:2236467-2236468 | + | TS | 0.51516 | 0.12599 | 0.38916 |
| DNHD1 | ENSG00000179532 | 11:6519965-6520858 | + | TE | 0.54524 | 0.026538 | 0.51871 |
| DOCK2 | ENSG00000134516 | 5:170082796-170082934 | + | TE | 0.25968 | 0.001892 | 0.25779 |
| DOCK4 | ENSG00000128512 | 7:111728220-111728720 | - | TE | 0.004411 | 0.042585 | -0.03817 |
| DOCK8 | ENSG00000107099 | 9:464159-464526 | + | TE | 0.00541 | 0.037347 | -0.03194 |
| DOLPP1 | ENSG00000167130 | 9:129090436-129090438 | + | TE | 0.062429 | 0.40579 | -0.34337 |
| DPP8 | ENSG00000074603 | 15:65517700-65517704 | - | TS | 0.7145 | 0.14202 | 0.57247 |
| DRAP1 | ENSG00000175550 | 11:65921555-65921561 | + | TE | 0.12105 | 0.5202 | -0.39916 |
| DSE | ENSG00000111817 | 6:116438244-116438277 | + | TE | 0.006013 | 0.000808 | 0.005205 |
| DSN1 | ENSG00000149636 | 20:36751791-36751794 | - | TE | 0.1591 | 0.015988 | 0.14311 |
| DTNA | ENSG00000134769 | 18:34866370-34866980 | + | TE | 0.1246 | 0.67871 | -0.55411 |
| DUSP10 | ENSG00000143507 | 1:221702427-221702677 | - | TE | 0.04526 | 0.010792 | 0.034468 |
| DUSP18 | ENSG00000167065 | 22:30662031-30663403 | - | TE | 0.00147 | 0.070408 | -0.06894 |
| DUSP19 | ENSG00000162999 | 2:183095431-183095899 | + | TE | 0.042855 | 0.37936 | -0.33651 |
| DUSP22 | ENSG00000112679 | 6:351354-351355 | + | TE | 0.00384 | 0.3422 | -0.33836 |
| DUT | ENSG00000128951 | 15:48342294-48342395 | + | TE | 0.22004 | 0.019966 | 0.20008 |
| DVL1 | ENSG00000107404 | 1:1335276-1335278 | - | TE | 0.30302 | 0.024087 | 0.27893 |
| DYNC1H1 | ENSG00000197102 | 14:102050672-102050716 | + | TE | 0.004666 | 0.073115 | -0.06845 |
| DYNC1H1 | ENSG00000197102 | 14:101964573-101964687 | + | TS | 0.81181 | 0.16603 | 0.64577 |
| DYNC1I2 | ENSG00000077380 | 2:171748410-171748411 | + | TE | 0.01088 | 0.099493 | -0.08861 |
| DYNC2H1 | ENSG00000187240 | 11:103109431-103109513 | + | TS | 0.11561 | 0.51138 | -0.39577 |
| DYRK2 | ENSG00000127334 | 12:67648778-67649182 | + | TS | 0.082075 | 0.453 | -0.37092 |
| EBAG9 | ENSG00000147654 | 8:109564443-109564559 | + | TE | 0.012747 | 0.10112 | -0.08837 |
| EBF1 | ENSG00000164330 | 5:158698664-158699142 | - | TE | 0.17662 | 0.02239 | 0.15423 |
| ECE1 | ENSG00000117298 | 1:21219894-21219936 | - | TE | 0.008543 | 0.000416 | 0.008127 |
| ECHDC1 | ENSG00000093144 | 6:127288712-127289266 | - | TE | 0.37213 | 0.072578 | 0.29955 |
| ECHDC1 | ENSG00000093144 | 6:127289634-127289779 | - | TE | 0.006274 | 0.039139 | -0.03287 |
| ECSIT | ENSG00000130159 | 19:11506075-11506183 | - | TE | 0.025091 | 0.13492 | -0.10983 |
| EDNRB | ENSG00000136160 | 13:77895526-77896407 | - | TE | 0.063901 | 0.29742 | -0.23352 |
| EEF1AKMT2 | ENSG00000203791 | 10:124760146-124760317 | - | TE | 0.01441 | 0.069039 | -0.05463 |
| EEF1G | ENSG00000254772 | 11:62559603-62559837 | - | TE | 0.051252 | 0.003877 | 0.047375 |
| EFCAB11 | ENSG00000140025 | 14:89923308-89923325 | - | TE | 0.31366 | 0.049967 | 0.2637 |
| EFCAB11 | ENSG00000140025 | 14:89795147-89797324 | - | TE | 0.14651 | 0.89009 | -0.74359 |
| EFCAB11 | ENSG00000140025 | 14:89794669-89795146 | - | TE | 0.85647 | 0.11205 | 0.74442 |
| EFCAB2 | ENSG00000203666 | 1:245121920-245121981 | + | TE | 0.002203 | 0.11285 | -0.11065 |
| EFEMP2 | ENSG00000172638 | 11:65866447-65866712 | - | TE | 0.40333 | 0.087097 | 0.31623 |
| EFNB2 | ENSG00000125266 | 13:106535115-106535662 | - | TS | 0.62085 | 0.14122 | 0.47963 |
| EFTUD2 | ENSG00000108883 | 17:44850917-44851119 | - | TE | 0.052461 | 0.004331 | 0.04813 |
| EIF2B4 | ENSG00000115211 | 2:27364358-27364376 | - | TE | 0.31725 | 0.070078 | 0.24717 |
| EIF2B5 | ENSG00000145191 | 3:184145114-184145123 | + | TE | 0.038794 | 0.19201 | -0.15321 |
| EIF2B5 | ENSG00000145191 | 3:184145303-184145311 | + | TE | 0.30022 | 0.059511 | 0.2407 |
| EIF4A2 | ENSG00000156976 | 3:186789125-186789291 | + | TE | 0.002357 | 0.067466 | -0.06511 |
| EIF4G1 | ENSG00000114867 | 3:184321282-184321534 | + | TE | 0.005132 | 0.000664 | 0.004468 |
| ELF1 | ENSG00000120690 | 13:40932028-40932033 | - | TE | 0.07205 | 0.00986 | 0.062191 |
| ELOC | ENSG00000154582 | 8:73946264-73946472 | - | TE | 0.004961 | 0.034628 | -0.02967 |
| ELP3 | ENSG00000134014 | 8:28190095-28190203 | + | TE | 0.045609 | 0.011078 | 0.03453 |
| EMC1 | ENSG00000127463 | 1:19218945-19219369 | - | TE | 0.0022 | 0.039012 | -0.03681 |
| EMC4 | ENSG00000128463 | 15:34229753-34229980 | + | TE | 0.006268 | 0.054662 | -0.04839 |
| EMC9 | ENSG00000100908 | 14:24138965-24139196 | - | TE | 0.013842 | 0.16908 | -0.15524 |
| EME2 | ENSG00000197774 | 16:1778455-1780627 | + | TE | 0.004862 | 0.029041 | -0.02418 |
| EMP3 | ENSG00000142227 | 19:48330356-48330483 | + | TE | 0.074829 | 0.005092 | 0.069737 |
| EMSY | ENSG00000158636 | 11:76550344-76550363 | + | TE | 0.024849 | 0.003891 | 0.020957 |
| ENG | ENSG00000106991 | 9:127815017-127815806 | - | TE | 0.68938 | 0.14756 | 0.54182 |
| ENPP2 | ENSG00000136960 | 8:119557086-119557086 | - | TE | 0.1081 | 0.022786 | 0.085315 |
| ENTPD4 | ENSG00000197217 | 8:23432926-23433154 | - | TE | 0.000438 | 0.017508 | -0.01707 |
| EOGT | ENSG00000163378 | 3:68977618-68977764 | - | TE | 0.015613 | 0.42358 | -0.40797 |
| EPAS1 | ENSG00000116016 | 2:46386697-46386703 | + | TE | 0.14152 | 0.011753 | 0.12977 |
| EPB41 | ENSG00000159023 | 1:29072315-29072350 | + | TE | 0.014991 | 0.12328 | -0.10829 |
| EPB41L4A | ENSG00000129595 | 5:112419314-112419316 | - | TS | 0.61466 | 0.076261 | 0.5384 |
| EPHA1 | ENSG00000146904 | 7:143391411-143391535 | - | TE | 0.094435 | 0.007875 | 0.08656 |
| EPN2 | ENSG00000072134 | 17:19335175-19335194 | + | TE | 0.074422 | 0.30713 | -0.2327 |
| EPS15 | ENSG00000085832 | 1:51519242-51519245 | - | TS | 0.060891 | 0.2441 | -0.18321 |
| EPS8 | ENSG00000151491 | 12:15620412-15620982 | - | TE | 0.40586 | 0.055566 | 0.3503 |
| ERAL1 | ENSG00000132591 | 17:28861060-28861067 | + | TE | 0.014451 | 0.11321 | -0.09876 |
| ERC1 | ENSG00000082805 | 12:1491568-1492487 | + | TE | 0.014366 | 0.001944 | 0.012422 |
| ERCC1 | ENSG00000012061 | 19:45423873-45423938 | - | TS | 0.17349 | 0.041715 | 0.13177 |
| ERCC3 | ENSG00000163161 | 2:127257297-127257302 | - | TE | 0.085214 | 0.37978 | -0.29456 |
| ERGIC2 | ENSG00000087502 | 12:29341033-29341218 | - | TE | 0.000586 | 0.077829 | -0.07724 |
| ERRFI1 | ENSG00000116285 | 1:8014040-8014396 | - | TE | 0.19312 | 0.04073 | 0.15239 |
| ERVK3-1 | ENSG00000142396 | 19:58315203-58315203 | + | TE | 0.139 | 0.009775 | 0.12922 |
| ERVW-1 | ENSG00000242950 | 7:92477947-92477986 | - | TS | 0.61202 | 0.097764 | 0.51426 |
| ESCO1 | ENSG00000141446 | 18:21529303-21529309 | - | TE | 0.10223 | 0.005953 | 0.096279 |
| ESPL1 | ENSG00000135476 | 12:53293636-53293638 | + | TE | 0.12695 | 0.88605 | -0.7591 |
| ESPL1 | ENSG00000135476 | 12:53293273-53293635 | + | TE | 0.86555 | 0.1135 | 0.75205 |
| ESR1 | ENSG00000091831 | 6:151807858-151808166 | + | TE | 0.14599 | 0.72621 | -0.58022 |
| ESRRA | ENSG00000173153 | 11:64316030-64316738 | + | TE | 0.34228 | 0.064698 | 0.27758 |
| EXOC6 | ENSG00000138190 | 10:93058525-93059490 | + | TE | 0.060218 | 0.40236 | -0.34215 |
| EXOC7 | ENSG00000182473 | 17:76083577-76083647 | - | TE | 0.009776 | 0.052823 | -0.04305 |
| EXOSC3 | ENSG00000107371 | 9:37780588-37780606 | - | TE | 0.59966 | 0.074297 | 0.52536 |
| EXPH5 | ENSG00000110723 | 11:108511367-108514875 | - | TE | 0.11187 | 0.022079 | 0.089791 |
| EZH1 | ENSG00000108799 | 17:42702385-42702592 | - | TE | 0.074872 | 0.011036 | 0.063836 |
| EZH1 | ENSG00000108799 | 17:42702292-42702324 | - | TE | 0.002362 | 0.015433 | -0.01307 |
| F8 | ENSG00000185010 | X:154835788-154835794 | - | TE | 0.15562 | 0.018822 | 0.1368 |
| FADS2 | ENSG00000134824 | 11:61867348-61867354 | + | TE | 0.01105 | 0.31575 | -0.3047 |
| FAF1 | ENSG00000185104 | 1:50441276-50441523 | - | TE | 0.009382 | 0.044045 | -0.03466 |
| FAH | ENSG00000103876 | 15:80162337-80163131 | + | TE | 0.077118 | 0.46715 | -0.39003 |
| FAHD1 | ENSG00000180185 | 16:1826967-1827865 | + | TS | 0.013248 | 0.06379 | -0.05054 |
| FAHD2A | ENSG00000115042 | 2:95402721-95402725 | + | TS | 0.14996 | 0.86338 | -0.71341 |
| FAHD2A | ENSG00000115042 | 2:95402726-95402872 | + | TS | 0.85772 | 0.14015 | 0.71757 |
| FAM104B | ENSG00000182518 | X:55145356-55145386 | - | TE | 0.067367 | 0.38761 | -0.32024 |
| FAM107A | ENSG00000168309 | 3:58565461-58565851 | - | TE | 0.11786 | 0.51441 | -0.39655 |
| FAM114A2 | ENSG00000055147 | 5:153992955-153993110 | - | TE | 0.017854 | 0.36405 | -0.3462 |
| FAM122C | ENSG00000156500 | X:134810976-134811150 | + | TE | 0.024657 | 0.48523 | -0.46058 |
| FAM129C | ENSG00000167483 | 19:17553635-17553639 | + | TE | 0.24456 | 0.036097 | 0.20847 |
| FAM129C | ENSG00000167483 | 19:17553640-17553645 | + | TE | 0.093955 | 0.014418 | 0.079537 |
| FAM13A | ENSG00000138640 | 4:88728179-88728659 | - | TE | 0.011886 | 0.0011 | 0.010786 |
| FAM13A | ENSG00000138640 | 4:88727069-88727425 | - | TE | 0.013792 | 0.001097 | 0.012695 |
| FAM13B | ENSG00000031003 | 5:137940172-137940180 | - | TE | 0.006521 | 0.032375 | -0.02585 |
| FAM13B | ENSG00000031003 | 5:138033086-138033086 | - | TS | 0.27444 | 0.049975 | 0.22446 |
| FAM160B2 | ENSG00000158863 | 8:22097035-22097045 | + | TE | 0.17781 | 0.031332 | 0.14648 |
| FAM167A | ENSG00000154319 | 8:11424018-11424636 | - | TE | 0.13695 | 0.031902 | 0.10505 |
| FAM172A | ENSG00000113391 | 5:93620883-93620928 | - | TE | 0.01134 | 0.076217 | -0.06488 |
| FAM193A | ENSG00000125386 | 4:2731775-2731889 | + | TE | 0.074137 | 0.004298 | 0.069839 |
| FAM199X | ENSG00000123575 | X:104189608-104191446 | + | TE | 0.000649 | 0.014737 | -0.01409 |
| FAM208A | ENSG00000163946 | 3:56620545-56622858 | - | TE | 0.00634 | 0.000957 | 0.005384 |
| FAM214B | ENSG00000005238 | 9:35104123-35104141 | - | TE | 0.023336 | 0.000631 | 0.022705 |
| FAM217B | ENSG00000196227 | 20:59944396-59945000 | + | TE | 0.000804 | 0.014543 | -0.01374 |
| FAM234A | ENSG00000167930 | 16:266121-266125 | + | TE | 0.14973 | 0.011913 | 0.13782 |
| FAM90A1 | ENSG00000171847 | 12:8221260-8221262 | - | TE | 0.11053 | 0.5348 | -0.42427 |
| FANCM | ENSG00000187790 | 14:45200888-45200890 | + | TE | 0.050618 | 0.32705 | -0.27644 |
| FAR2 | ENSG00000064763 | 12:29334073-29334073 | + | TE | 0.004788 | 0.073958 | -0.06917 |
| FARS2 | ENSG00000145982 | 6:5771291-5771429 | + | TE | 0.50965 | 0.12489 | 0.38475 |
| FAS | ENSG00000026103 | 10:89014620-89015532 | + | TE | 0.000376 | 0.083744 | -0.08337 |
| FBLN5 | ENSG00000140092 | 14:91869412-91869440 | - | TE | 0.051659 | 0.27432 | -0.22266 |
| FBXL13 | ENSG00000161040 | 7:102813230-102813232 | - | TE | 0.11403 | 0.85594 | -0.74192 |
| FBXL13 | ENSG00000161040 | 7:102813233-102813274 | - | TE | 0.89404 | 0.1429 | 0.75114 |
| FBXL14 | ENSG00000171823 | 12:1593039-1594165 | - | TS | 0.11234 | 0.004724 | 0.10761 |
| FBXL19 | ENSG00000099364 | 16:30948777-30948783 | + | TE | 0.066981 | 0.31313 | -0.24615 |
| FBXO24 | ENSG00000106336 | 7:100600534-100601025 | + | TE | 0.61514 | 0.060702 | 0.55444 |
| FBXO25 | ENSG00000147364 | 8:469691-469873 | + | TE | 0.002905 | 0.012657 | -0.00975 |
| FBXO32 | ENSG00000156804 | 8:123503127-123503462 | - | TE | 0.076423 | 0.007486 | 0.068937 |
| FBXO42 | ENSG00000037637 | 1:16251082-16251785 | - | TE | 0.063558 | 0.004736 | 0.058822 |
| FBXO5 | ENSG00000112029 | 6:152970519-152970528 | - | TE | 0.29872 | 0.052673 | 0.24604 |
| FBXW11 | ENSG00000072803 | 5:171863621-171864101 | - | TE | 0.008727 | 0.001452 | 0.007275 |
| FCHSD1 | ENSG00000197948 | 5:141639308-141640981 | - | TE | 0.10457 | 0.011828 | 0.092746 |
| FCRLA | ENSG00000132185 | 1:161713453-161713468 | + | TE | 0.11041 | 0.006128 | 0.10428 |
| FDFT1 | ENSG00000079459 | 8:11838968-11839114 | + | TE | 0.020122 | 0.004645 | 0.015478 |
| FEZ2 | ENSG00000171055 | 2:36552304-36552946 | - | TE | 0.035918 | 0.17071 | -0.13479 |
| FGF11 | ENSG00000161958 | 17:7444903-7444937 | + | TE | 0.24058 | 0.047663 | 0.19292 |
| FGFBP2 | ENSG00000137441 | 4:15960248-15960611 | - | TE | 0.0621 | 0.30165 | -0.23955 |
| FGFR1 | ENSG00000077782 | 8:38413522-38413804 | - | TE | 0.005243 | 0.13354 | -0.1283 |
| FGFR1OP2 | ENSG00000111790 | 12:26938497-26938499 | + | TS | 0.064145 | 0.27952 | -0.21537 |
| FHL2 | ENSG00000115641 | 2:105360857-105360860 | - | TE | 0.10203 | 0.006422 | 0.095612 |
| FHL3 | ENSG00000183386 | 1:37997345-37997559 | - | TE | 0.001001 | 0.0096 | -0.0086 |
| FKBP10 | ENSG00000141756 | 17:41822294-41822540 | + | TE | 0.11855 | 0.69033 | -0.57178 |
| FKBP14 | ENSG00000106080 | 7:30026312-30026419 | - | TS | 0.083382 | 0.34908 | -0.2657 |
| FKBP8 | ENSG00000105701 | 19:18531752-18531761 | - | TE | 0.001617 | 0.000226 | 0.001391 |
| FLNB | ENSG00000136068 | 3:58170575-58170774 | + | TE | 0.060178 | 0.012338 | 0.04784 |
| FLT1 | ENSG00000102755 | 13:28385548-28385566 | - | TE | 0.038958 | 0.26975 | -0.23079 |
| FMC1 | ENSG00000164898 | 7:139345501-139345777 | + | TE | 0.12012 | 0.8239 | -0.70378 |
| FMC1 | ENSG00000164898 | 7:139345791-139346319 | + | TE | 0.876 | 0.18643 | 0.68957 |
| FMR1 | ENSG00000102081 | X:147951113-147951119 | + | TE | 0.061376 | 0.01219 | 0.049186 |
| FN3KRP | ENSG00000141560 | 17:82726833-82727097 | + | TE | 0.108 | 0.011223 | 0.096777 |
| FNBP4 | ENSG00000109920 | 11:47724901-47726904 | - | TS | 0.13593 | 0.002684 | 0.13324 |
| FNDC5 | ENSG00000160097 | 1:32863556-32863776 | - | TE | 0.024738 | 0.11403 | -0.08929 |
| FNDC5 | ENSG00000160097 | 1:32862307-32863555 | - | TE | 0.366 | 0.071383 | 0.29462 |
| FOCAD | ENSG00000188352 | 9:20995556-20995948 | + | TE | 0.006072 | 0.14916 | -0.14309 |
| FOXK2 | ENSG00000141568 | 17:82602393-82602405 | + | TE | 0.09712 | 0.002086 | 0.095034 |
| FOXN3 | ENSG00000053254 | 14:89161385-89161399 | - | TE | 0.004865 | 0.000255 | 0.00461 |
| FOXP1 | ENSG00000114861 | 3:70959247-70959247 | - | TE | 0.014592 | 0.10645 | -0.09186 |
| FOXP1 | ENSG00000114861 | 3:70959248-70959315 | - | TE | 0.014564 | 0.1065 | -0.09194 |
| FOXP1 | ENSG00000114861 | 3:70959242-70959246 | - | TE | 0.50408 | 0.1065 | 0.39758 |
| FOXP1 | ENSG00000114861 | 3:71003408-71005336 | - | TS | 0.003556 | 0.031542 | -0.02799 |
| FOXRED2 | ENSG00000100350 | 22:36489987-36490267 | - | TE | 0.099832 | 0.021854 | 0.077978 |
| FRMD6 | ENSG00000139926 | 14:51730460-51730727 | + | TE | 0.44294 | 0.095782 | 0.34716 |
| FRMD8 | ENSG00000126391 | 11:65411781-65413522 | + | TE | 0.11381 | 0.027034 | 0.086777 |
| FSTL3 | ENSG00000070404 | 19:683393-683399 | + | TE | 0.02494 | 0.13906 | -0.11412 |
| FTSJ1 | ENSG00000068438 | X:48486300-48486305 | + | TE | 0.1554 | 0.013873 | 0.14152 |
| FUS | ENSG00000089280 | 16:31191603-31191605 | + | TE | 0.018431 | 0.1462 | -0.12776 |
| FUT10 | ENSG00000172728 | 8:33370824-33371060 | - | TE | 0.2875 | 0.071478 | 0.21602 |
| FUZ | ENSG00000010361 | 19:49806897-49807374 | - | TE | 0.5484 | 0.038843 | 0.50956 |
| FXR1 | ENSG00000114416 | 3:180976546-180976579 | + | TE | 0.005462 | 0.001174 | 0.004289 |
| GABARAPL1 | ENSG00000139112 | 12:10221787-10223123 | + | TE | 0.012756 | 0.081384 | -0.06863 |
| GABBR1 | ENSG00000204681 | 6:29602235-29602237 | - | TE | 0.090648 | 0.021711 | 0.068937 |
| GABRB3 | ENSG00000166206 | 15:26547633-26548134 | - | TE | 0.10921 | 0.51175 | -0.40253 |
| GALNT10 | ENSG00000164574 | 5:154419013-154419033 | + | TE | 0.001258 | 0.028919 | -0.02766 |
| GAR1 | ENSG00000109534 | 4:109824738-109824740 | + | TE | 0.038644 | 0.15762 | -0.11898 |
| GAS2 | ENSG00000148935 | 11:22812997-22813001 | + | TE | 0.076717 | 0.33613 | -0.25942 |
| GAS7 | ENSG00000007237 | 17:9915255-9915977 | - | TE | 0.004865 | 9.59E-05 | 0.004769 |
| GAS8 | ENSG00000141013 | 16:90036767-90036926 | + | TE | 0.11221 | 0.57935 | -0.46714 |
| GATD3B | ENSG00000280071 | 21:5128373-5128378 | - | TS | 0.32277 | 0.072809 | 0.24996 |
| GBA2 | ENSG00000070610 | 9:35748346-35748973 | - | TS | 0.11283 | 0.027187 | 0.085644 |
| GBGT1 | ENSG00000148288 | 9:133152948-133152952 | - | TE | 0.003645 | 0.041501 | -0.03786 |
| GBP1 | ENSG00000117228 | 1:89053362-89053468 | - | TE | 0.000389 | 0.00258 | -0.00219 |
| GBP5 | ENSG00000154451 | 1:89260582-89260817 | - | TE | 0.000167 | 0.000679 | -0.00051 |
| GCLC | ENSG00000001084 | 6:53497341-53497383 | - | TE | 0.13329 | 0.006126 | 0.12716 |
| GCNT1 | ENSG00000187210 | 9:76502470-76507414 | + | TE | 0.067876 | 0.014609 | 0.053267 |
| GCSAM | ENSG00000174500 | 3:112123096-112123224 | - | TE | 0.049533 | 0.010149 | 0.039384 |
| GDAP1L1 | ENSG00000124194 | 20:44280374-44280917 | + | TE | 0.89345 | 0.20183 | 0.69162 |
| GDAP1L1 | ENSG00000124194 | 20:44279236-44280373 | + | TE | 0.10615 | 0.76157 | -0.65542 |
| GDE1 | ENSG00000006007 | 16:19503101-19503617 | - | TE | 0.003622 | 0.035593 | -0.03197 |
| GDI2 | ENSG00000057608 | 10:5765229-5765953 | - | TE | 0.004999 | 0.082609 | -0.07761 |
| GEMIN4 | ENSG00000179409 | 17:744623-746630 | - | TE | 0.032916 | 0.15076 | -0.11785 |
| GET4 | ENSG00000239857 | 7:895334-896434 | + | TE | 0.005103 | 0.024884 | -0.01978 |
| GGACT | ENSG00000134864 | 13:100532495-100532513 | - | TE | 0.023176 | 0.13342 | -0.11025 |
| GIGYF2 | ENSG00000204120 | 2:232858609-232858612 | + | TE | 0.029473 | 0.22994 | -0.20046 |
| GIMAP4 | ENSG00000133574 | 7:150572129-150572938 | + | TE | 0.000113 | 0.021543 | -0.02143 |
| GIPC3 | ENSG00000179855 | 19:3585553-3585575 | + | TS | 0.3462 | 0.071645 | 0.27455 |
| GIT1 | ENSG00000108262 | 17:29574451-29574914 | - | TE | 0.004256 | 0.035643 | -0.03139 |
| GIT1 | ENSG00000108262 | 17:29573469-29573474 | - | TE | 0.067006 | 0.004461 | 0.062545 |
| GIT2 | ENSG00000139436 | 12:109929804-109933037 | - | TE | 0.000271 | 0.002563 | -0.00229 |
| GK5 | ENSG00000175066 | 3:142163572-142163822 | - | TE | 0.12446 | 0.003803 | 0.12066 |
| GLCE | ENSG00000138604 | 15:69160634-69160757 | + | TS | 0.19914 | 0.8918 | -0.69266 |
| GLCE | ENSG00000138604 | 15:69160584-69160633 | + | TS | 0.80788 | 0.10402 | 0.70386 |
| GLDN | ENSG00000186417 | 15:51383893-51384222 | + | TE | 0.13815 | 0.67517 | -0.53702 |
| GLG1 | ENSG00000090863 | 16:74607106-74607112 | - | TS | 0.35505 | 0.057447 | 0.29761 |
| GLS | ENSG00000115419 | 2:190932716-190933008 | + | TE | 0.035165 | 0.002789 | 0.032376 |
| GLT8D1 | ENSG00000016864 | 3:52694492-52695035 | - | TE | 0.15545 | 0.016448 | 0.13901 |
| GMNN | ENSG00000112312 | 6:24785638-24786095 | + | TE | 0.022252 | 0.25874 | -0.23648 |
| GMPR | ENSG00000137198 | 6:16295006-16295121 | + | TE | 0.004247 | 0.050378 | -0.04613 |
| GMPR2 | ENSG00000100938 | 14:24239240-24239242 | + | TE | 0.002303 | 0.028895 | -0.02659 |
| GMPS | ENSG00000163655 | 3:155937591-155937710 | + | TE | 0.13984 | 0.003111 | 0.13673 |
| GNA12 | ENSG00000146535 | 7:2728561-2728565 | - | TE | 0.33399 | 0.045551 | 0.28844 |
| GNAL | ENSG00000141404 | 18:11881136-11881172 | + | TE | 0.026368 | 0.002144 | 0.024224 |
| GNAS | ENSG00000087460 | 20:58841644-58841875 | + | TS | 0.089302 | 0.37739 | -0.28809 |
| GNB4 | ENSG00000114450 | 3:179400849-179401086 | - | TE | 0.011722 | 0.053514 | -0.04179 |
| GNG2 | ENSG00000186469 | 14:51860646-51860790 | + | TS | 0.12225 | 0.50151 | -0.37926 |
| GNPDA1 | ENSG00000113552 | 5:142000678-142002129 | - | TE | 0.003876 | 0.12973 | -0.12586 |
| GNS | ENSG00000135677 | 12:64716475-64716819 | - | TE | 0.000216 | 0.009588 | -0.00937 |
| GOLGA2 | ENSG00000167110 | 9:128257067-128257086 | - | TE | 0.06221 | 0.010724 | 0.051485 |
| GOLGB1 | ENSG00000173230 | 3:121663199-121663202 | - | TE | 0.15929 | 0.009657 | 0.14964 |
| GOLT1B | ENSG00000111711 | 12:21516185-21516231 | + | TE | 0.002862 | 0.035476 | -0.03261 |
| GOLT1B | ENSG00000111711 | 12:21516063-21516184 | + | TE | 0.002848 | 0.024279 | -0.02143 |
| GORASP1 | ENSG00000114745 | 3:39107479-39107569 | - | TS | 0.67297 | 0.14987 | 0.5231 |
| GORASP2 | ENSG00000115806 | 2:170966602-170966623 | + | TE | 0.004377 | 0.062839 | -0.05846 |
| GOSR2 | ENSG00000108433 | 17:46941601-46941606 | + | TE | 0.075627 | 0.012909 | 0.062719 |
| GOSR2 | ENSG00000108433 | 17:46932200-46933245 | + | TE | 0.08384 | 0.017686 | 0.066154 |
| GPBAR1 | ENSG00000179921 | 2:218262680-218263848 | + | TE | 0.007543 | 0.13393 | -0.12639 |
| GPER1 | ENSG00000164850 | 7:1086807-1086875 | + | TS | 0.87687 | 0.11952 | 0.75735 |
| GPER1 | ENSG00000164850 | 7:1086876-1087244 | + | TS | 0.12709 | 0.86727 | -0.74018 |
| GPI | ENSG00000105220 | 19:34400333-34400335 | + | TE | 0.004529 | 0.02472 | -0.02019 |
| GPI | ENSG00000105220 | 19:34400328-34400328 | + | TE | 0.00894 | 0.064015 | -0.05508 |
| GPR1 | ENSG00000183671 | 2:206176180-206176835 | - | TE | 0.71142 | 0.155 | 0.55642 |
| GPR171 | ENSG00000174946 | 3:151198427-151199386 | - | TE | 0.001249 | 0.025471 | -0.02422 |
| GPR18 | ENSG00000125245 | 13:99254732-99255578 | - | TE | 0.032201 | 0.17868 | -0.14648 |
| GPR18 | ENSG00000125245 | 13:99255579-99255906 | - | TE | 0.024573 | 0.003233 | 0.021339 |
| GPR34 | ENSG00000171659 | X:41697274-41697277 | + | TE | 0.024792 | 0.17069 | -0.1459 |
| GPR85 | ENSG00000164604 | 7:113084731-113084891 | - | TE | 0.46818 | 0.10785 | 0.36033 |
| GPR85 | ENSG00000164604 | 7:113083609-113084508 | - | TE | 0.10332 | 0.42017 | -0.31685 |
| GPRC5C | ENSG00000170412 | 17:74447428-74447429 | + | TE | 0.51954 | 0.03505 | 0.48449 |
| GPS1 | ENSG00000169727 | 17:82057425-82057463 | + | TE | 0.067421 | 0.015999 | 0.051422 |
| GPX4 | ENSG00000167468 | 19:1106789-1106790 | + | TE | 0.022551 | 0.003871 | 0.018681 |
| GRAMD1C | ENSG00000178075 | 3:113945561-113945760 | + | TE | 0.025545 | 0.17479 | -0.14925 |
| GRK2 | ENSG00000173020 | 11:67286056-67286372 | + | TE | 0.041016 | 0.001169 | 0.039846 |
| GSDME | ENSG00000105928 | 7:24698351-24698354 | - | TE | 0.18096 | 0.043782 | 0.13718 |
| GSG1 | ENSG00000111305 | 12:13084825-13085243 | - | TE | 0.099439 | 0.72306 | -0.62362 |
| GSG1 | ENSG00000111305 | 12:13084577-13084586 | - | TE | 0.49934 | 0.11752 | 0.38183 |
| GSTO2 | ENSG00000065621 | 10:104299128-104299411 | + | TE | 0.001017 | 0.04433 | -0.04331 |
| GTF2I | ENSG00000263001 | 7:74759960-74760690 | + | TE | 0.063695 | 0.0099 | 0.053795 |
| GTF3C5 | ENSG00000148308 | 9:133058502-133058503 | + | TE | 0.009585 | 0.4705 | -0.46091 |
| GTPBP6 | ENSG00000178605 | X:304750-305197 | - | TE | 0.051129 | 0.012369 | 0.038761 |
| GTPBP8 | ENSG00000163607 | 3:112990987-112990990 | + | TS | 0.33907 | 0.047966 | 0.2911 |
| GYPA | ENSG00000170180 | 4:144140673-144140674 | - | TS | 0.050442 | 0.4961 | -0.44566 |
| H2AFX | ENSG00000188486 | 11:119093854-119093878 | - | TE | 0.015153 | 0.07828 | -0.06313 |
| H3F3B | ENSG00000132475 | 17:75783107-75785893 | - | TS | 0.00549 | 0.03968 | -0.03419 |
| HACE1 | ENSG00000085382 | 6:104729170-104729317 | - | TE | 0.013731 | 0.099022 | -0.08529 |
| HADHA | ENSG00000084754 | 2:26190889-26190921 | - | TE | 0.011262 | 0.077558 | -0.0663 |
| HARS | ENSG00000170445 | 5:140673920-140673931 | - | TE | 0.037203 | 0.16254 | -0.12534 |
| HARS2 | ENSG00000112855 | 5:140699306-140699318 | + | TE | 0.062114 | 0.010652 | 0.051462 |
| HARS2 | ENSG00000112855 | 5:140699236-140699248 | + | TE | 0.10348 | 0.010879 | 0.092603 |
| HAUS1 | ENSG00000152240 | 18:46128331-46128333 | + | TE | 0.47613 | 0.10068 | 0.37546 |
| HBS1L | ENSG00000112339 | 6:134964633-134964841 | - | TE | 0.007742 | 0.042973 | -0.03523 |
| HDAC8 | ENSG00000147099 | X:72457402-72457405 | - | TE | 0.15334 | 0.019315 | 0.13402 |
| HDAC8 | ENSG00000147099 | X:72474535-72474729 | - | TE | 0.084571 | 0.41632 | -0.33175 |
| HDAC9 | ENSG00000048052 | 7:18666980-18667105 | + | TE | 0.012379 | 0.05141 | -0.03903 |
| HDAC9 | ENSG00000048052 | 7:18668838-18668843 | + | TE | 0.027762 | 0.11784 | -0.09008 |
| HDLBP | ENSG00000115677 | 2:241229483-241229687 | - | TE | 0.16195 | 0.039184 | 0.12277 |
| HECW2 | ENSG00000138411 | 2:196199275-196199336 | - | TE | 0.017937 | 0.002626 | 0.015312 |
| HELB | ENSG00000127311 | 12:66343456-66343643 | + | TE | 0.014939 | 0.001262 | 0.013677 |
| HEMK1 | ENSG00000114735 | 3:50580674-50584748 | + | TE | 0.00066 | 0.026681 | -0.02602 |
| HENMT1 | ENSG00000162639 | 1:108648290-108648294 | - | TE | 0.16617 | 0.029393 | 0.13677 |
| HEPH | ENSG00000089472 | X:66267347-66267383 | + | TE | 0.061159 | 0.62765 | -0.56649 |
| HERC2 | ENSG00000128731 | 15:28111079-28111278 | - | TE | 0.090646 | 0.51232 | -0.42167 |
| HERC5 | ENSG00000138646 | 4:88506023-88506161 | + | TE | 0.012074 | 0.24671 | -0.23464 |
| HERPUD1 | ENSG00000051108 | 16:56932142-56932146 | + | TS | 0.014758 | 0.19039 | -0.17563 |
| HES4 | ENSG00000188290 | 1:1000097-1000111 | - | TS | 0.19303 | 0.020833 | 0.17219 |
| HESX1 | ENSG00000163666 | 3:57197843-57198295 | - | TE | 0.083437 | 0.35547 | -0.27203 |
| HGH1 | ENSG00000235173 | 8:144140714-144140724 | + | TE | 0.14489 | 0.012216 | 0.13268 |
| HIC1 | ENSG00000177374 | 17:2056890-2056943 | + | TE | 0.057856 | 0.004572 | 0.053284 |
| HIP1R | ENSG00000130787 | 12:122857216-122857384 | + | TE | 0.004306 | 0.020739 | -0.01643 |
| HLA-DOB | ENSG00000241106 | 6:32812767-32813046 | - | TE | 0.15446 | 0.020935 | 0.13353 |
| HMGB2 | ENSG00000164104 | 4:173331705-173332238 | - | TE | 0.005523 | 0.12309 | -0.11757 |
| HMGB3 | ENSG00000029993 | X:150987893-150987932 | + | TE | 0.12946 | 0.02568 | 0.10378 |
| HMMR | ENSG00000072571 | 5:163491112-163491238 | + | TE | 0.030293 | 0.12474 | -0.09444 |
| HNRNPA3 | ENSG00000170144 | 2:177219408-177219806 | + | TE | 0.000225 | 0.03639 | -0.03617 |
| HNRNPC | ENSG00000092199 | 14:21210550-21210555 | - | TE | 0.001745 | 0.000347 | 0.001398 |
| HNRNPD | ENSG00000138668 | 4:82355249-82355303 | - | TE | 0.005101 | 0.052062 | -0.04696 |
| HNRNPF | ENSG00000169813 | 10:43386022-43387588 | - | TE | 0.011896 | 0.079209 | -0.06731 |
| HNRNPH1 | ENSG00000169045 | 5:179623636-179623646 | - | TS | 0.30303 | 0.053258 | 0.24977 |
| HNRNPLL | ENSG00000143889 | 2:38564032-38564237 | - | TE | 0.001517 | 0.016963 | -0.01545 |
| HNRNPU | ENSG00000153187 | 1:244853418-244853466 | - | TE | 0.000452 | 0.001856 | -0.0014 |
| HNRNPU | ENSG00000153187 | 1:244864538-244864538 | - | TS | 0.077347 | 0.017499 | 0.059847 |
| HNRNPUL1 | ENSG00000105323 | 19:41307039-41307050 | + | TE | 0.014337 | 0.001307 | 0.01303 |
| HOOK1 | ENSG00000134709 | 1:59872795-59872978 | + | TE | 0.028888 | 0.22461 | -0.19572 |
| HOPX | ENSG00000171476 | 4:56647998-56647999 | - | TE | 0.07861 | 0.011758 | 0.066852 |
| HOXA10 | ENSG00000253293 | 7:27171220-27172173 | - | TE | 0.27525 | 0.064053 | 0.2112 |
| HOXA3 | ENSG00000105997 | 7:27106184-27106189 | - | TE | 0.45387 | 0.06078 | 0.39309 |
| HPGD | ENSG00000164120 | 4:174491870-174492094 | - | TE | 0.005653 | 0.067854 | -0.0622 |
| HPGD | ENSG00000164120 | 4:174491755-174491784 | - | TE | 0.056912 | 0.008645 | 0.048267 |
| HPS1 | ENSG00000107521 | 10:98446911-98446913 | - | TS | 0.37584 | 0.08833 | 0.28751 |
| HPS5 | ENSG00000110756 | 11:18279059-18279942 | - | TE | 0.005961 | 0.054519 | -0.04856 |
| HPSE | ENSG00000173083 | 4:83295320-83295343 | - | TE | 0.000834 | 0.023996 | -0.02316 |
| HPSE | ENSG00000173083 | 4:83295344-83295503 | - | TE | 0.000848 | 0.034934 | -0.03409 |
| HRAS | ENSG00000174775 | 11:532246-532375 | - | TE | 0.11874 | 0.86398 | -0.74524 |
| HRAS | ENSG00000174775 | 11:532243-532245 | - | TE | 0.87685 | 0.14071 | 0.73614 |
| HRASLS5 | ENSG00000168004 | 11:63463260-63463336 | - | TE | 0.089927 | 0.46534 | -0.37542 |
| HS6ST1 | ENSG00000136720 | 2:128268690-128268870 | - | TE | 0.17716 | 0.004005 | 0.17316 |
| HSD17B1 | ENSG00000108786 | 17:42552923-42552933 | + | TS | 0.13977 | 0.00297 | 0.1368 |
| HSD17B12 | ENSG00000149084 | 11:43856405-43856610 | + | TE | 0.002124 | 0.010467 | -0.00834 |
| HSD3B7 | ENSG00000099377 | 16:30989147-30989152 | + | TE | 0.011958 | 0.26498 | -0.25302 |
| HSF1 | ENSG00000185122 | 8:144314717-144314719 | + | TE | 0.42346 | 0.006134 | 0.41732 |
| HSF2 | ENSG00000025156 | 6:122431993-122432966 | + | TE | 0.011566 | 0.15938 | -0.14782 |
| HSPA4L | ENSG00000164070 | 4:127832969-127833042 | + | TE | 0.094845 | 0.014287 | 0.080559 |
| HSPE1 | ENSG00000115541 | 2:197500140-197500348 | + | TS | 0.49495 | 0.12052 | 0.37443 |
| HTATIP2 | ENSG00000109854 | 11:20383369-20383778 | + | TE | 0.006628 | 0.059634 | -0.05301 |
| IBA57 | ENSG00000181873 | 1:228175122-228175575 | + | TE | 0.06372 | 0.006549 | 0.057171 |
| ICAM4 | ENSG00000105371 | 19:10288503-10288520 | + | TE | 0.14083 | 0.88638 | -0.74555 |
| ICAM4 | ENSG00000105371 | 19:10287986-10288502 | + | TE | 0.85267 | 0.1102 | 0.74248 |
| IDE | ENSG00000119912 | 10:92454269-92454539 | - | TE | 0.050384 | 0.002754 | 0.04763 |
| IFI16 | ENSG00000163565 | 1:159054821-159055148 | + | TE | 0.003453 | 0.020991 | -0.01754 |
| IFI27 | ENSG00000165949 | 14:94116442-94116447 | + | TE | 0.005963 | 0.02884 | -0.02288 |
| IFT22 | ENSG00000128581 | 7:101315072-101315282 | - | TE | 0.082351 | 0.64669 | -0.56434 |
| IFT80 | ENSG00000068885 | 3:160258403-160258635 | - | TE | 0.016585 | 0.10358 | -0.08699 |
| IFT88 | ENSG00000032742 | 13:20691363-20691437 | + | TE | 0.50468 | 0.12151 | 0.38317 |
| IGF1 | ENSG00000017427 | 12:102402328-102402458 | - | TE | 0.044352 | 0.25322 | -0.20887 |
| IGF2 | ENSG00000167244 | 11:2132716-2133223 | - | TE | 0.019197 | 0.22505 | -0.20585 |
| IGF2BP2 | ENSG00000073792 | 3:185643975-185645500 | - | TE | 0.11132 | 0.004356 | 0.10697 |
| IGFBP2 | ENSG00000115457 | 2:216633429-216633965 | + | TS | 0.052421 | 0.26426 | -0.21184 |
| IGFLR1 | ENSG00000126246 | 19:35739528-35739626 | - | TE | 0.1234 | 0.01896 | 0.10444 |
| IKZF1 | ENSG00000185811 | 7:50308680-50308720 | + | TS | 0.12627 | 0.79465 | -0.66838 |
| IKZF1 | ENSG00000185811 | 7:50308721-50308747 | + | TS | 0.87492 | 0.20714 | 0.66778 |
| IKZF3 | ENSG00000161405 | 17:39765284-39765495 | - | TE | 0.000295 | 0.004146 | -0.00385 |
| IL15 | ENSG00000164136 | 4:141733436-141733458 | + | TE | 0.051144 | 0.22796 | -0.17681 |
| IL15 | ENSG00000164136 | 4:141733986-141733987 | + | TE | 0.35667 | 0.086425 | 0.27024 |
| IL15RA | ENSG00000134470 | 10:5978180-5978187 | - | TS | 0.11184 | 0.60832 | -0.49648 |
| IL2RA | ENSG00000134460 | 10:6062092-6062309 | - | TS | 0.10348 | 0.54184 | -0.43836 |
| IL7 | ENSG00000104432 | 8:78733665-78733828 | - | TE | 0.30829 | 0.047882 | 0.26041 |
| ILK | ENSG00000166333 | 11:6610462-6610611 | + | TE | 0.10792 | 0.004804 | 0.10312 |
| ILKAP | ENSG00000132323 | 2:238203684-238203729 | - | TS | 0.091619 | 0.61083 | -0.51921 |
| IMMT | ENSG00000132305 | 2:86143940-86143948 | - | TE | 0.05285 | 0.26024 | -0.20739 |
| IMPA1 | ENSG00000133731 | 8:81659262-81659466 | - | TE | 0.001675 | 0.013233 | -0.01156 |
| INIP | ENSG00000148153 | 9:112687117-112687144 | - | TE | 0.034222 | 0.002522 | 0.031701 |
| INO80E | ENSG00000169592 | 16:29996274-29996391 | + | TS | 0.071614 | 0.43551 | -0.36389 |
| INPP5F | ENSG00000198825 | 10:119828361-119828620 | + | TE | 0.020767 | 0.14742 | -0.12665 |
| INPP5F | ENSG00000198825 | 10:119818727-119819539 | + | TS | 0.10519 | 0.7368 | -0.63161 |
| INPPL1 | ENSG00000165458 | 11:72237206-72237659 | + | TS | 0.002979 | 0.12728 | -0.1243 |
| INSL3 | ENSG00000248099 | 19:17821524-17821574 | - | TS | 0.50506 | 0.10954 | 0.39552 |
| INTS11 | ENSG00000127054 | 1:1311602-1311605 | - | TE | 0.29437 | 0.063258 | 0.23111 |
| INTS2 | ENSG00000108506 | 17:61865370-61865389 | - | TE | 0.007439 | 0.055597 | -0.04816 |
| INTS3 | ENSG00000143624 | 1:153773193-153773319 | + | TE | 0.099974 | 0.005302 | 0.094672 |
| INTS3 | ENSG00000143624 | 1:153773536-153773728 | + | TE | 0.017854 | 0.16645 | -0.1486 |
| INTS6L | ENSG00000165359 | X:135520643-135520658 | + | TS | 0.041381 | 0.43579 | -0.39441 |
| INTS6L | ENSG00000165359 | X:135582241-135582493 | + | TE | 0.52928 | 0.12668 | 0.40261 |
| IP6K2 | ENSG00000068745 | 3:48693456-48693487 | - | TE | 0.0375 | 0.003983 | 0.033517 |
| IP6K2 | ENSG00000068745 | 3:48694051-48694240 | - | TE | 0.017471 | 0.004018 | 0.013452 |
| IPO11 | ENSG00000086200 | 5:62628578-62628582 | + | TE | 0.1547 | 0.01706 | 0.13764 |
| IQCB1 | ENSG00000173226 | 3:121769763-121769993 | - | TE | 0.07417 | 0.30812 | -0.23395 |
| IQCC | ENSG00000160051 | 1:32208683-32208687 | + | TE | 0.010755 | 0.056315 | -0.04556 |
| IQGAP1 | ENSG00000140575 | 15:90499995-90501389 | + | TE | 0.000146 | 0.062276 | -0.06213 |
| IQSEC1 | ENSG00000144711 | 3:12900753-12900848 | - | TE | 0.000351 | 0.020592 | -0.02024 |
| IQSEC2 | ENSG00000124313 | X:53225831-53227865 | - | TE | 0.098062 | 0.85395 | -0.75589 |
| IQSEC2 | ENSG00000124313 | X:53225828-53225830 | - | TE | 0.90659 | 0.14448 | 0.76211 |
| IRAK1 | ENSG00000184216 | X:154010500-154010505 | - | TE | 0.004152 | 0.23214 | -0.22798 |
| IRF3 | ENSG00000126456 | 19:49659579-49659833 | - | TE | 0.22441 | 0.016547 | 0.20787 |
| IRF9 | ENSG00000213928 | 14:24166122-24166254 | + | TE | 0.001734 | 0.11041 | -0.10868 |
| IRX3 | ENSG00000177508 | 16:54283304-54283435 | - | TE | 0.081121 | 0.40928 | -0.32816 |
| ISCU | ENSG00000136003 | 12:108567269-108567444 | + | TE | 0.18243 | 0.039088 | 0.14334 |
| ISG20 | ENSG00000172183 | 15:88646912-88647819 | + | TS | 0.010192 | 0.001871 | 0.008321 |
| ISOC2 | ENSG00000063241 | 19:55461625-55461626 | - | TS | 0.49297 | 0.084634 | 0.40833 |
| ISYNA1 | ENSG00000105655 | 19:18434815-18434820 | - | TE | 0.054281 | 0.39075 | -0.33646 |
| ITGA2 | ENSG00000164171 | 5:53090654-53090657 | + | TE | 0.079775 | 0.010363 | 0.069411 |
| ITGA2 | ENSG00000164171 | 5:53090647-53090653 | + | TE | 0.079372 | 0.010525 | 0.068847 |
| ITGA4 | ENSG00000115232 | 2:181457202-181457206 | + | TS | 0.061711 | 0.004418 | 0.057294 |
| ITGAV | ENSG00000138448 | 2:186590065-186590079 | + | TS | 0.54729 | 0.13524 | 0.41205 |
| ITGB4 | ENSG00000132470 | 17:75721509-75721563 | + | TS | 0.8593 | 0.1434 | 0.7159 |
| ITGB4 | ENSG00000132470 | 17:75721497-75721508 | + | TS | 0.14231 | 0.85214 | -0.70983 |
| ITGB8 | ENSG00000105855 | 7:20409875-20410330 | + | TE | 0.038133 | 0.28114 | -0.243 |
| ITK | ENSG00000113263 | 5:157180840-157180842 | + | TS | 0.005173 | 0.021221 | -0.01605 |
| ITPR1 | ENSG00000150995 | 3:4847249-4847256 | + | TE | 0.077011 | 0.00832 | 0.068691 |
| ITPR1 | ENSG00000150995 | 3:4847349-4847353 | + | TE | 0.025882 | 0.18512 | -0.15924 |
| ITPR1 | ENSG00000150995 | 3:4847327-4847348 | + | TE | 0.048111 | 0.00909 | 0.039021 |
| ITPR1 | ENSG00000150995 | 3:4847452-4847498 | + | TE | 0.045024 | 0.008265 | 0.036759 |
| ITPR1 | ENSG00000150995 | 3:4847435-4847451 | + | TE | 0.066297 | 0.015143 | 0.051153 |
| ITPR1 | ENSG00000150995 | 3:4847287-4847293 | + | TE | 0.013655 | 0.11166 | -0.098 |
| ITPR1 | ENSG00000150995 | 3:4847373-4847380 | + | TE | 0.070935 | 0.008346 | 0.062589 |
| ITPR1 | ENSG00000150995 | 3:4847104-4847154 | + | TE | 0.06566 | 0.008288 | 0.057372 |
| IWS1 | ENSG00000163166 | 2:127505040-127505190 | - | TE | 0.025219 | 0.005044 | 0.020175 |
| JAKMIP1 | ENSG00000152969 | 4:6200253-6200555 | - | TS | 0.1226 | 0.79348 | -0.67088 |
| JAKMIP1 | ENSG00000152969 | 4:6200556-6200591 | - | TS | 0.8784 | 0.21016 | 0.66824 |
| JAM2 | ENSG00000154721 | 21:25714890-25717552 | + | TE | 0.012422 | 0.13996 | -0.12754 |
| JAM3 | ENSG00000166086 | 11:134151752-134152001 | + | TE | 0.042446 | 0.17452 | -0.13208 |
| JAZF1 | ENSG00000153814 | 7:27830593-27832976 | - | TE | 0.001173 | 0.054171 | -0.053 |
| JMJD4 | ENSG00000081692 | 1:227731189-227731192 | - | TE | 0.037155 | 0.007656 | 0.029498 |
| JMJD8 | ENSG00000161999 | 16:681703-682073 | - | TE | 0.091975 | 0.016084 | 0.07589 |
| JPT1 | ENSG00000189159 | 17:75135286-75135306 | - | TE | 0.020502 | 0.004468 | 0.016034 |
| JPT2 | ENSG00000206053 | 16:1698811-1698976 | + | TE | 0.015471 | 0.002789 | 0.012682 |
| KAT6B | ENSG00000156650 | 10:75022134-75022137 | + | TE | 0.90913 | 0.12143 | 0.7877 |
| KAT6B | ENSG00000156650 | 10:75021968-75021987 | + | TE | 0.088418 | 0.8847 | -0.79628 |
| KAT6B | ENSG00000156650 | 10:75032222-75032237 | + | TE | 0.030603 | 0.003278 | 0.027325 |
| KAT6B | ENSG00000156650 | 10:74843479-74844982 | + | TE | 0.054318 | 0.006142 | 0.048176 |
| KCND3 | ENSG00000171385 | 1:111775809-111776278 | - | TE | 0.11876 | 0.021905 | 0.096854 |
| KCNMA1 | ENSG00000156113 | 10:77637948-77637993 | - | TS | 0.48886 | 0.11549 | 0.37336 |
| KCNMA1 | ENSG00000156113 | 10:77637807-77637819 | - | TS | 0.15555 | 0.031058 | 0.12449 |
| KCNMA1 | ENSG00000156113 | 10:77636199-77636515 | - | TE | 0.23392 | 0.025897 | 0.20802 |
| KCNQ3 | ENSG00000184156 | 8:132120858-132126540 | - | TE | 0.19566 | 0.8153 | -0.61963 |
| KCNQ3 | ENSG00000184156 | 8:132129145-132129405 | - | TE | 0.4683 | 0.083292 | 0.38501 |
| KCTD10 | ENSG00000110906 | 12:109449961-109449970 | - | TE | 0.081004 | 0.002816 | 0.078188 |
| KDM1B | ENSG00000165097 | 6:18223849-18223853 | + | TE | 0.37618 | 0.079346 | 0.29684 |
| KDM2A | ENSG00000173120 | 11:67254874-67255062 | + | TE | 0.023882 | 0.000408 | 0.023474 |
| KDM2B | ENSG00000089094 | 12:121429509-121429776 | - | TE | 0.023376 | 0.003405 | 0.01997 |
| KDM5A | ENSG00000073614 | 12:285456-285662 | - | TE | 0.006653 | 0.000545 | 0.006108 |
| KDM5B | ENSG00000117139 | 1:202727417-202727426 | - | TE | 0.002212 | 0.035706 | -0.03349 |
| KDM5D | ENSG00000012817 | Y:19705417-19705419 | - | TE | 0.019692 | 0.001171 | 0.018521 |
| KDM5D | ENSG00000012817 | Y:19705415-19705416 | - | TE | 0.020833 | 0.00118 | 0.019653 |
| KDM5D | ENSG00000012817 | Y:19705420-19705424 | - | TE | 0.019633 | 0.001277 | 0.018356 |
| KDM8 | ENSG00000155666 | 16:27220860-27221084 | + | TE | 0.18533 | 0.035246 | 0.15008 |
| KIAA0895L | ENSG00000196123 | 16:67183262-67183941 | - | TS | 0.034116 | 0.14772 | -0.1136 |
| KIAA0930 | ENSG00000100364 | 22:45196749-45197216 | - | TE | 0.095258 | 0.004975 | 0.090284 |
| KIAA1586 | ENSG00000168116 | 6:57055226-57055239 | + | TE | 0.007218 | 0.13048 | -0.12326 |
| KIF21A | ENSG00000139116 | 12:39294387-39294420 | - | TE | 0.071954 | 0.31778 | -0.24582 |
| KLC4 | ENSG00000137171 | 6:43059857-43059882 | + | TS | 0.093358 | 0.42168 | -0.32832 |
| KLF10 | ENSG00000155090 | 8:102648786-102650391 | - | TE | 0.42979 | 0.008845 | 0.42095 |
| KLF6 | ENSG00000067082 | 10:3779047-3779549 | - | TE | 0.000183 | 0.013869 | -0.01369 |
| KLF8 | ENSG00000102349 | X:56232437-56232473 | + | TS | 0.13787 | 0.67943 | -0.54156 |
| KLHDC1 | ENSG00000197776 | 14:49751848-49751852 | + | TE | 0.022842 | 0.10095 | -0.07811 |
| KLHDC4 | ENSG00000104731 | 16:87707813-87707815 | - | TE | 0.85922 | 0.1022 | 0.75702 |
| KLHDC4 | ENSG00000104731 | 16:87707816-87708072 | - | TE | 0.14455 | 0.89601 | -0.75145 |
| KLHL20 | ENSG00000076321 | 1:173733713-173733828 | + | TE | 0.080708 | 0.006905 | 0.073803 |
| KLHL21 | ENSG00000162413 | 1:6593381-6593658 | - | TE | 0.000624 | 0.010716 | -0.01009 |
| KLHL28 | ENSG00000179454 | 14:44928813-44929191 | - | TE | 0.001721 | 0.011626 | -0.00991 |
| KLHL35 | ENSG00000149243 | 11:75430192-75430629 | - | TS | 0.79748 | 0.10903 | 0.68845 |
| KLHL5 | ENSG00000109790 | 4:39121010-39121205 | + | TE | 0.03711 | 0.005051 | 0.032058 |
| KLHL7 | ENSG00000122550 | 7:23174015-23174192 | + | TE | 0.077237 | 0.015855 | 0.061382 |
| KLHL8 | ENSG00000145332 | 4:87161043-87162443 | - | TE | 0.001172 | 0.01961 | -0.01844 |
| KMT2E | ENSG00000005483 | 7:105014190-105014207 | + | TS | 0.66072 | 0.11315 | 0.54757 |
| KRBA2 | ENSG00000184619 | 17:8376668-8376681 | - | TS | 0.38732 | 0.059669 | 0.32766 |
| KRIT1 | ENSG00000001631 | 7:92200178-92200357 | - | TE | 0.01558 | 0.09505 | -0.07947 |
| KRT72 | ENSG00000170486 | 12:52585589-52585713 | - | TE | 0.50503 | 0.087516 | 0.41752 |
| KRT73 | ENSG00000186049 | 12:52607736-52608452 | - | TE | 0.031241 | 0.58215 | -0.5509 |
| L2HGDH | ENSG00000087299 | 14:50312011-50312168 | - | TS | 0.89604 | 0.20715 | 0.68889 |
| L2HGDH | ENSG00000087299 | 14:50312230-50312548 | - | TS | 0.10742 | 0.79619 | -0.68877 |
| L3MBTL1 | ENSG00000185513 | 20:43550942-43550950 | + | TE | 0.016105 | 0.25467 | -0.23856 |
| L3MBTL1 | ENSG00000185513 | 20:43550911-43550927 | + | TE | 0.055902 | 0.010221 | 0.045681 |
| LAMA5 | ENSG00000130702 | 20:62309065-62309144 | - | TE | 0.058773 | 0.67765 | -0.61888 |
| LAMP3 | ENSG00000078081 | 3:183123925-183124214 | - | TE | 0.056311 | 0.34892 | -0.29261 |
| LAMTOR5 | ENSG00000134248 | 1:110401253-110401256 | - | TE | 0.0278 | 0.003284 | 0.024516 |
| LARGE1 | ENSG00000133424 | 22:33273089-33274624 | - | TE | 0.46553 | 0.05113 | 0.4144 |
| LARGE1 | ENSG00000133424 | 22:33919995-33920414 | - | TS | 0.092844 | 0.51273 | -0.41989 |
| LAT2 | ENSG00000086730 | 7:74228944-74229072 | + | TE | 0.000834 | 0.00659 | -0.00576 |
| LBH | ENSG00000213626 | 2:30257671-30257700 | + | TE | 0.00353 | 0.000665 | 0.002865 |
| LBX2 | ENSG00000179528 | 2:74497517-74497581 | - | TE | 0.14307 | 0.65937 | -0.51629 |
| LCMT1 | ENSG00000205629 | 16:25111932-25111996 | + | TS | 0.033536 | 0.1563 | -0.12277 |
| LCN10 | ENSG00000187922 | 9:136738167-136739460 | - | TE | 0.059342 | 0.4966 | -0.43726 |
| LCORL | ENSG00000178177 | 4:18021598-18021757 | - | TS | 0.14147 | 0.87684 | -0.73538 |
| LCORL | ENSG00000178177 | 4:18021758-18021876 | - | TS | 0.86094 | 0.12664 | 0.7343 |
| LDLRAD4 | ENSG00000168675 | 18:13645660-13645712 | + | TE | 0.00683 | 0.001468 | 0.005361 |
| LEPR | ENSG00000116678 | 1:65635410-65635428 | + | TE | 0.25623 | 0.013516 | 0.24272 |
| LGALS3BP | ENSG00000108679 | 17:78971238-78971255 | - | TE | 0.061299 | 0.002747 | 0.058551 |
| LGALS3BP | ENSG00000108679 | 17:78972470-78972704 | - | TE | 0.001424 | 0.006103 | -0.00468 |
| LGALS9C | ENSG00000171916 | 17:18489481-18489541 | + | TS | 0.29385 | 0.023293 | 0.27055 |
| LGR6 | ENSG00000133067 | 1:202319691-202319781 | + | TE | 0.064369 | 0.34497 | -0.2806 |
| LGR6 | ENSG00000133067 | 1:202317952-202319207 | + | TE | 0.25568 | 0.057607 | 0.19807 |
| LIG3 | ENSG00000005156 | 17:35004273-35004804 | + | TE | 0.047722 | 0.004899 | 0.042823 |
| LIG4 | ENSG00000174405 | 13:108207439-108207445 | - | TE | 0.05349 | 0.33818 | -0.28469 |
| LILRB2 | ENSG00000131042 | 19:54280961-54281097 | - | TS | 0.14693 | 0.018255 | 0.12868 |
| LILRB5 | ENSG00000105609 | 19:54250496-54250545 | - | TE | 0.098671 | 0.40757 | -0.3089 |
| LIMCH1 | ENSG00000064042 | 4:41699021-41700043 | + | TE | 0.61446 | 0.14393 | 0.47053 |
| LIMK2 | ENSG00000182541 | 22:31248358-31248401 | + | TS | 0.085318 | 0.36786 | -0.28254 |
| LIN54 | ENSG00000189308 | 4:83010711-83010731 | - | TS | 0.13958 | 0.89119 | -0.75161 |
| LIN54 | ENSG00000189308 | 4:82925715-82927559 | - | TE | 0.020791 | 0.004909 | 0.015881 |
| LIN54 | ENSG00000189308 | 4:83010484-83010709 | - | TS | 0.85799 | 0.11077 | 0.74722 |
| LIPC | ENSG00000166035 | 15:58410569-58410571 | + | TS | 0.025105 | 0.25526 | -0.23015 |
| LIPT1 | ENSG00000144182 | 2:99161957-99162075 | + | TE | 0.13322 | 0.008869 | 0.12435 |
| LMBRD1 | ENSG00000168216 | 6:69675946-69675950 | - | TE | 0.002762 | 0.020681 | -0.01792 |
| LMBRD1 | ENSG00000168216 | 6:69675926-69675930 | - | TE | 0.15169 | 0.037576 | 0.11412 |
| LMF1 | ENSG00000103227 | 16:854501-854706 | - | TE | 0.17609 | 0.035372 | 0.14071 |
| LMF1 | ENSG00000103227 | 16:970961-970963 | - | TS | 0.13897 | 0.87225 | -0.73327 |
| LMF1 | ENSG00000103227 | 16:970964-970982 | - | TS | 0.86185 | 0.12645 | 0.7354 |
| LMLN | ENSG00000185621 | 3:198038698-198039011 | + | TE | 0.13216 | 0.019988 | 0.11218 |
| LMO4 | ENSG00000143013 | 1:87345016-87345408 | + | TE | 0.000916 | 0.003884 | -0.00297 |
| LNPK | ENSG00000144320 | 2:175923897-175929172 | - | TE | 0.001546 | 0.031929 | -0.03038 |
| LOXHD1 | ENSG00000167210 | 18:46477273-46477639 | - | TE | 0.021401 | 0.13363 | -0.11223 |
| LPAR1 | ENSG00000198121 | 9:111038044-111038085 | - | TS | 0.09271 | 0.75815 | -0.66544 |
| LPAR3 | ENSG00000171517 | 1:84813404-84814171 | - | TE | 0.092036 | 0.76497 | -0.67293 |
| LPAR6 | ENSG00000139679 | 13:48411741-48412205 | - | TE | 0.00252 | 0.019492 | -0.01697 |
| LPCAT1 | ENSG00000153395 | 5:1461423-1461428 | - | TE | 0.051006 | 0.24902 | -0.19801 |
| LPIN1 | ENSG00000134324 | 2:11826514-11826530 | + | TE | 0.033161 | 0.003704 | 0.029456 |
| LRCH3 | ENSG00000186001 | 3:197883541-197883772 | + | TE | 0.001723 | 0.062469 | -0.06075 |
| LRCH3 | ENSG00000186001 | 3:197883773-197883813 | + | TE | 0.014475 | 0.001518 | 0.012956 |
| LRP1 | ENSG00000123384 | 12:57128499-57128715 | + | TS | 0.44438 | 0.06439 | 0.37999 |
| LRPPRC | ENSG00000138095 | 2:43995977-43995978 | - | TS | 0.49951 | 0.10912 | 0.3904 |
| LRRC2 | ENSG00000163827 | 3:46518349-46519063 | - | TE | 0.29507 | 0.058503 | 0.23657 |
| LRRC37B | ENSG00000185158 | 17:32007988-32008132 | + | TS | 0.13918 | 0.85801 | -0.71883 |
| LRRC37B | ENSG00000185158 | 17:32007872-32007987 | + | TS | 0.85162 | 0.14266 | 0.70896 |
| LRRC6 | ENSG00000129295 | 8:132572201-132572424 | - | TE | 0.26459 | 0.036649 | 0.22794 |
| LRRIQ3 | ENSG00000162620 | 1:74179907-74180222 | - | TE | 0.11109 | 0.74781 | -0.63672 |
| LRRN3 | ENSG00000173114 | 7:111124933-111125451 | + | TE | 0.21498 | 0.012893 | 0.20209 |
| LSM14A | ENSG00000257103 | 19:34228097-34229279 | + | TE | 0.011368 | 0.002173 | 0.009196 |
| LTA4H | ENSG00000111144 | 12:96000828-96000829 | - | TE | 0.028835 | 0.006336 | 0.022499 |
| LTBP1 | ENSG00000049323 | 2:33398364-33399363 | + | TE | 0.04072 | 0.26465 | -0.22393 |
| LTF | ENSG00000012223 | 3:46435645-46436018 | - | TE | 0.034447 | 0.14436 | -0.10991 |
| LTK | ENSG00000062524 | 15:41503642-41503950 | - | TE | 0.6875 | 0.065613 | 0.62189 |
| LTK | ENSG00000062524 | 15:41503638-41503641 | - | TE | 0.10641 | 0.4585 | -0.35209 |
| LUC7L | ENSG00000007392 | 16:188969-188970 | - | TE | 0.038098 | 0.1556 | -0.1175 |
| LY6E | ENSG00000160932 | 8:143021566-143021640 | + | TE | 0.000128 | 0.00062 | -0.00049 |
| LY6E | ENSG00000160932 | 8:143022411-143022412 | + | TE | 0.000216 | 0.002386 | -0.00217 |
| LY6G5B | ENSG00000240053 | 6:31671864-31672462 | + | TE | 0.056605 | 0.004625 | 0.05198 |
| LYG1 | ENSG00000144214 | 2:99284238-99284256 | - | TE | 0.63599 | 0.10433 | 0.53166 |
| M1AP | ENSG00000159374 | 2:74557883-74557891 | - | TE | 0.020674 | 0.13725 | -0.11657 |
| MACF1 | ENSG00000127603 | 1:39484601-39486217 | + | TE | 0.005665 | 0.075389 | -0.06972 |
| MAF1 | ENSG00000179632 | 8:144107088-144107392 | + | TE | 0.001377 | 0.028203 | -0.02683 |
| MAK | ENSG00000111837 | 6:10762740-10764200 | - | TE | 0.077819 | 0.004604 | 0.073215 |
| MAMDC4 | ENSG00000177943 | 9:136860798-136860799 | + | TE | 0.88292 | 0.12079 | 0.76213 |
| MAMDC4 | ENSG00000177943 | 9:136860562-136860797 | + | TE | 0.11221 | 0.87847 | -0.76627 |
| MAML3 | ENSG00000196782 | 4:140152970-140154184 | - | TS | 0.002082 | 0.13742 | -0.13534 |
| MAML3 | ENSG00000196782 | 4:139720212-139720323 | - | TE | 0.11975 | 0.002155 | 0.1176 |
| MAN2A2 | ENSG00000196547 | 15:90921192-90922579 | + | TE | 0.000983 | 0.031249 | -0.03027 |
| MAN2B2 | ENSG00000013288 | 4:6621188-6621354 | + | TE | 0.11474 | 0.001868 | 0.11287 |
| MANBA | ENSG00000109323 | 4:102760973-102760982 | - | TS | 0.053664 | 0.54962 | -0.49595 |
| MAP2K5 | ENSG00000137764 | 15:67542709-67543330 | + | TS | 0.24441 | 0.007609 | 0.2368 |
| MAP2K6 | ENSG00000108984 | 17:69542012-69542017 | + | TE | 0.046044 | 0.000197 | 0.045847 |
| MAP3K11 | ENSG00000173327 | 11:65614237-65614242 | - | TS | 0.24225 | 0.004338 | 0.23792 |
| MAP3K12 | ENSG00000139625 | 12:53481121-53481121 | - | TE | 0.00264 | 0.083639 | -0.081 |
| MAP3K14 | ENSG00000006062 | 17:45263128-45263134 | - | TE | 0.007969 | 0.19949 | -0.19152 |
| MAP3K15 | ENSG00000180815 | X:19360058-19360059 | - | TE | 0.43749 | 0.038029 | 0.39946 |
| MAP3K3 | ENSG00000198909 | 17:63693961-63695089 | + | TE | 0.01243 | 0.001116 | 0.011315 |
| MAP3K3 | ENSG00000198909 | 17:63622710-63622716 | + | TS | 0.022738 | 0.26897 | -0.24623 |
| MAP3K4 | ENSG00000085511 | 6:161116850-161117308 | + | TE | 0.88568 | 0.12074 | 0.76493 |
| MAP3K4 | ENSG00000085511 | 6:160991727-160991783 | + | TS | 0.43411 | 0.088197 | 0.34592 |
| MAP3K4 | ENSG00000085511 | 6:161117309-161117384 | + | TE | 0.11393 | 0.86648 | -0.75255 |
| MAPK14 | ENSG00000112062 | 6:36108380-36108509 | + | TE | 0.012991 | 0.000111 | 0.012879 |
| MAPKAPK5 | ENSG00000089022 | 12:111892967-111893157 | + | TE | 0.13791 | 0.017255 | 0.12065 |
| MARK4 | ENSG00000007047 | 19:45303370-45303693 | + | TE | 0.010877 | 0.073483 | -0.06261 |
| MASTL | ENSG00000120539 | 10:27186920-27186924 | + | TE | 0.1706 | 0.042145 | 0.12845 |
| MAT2A | ENSG00000168906 | 2:85543035-85543218 | + | TE | 0.095973 | 0.015307 | 0.080666 |
| MATR3 | ENSG00000015479 | 5:139330438-139330441 | + | TE | 0.11301 | 0.003184 | 0.10983 |
| MAU2 | ENSG00000129933 | 19:19357756-19357760 | + | TE | 0.001134 | 0.005944 | -0.00481 |
| MBD1 | ENSG00000141644 | 18:50281460-50281466 | - | TS | 0.089005 | 0.49526 | -0.40626 |
| MBD1 | ENSG00000141644 | 18:50266886-50267516 | - | TE | 0.03162 | 0.2675 | -0.23588 |
| MBNL1 | ENSG00000152601 | 3:152463160-152465203 | + | TE | 0.000121 | 0.018378 | -0.01826 |
| MBNL1 | ENSG00000152601 | 3:152462385-152463159 | + | TE | 0.008853 | 0.042848 | -0.034 |
| MBOAT2 | ENSG00000143797 | 2:9003659-9003813 | - | TS | 0.018184 | 0.25207 | -0.23388 |
| MBP | ENSG00000197971 | 18:76978861-76979766 | - | TE | 0.021229 | 0.14072 | -0.11949 |
| MC1R | ENSG00000258839 | 16:89918851-89920951 | + | TE | 0.16609 | 0.75097 | -0.58489 |
| MCC | ENSG00000171444 | 5:113027302-113027482 | - | TE | 0.00052 | 0.061197 | -0.06068 |
| MCF2 | ENSG00000101977 | X:139582443-139582470 | - | TE | 0.48002 | 0.078999 | 0.40102 |
| MCM3 | ENSG00000112118 | 6:52264009-52264013 | - | TE | 0.018682 | 0.13704 | -0.11836 |
| MCMBP | ENSG00000197771 | 10:119831201-119831600 | - | TE | 0.001041 | 0.004583 | -0.00354 |
| MCOLN2 | ENSG00000153898 | 1:84996796-84996940 | - | TS | 0.85783 | 0.16231 | 0.69552 |
| MCOLN2 | ENSG00000153898 | 1:84996941-84997113 | - | TS | 0.13568 | 0.84257 | -0.70689 |
| MDGA1 | ENSG00000112139 | 6:37696915-37697990 | - | TS | 0.28193 | 0.047445 | 0.23448 |
| MDM2 | ENSG00000135679 | 12:68843934-68844190 | + | TE | 0.001032 | 0.070109 | -0.06908 |
| MEAF6 | ENSG00000163875 | 1:37493335-37493397 | - | TE | 0.002905 | 0.018187 | -0.01528 |
| MED12 | ENSG00000184634 | X:71142448-71142454 | + | TE | 0.01042 | 0.15534 | -0.14492 |
| MED14 | ENSG00000180182 | X:40649543-40649672 | - | TE | 0.14944 | 0.019014 | 0.13042 |
| MED16 | ENSG00000175221 | 19:867972-868251 | - | TE | 0.008628 | 0.077307 | -0.06868 |
| MEF2D | ENSG00000116604 | 1:156466218-156466498 | - | TE | 0.033554 | 0.001091 | 0.032464 |
| MEF2D | ENSG00000116604 | 1:156466210-156466217 | - | TE | 0.056323 | 0.001121 | 0.055202 |
| MEGF6 | ENSG00000162591 | 1:3489926-3490311 | - | TE | 0.061593 | 0.30851 | -0.24692 |
| MEMO1 | ENSG00000162959 | 2:31867827-31867977 | - | TE | 0.00135 | 0.03389 | -0.03254 |
| MEMO1 | ENSG00000162959 | 2:31867978-31868492 | - | TE | 0.001296 | 0.021553 | -0.02026 |
| MEOX1 | ENSG00000005102 | 17:43640388-43640401 | - | TE | 0.03613 | 0.20536 | -0.16923 |
| METRN | ENSG00000103260 | 16:717071-717390 | + | TE | 0.10618 | 0.021871 | 0.084306 |
| METTL8 | ENSG00000123600 | 2:171323933-171323997 | - | TE | 0.050653 | 0.004654 | 0.045999 |
| MFSD1 | ENSG00000118855 | 3:158822443-158823193 | + | TS | 0.083206 | 0.013562 | 0.069644 |
| MFSD1 | ENSG00000118855 | 3:158828979-158829116 | + | TE | 0.037983 | 0.36315 | -0.32517 |
| MFSD10 | ENSG00000109736 | 4:2930568-2930953 | - | TE | 0.059435 | 0.006886 | 0.052549 |
| MFSD14C | ENSG00000196312 | 9:97013581-97013600 | - | TS | 0.11059 | 0.47746 | -0.36686 |
| MFSD8 | ENSG00000164073 | 4:127917898-127917924 | - | TE | 0.3411 | 0.053211 | 0.28788 |
| MGLL | ENSG00000074416 | 3:127692132-127692138 | - | TE | 0.00254 | 0.054462 | -0.05192 |
| MGRN1 | ENSG00000102858 | 16:4688909-4690972 | + | TE | 0.11594 | 0.002413 | 0.11352 |
| MICAL2 | ENSG00000133816 | 11:12263560-12263781 | + | TE | 0.032073 | 0.14546 | -0.11339 |
| MICAL2 | ENSG00000133816 | 11:12263783-12263785 | + | TE | 0.031483 | 0.13693 | -0.10545 |
| MICAL2 | ENSG00000133816 | 11:12358295-12358479 | + | TE | 0.023299 | 0.11448 | -0.09118 |
| MINDY2 | ENSG00000128923 | 15:58771286-58771313 | + | TS | 0.03284 | 0.13882 | -0.10598 |
| MINOS1 | ENSG00000173436 | 1:19626607-19626623 | + | TE | 0.054845 | 0.27511 | -0.22026 |
| MINOS1 | ENSG00000173436 | 1:19626387-19626587 | + | TE | 0.017504 | 0.18791 | -0.17041 |
| MIS18BP1 | ENSG00000129534 | 14:45253035-45253177 | - | TS | 0.49568 | 0.095883 | 0.3998 |
| MITF | ENSG00000187098 | 3:69965249-69965647 | + | TE | 0.022743 | 0.13744 | -0.11469 |
| MKRN1 | ENSG00000133606 | 7:140456186-140456191 | - | TE | 0.083844 | 0.012332 | 0.071513 |
| MLH1 | ENSG00000076242 | 3:37050707-37050842 | + | TE | 0.07801 | 0.016464 | 0.061546 |
| MLX | ENSG00000108788 | 17:42571547-42571729 | + | TE | 0.014431 | 0.058968 | -0.04454 |
| MMAA | ENSG00000151611 | 4:145655569-145658424 | + | TE | 0.010483 | 0.11138 | -0.1009 |
| MMP17 | ENSG00000198598 | 12:131851772-131851783 | + | TE | 0.17651 | 0.029603 | 0.14691 |
| MOB4 | ENSG00000115540 | 2:197550515-197551012 | + | TE | 0.10067 | 0.010955 | 0.08972 |
| MOGS | ENSG00000115275 | 2:74461070-74461101 | - | TE | 0.038142 | 0.009101 | 0.029041 |
| MOSPD1 | ENSG00000101928 | X:134889015-134889192 | - | TE | 0.013929 | 0.10581 | -0.09188 |
| MOSPD2 | ENSG00000130150 | X:14873491-14873528 | + | TS | 0.087969 | 0.50018 | -0.41221 |
| MPDU1 | ENSG00000129255 | 17:7587426-7587643 | + | TE | 0.070708 | 0.01766 | 0.053047 |
| MPI | ENSG00000178802 | 15:74900484-74900689 | + | TE | 0.053911 | 0.007684 | 0.046227 |
| MPI | ENSG00000178802 | 15:74897512-74898077 | + | TE | 0.18933 | 0.024044 | 0.16528 |
| MPND | ENSG00000008382 | 19:4360082-4360086 | + | TE | 0.90265 | 0.08664 | 0.81601 |
| MPND | ENSG00000008382 | 19:4359916-4360068 | + | TE | 0.10053 | 0.46871 | -0.36818 |
| MPND | ENSG00000008382 | 19:4360069-4360081 | + | TE | 0.10119 | 0.53759 | -0.4364 |
| MPP2 | ENSG00000108852 | 17:43877762-43877983 | - | TE | 0.33405 | 0.081425 | 0.25262 |
| MPPE1 | ENSG00000154889 | 18:11908316-11908317 | - | TS | 0.012277 | 0.099417 | -0.08714 |
| MRO | ENSG00000134042 | 18:50798204-50798461 | - | TE | 0.07074 | 0.42068 | -0.34994 |
| MRPL43 | ENSG00000055950 | 10:100986276-100986279 | - | TE | 0.079845 | 0.55449 | -0.47464 |
| MRPL48 | ENSG00000175581 | 11:73864599-73864599 | + | TE | 0.25017 | 0.031963 | 0.2182 |
| MRPL48 | ENSG00000175581 | 11:73864600-73864611 | + | TE | 0.22699 | 0.032152 | 0.19484 |
| MRPL53 | ENSG00000204822 | 2:74471993-74472252 | - | TE | 0.15426 | 0.018028 | 0.13623 |
| MRPS2 | ENSG00000122140 | 9:135503977-135503986 | + | TE | 0.14152 | 0.034857 | 0.10667 |
| MRPS34 | ENSG00000074071 | 16:1772799-1773150 | - | TS | 0.020868 | 0.20659 | -0.18572 |
| MS4A1 | ENSG00000156738 | 11:60470753-60470760 | + | TE | 0.002399 | 0.015174 | -0.01278 |
| MS4A4A | ENSG00000110079 | 11:60308969-60308972 | + | TE | 0.079074 | 0.00601 | 0.073064 |
| MSL1 | ENSG00000188895 | 17:40122614-40122760 | + | TS | 0.064928 | 0.30014 | -0.23521 |
| MSL3 | ENSG00000005302 | X:11775754-11775760 | + | TE | 0.029305 | 0.004841 | 0.024464 |
| MSL3 | ENSG00000005302 | X:11775761-11775772 | + | TE | 0.004179 | 0.019655 | -0.01548 |
| MSR1 | ENSG00000038945 | 8:16110046-16110084 | - | TE | 0.10388 | 0.64493 | -0.54105 |
| MSRB3 | ENSG00000174099 | 12:65463155-65463376 | + | TE | 0.024817 | 0.005942 | 0.018874 |
| MST1 | ENSG00000173531 | 3:49684049-49684189 | - | TE | 0.50334 | 0.11365 | 0.3897 |
| MT1F | ENSG00000198417 | 16:56659304-56659305 | + | TE | 0.022703 | 0.095908 | -0.0732 |
| MTCH2 | ENSG00000109919 | 11:47618704-47618838 | - | TE | 0.008578 | 0.1115 | -0.10292 |
| MTFR2 | ENSG00000146410 | 6:136231024-136231029 | - | TE | 0.92971 | 0.099075 | 0.83064 |
| MTFR2 | ENSG00000146410 | 6:136231030-136231388 | - | TE | 0.071011 | 0.90329 | -0.83228 |
| MTHFD1 | ENSG00000100714 | 14:64460004-64460004 | + | TE | 0.24451 | 0.013348 | 0.23116 |
| MTHFD2L | ENSG00000163738 | 4:74160089-74160514 | + | TE | 0.23889 | 0.054726 | 0.18417 |
| MTMR10 | ENSG00000166912 | 15:30938948-30939524 | - | TE | 0.002873 | 0.000373 | 0.0025 |
| MTMR12 | ENSG00000150712 | 5:32226994-32227005 | - | TE | 0.00509 | 0.18639 | -0.1813 |
| MTMR14 | ENSG00000163719 | 3:9702217-9702387 | + | TE | 0.082792 | 0.005879 | 0.076914 |
| MTR | ENSG00000116984 | 1:236795310-236795737 | + | TS | 0.084598 | 0.3633 | -0.2787 |
| MTRF1 | ENSG00000120662 | 13:41252284-41252644 | - | TE | 0.14398 | 0.62622 | -0.48224 |
| MTSS1 | ENSG00000170873 | 8:124727884-124727998 | - | TS | 0.30205 | 0.04163 | 0.26042 |
| MTSS1 | ENSG00000170873 | 8:124550976-124552799 | - | TE | 0.001702 | 0.010475 | -0.00877 |
| MTURN | ENSG00000180354 | 7:30157970-30162760 | + | TE | 0.002661 | 0.000466 | 0.002195 |
| MVK | ENSG00000110921 | 12:109597237-109597261 | + | TE | 0.045502 | 0.23937 | -0.19386 |
| MXD1 | ENSG00000059728 | 2:69942943-69942945 | + | TE | 0.002048 | 0.066326 | -0.06428 |
| MXI1 | ENSG00000119950 | 10:110225993-110226083 | + | TS | 0.15142 | 0.009272 | 0.14214 |
| MYB | ENSG00000118513 | 6:135217981-135218006 | + | TE | 0.1841 | 0.042896 | 0.14121 |
| MYCBP2 | ENSG00000005810 | 13:77044655-77044657 | - | TE | 0.10533 | 0.006379 | 0.098952 |
| MYO7A | ENSG00000137474 | 11:77215240-77215241 | + | TE | 0.25038 | 0.060712 | 0.18967 |
| MYRIP | ENSG00000170011 | 3:40259966-40260313 | + | TE | 0.010892 | 0.10061 | -0.08972 |
| MZT2B | ENSG00000152082 | 2:130182313-130182414 | + | TS | 0.13758 | 0.88278 | -0.7452 |
| MZT2B | ENSG00000152082 | 2:130182415-130182452 | + | TS | 0.8598 | 0.12035 | 0.73945 |
| N4BP1 | ENSG00000102921 | 16:48543164-48543261 | - | TE | 0.077969 | 0.000494 | 0.077475 |
| NAA60 | ENSG00000122390 | 16:3486915-3486916 | + | TE | 0.079478 | 0.012786 | 0.066692 |
| NAA60 | ENSG00000122390 | 16:3486953-3486963 | + | TE | 0.077149 | 0.008996 | 0.068153 |
| NAA80 | ENSG00000243477 | 3:50296418-50296451 | - | TE | 0.076648 | 0.018302 | 0.058347 |
| NADSYN1 | ENSG00000172890 | 11:71498529-71498728 | + | TE | 0.003986 | 0.032616 | -0.02863 |
| NAGK | ENSG00000124357 | 2:71078621-71078635 | + | TE | 0.001862 | 0.043012 | -0.04115 |
| NCAPG | ENSG00000109805 | 4:17843302-17843448 | + | TE | 0.14291 | 0.010336 | 0.13257 |
| NCEH1 | ENSG00000144959 | 3:172633873-172633998 | - | TE | 0.1118 | 0.002622 | 0.10917 |
| NCF2 | ENSG00000116701 | 1:183555725-183556230 | - | TE | 0.12992 | 0.000292 | 0.12963 |
| NDUFA10 | ENSG00000130414 | 2:239960741-239961186 | - | TE | 0.076794 | 0.018831 | 0.057963 |
| NDUFA13 | ENSG00000186010 | 19:19516241-19516332 | + | TS | 0.13624 | 0.028453 | 0.10779 |
| NDUFAF2 | ENSG00000164182 | 5:61153027-61153037 | + | TE | 0.47983 | 0.07745 | 0.40238 |
| NDUFAF3 | ENSG00000178057 | 3:49023288-49023288 | + | TE | 0.19505 | 0.018963 | 0.17609 |
| NDUFB3 | ENSG00000119013 | 2:201085745-201085750 | + | TE | 0.2287 | 0.031995 | 0.1967 |
| NDUFB5 | ENSG00000136521 | 3:179624500-179624500 | + | TE | 0.002709 | 0.011902 | -0.00919 |
| NDUFB5 | ENSG00000136521 | 3:179624378-179624497 | + | TE | 0.011341 | 0.002561 | 0.00878 |
| NDUFB6 | ENSG00000165264 | 9:32573160-32573162 | - | TS | 0.030086 | 0.16443 | -0.13434 |
| NDUFS1 | ENSG00000023228 | 2:206124101-206124276 | - | TE | 0.000606 | 0.036314 | -0.03571 |
| NECAB1 | ENSG00000123119 | 8:90791550-90791740 | + | TS | 0.24789 | 0.046232 | 0.20165 |
| NECAB3 | ENSG00000125967 | 20:33657730-33657857 | - | TE | 0.13868 | 0.014665 | 0.12401 |
| NECAP1 | ENSG00000089818 | 12:8097731-8097735 | + | TE | 0.00182 | 0.012035 | -0.01021 |
| NECAP1 | ENSG00000089818 | 12:8097736-8097738 | + | TE | 0.030697 | 0.002573 | 0.028124 |
| NEDD4L | ENSG00000049759 | 18:58396552-58396573 | + | TE | 0.024172 | 0.14919 | -0.12502 |
| NEDD4L | ENSG00000049759 | 18:58396579-58396580 | + | TE | 0.14969 | 0.022987 | 0.1267 |
| NEDD4L | ENSG00000049759 | 18:58044548-58044708 | + | TS | 0.10697 | 0.025125 | 0.081841 |
| NEDD4L | ENSG00000049759 | 18:58044367-58044377 | + | TS | 0.10628 | 0.025146 | 0.081139 |
| NEFM | ENSG00000104722 | 8:24919094-24919098 | + | TE | 0.68119 | 0.084308 | 0.59688 |
| NELFA | ENSG00000185049 | 4:1982723-1982729 | - | TE | 0.025885 | 0.35629 | -0.3304 |
| NEXN | ENSG00000162614 | 1:77943225-77943891 | + | TE | 0.006777 | 0.078939 | -0.07216 |
| NF2 | ENSG00000186575 | 22:29698595-29698598 | + | TE | 0.18504 | 0.037421 | 0.14762 |
| NFATC2 | ENSG00000101096 | 20:51388626-51391227 | - | TE | 0.001325 | 0.006387 | -0.00506 |
| NFATC3 | ENSG00000072736 | 16:68085389-68085440 | + | TS | 0.61135 | 0.042749 | 0.5686 |
| NFATC3 | ENSG00000072736 | 16:68226350-68226471 | + | TE | 0.12796 | 0.005931 | 0.12202 |
| NFIA | ENSG00000162599 | 1:61455303-61455562 | + | TE | 0.073779 | 0.005529 | 0.06825 |
| NFKB1 | ENSG00000109320 | 4:102501331-102501788 | + | TS | 0.14864 | 0.008154 | 0.14048 |
| NFYC | ENSG00000066136 | 1:40770712-40771102 | + | TE | 0.003111 | 0.050874 | -0.04776 |
| NFYC | ENSG00000066136 | 1:40771103-40771106 | + | TE | 0.031604 | 0.004679 | 0.026925 |
| NGDN | ENSG00000129460 | 14:23478007-23478178 | + | TE | 0.064263 | 0.38689 | -0.32262 |
| NGLY1 | ENSG00000151092 | 3:25783499-25783551 | - | TS | 0.50925 | 0.11459 | 0.39466 |
| NISCH | ENSG00000010322 | 3:52493069-52493071 | + | TE | 0.018264 | 0.10727 | -0.08901 |
| NKAP | ENSG00000101882 | X:119925051-119925394 | - | TE | 0.00733 | 0.034292 | -0.02696 |
| NLRC3 | ENSG00000167984 | 16:3539038-3541915 | - | TE | 0.080175 | 0.49952 | -0.41934 |
| NME3 | ENSG00000103024 | 16:1771489-1771510 | - | TS | 0.092087 | 0.37564 | -0.28355 |
| NME3 | ENSG00000103024 | 16:1770377-1770480 | - | TE | 0.16481 | 0.029387 | 0.13543 |
| NME3 | ENSG00000103024 | 16:1770342-1770343 | - | TE | 0.16426 | 0.028719 | 0.13554 |
| NME7 | ENSG00000143156 | 1:169132531-169132536 | - | TE | 0.37865 | 0.091889 | 0.28676 |
| NMRK1 | ENSG00000106733 | 9:75061202-75061567 | - | TE | 0.017532 | 0.1291 | -0.11157 |
| NOL4L | ENSG00000197183 | 20:32483362-32483471 | - | TS | 0.87132 | 0.13935 | 0.73197 |
| NOL4L | ENSG00000197183 | 20:32483472-32483582 | - | TS | 0.12404 | 0.85867 | -0.73463 |
| NOP10 | ENSG00000182117 | 15:34341713-34341718 | - | TE | 0.081192 | 0.007832 | 0.073361 |
| NOP16 | ENSG00000048162 | 5:176388763-176388927 | - | TS | 0.1624 | 0.03553 | 0.12687 |
| NOP53 | ENSG00000105373 | 19:47745551-47745564 | + | TS | 0.11326 | 0.76678 | -0.65352 |
| NOVA1 | ENSG00000139910 | 14:26445889-26448963 | - | TE | 0.10959 | 0.84528 | -0.73569 |
| NOVA1 | ENSG00000139910 | 14:26443093-26445888 | - | TE | 0.89055 | 0.15659 | 0.73396 |
| NPHP3 | ENSG00000113971 | 3:132722008-132722414 | - | TS | 0.1284 | 0.67736 | -0.54897 |
| NPHP4 | ENSG00000131697 | 1:5862813-5862817 | - | TE | 0.073708 | 0.38583 | -0.31212 |
| NPM1 | ENSG00000181163 | 5:171410884-171410884 | + | TE | 0.094564 | 0.005555 | 0.089009 |
| NPRL2 | ENSG00000114388 | 3:50347491-50347673 | - | TE | 0.099779 | 0.86081 | -0.76103 |
| NPRL2 | ENSG00000114388 | 3:50347488-50347489 | - | TE | 0.90324 | 0.13692 | 0.76633 |
| NPRL3 | ENSG00000103148 | 16:84274-84595 | - | TE | 0.042007 | 0.007649 | 0.034359 |
| NR2C2 | ENSG00000177463 | 3:14947584-14947728 | + | TS | 0.10649 | 0.46883 | -0.36235 |
| NR3C1 | ENSG00000113580 | 5:143403690-143403700 | - | TS | 0.15547 | 0.031337 | 0.12413 |
| NR3C2 | ENSG00000151623 | 4:148080193-148080866 | - | TE | 0.020278 | 0.089566 | -0.06929 |
| NR6A1 | ENSG00000148200 | 9:124771278-124771297 | - | TS | 0.37241 | 0.088697 | 0.28372 |
| NRBP1 | ENSG00000115216 | 2:27442256-27442259 | + | TE | 0.043821 | 0.001942 | 0.041879 |
| NRG1 | ENSG00000157168 | 8:32742734-32743207 | + | TE | 0.90325 | 0.20894 | 0.69431 |
| NRG1 | ENSG00000157168 | 8:32743208-32743224 | + | TE | 0.094762 | 0.79013 | -0.69537 |
| NRGN | ENSG00000154146 | 11:124747207-124747210 | + | TE | 0.000263 | 0.007505 | -0.00724 |
| NRN1 | ENSG00000124785 | 6:5997999-5998001 | - | TE | 0.57916 | 0.061294 | 0.51786 |
| NRP1 | ENSG00000099250 | 10:33179897-33180365 | - | TE | 0.35373 | 0.057866 | 0.29587 |
| NRP2 | ENSG00000118257 | 2:205794754-205795058 | + | TE | 0.83605 | 0.12583 | 0.71022 |
| NRP2 | ENSG00000118257 | 2:205795173-205798133 | + | TE | 0.16508 | 0.87177 | -0.70668 |
| NSL1 | ENSG00000117697 | 1:212738562-212738686 | - | TE | 0.003351 | 0.019872 | -0.01652 |
| NSMAF | ENSG00000035681 | 8:58583532-58583769 | - | TE | 0.000651 | 0.009422 | -0.00877 |
| NT5C3B | ENSG00000141698 | 17:41825181-41825237 | - | TE | 0.37571 | 0.046566 | 0.32914 |
| NT5E | ENSG00000135318 | 6:85493841-85495778 | + | TE | 0.15007 | 0.008345 | 0.14172 |
| NTPCR | ENSG00000135778 | 1:232978652-232978722 | + | TE | 0.02647 | 0.006611 | 0.019859 |
| NUDCD1 | ENSG00000120526 | 8:109242675-109243301 | - | TE | 0.19258 | 0.032422 | 0.16016 |
| NUDT16L1 | ENSG00000168101 | 16:4693694-4693706 | + | TS | 0.71263 | 0.13517 | 0.57747 |
| NUDT4 | ENSG00000173598 | 12:93377896-93378421 | + | TS | 0.010133 | 0.001251 | 0.008882 |
| NUMB | ENSG00000133961 | 14:73275107-73275215 | - | TE | 0.000308 | 0.001563 | -0.00125 |
| NUP214 | ENSG00000126883 | 9:131197844-131197886 | + | TE | 0.030711 | 0.12724 | -0.09653 |
| NUP50 | ENSG00000093000 | 22:45184453-45184830 | + | TE | 0.006983 | 0.028448 | -0.02147 |
| NUP50 | ENSG00000093000 | 22:45184831-45185624 | + | TE | 0.000317 | 0.023134 | -0.02282 |
| NUP62CL | ENSG00000198088 | X:107123427-107123471 | - | TE | 0.37785 | 0.093173 | 0.28467 |
| NUPL2 | ENSG00000136243 | 7:23201007-23201009 | + | TE | 0.012786 | 0.06362 | -0.05083 |
| OAS1 | ENSG00000089127 | 12:112919616-112919875 | + | TE | 0.012659 | 0.12898 | -0.11632 |
| OAS2 | ENSG00000111335 | 12:113011719-113011723 | + | TE | 0.013952 | 0.002815 | 0.011138 |
| OAT | ENSG00000065154 | 10:124397303-124397306 | - | TE | 0.43484 | 0.010255 | 0.42458 |
| OAZ1 | ENSG00000104904 | 19:2272972-2273269 | + | TE | 0.014548 | 0.001299 | 0.013249 |
| OAZ2 | ENSG00000180304 | 15:64688485-64688834 | - | TE | 0.005386 | 0.000812 | 0.004574 |
| ODF2L | ENSG00000122417 | 1:86348738-86348878 | - | TE | 0.006084 | 0.47354 | -0.46746 |
| OGFOD3 | ENSG00000181396 | 17:82390116-82392009 | - | TE | 0.17429 | 0.039536 | 0.13475 |
| OGG1 | ENSG00000114026 | 3:9757408-9757513 | + | TE | 0.014269 | 0.001382 | 0.012887 |
| OPA1 | ENSG00000198836 | 3:193697284-193697384 | + | TE | 0.009452 | 0.001818 | 0.007634 |
| OPA1 | ENSG00000198836 | 3:193695734-193696496 | + | TE | 0.001865 | 0.017546 | -0.01568 |
| OPA1 | ENSG00000198836 | 3:193697441-193697523 | + | TE | 0.008799 | 0.001805 | 0.006993 |
| OR2AT4 | ENSG00000171561 | 11:75081753-75088712 | - | TE | 0.14229 | 0.63459 | -0.4923 |
| OR2T2 | ENSG00000196240 | 1:248452776-248453829 | + | TE | 0.26923 | 0.050274 | 0.21895 |
| OR2T33 | ENSG00000177212 | 1:248269917-248272770 | - | TE | 0.62744 | 0.066552 | 0.56088 |
| OR3A3 | ENSG00000159961 | 17:3420580-3421533 | + | TE | 0.031578 | 0.51953 | -0.48795 |
| OR51I2 | ENSG00000187918 | 11:5453408-5454477 | + | TE | 0.65546 | 0.086137 | 0.56932 |
| OR52N5 | ENSG00000181009 | 11:5777634-5778657 | - | TE | 0.077801 | 0.40864 | -0.33084 |
| ORAI2 | ENSG00000160991 | 7:102448733-102451609 | + | TE | 0.000459 | 0.021844 | -0.02139 |
| OSBP2 | ENSG00000184792 | 22:30906407-30906554 | + | TE | 0.046024 | 0.005805 | 0.040219 |
| OSBPL2 | ENSG00000130703 | 20:62294852-62294856 | + | TE | 0.020333 | 0.001237 | 0.019096 |
| OSBPL3 | ENSG00000070882 | 7:24796539-24799765 | - | TE | 0.02239 | 0.2728 | -0.25041 |
| OSCAR | ENSG00000170909 | 19:54094670-54095357 | - | TE | 0.1989 | 0.018795 | 0.1801 |
| OVOL1 | ENSG00000172818 | 11:65795046-65796089 | + | TE | 0.10566 | 0.47618 | -0.37052 |
| P2RX5 | ENSG00000083454 | 17:3673230-3673599 | - | TE | 0.32048 | 0.031429 | 0.28905 |
| P2RX7 | ENSG00000089041 | 12:121184803-121185081 | + | TE | 0.002795 | 0.017943 | -0.01515 |
| P2RX7 | ENSG00000089041 | 12:121184678-121184802 | + | TE | 0.002833 | 0.01237 | -0.00954 |
| P2RY10 | ENSG00000078589 | X:78961941-78961954 | + | TE | 0.010843 | 0.049451 | -0.03861 |
| P4HB | ENSG00000185624 | 17:81843166-81843491 | - | TE | 0.051864 | 0.25817 | -0.20631 |
| PAAF1 | ENSG00000175575 | 11:73927729-73927745 | + | TE | 0.041168 | 0.26003 | -0.21886 |
| PACRGL | ENSG00000163138 | 4:20728322-20728357 | + | TE | 0.095301 | 0.48006 | -0.38476 |
| PAM | ENSG00000145730 | 5:103029111-103029165 | + | TE | 0.068304 | 0.008047 | 0.060257 |
| PANK2 | ENSG00000125779 | 20:3923632-3923702 | + | TE | 0.000436 | 0.005498 | -0.00506 |
| PAQR4 | ENSG00000162073 | 16:2973483-2973489 | + | TE | 0.14541 | 0.027903 | 0.11751 |
| PAQR6 | ENSG00000160781 | 1:156243415-156243415 | - | TE | 0.012956 | 0.12738 | -0.11442 |
| PARP14 | ENSG00000173193 | 3:122720828-122720968 | + | TE | 0.066058 | 0.28825 | -0.2222 |
| PARP15 | ENSG00000173200 | 3:122636040-122636153 | + | TE | 0.001144 | 0.011542 | -0.0104 |
| PARP8 | ENSG00000151883 | 5:50842342-50842342 | + | TE | 0.000196 | 0.02777 | -0.02757 |
| PARPBP | ENSG00000185480 | 12:102195951-102196368 | + | TE | 0.18053 | 0.028165 | 0.15237 |
| PATZ1 | ENSG00000100105 | 22:31325809-31327309 | - | TE | 0.31181 | 0.062991 | 0.24882 |
| PAX8 | ENSG00000125618 | 2:113218469-113218609 | - | TE | 0.04159 | 0.18454 | -0.14295 |
| PBK | ENSG00000168078 | 8:27810201-27810501 | - | TE | 0.07813 | 0.3242 | -0.24607 |
| PBX1 | ENSG00000185630 | 1:164851822-164851831 | + | TE | 0.057158 | 0.011873 | 0.045285 |
| PCDH1 | ENSG00000156453 | 5:141863013-141863056 | - | TE | 0.035747 | 0.24947 | -0.21372 |
| PCDHGA9 | ENSG00000261934 | 5:141402953-141405376 | + | TS | 0.3209 | 0.061205 | 0.2597 |
| PCF11 | ENSG00000165494 | 11:83156988-83157126 | + | TS | 0.010819 | 0.064291 | -0.05347 |
| PCGF3 | ENSG00000185619 | 4:766674-767864 | + | TE | 0.000448 | 0.021192 | -0.02074 |
| PCID2 | ENSG00000126226 | 13:113177611-113177611 | - | TE | 0.15288 | 0.025586 | 0.1273 |
| PCID2 | ENSG00000126226 | 13:113177613-113177616 | - | TE | 0.025404 | 0.14389 | -0.11848 |
| PCK2 | ENSG00000100889 | 14:24100304-24100562 | + | TE | 0.14344 | 0.86597 | -0.72252 |
| PCK2 | ENSG00000100889 | 14:24100244-24100303 | + | TE | 0.86077 | 0.12687 | 0.7339 |
| PCMTD1 | ENSG00000168300 | 8:51820340-51820718 | - | TE | 0.007332 | 0.034053 | -0.02672 |
| PCMTD2 | ENSG00000203880 | 20:64273221-64274842 | + | TE | 0.001918 | 0.07505 | -0.07313 |
| PCYT2 | ENSG00000185813 | 17:81904424-81904635 | - | TE | 0.10214 | 0.008811 | 0.093331 |
| PDCD10 | ENSG00000114209 | 3:167683912-167684389 | - | TE | 0.091999 | 0.013743 | 0.078256 |
| PDCD2 | ENSG00000071994 | 6:170575295-170575327 | - | TE | 0.059346 | 0.002524 | 0.056822 |
| PDCD2 | ENSG00000071994 | 6:170581748-170581791 | - | TE | 0.10643 | 0.023198 | 0.083234 |
| PDCD6IP | ENSG00000170248 | 3:33798571-33798574 | + | TS | 0.0228 | 0.13775 | -0.11495 |
| PDE4DIP | ENSG00000178104 | 1:149032498-149033016 | + | TE | 0.16644 | 0.032482 | 0.13395 |
| PDE8A | ENSG00000073417 | 15:85139137-85139142 | + | TE | 0.17199 | 0.033895 | 0.1381 |
| PDE8B | ENSG00000113231 | 5:77426445-77426633 | + | TE | 0.070872 | 0.007431 | 0.063441 |
| PDGFC | ENSG00000145431 | 4:156762948-156763206 | - | TE | 0.02087 | 0.11526 | -0.09439 |
| PDHA1 | ENSG00000131828 | X:19359845-19361685 | + | TE | 0.042934 | 0.006434 | 0.0365 |
| PDLIM5 | ENSG00000163110 | 4:94664036-94664186 | + | TE | 0.003943 | 0.10249 | -0.09854 |
| PDLIM5 | ENSG00000163110 | 4:94665501-94665501 | + | TE | 0.059503 | 0.011751 | 0.047752 |
| PDLIM5 | ENSG00000163110 | 4:94586989-94588127 | + | TE | 0.056931 | 0.30124 | -0.24431 |
| PDLIM5 | ENSG00000163110 | 4:94668224-94668227 | + | TE | 0.47074 | 0.099005 | 0.37173 |
| PEX1 | ENSG00000127980 | 7:92487020-92487025 | - | TE | 0.078226 | 0.35929 | -0.28106 |
| PEX26 | ENSG00000215193 | 22:18088461-18105396 | + | TE | 0.55248 | 0.12483 | 0.42766 |
| PEX5 | ENSG00000139197 | 12:7210257-7210262 | + | TE | 0.13918 | 0.70696 | -0.56778 |
| PEX5L | ENSG00000114757 | 3:179801817-179802032 | - | TE | 0.26902 | 0.056276 | 0.21275 |
| PFN2 | ENSG00000070087 | 3:149964935-149965347 | - | TE | 0.084377 | 0.015308 | 0.069068 |
| PGA4 | ENSG00000229183 | 11:61231453-61231566 | + | TE | 0.047095 | 0.19008 | -0.14298 |
| PGAP1 | ENSG00000197121 | 2:196835590-196841338 | - | TE | 0.03142 | 0.004522 | 0.026898 |
| PGAP2 | ENSG00000148985 | 11:3826331-3826334 | + | TE | 0.072913 | 0.002419 | 0.070494 |
| PGAP2 | ENSG00000148985 | 11:3826335-3826352 | + | TE | 0.007884 | 0.001596 | 0.006289 |
| PGRMC2 | ENSG00000164040 | 4:128269242-128269255 | - | TE | 0.002769 | 0.041294 | -0.03853 |
| PHACTR2 | ENSG00000112419 | 6:143824122-143826177 | + | TE | 0.038795 | 0.006217 | 0.032579 |
| PHACTR4 | ENSG00000204138 | 1:28500365-28500369 | + | TE | 0.016406 | 0.13509 | -0.11869 |
| PHF14 | ENSG00000106443 | 7:11102486-11102646 | + | TE | 0.15349 | 0.037715 | 0.11577 |
| PHF20L1 | ENSG00000129292 | 8:132811129-132812316 | + | TE | 0.000926 | 0.009467 | -0.00854 |
| PHF23 | ENSG00000040633 | 17:7235038-7235286 | - | TE | 0.13435 | 0.032418 | 0.10193 |
| PHF8 | ENSG00000172943 | X:53943256-53943402 | - | TE | 0.012495 | 0.12119 | -0.10869 |
| PHIP | ENSG00000146247 | 6:78940546-78941330 | - | TE | 0.000277 | 0.011917 | -0.01164 |
| PIAS2 | ENSG00000078043 | 18:46812280-46812612 | - | TE | 0.010234 | 0.075699 | -0.06547 |
| PIEZO1 | ENSG00000103335 | 16:88715346-88715448 | - | TE | 0.21807 | 0.034743 | 0.18332 |
| PIGG | ENSG00000174227 | 4:539529-539529 | + | TE | 0.29389 | 0.035944 | 0.25794 |
| PIGG | ENSG00000174227 | 4:522838-522842 | + | TE | 0.15018 | 0.032555 | 0.11762 |
| PIGN | ENSG00000197563 | 18:62044311-62044323 | - | TE | 0.039787 | 0.007813 | 0.031975 |
| PIGN | ENSG00000197563 | 18:62045187-62045341 | - | TE | 0.041403 | 0.00777 | 0.033633 |
| PIGO | ENSG00000165282 | 9:35088691-35088692 | - | TE | 0.088624 | 0.58104 | -0.49242 |
| PIGQ | ENSG00000007541 | 16:582883-583035 | + | TE | 0.013063 | 0.082086 | -0.06902 |
| PIGW | ENSG00000277161 | 17:36537935-36538854 | + | TE | 0.098389 | 0.023729 | 0.07466 |
| PIGW | ENSG00000277161 | 17:36537094-36537934 | + | TE | 0.004547 | 0.018219 | -0.01367 |
| PIH1D1 | ENSG00000104872 | 19:49451818-49451901 | - | TS | 0.90192 | 0.12361 | 0.77831 |
| PIH1D1 | ENSG00000104872 | 19:49451811-49451817 | - | TS | 0.10225 | 0.87824 | -0.776 |
| PIK3R1 | ENSG00000145675 | 5:68298584-68298605 | + | TE | 0.00563 | 0.000603 | 0.005027 |
| PIK3R3 | ENSG00000117461 | 1:46040142-46042640 | - | TE | 0.353 | 0.082366 | 0.27064 |
| PILRB | ENSG00000121716 | 7:100352176-100352267 | + | TS | 0.11457 | 0.47883 | -0.36426 |
| PIM2 | ENSG00000102096 | X:48914104-48914294 | - | TE | 0.008619 | 0.070615 | -0.062 |
| PIM3 | ENSG00000198355 | 22:49960513-49960767 | + | TS | 0.45404 | 0.10894 | 0.3451 |
| PITPNM3 | ENSG00000091622 | 17:6455391-6455399 | - | TE | 0.24262 | 0.058026 | 0.1846 |
| PITRM1 | ENSG00000107959 | 10:3137730-3138124 | - | TE | 0.57053 | 0.14033 | 0.4302 |
| PKD2 | ENSG00000118762 | 4:88075859-88075863 | + | TE | 0.064161 | 0.014615 | 0.049547 |
| PKLR | ENSG00000143627 | 1:155289293-155289838 | - | TE | 0.86322 | 0.20024 | 0.66297 |
| PKLR | ENSG00000143627 | 1:155289839-155290678 | - | TE | 0.13626 | 0.7945 | -0.65824 |
| PKP4 | ENSG00000144283 | 2:158681427-158681429 | + | TE | 0.016367 | 0.13296 | -0.11659 |
| PLA2G12A | ENSG00000123739 | 4:109713917-109714261 | - | TE | 0.013452 | 0.002017 | 0.011435 |
| PLA2G4B | ENSG00000243708 | 15:41848141-41848147 | + | TE | 0.067072 | 0.35052 | -0.28345 |
| PLA2G6 | ENSG00000184381 | 22:38111495-38111499 | - | TE | 0.007085 | 0.091041 | -0.08396 |
| PLAC8 | ENSG00000145287 | 4:83114716-83114736 | - | TS | 0.21938 | 0.053824 | 0.16556 |
| PLAGL1 | ENSG00000118495 | 6:143940302-143940334 | - | TE | 0.02658 | 0.002101 | 0.024479 |
| PLAGL1 | ENSG00000118495 | 6:143941284-143941423 | - | TE | 0.025392 | 0.004147 | 0.021245 |
| PLCB3 | ENSG00000149782 | 11:64267922-64267923 | + | TE | 0.87032 | 0.12848 | 0.74184 |
| PLCB3 | ENSG00000149782 | 11:64267571-64267921 | + | TE | 0.13348 | 0.87723 | -0.74375 |
| PLEKHF1 | ENSG00000166289 | 19:29674224-29674843 | + | TE | 0.006035 | 0.14682 | -0.14078 |
| PLEKHG7 | ENSG00000187510 | 12:92708548-92708549 | + | TE | 0.12875 | 0.9044 | -0.77565 |
| PLEKHG7 | ENSG00000187510 | 12:92707673-92708547 | + | TE | 0.87092 | 0.09829 | 0.77263 |
| PLIN2 | ENSG00000147872 | 9:19123313-19123476 | - | TE | 0.12819 | 0.56859 | -0.4404 |
| PLOD2 | ENSG00000152952 | 3:146070241-146070872 | - | TE | 0.040531 | 0.17687 | -0.13634 |
| PLSCR3 | ENSG00000187838 | 17:7389727-7389729 | - | TE | 0.05539 | 0.01278 | 0.04261 |
| PLSCR3 | ENSG00000187838 | 17:7389733-7389734 | - | TE | 0.1852 | 0.016895 | 0.16831 |
| PLXNA3 | ENSG00000130827 | X:154472590-154472696 | + | TE | 0.006502 | 0.24634 | -0.23984 |
| PML | ENSG00000140464 | 15:73994673-73994716 | + | TS | 0.019421 | 0.095305 | -0.07588 |
| PNISR | ENSG00000132424 | 6:99401461-99401630 | - | TE | 0.000648 | 0.034012 | -0.03336 |
| PNP | ENSG00000198805 | 14:20477088-20477094 | + | TE | 0.001822 | 0.034813 | -0.03299 |
| PNPLA8 | ENSG00000135241 | 7:108472531-108472675 | - | TE | 0.088357 | 0.00473 | 0.083627 |
| PNRC2 | ENSG00000189266 | 1:23961444-23961880 | + | TE | 0.000656 | 0.023359 | -0.0227 |
| POLD4 | ENSG00000175482 | 11:67351548-67351552 | - | TE | 0.11615 | 0.006003 | 0.11015 |
| POLD4 | ENSG00000175482 | 11:67351553-67351564 | - | TE | 0.047865 | 0.005856 | 0.042009 |
| POLI | ENSG00000101751 | 18:54293649-54293776 | + | TE | 0.055103 | 0.22308 | -0.16797 |
| POLM | ENSG00000122678 | 7:44082512-44082514 | - | TS | 0.10466 | 0.48156 | -0.37691 |
| POLM | ENSG00000122678 | 7:44072262-44073217 | - | TE | 0.003436 | 0.034412 | -0.03098 |
| POLR1B | ENSG00000125630 | 2:112574847-112576029 | + | TE | 0.007019 | 0.031308 | -0.02429 |
| POLR1D | ENSG00000186184 | 13:27622875-27623449 | + | TE | 0.004714 | 0.12682 | -0.12211 |
| POLR2G | ENSG00000168002 | 11:62766494-62766707 | + | TE | 0.080502 | 0.39274 | -0.31224 |
| POLR2M | ENSG00000255529 | 15:57708714-57708746 | + | TE | 0.013117 | 0.067309 | -0.05419 |
| POLR3E | ENSG00000058600 | 16:22334442-22335101 | + | TE | 0.1112 | 0.026445 | 0.084751 |
| POLR3E | ENSG00000058600 | 16:22333697-22333935 | + | TE | 0.013345 | 0.093998 | -0.08065 |
| POLR3H | ENSG00000100413 | 22:41529212-41529336 | - | TE | 0.004255 | 0.21613 | -0.21187 |
| POMC | ENSG00000115138 | 2:25160853-25160914 | - | TE | 0.070981 | 0.38451 | -0.31353 |
| POP4 | ENSG00000105171 | 19:29615704-29615800 | + | TE | 0.13243 | 0.006573 | 0.12586 |
| POU5F1 | ENSG00000204531 | 6:31164348-31164350 | - | TE | 0.172 | 0.038792 | 0.13321 |
| PPARGC1B | ENSG00000155846 | 5:149847705-149847718 | + | TE | 0.035912 | 0.005941 | 0.029971 |
| PPCDC | ENSG00000138621 | 15:75044515-75044623 | + | TE | 0.002964 | 0.072413 | -0.06945 |
| PPCS | ENSG00000127125 | 1:42460413-42460414 | + | TE | 0.023368 | 0.1439 | -0.12053 |
| PPHLN1 | ENSG00000134283 | 12:42448619-42448623 | + | TE | 0.075268 | 0.34451 | -0.26924 |
| PPIA | ENSG00000196262 | 7:44801631-44801631 | + | TE | 0.004485 | 0.10949 | -0.10501 |
| PPIA | ENSG00000196262 | 7:44801287-44801624 | + | TE | 0.18143 | 0.003161 | 0.17827 |
| PPIE | ENSG00000084072 | 1:39753287-39753707 | + | TE | 0.051566 | 0.49936 | -0.44779 |
| PPIF | ENSG00000108179 | 10:79347658-79347743 | + | TS | 0.11658 | 0.029118 | 0.087466 |
| PPM1L | ENSG00000163590 | 3:161069845-161069856 | + | TE | 0.001343 | 0.006509 | -0.00517 |
| PPM1L | ENSG00000163590 | 3:161068811-161069844 | + | TE | 0.001353 | 0.014044 | -0.01269 |
| PPM1N | ENSG00000213889 | 19:45498607-45499609 | + | TS | 0.36061 | 0.081847 | 0.27877 |
| PPP1CC | ENSG00000186298 | 12:110720748-110721104 | - | TE | 0.02236 | 0.13124 | -0.10888 |
| PPP2CB | ENSG00000104695 | 8:30786109-30786307 | - | TE | 0.011686 | 0.14145 | -0.12977 |
| PPP2R1A | ENSG00000105568 | 19:52226402-52226417 | + | TE | 0.030483 | 0.004463 | 0.02602 |
| PPP2R1B | ENSG00000137713 | 11:111741055-111741320 | - | TE | 0.041625 | 0.009393 | 0.032231 |
| PPP2R3A | ENSG00000073711 | 3:136145043-136145384 | + | TE | 0.030633 | 0.004991 | 0.025643 |
| PPP3CA | ENSG00000138814 | 4:101346843-101347327 | - | TS | 0.017 | 0.13558 | -0.11858 |
| PPP6R3 | ENSG00000110075 | 11:68613066-68613117 | + | TE | 0.000901 | 0.073798 | -0.0729 |
| PQLC3 | ENSG00000162976 | 2:11178871-11178874 | + | TE | 0.093677 | 0.005348 | 0.088328 |
| PRDM1 | ENSG00000057657 | 6:106107509-106108282 | + | TE | 0.000524 | 0.017429 | -0.01691 |
| PRDM10 | ENSG00000170325 | 11:129902340-129902516 | - | TE | 0.083133 | 0.00255 | 0.080583 |
| PRELID3A | ENSG00000141391 | 18:12431150-12431463 | + | TE | 0.43748 | 0.10424 | 0.33324 |
| PRICKLE1 | ENSG00000139174 | 12:42459265-42459279 | - | TE | 0.070875 | 0.012433 | 0.058442 |
| PRICKLE1 | ENSG00000139174 | 12:42458397-42458824 | - | TE | 0.030731 | 0.12838 | -0.09765 |
| PRICKLE2 | ENSG00000163637 | 3:64093871-64095740 | - | TE | 0.11862 | 0.48887 | -0.37025 |
| PRIMPOL | ENSG00000164306 | 4:184694759-184694952 | + | TE | 0.13935 | 0.88944 | -0.75009 |
| PRIMPOL | ENSG00000164306 | 4:184694953-184694963 | + | TE | 0.8614 | 0.11089 | 0.7505 |
| PRKAG2 | ENSG00000106617 | 7:151556111-151556123 | - | TE | 0.002888 | 0.021818 | -0.01893 |
| PRKD3 | ENSG00000115825 | 2:37317024-37317079 | - | TS | 0.14712 | 0.001947 | 0.14517 |
| PRLR | ENSG00000113494 | 5:35065089-35065376 | - | TE | 0.30486 | 0.047095 | 0.25777 |
| PRMT3 | ENSG00000185238 | 11:20509254-20509282 | + | TE | 0.31563 | 0.027802 | 0.28783 |
| PRMT9 | ENSG00000164169 | 4:147683799-147684125 | - | TS | 0.65256 | 0.12542 | 0.52714 |
| PRPF38B | ENSG00000134186 | 1:108692323-108692323 | + | TS | 0.23039 | 0.017693 | 0.21269 |
| PRPS2 | ENSG00000101911 | X:12822704-12823086 | + | TE | 0.006608 | 0.053621 | -0.04701 |
| PRPSAP2 | ENSG00000141127 | 17:18930540-18930734 | + | TE | 0.14395 | 0.033213 | 0.11073 |
| PRR14 | ENSG00000156858 | 16:30656032-30656408 | + | TE | 0.16661 | 0.73176 | -0.56514 |
| PRR29 | ENSG00000224383 | 17:64003114-64004285 | + | TE | 0.049052 | 0.004465 | 0.044587 |
| PRR3 | ENSG00000204576 | 6:30556886-30557279 | + | TS | 0.070737 | 0.49596 | -0.42522 |
| PRR5L | ENSG00000135362 | 11:36462342-36463644 | + | TE | 0.002712 | 0.015252 | -0.01254 |
| PRRC2B | ENSG00000130723 | 9:131495740-131495874 | + | TE | 0.001194 | 0.42605 | -0.42486 |
| PRRC2C | ENSG00000117523 | 1:171591587-171593258 | + | TE | 0.016833 | 0.12153 | -0.1047 |
| PRRG2 | ENSG00000126460 | 19:49590371-49591004 | + | TE | 0.059858 | 0.49615 | -0.4363 |
| PRSS51 | ENSG00000253649 | 8:10496326-10496466 | - | TE | 0.02307 | 0.25021 | -0.22714 |
| PRXL2B | ENSG00000157870 | 1:2589514-2589632 | + | TE | 0.056149 | 0.009886 | 0.046263 |
| PSAT1 | ENSG00000135069 | 9:78328981-78329454 | + | TE | 0.019072 | 0.091605 | -0.07253 |
| PSENEN | ENSG00000205155 | 19:35745600-35745618 | + | TS | 0.48704 | 0.095752 | 0.39129 |
| PSMA5 | ENSG00000143106 | 1:109402013-109402090 | - | TE | 0.010831 | 0.27244 | -0.26161 |
| PSMA6 | ENSG00000100902 | 14:35317473-35317474 | + | TE | 0.07939 | 0.43498 | -0.35559 |
| PSMB9 | ENSG00000240065 | 6:32854148-32854160 | + | TS | 0.12292 | 0.49263 | -0.36971 |
| PSMC6 | ENSG00000100519 | 14:52721500-52723744 | + | TS | 0.14509 | 0.010167 | 0.13492 |
| PSMC6 | ENSG00000100519 | 14:52727992-52727995 | + | TE | 0.026788 | 0.19023 | -0.16344 |
| PSPN | ENSG00000125650 | 19:6375850-6379058 | - | TS | 0.01185 | 0.13124 | -0.11939 |
| PSTPIP1 | ENSG00000140368 | 15:76995122-76995125 | + | TS | 0.017007 | 0.070628 | -0.05362 |
| PTAR1 | ENSG00000188647 | 9:69718138-69718568 | - | TE | 0.000137 | 0.021718 | -0.02158 |
| PTBP1 | ENSG00000011304 | 19:811290-812222 | + | TE | 0.018192 | 0.003407 | 0.014785 |
| PTBP2 | ENSG00000117569 | 1:96813276-96813447 | + | TE | 0.15909 | 0.036582 | 0.12251 |
| PTCD2 | ENSG00000049883 | 5:72358839-72358912 | + | TE | 0.41708 | 0.090649 | 0.32643 |
| PTDSS2 | ENSG00000174915 | 11:491388-491399 | + | TE | 0.1245 | 0.87819 | -0.75369 |
| PTDSS2 | ENSG00000174915 | 11:491121-491387 | + | TE | 0.87315 | 0.12305 | 0.75011 |
| PTGS2 | ENSG00000073756 | 1:186672893-186674352 | - | TE | 0.001198 | 0.014752 | -0.01355 |
| PTMA | ENSG00000187514 | 2:231708516-231708529 | + | TS | 0.000535 | 0.004525 | -0.00399 |
| PTMS | ENSG00000159335 | 12:6770392-6770498 | + | TE | 0.006118 | 0.16312 | -0.15701 |
| PTOV1 | ENSG00000104960 | 19:49851191-49851224 | + | TS | 0.036284 | 0.23428 | -0.198 |
| PTP4A2 | ENSG00000184007 | 1:31919110-31919172 | - | TS | 0.027545 | 0.002813 | 0.024731 |
| PTPA | ENSG00000119383 | 9:129147773-129147957 | + | TE | 0.002871 | 0.022871 | -0.02 |
| PTPMT1 | ENSG00000110536 | 11:47571928-47572119 | + | TE | 0.052805 | 0.005902 | 0.046903 |
| PTPN22 | ENSG00000134242 | 1:113871754-113871759 | - | TS | 0.72847 | 0.11094 | 0.61753 |
| PTPN22 | ENSG00000134242 | 1:113814679-113814969 | - | TE | 0.025442 | 0.10637 | -0.08093 |
| PTPRA | ENSG00000132670 | 20:3038669-3038674 | + | TE | 0.046215 | 0.003504 | 0.042711 |
| PTPRN2 | ENSG00000155093 | 7:157539061-157539065 | - | TE | 0.005122 | 0.19139 | -0.18627 |
| PUS7L | ENSG00000129317 | 12:43729763-43730702 | - | TE | 0.000754 | 0.01124 | -0.01049 |
| PXK | ENSG00000168297 | 3:58425059-58425151 | + | TE | 0.000768 | 0.008647 | -0.00788 |
| PYCR1 | ENSG00000183010 | 17:81932921-81932934 | - | TE | 0.51933 | 0.094165 | 0.42516 |
| PYGL | ENSG00000100504 | 14:50944484-50944736 | - | TS | 0.12298 | 0.005247 | 0.11773 |
| PYM1 | ENSG00000170473 | 12:55901971-55902355 | - | TE | 0.089379 | 0.51768 | -0.42831 |
| R3HDM1 | ENSG00000048991 | 2:135725267-135725268 | + | TE | 0.19196 | 0.047308 | 0.14465 |
| RAB12 | ENSG00000206418 | 18:8609445-8609953 | + | TS | 0.063619 | 0.26681 | -0.20319 |
| RAB13 | ENSG00000143545 | 1:153981654-153982176 | - | TE | 0.006867 | 0.0383 | -0.03143 |
| RAB15 | ENSG00000139998 | 14:64945828-64945838 | - | TE | 0.004265 | 0.13783 | -0.13356 |
| RAB15 | ENSG00000139998 | 14:64945827-64945827 | - | TE | 0.00366 | 0.076515 | -0.07286 |
| RAB29 | ENSG00000117280 | 1:205770300-205770453 | - | TE | 0.030513 | 0.001297 | 0.029216 |
| RAB29 | ENSG00000117280 | 1:205769703-205769717 | - | TE | 0.001526 | 0.013425 | -0.0119 |
| RAB2A | ENSG00000104388 | 8:60516936-60517175 | + | TS | 0.091653 | 0.019391 | 0.072262 |
| RAB34 | ENSG00000109113 | 17:28716756-28716902 | - | TE | 0.69362 | 0.11389 | 0.57973 |
| RAB38 | ENSG00000123892 | 11:88175183-88175433 | - | TS | 0.14446 | 0.77802 | -0.63356 |
| RAB38 | ENSG00000123892 | 11:88175434-88175467 | - | TS | 0.85838 | 0.21254 | 0.64584 |
| RABL6 | ENSG00000196642 | 9:136840485-136841177 | + | TE | 0.18952 | 0.014802 | 0.17472 |
| RACK1 | ENSG00000204628 | 5:181243743-181243889 | - | TS | 0.002688 | 0.011921 | -0.00923 |
| RAMP1 | ENSG00000132329 | 2:237912011-237912114 | + | TE | 0.13226 | 0.71834 | -0.58608 |
| RANBP1 | ENSG00000099901 | 22:20127354-20127355 | + | TE | 0.071803 | 0.39732 | -0.32551 |
| RANBP10 | ENSG00000141084 | 16:67806558-67806560 | - | TS | 0.030733 | 0.14933 | -0.11859 |
| RANBP3 | ENSG00000031823 | 19:5917323-5917351 | - | TE | 0.009794 | 0.055964 | -0.04617 |
| RANBP6 | ENSG00000137040 | 9:6015613-6015618 | - | TS | 0.50478 | 0.12612 | 0.37866 |
| RAPGEF3 | ENSG00000079337 | 12:47737257-47737539 | - | TE | 0.01888 | 0.085507 | -0.06663 |
| RASA4 | ENSG00000105808 | 7:102582691-102582892 | - | TE | 0.006001 | 0.036399 | -0.0304 |
| RASGRP1 | ENSG00000172575 | 15:38490480-38490553 | - | TE | 0.056904 | 0.007869 | 0.049035 |
| RASGRP3 | ENSG00000152689 | 2:33564606-33564731 | + | TE | 0.025122 | 0.25867 | -0.23354 |
| RASSF1 | ENSG00000068028 | 3:50336800-50337172 | - | TE | 0.041328 | 0.007525 | 0.033803 |
| RBAK | ENSG00000146587 | 7:5063695-5063875 | + | TE | 0.09454 | 0.002188 | 0.092352 |
| RBBP6 | ENSG00000122257 | 16:24540574-24540582 | + | TS | 0.002587 | 0.011684 | -0.0091 |
| RBM14 | ENSG00000239306 | 11:66627344-66627347 | + | TE | 0.053739 | 0.001562 | 0.052177 |
| RBM17 | ENSG00000134453 | 10:6115453-6115766 | + | TE | 0.013529 | 0.25598 | -0.24245 |
| RBM23 | ENSG00000100461 | 14:22900777-22901335 | - | TE | 0.008623 | 0.000471 | 0.008152 |
| RBM28 | ENSG00000106344 | 7:128310480-128310931 | - | TE | 0.13148 | 0.00158 | 0.1299 |
| RBM7 | ENSG00000076053 | 11:114408910-114408924 | + | TE | 0.001267 | 0.028745 | -0.02748 |
| RBMS1 | ENSG00000153250 | 2:160274518-160274539 | - | TE | 0.000532 | 0.002953 | -0.00242 |
| RBMS2 | ENSG00000076067 | 12:56589596-56589652 | + | TE | 0.009043 | 0.11502 | -0.10598 |
| RBMS2 | ENSG00000076067 | 12:56589140-56589503 | + | TE | 0.030341 | 0.003361 | 0.02698 |
| RBPJ | ENSG00000168214 | 4:26431008-26431066 | + | TE | 0.002791 | 0.000155 | 0.002637 |
| RBPJ | ENSG00000168214 | 4:26430692-26431007 | + | TE | 0.000136 | 0.002021 | -0.00189 |
| RBPMS | ENSG00000157110 | 8:30570995-30572256 | + | TE | 0.28965 | 0.008171 | 0.28148 |
| RBPMS | ENSG00000157110 | 8:30570637-30570761 | + | TE | 0.13837 | 0.007328 | 0.13105 |
| RCOR3 | ENSG00000117625 | 1:211314020-211314587 | + | TE | 0.000914 | 0.005753 | -0.00484 |
| RCOR3 | ENSG00000117625 | 1:211314588-211314591 | + | TE | 0.012366 | 0.000822 | 0.011544 |
| RDX | ENSG00000137710 | 11:110231550-110231550 | - | TE | 0.009438 | 0.066808 | -0.05737 |
| REEP1 | ENSG00000068615 | 2:86216554-86216580 | - | TE | 0.20674 | 0.048373 | 0.15836 |
| REEP1 | ENSG00000068615 | 2:86216517-86216553 | - | TE | 0.041905 | 0.30097 | -0.25906 |
| REEP4 | ENSG00000168476 | 8:22141451-22141768 | - | TS | 0.10308 | 0.025096 | 0.077987 |
| REEP5 | ENSG00000129625 | 5:112878577-112878835 | - | TE | 0.00078 | 0.008819 | -0.00804 |
| REL | ENSG00000162924 | 2:60921763-60922984 | + | TE | 0.001041 | 9.98E-05 | 0.000941 |
| RELT | ENSG00000054967 | 11:73395444-73396640 | + | TE | 0.000579 | 0.004699 | -0.00412 |
| REST | ENSG00000084093 | 4:56931794-56932184 | + | TE | 0.003377 | 0.050421 | -0.04705 |
| RETREG1 | ENSG00000154153 | 5:16508687-16508998 | - | TS | 0.50233 | 0.12449 | 0.37784 |
| RETREG2 | ENSG00000144567 | 2:219178227-219178633 | + | TS | 0.43509 | 0.094745 | 0.34035 |
| RFC2 | ENSG00000049541 | 7:74231537-74232216 | - | TE | 0.049718 | 0.2751 | -0.22539 |
| RFK | ENSG00000135002 | 9:76385517-76385545 | - | TE | 0.012171 | 0.050487 | -0.03832 |
| RFTN2 | ENSG00000162944 | 2:197571735-197572280 | - | TE | 0.003277 | 0.032688 | -0.02941 |
| RFX2 | ENSG00000087903 | 19:5994816-5994950 | - | TE | 0.24446 | 0.004147 | 0.24031 |
| RGMA | ENSG00000182175 | 15:93044749-93044817 | - | TE | 0.021361 | 0.089395 | -0.06803 |
| RGMA | ENSG00000182175 | 15:93044956-93045705 | - | TE | 0.084093 | 0.020011 | 0.064082 |
| RGPD1 | ENSG00000187627 | 2:87012664-87012741 | + | TE | 0.00468 | 0.023906 | -0.01923 |
| RGPD1 | ENSG00000187627 | 2:87012513-87012547 | + | TE | 0.004717 | 0.021215 | -0.0165 |
| RGPD5 | ENSG00000015568 | 2:109794274-109794307 | + | TS | 0.69602 | 0.09027 | 0.60575 |
| RGPD6 | ENSG00000183054 | 2:110513821-110515645 | - | TE | 0.14913 | 0.014017 | 0.13511 |
| RGS14 | ENSG00000169220 | 5:177372299-177372596 | + | TE | 0.000909 | 0.028546 | -0.02764 |
| RGS6 | ENSG00000182732 | 14:72562468-72562476 | + | TE | 0.08469 | 0.79045 | -0.70575 |
| RGS6 | ENSG00000182732 | 14:72562590-72562592 | + | TE | 0.91894 | 0.20723 | 0.71171 |
| RHEX | ENSG00000263961 | 1:206101752-206102448 | + | TE | 0.702 | 0.096132 | 0.60587 |
| RHNO1 | ENSG00000171792 | 12:2889272-2889440 | + | TE | 0.042971 | 0.007864 | 0.035108 |
| RHOC | ENSG00000155366 | 1:112701131-112701132 | - | TE | 0.069216 | 0.004849 | 0.064367 |
| RHOH | ENSG00000168421 | 4:40243406-40243695 | + | TE | 0.002535 | 0.049184 | -0.04665 |
| RHOQ | ENSG00000119729 | 2:46580928-46581046 | + | TE | 0.007571 | 0.058714 | -0.05114 |
| RHOT1 | ENSG00000126858 | 17:32225504-32225666 | + | TE | 0.00278 | 0.050942 | -0.04816 |
| RHOT1 | ENSG00000126858 | 17:32225728-32225770 | + | TE | 0.012761 | 0.002629 | 0.010132 |
| RIC3 | ENSG00000166405 | 11:8110698-8111137 | - | TE | 0.057149 | 0.22881 | -0.17166 |
| RIPOR1 | ENSG00000039523 | 16:67546638-67546786 | + | TE | 0.020965 | 0.15898 | -0.13802 |
| RIPOR2 | ENSG00000111913 | 6:24840272-24840296 | - | TE | 0.000281 | 0.002349 | -0.00207 |
| RMDN1 | ENSG00000176623 | 8:86474112-86474245 | - | TE | 0.16327 | 0.032872 | 0.13039 |
| RMDN2 | ENSG00000115841 | 2:38017742-38017744 | + | TE | 0.44093 | 0.084937 | 0.35599 |
| RMDN3 | ENSG00000137824 | 15:40755199-40755246 | - | TS | 0.85278 | 0.15308 | 0.6997 |
| RMI2 | ENSG00000175643 | 16:11351366-11351755 | + | TE | 0.1208 | 0.56448 | -0.44368 |
| RMND5B | ENSG00000145916 | 5:178148567-178148568 | + | TE | 0.15114 | 0.00214 | 0.149 |
| RNF111 | ENSG00000157450 | 15:58987655-58987951 | + | TS | 0.022554 | 0.11387 | -0.09132 |
| RNF111 | ENSG00000157450 | 15:59096427-59096431 | + | TE | 0.001672 | 0.016991 | -0.01532 |
| RNF125 | ENSG00000101695 | 18:32068298-32068489 | + | TE | 0.012333 | 0.001063 | 0.01127 |
| RNF144B | ENSG00000137393 | 6:18387350-18387513 | + | TS | 0.11589 | 0.013834 | 0.10205 |
| RNF146 | ENSG00000118518 | 6:127286616-127286891 | + | TE | 0.000836 | 0.003528 | -0.00269 |
| RNF146 | ENSG00000118518 | 6:127266849-127266850 | + | TS | 0.49777 | 0.12298 | 0.3748 |
| RNF149 | ENSG00000163162 | 2:101308129-101308696 | - | TS | 0.015462 | 0.14405 | -0.12859 |
| RNF4 | ENSG00000063978 | 4:2514475-2514503 | + | TE | 0.000627 | 0.010032 | -0.00941 |
| RNF7 | ENSG00000114125 | 3:141738204-141738248 | + | TS | 0.38936 | 0.088277 | 0.30108 |
| RNH1 | ENSG00000023191 | 11:494512-494514 | - | TE | 0.041845 | 0.010029 | 0.031817 |
| RNPEPL1 | ENSG00000142327 | 2:240568587-240568860 | + | TS | 0.012283 | 0.049363 | -0.03708 |
| RNPS1 | ENSG00000205937 | 16:2253126-2253129 | - | TE | 0.022005 | 0.002319 | 0.019686 |
| ROCK2 | ENSG00000134318 | 2:11183331-11183440 | - | TE | 0.001319 | 0.10064 | -0.09932 |
| ROM1 | ENSG00000149489 | 11:62614621-62615100 | + | TE | 0.64386 | 0.10667 | 0.53719 |
| RPAIN | ENSG00000129197 | 17:5420189-5420199 | + | TS | 0.081158 | 0.61255 | -0.53139 |
| RPAP1 | ENSG00000103932 | 15:41517176-41517178 | - | TE | 0.19558 | 0.045612 | 0.14997 |
| RPE | ENSG00000197713 | 2:210021227-210021576 | + | TE | 0.002348 | 0.014323 | -0.01198 |
| RPE | ENSG00000197713 | 2:210019941-210019961 | + | TE | 0.002312 | 0.073579 | -0.07127 |
| RPH3A | ENSG00000089169 | 12:112896915-112897177 | + | TE | 0.00035 | 0.001413 | -0.00106 |
| RPH3A | ENSG00000089169 | 12:112897178-112897453 | + | TE | 0.000343 | 0.001482 | -0.00114 |
| RPH3A | ENSG00000089169 | 12:112896650-112896914 | + | TE | 0.000346 | 0.00147 | -0.00112 |
| RPL18 | ENSG00000063177 | 19:48619141-48619175 | - | TS | 0.88717 | 0.15124 | 0.73594 |
| RPL18 | ENSG00000063177 | 19:48619176-48619177 | - | TS | 0.10593 | 0.86038 | -0.75445 |
| RPL31 | ENSG00000071082 | 2:101017889-101018289 | + | TE | 0.1849 | 0.76656 | -0.58166 |
| RPL32 | ENSG00000144713 | 3:12836102-12836223 | - | TE | 0.0584 | 0.004349 | 0.054051 |
| RPL36A | ENSG00000241343 | X:101391823-101391969 | + | TE | 0.014249 | 0.13326 | -0.11901 |
| RPL5 | ENSG00000122406 | 1:92832040-92832056 | + | TS | 0.14657 | 0.004589 | 0.14198 |
| RPL6 | ENSG00000089009 | 12:112405203-112405376 | - | TE | 0.06768 | 0.01267 | 0.05501 |
| RPRD1A | ENSG00000141425 | 18:36067558-36067559 | - | TS | 0.072988 | 0.30906 | -0.23607 |
| RPS15A | ENSG00000134419 | 16:18781295-18782955 | - | TE | 0.035837 | 0.26533 | -0.22949 |
| RPS6KA1 | ENSG00000117676 | 1:26575026-26575030 | + | TE | 0.000672 | 0.043141 | -0.04247 |
| RPS6KA2 | ENSG00000071242 | 6:166412748-166412887 | - | TE | 0.009614 | 0.12938 | -0.11976 |
| RPS6KA5 | ENSG00000100784 | 14:90870849-90871503 | - | TE | 0.047017 | 0.007298 | 0.039719 |
| RPUSD1 | ENSG00000007376 | 16:785983-786193 | - | TE | 0.004892 | 0.033482 | -0.02859 |
| RPUSD3 | ENSG00000156990 | 3:9841854-9841982 | - | TE | 0.14873 | 0.024329 | 0.1244 |
| RSBN1L | ENSG00000187257 | 7:77778634-77778818 | + | TE | 0.053785 | 0.22713 | -0.17334 |
| RSF1 | ENSG00000048649 | 11:77666230-77667491 | - | TE | 0.04656 | 0.001016 | 0.045544 |
| RSPH3 | ENSG00000130363 | 6:158999782-159000166 | - | TS | 0.007144 | 0.033928 | -0.02678 |
| RSPH4A | ENSG00000111834 | 6:116616487-116617309 | + | TS | 0.51894 | 0.10106 | 0.41788 |
| RSPRY1 | ENSG00000159579 | 16:57240447-57240475 | + | TE | 0.020624 | 0.001918 | 0.018706 |
| RSPRY1 | ENSG00000159579 | 16:57238879-57239957 | + | TE | 0.002016 | 0.01038 | -0.00836 |
| RTEL1 | ENSG00000258366 | 20:63695778-63695980 | + | TE | 0.12614 | 0.60969 | -0.48355 |
| RUNX2 | ENSG00000124813 | 6:45546827-45547305 | + | TE | 0.000638 | 0.01905 | -0.01841 |
| RUSC1 | ENSG00000160753 | 1:155330730-155330730 | + | TE | 0.006268 | 0.078038 | -0.07177 |
| RUSC2 | ENSG00000198853 | 9:35561893-35561898 | + | TE | 0.03731 | 0.25188 | -0.21457 |
| S100A6 | ENSG00000197956 | 1:153535992-153536215 | - | TS | 0.10602 | 0.5052 | -0.39918 |
| SACM1L | ENSG00000211456 | 3:45743805-45743935 | + | TE | 0.002764 | 0.034451 | -0.03169 |
| SACS | ENSG00000151835 | 13:23328830-23328832 | - | TE | 0.21824 | 0.050626 | 0.16762 |
| SAMD4A | ENSG00000020577 | 14:54788916-54789127 | + | TE | 0.16767 | 0.014835 | 0.15283 |
| SAMHD1 | ENSG00000101347 | 20:36892932-36893066 | - | TE | 0.000625 | 0.035696 | -0.03507 |
| SAMM50 | ENSG00000100347 | 22:43996377-43996529 | + | TE | 0.070283 | 0.28886 | -0.21858 |
| SART1 | ENSG00000175467 | 11:65961728-65962093 | + | TS | 0.58698 | 0.10286 | 0.48412 |
| SBF2 | ENSG00000133812 | 11:9779154-9780516 | - | TE | 0.001093 | 0.038562 | -0.03747 |
| SC5D | ENSG00000109929 | 11:121307844-121308633 | + | TE | 0.002498 | 0.041426 | -0.03893 |
| SC5D | ENSG00000109929 | 11:121307057-121307843 | + | TE | 0.002524 | 0.01851 | -0.01599 |
| SCAF11 | ENSG00000139218 | 12:45990566-45990576 | - | TS | 0.27205 | 0.062418 | 0.20963 |
| SCAMP5 | ENSG00000198794 | 15:75018789-75019061 | + | TE | 0.1232 | 0.027303 | 0.095894 |
| SCARB2 | ENSG00000138760 | 4:76172700-76172727 | - | TE | 0.020379 | 0.099569 | -0.07919 |
| SCFD2 | ENSG00000184178 | 4:53366062-53366075 | - | TS | 0.020567 | 0.13902 | -0.11845 |
| SCMH1 | ENSG00000010803 | 1:41027202-41028191 | - | TE | 0.021517 | 0.13022 | -0.1087 |
| SCN1B | ENSG00000105711 | 19:35037698-35037720 | + | TE | 0.02864 | 0.14058 | -0.11194 |
| SCN9A | ENSG00000169432 | 2:166198611-166199864 | - | TE | 0.00963 | 0.054708 | -0.04508 |
| SCNM1 | ENSG00000163156 | 1:151168986-151169111 | + | TE | 0.011014 | 0.12989 | -0.11887 |
| SCRN1 | ENSG00000136193 | 7:29923562-29923673 | - | TE | 0.071997 | 0.004194 | 0.067803 |
| SCRN3 | ENSG00000144306 | 2:174427865-174427895 | + | TE | 0.013433 | 0.060556 | -0.04712 |
| SCUBE2 | ENSG00000175356 | 11:9019498-9020444 | - | TE | 0.8986 | 0.17944 | 0.71917 |
| SCUBE2 | ENSG00000175356 | 11:9020890-9021197 | - | TE | 0.097778 | 0.82087 | -0.72309 |
| SCYL2 | ENSG00000136021 | 12:100338528-100338633 | + | TE | 0.07016 | 0.000535 | 0.069625 |
| SDAD1 | ENSG00000198301 | 4:75949906-75949915 | - | TE | 0.006698 | 0.04918 | -0.04248 |
| SDC1 | ENSG00000115884 | 2:20225097-20225112 | - | TS | 0.4746 | 0.042416 | 0.43219 |
| SDF4 | ENSG00000078808 | 1:1216932-1216933 | - | TE | 0.009171 | 0.046166 | -0.037 |
| SDR39U1 | ENSG00000100445 | 14:24439768-24439779 | - | TE | 0.043314 | 0.002627 | 0.040688 |
| SEC11C | ENSG00000166562 | 18:59139477-59139865 | + | TS | 0.07908 | 0.49802 | -0.41894 |
| SEC14L1 | ENSG00000129657 | 17:77140932-77140978 | + | TS | 0.059707 | 0.27266 | -0.21295 |
| SEC31A | ENSG00000138674 | 4:82819022-82819037 | - | TE | 0.031214 | 0.003531 | 0.027682 |
| SEC31B | ENSG00000075826 | 10:100486647-100487795 | - | TE | 0.85094 | 0.13727 | 0.71367 |
| SEC31B | ENSG00000075826 | 10:100486642-100486646 | - | TE | 0.14957 | 0.86273 | -0.71316 |
| SEC61A2 | ENSG00000065665 | 10:12169258-12169956 | + | TE | 0.010136 | 0.13341 | -0.12327 |
| SELENBP1 | ENSG00000143416 | 1:151364309-151364705 | - | TE | 0.27483 | 0.055714 | 0.21912 |
| SELENOF | ENSG00000183291 | 1:86863301-86863605 | - | TE | 0.000298 | 0.02037 | -0.02007 |
| SELP | ENSG00000174175 | 1:169588852-169589076 | - | TE | 0.23886 | 0.00591 | 0.23295 |
| SELPLG | ENSG00000110876 | 12:108622829-108624312 | - | TE | 7.48E-05 | 0.009096 | -0.00902 |
| SEMA4B | ENSG00000185033 | 15:90229661-90229670 | + | TE | 0.000708 | 0.004 | -0.00329 |
| SEMA4C | ENSG00000168758 | 2:96859742-96859747 | - | TE | 0.2998 | 0.040706 | 0.25909 |
| SEMA4D | ENSG00000187764 | 9:89361601-89361978 | - | TE | 0.000717 | 0.007101 | -0.00638 |
| SEMA6A | ENSG00000092421 | 5:116446134-116446463 | - | TE | 0.11242 | 0.022005 | 0.09042 |
| SENP3 | ENSG00000161956 | 17:7571373-7571686 | + | TE | 0.028882 | 0.12585 | -0.09697 |
| SEPHS1 | ENSG00000086475 | 10:13317424-13317438 | - | TE | 0.008338 | 0.047654 | -0.03932 |
| Sept2 | ENSG00000168385 | 2:241351967-241352109 | + | TE | 0.000456 | 0.022671 | -0.02222 |
| Sept2 | ENSG00000168385 | 2:241336099-241336336 | + | TE | 0.10443 | 0.021616 | 0.082809 |
| Sept3 | ENSG00000100167 | 22:41996902-41998066 | + | TE | 0.06983 | 0.30743 | -0.2376 |
| Sept9 | ENSG00000184640 | 17:77498825-77498941 | + | TE | 0.01098 | 0.00111 | 0.00987 |
| SERBP1 | ENSG00000142864 | 1:67412867-67413215 | - | TE | 0.000353 | 0.006338 | -0.00599 |
| SERGEF | ENSG00000129158 | 11:17788049-17788051 | - | TE | 0.32181 | 0.053985 | 0.26783 |
| SERPINB6 | ENSG00000124570 | 6:2948245-2948699 | - | TE | 0.003126 | 0.036843 | -0.03372 |
| SERPING1 | ENSG00000149131 | 11:57614328-57614529 | + | TE | 0.001384 | 0.062647 | -0.06126 |
| SERPING1 | ENSG00000149131 | 11:57614582-57614739 | + | TE | 0.001385 | 0.017806 | -0.01642 |
| SET | ENSG00000119335 | 9:128694665-128694894 | + | TE | 0.001036 | 0.022954 | -0.02192 |
| SGF29 | ENSG00000176476 | 16:28553920-28553925 | + | TS | 0.23505 | 0.055582 | 0.17947 |
| SGK1 | ENSG00000118515 | 6:134169353-134169880 | - | TE | 0.001829 | 0.027611 | -0.02578 |
| SGSM3 | ENSG00000100359 | 22:40409812-40410103 | + | TE | 0.21229 | 0.047708 | 0.16458 |
| SH3BP2 | ENSG00000087266 | 4:2812219-2812300 | + | TS | 0.63724 | 0.11642 | 0.52082 |
| SIGLEC15 | ENSG00000197046 | 18:45842106-45842224 | + | TE | 0.006726 | 0.24196 | -0.23523 |
| SIGLEC7 | ENSG00000168995 | 19:51142307-51142369 | + | TS | 0.11517 | 0.024252 | 0.090916 |
| SIVA1 | ENSG00000184990 | 14:104756761-104756925 | + | TE | 0.37029 | 0.07125 | 0.29904 |
| SIVA1 | ENSG00000184990 | 14:104759658-104759659 | + | TE | 0.093455 | 0.52601 | -0.43255 |
| SIX5 | ENSG00000177045 | 19:45768852-45769226 | - | TS | 0.51545 | 0.063105 | 0.45235 |
| SKA2 | ENSG00000182628 | 17:59111943-59111955 | - | TE | 0.024862 | 0.005208 | 0.019654 |
| SKA2 | ENSG00000182628 | 17:59111956-59112046 | - | TE | 0.024208 | 0.005295 | 0.018913 |
| SLBP | ENSG00000163950 | 4:1712318-1712318 | - | TS | 0.044007 | 0.2721 | -0.2281 |
| SLC11A1 | ENSG00000018280 | 2:218396027-218396027 | + | TE | 0.002692 | 0.031375 | -0.02868 |
| SLC11A2 | ENSG00000110911 | 12:50986071-50986080 | - | TE | 0.00735 | 0.048245 | -0.0409 |
| SLC12A4 | ENSG00000124067 | 16:67944485-67944592 | - | TE | 0.044896 | 0.009233 | 0.035662 |
| SLC12A5 | ENSG00000124140 | 20:46060151-46060152 | + | TE | 0.19893 | 0.83685 | -0.63793 |
| SLC12A5 | ENSG00000124140 | 20:46059599-46060150 | + | TE | 0.80156 | 0.16558 | 0.63599 |
| SLC14A1 | ENSG00000141469 | 18:45749778-45749951 | + | TE | 0.034447 | 0.15387 | -0.11942 |
| SLC14A1 | ENSG00000141469 | 18:45750331-45750335 | + | TE | 0.007468 | 0.043508 | -0.03604 |
| SLC15A4 | ENSG00000139370 | 12:128793895-128794172 | - | TE | 0.000941 | 0.010974 | -0.01003 |
| SLC17A9 | ENSG00000101194 | 20:62952647-62952706 | + | TS | 0.87479 | 0.067349 | 0.80744 |
| SLC17A9 | ENSG00000101194 | 20:62952707-62952710 | + | TS | 0.12549 | 0.85475 | -0.72925 |
| SLC19A2 | ENSG00000117479 | 1:169463932-169464047 | - | TE | 0.25609 | 0.028154 | 0.22793 |
| SLC19A2 | ENSG00000117479 | 1:169463931-169463931 | - | TE | 0.12449 | 0.021461 | 0.10303 |
| SLC1A2 | ENSG00000110436 | 11:35260239-35260685 | - | TE | 0.13945 | 0.74384 | -0.60439 |
| SLC1A3 | ENSG00000079215 | 5:36606439-36606610 | + | TS | 0.84366 | 0.080571 | 0.76309 |
| SLC1A3 | ENSG00000079215 | 5:36686065-36686233 | + | TE | 0.14924 | 0.00995 | 0.13929 |
| SLC1A3 | ENSG00000079215 | 5:36606617-36606735 | + | TS | 0.15939 | 0.92089 | -0.7615 |
| SLC20A1 | ENSG00000144136 | 2:112662864-112663825 | + | TE | 0.11726 | 0.49622 | -0.37896 |
| SLC22A18 | ENSG00000110628 | 11:2925241-2925241 | + | TE | 0.094704 | 0.018367 | 0.076337 |
| SLC24A1 | ENSG00000074621 | 15:65624081-65625970 | + | TS | 0.19512 | 0.010698 | 0.18442 |
| SLC25A13 | ENSG00000004864 | 7:96322094-96322147 | - | TS | 0.13774 | 0.80268 | -0.66494 |
| SLC25A13 | ENSG00000004864 | 7:96321942-96322093 | - | TS | 0.85754 | 0.19652 | 0.66103 |
| SLC25A3 | ENSG00000075415 | 12:98601708-98601708 | + | TE | 0.12894 | 0.002334 | 0.12661 |
| SLC25A43 | ENSG00000077713 | X:119399060-119399379 | + | TS | 0.09162 | 0.83015 | -0.73853 |
| SLC25A43 | ENSG00000077713 | X:119399380-119399678 | + | TS | 0.90793 | 0.17 | 0.73793 |
| SLC25A46 | ENSG00000164209 | 5:110753302-110755119 | + | TS | 0.032366 | 0.005612 | 0.026753 |
| SLC26A7 | ENSG00000147606 | 8:91395062-91395649 | + | TE | 0.76952 | 0.10814 | 0.66138 |
| SLC27A3 | ENSG00000143554 | 1:153780125-153780151 | + | TE | 0.13259 | 0.88162 | -0.74903 |
| SLC27A3 | ENSG00000143554 | 1:153779931-153780124 | + | TE | 0.85198 | 0.1141 | 0.73788 |
| SLC2A4 | ENSG00000181856 | 17:7286426-7287833 | + | TE | 0.078407 | 0.010691 | 0.067715 |
| SLC33A1 | ENSG00000169359 | 3:155826526-155826819 | - | TE | 0.1159 | 0.013884 | 0.10201 |
| SLC35A3 | ENSG00000117620 | 1:100023943-100024014 | + | TE | 0.059971 | 0.012135 | 0.047836 |
| SLC35B2 | ENSG00000157593 | 6:44254102-44254105 | - | TE | 0.016898 | 0.090098 | -0.0732 |
| SLC35C1 | ENSG00000181830 | 11:45810776-45811558 | + | TE | 0.029936 | 0.004802 | 0.025134 |
| SLC35C2 | ENSG00000080189 | 20:46349528-46349529 | - | TE | 0.015831 | 0.12275 | -0.10692 |
| SLC35E2A | ENSG00000215790 | 1:1724838-1724839 | - | TE | 0.37939 | 0.028334 | 0.35106 |
| SLC35E2B | ENSG00000189339 | 1:1692645-1692709 | - | TS | 0.15792 | 0.020433 | 0.13748 |
| SLC38A2 | ENSG00000134294 | 12:46372772-46372773 | - | TS | 0.044618 | 0.005527 | 0.039091 |
| SLC38A9 | ENSG00000177058 | 5:55625847-55625857 | - | TE | 0.29288 | 0.025284 | 0.26759 |
| SLC3A2 | ENSG00000168003 | 11:62856111-62856143 | + | TS | 0.61446 | 0.10354 | 0.51092 |
| SLC47A2 | ENSG00000180638 | 17:19678319-19678520 | - | TE | 0.8563 | 0.097144 | 0.75915 |
| SLC47A2 | ENSG00000180638 | 17:19678288-19678314 | - | TE | 0.14658 | 0.90379 | -0.75721 |
| SLC48A1 | ENSG00000211584 | 12:47782218-47782751 | + | TE | 0.29398 | 0.022682 | 0.2713 |
| SLC6A6 | ENSG00000131389 | 3:14402620-14402620 | + | TS | 0.022108 | 0.4466 | -0.42449 |
| SLC6A6 | ENSG00000131389 | 3:14402576-14402583 | + | TS | 0.023252 | 0.19864 | -0.17538 |
| SLC8A1 | ENSG00000183023 | 2:40115045-40115112 | - | TE | 0.020108 | 0.001462 | 0.018646 |
| SLC8A3 | ENSG00000100678 | 14:70046075-70046323 | - | TE | 0.038789 | 0.264 | -0.22521 |
| SLC9A6 | ENSG00000198689 | X:136047131-136047261 | + | TE | 0.002286 | 0.01062 | -0.00833 |
| SLC9A7 | ENSG00000065923 | X:46605318-46607203 | - | TE | 0.003301 | 0.014491 | -0.01119 |
| SLF2 | ENSG00000119906 | 10:100912569-100912962 | + | TS | 0.051884 | 0.005576 | 0.046308 |
| SLFN5 | ENSG00000166750 | 17:35265072-35266360 | + | TE | 0.00026 | 0.003514 | -0.00325 |
| SLU7 | ENSG00000164609 | 5:160403153-160403464 | - | TE | 0.03823 | 0.007906 | 0.030324 |
| SMAD2 | ENSG00000175387 | 18:47930800-47930842 | - | TS | 0.069625 | 0.28875 | -0.21912 |
| SMARCA4 | ENSG00000127616 | 19:11062280-11062282 | + | TE | 0.069262 | 0.010104 | 0.059158 |
| SMARCAL1 | ENSG00000138375 | 2:216483018-216483053 | + | TE | 0.43558 | 0.089444 | 0.34614 |
| SMARCB1 | ENSG00000099956 | 22:23834354-23834358 | + | TE | 0.12311 | 0.013586 | 0.10952 |
| SMARCC2 | ENSG00000139613 | 12:56163345-56163765 | - | TE | 0.009911 | 0.064565 | -0.05465 |
| SMARCD2 | ENSG00000108604 | 17:63832502-63832603 | - | TE | 0.06975 | 0.011937 | 0.057813 |
| SMARCE1 | ENSG00000073584 | 17:40627743-40627743 | - | TE | 0.016923 | 0.003384 | 0.013539 |
| SMC6 | ENSG00000163029 | 2:17665447-17665613 | - | TE | 0.018156 | 0.14175 | -0.12359 |
| SMG6 | ENSG00000070366 | 17:2061405-2061622 | - | TE | 0.077081 | 0.00807 | 0.069011 |
| SMIM1 | ENSG00000235169 | 1:3775795-3775908 | + | TE | 0.064476 | 0.25967 | -0.19519 |
| SMIM30 | ENSG00000214194 | 7:113116718-113116860 | - | TE | 0.026951 | 0.12587 | -0.09892 |
| SMUG1 | ENSG00000123415 | 12:54181468-54181470 | - | TE | 0.11245 | 0.007175 | 0.10528 |
| SNAP23 | ENSG00000092531 | 15:42531828-42533059 | + | TE | 0.017285 | 0.077373 | -0.06009 |
| SNAP23 | ENSG00000092531 | 15:42531479-42531651 | + | TE | 0.00392 | 0.000668 | 0.003251 |
| SND1 | ENSG00000197157 | 7:128091993-128092607 | + | TE | 0.29443 | 0.010012 | 0.28441 |
| SNRNP35 | ENSG00000184209 | 12:123465538-123466299 | + | TE | 0.019414 | 0.10694 | -0.08752 |
| SNRNP70 | ENSG00000104852 | 19:49108604-49108605 | + | TE | 0.036795 | 0.15318 | -0.11638 |
| SNRPG | ENSG00000143977 | 2:70281505-70281506 | - | TE | 0.68165 | 0.07126 | 0.61039 |
| SNTB1 | ENSG00000172164 | 8:120538521-120538969 | - | TE | 0.001209 | 0.021706 | -0.0205 |
| SNX10 | ENSG00000086300 | 7:26372747-26372944 | + | TE | 0.000168 | 0.058366 | -0.0582 |
| SNX14 | ENSG00000135317 | 6:85505798-85505798 | - | TE | 0.090737 | 0.5482 | -0.45746 |
| SNX14 | ENSG00000135317 | 6:85505802-85505805 | - | TE | 0.091668 | 0.39257 | -0.3009 |
| SNX14 | ENSG00000135317 | 6:85505806-85506005 | - | TE | 0.71294 | 0.074287 | 0.63866 |
| SNX18 | ENSG00000178996 | 5:54517865-54519573 | + | TS | 0.00517 | 0.027897 | -0.02273 |
| SNX2 | ENSG00000205302 | 5:122829672-122829755 | + | TE | 0.009959 | 0.069974 | -0.06002 |
| SNX21 | ENSG00000124104 | 20:45840781-45840907 | + | TE | 0.011141 | 0.23424 | -0.2231 |
| SOS2 | ENSG00000100485 | 14:50231310-50231558 | - | TS | 0.032449 | 0.13719 | -0.10475 |
| SP1 | ENSG00000185591 | 12:53410927-53411630 | + | TE | 0.001882 | 0.019687 | -0.01781 |
| SP3 | ENSG00000172845 | 2:173908532-173909940 | - | TE | 0.067427 | 0.011891 | 0.055536 |
| SPAG9 | ENSG00000008294 | 17:50965627-50965643 | - | TE | 0.000459 | 0.001937 | -0.00148 |
| SPAST | ENSG00000021574 | 2:32063962-32063967 | + | TS | 0.005626 | 0.070534 | -0.06491 |
| SPATS2L | ENSG00000196141 | 2:200478210-200478529 | + | TE | 0.005532 | 0.059605 | -0.05407 |
| SPECC1 | ENSG00000128487 | 17:20313976-20314138 | + | TE | 0.009452 | 0.069571 | -0.06012 |
| SPECC1L | ENSG00000100014 | 22:24414753-24414773 | + | TE | 0.014631 | 0.002638 | 0.011993 |
| SPG11 | ENSG00000104133 | 15:44563121-44563301 | - | TE | 0.003651 | 0.072215 | -0.06856 |
| SPG7 | ENSG00000197912 | 16:89543576-89543684 | + | TS | 0.047721 | 0.009811 | 0.03791 |
| SPIDR | ENSG00000164808 | 8:47735914-47735916 | + | TE | 0.019686 | 0.000635 | 0.019051 |
| SPIN3 | ENSG00000204271 | X:56990946-56990953 | - | TE | 0.027684 | 0.13298 | -0.1053 |
| SPOCK1 | ENSG00000152377 | 5:136978454-136978844 | - | TE | 0.13139 | 0.032341 | 0.099044 |
| SPP1 | ENSG00000118785 | 4:87983327-87983410 | + | TE | 0.49928 | 0.12364 | 0.37564 |
| SPP1 | ENSG00000118785 | 4:87983128-87983297 | + | TE | 0.12602 | 0.87618 | -0.75016 |
| SQSTM1 | ENSG00000161011 | 5:179813025-179813598 | + | TE | 0.007954 | 0.042822 | -0.03487 |
| SREK1 | ENSG00000153914 | 5:66180812-66180879 | + | TE | 0.055893 | 0.003321 | 0.052572 |
| SRGAP3 | ENSG00000196220 | 3:9248992-9249213 | - | TS | 0.078071 | 0.32301 | -0.24493 |
| SRP19 | ENSG00000153037 | 5:112867404-112867724 | + | TE | 0.005656 | 0.058459 | -0.0528 |
| SRP54 | ENSG00000100883 | 14:35019359-35019391 | + | TE | 0.10558 | 0.88679 | -0.78121 |
| SRP54 | ENSG00000100883 | 14:35019075-35019358 | + | TE | 0.89142 | 0.10607 | 0.78535 |
| SRPK1 | ENSG00000096063 | 6:35835428-35835488 | - | TE | 0.000285 | 0.062488 | -0.0622 |
| SRPK2 | ENSG00000135250 | 7:105117526-105118022 | - | TE | 0.000992 | 0.008518 | -0.00753 |
| SRRM2 | ENSG00000167978 | 16:2760500-2760621 | + | TE | 0.1445 | 0.019432 | 0.12507 |
| SRSF11 | ENSG00000116754 | 1:70250611-70250725 | + | TE | 0.082056 | 0.001162 | 0.080894 |
| SRSF11 | ENSG00000116754 | 1:70250947-70251617 | + | TE | 0.001282 | 0.030762 | -0.02948 |
| SRSF2 | ENSG00000161547 | 17:76734119-76734146 | - | TE | 0.008083 | 0.001918 | 0.006165 |
| SRSF7 | ENSG00000115875 | 2:38743601-38744716 | - | TE | 0.002947 | 0.031436 | -0.02849 |
| SSBP1 | ENSG00000106028 | 7:141738365-141738395 | + | TS | 0.046243 | 0.010005 | 0.036239 |
| SSBP2 | ENSG00000145687 | 5:81419873-81419917 | - | TE | 0.057662 | 0.001772 | 0.05589 |
| SSBP3 | ENSG00000157216 | 1:54226639-54226919 | - | TE | 0.062605 | 0.009232 | 0.053373 |
| SSH2 | ENSG00000141298 | 17:29630602-29630748 | - | TE | 0.020847 | 0.002568 | 0.018279 |
| SSH3 | ENSG00000172830 | 11:67312606-67312607 | + | TE | 0.12338 | 0.51113 | -0.38774 |
| SSH3 | ENSG00000172830 | 11:67312383-67312592 | + | TE | 0.87241 | 0.11083 | 0.76158 |
| SSR4 | ENSG00000180879 | X:153798498-153798498 | + | TE | 0.20713 | 0.033962 | 0.17317 |
| ST3GAL1 | ENSG00000008513 | 8:133571889-133571925 | - | TS | 0.01449 | 0.063085 | -0.0486 |
| ST3GAL6 | ENSG00000064225 | 3:98794363-98794393 | + | TE | 0.04681 | 0.009524 | 0.037286 |
| STAC | ENSG00000144681 | 3:36380344-36380502 | + | TS | 0.033236 | 0.14606 | -0.11282 |
| STAT4 | ENSG00000138378 | 2:191029580-191029866 | - | TE | 0.18594 | 0.027432 | 0.1585 |
| STAT5A | ENSG00000126561 | 17:42310507-42310860 | + | TE | 0.055579 | 0.001873 | 0.053706 |
| STAU2 | ENSG00000040341 | 8:73653658-73653896 | - | TE | 0.1182 | 0.019698 | 0.098507 |
| STEAP3 | ENSG00000115107 | 2:119263057-119263698 | + | TE | 0.10854 | 0.018093 | 0.090445 |
| STEAP4 | ENSG00000127954 | 7:88278028-88279628 | - | TE | 3.07E-05 | 0.050185 | -0.05015 |
| STIM2 | ENSG00000109689 | 4:27023967-27024187 | + | TE | 0.010948 | 0.001873 | 0.009075 |
| STK17B | ENSG00000081320 | 2:196137447-196137729 | - | TE | 0.002784 | 0.000104 | 0.002679 |
| STK25 | ENSG00000115694 | 2:241495249-241495537 | - | TE | 0.060663 | 0.005479 | 0.055183 |
| STK25 | ENSG00000115694 | 2:241495017-241495018 | - | TE | 0.006497 | 0.049038 | -0.04254 |
| STMN3 | ENSG00000197457 | 20:63641086-63641397 | - | TE | 0.17377 | 0.00821 | 0.16556 |
| STMN3 | ENSG00000197457 | 20:63639705-63639707 | - | TE | 0.051155 | 0.40756 | -0.35641 |
| STOML1 | ENSG00000067221 | 15:73983220-73983936 | - | TE | 0.033599 | 0.21589 | -0.18229 |
| STOML2 | ENSG00000165283 | 9:35099776-35099890 | - | TE | 0.61533 | 0.14354 | 0.47179 |
| STRIP1 | ENSG00000143093 | 1:110054640-110054641 | + | TE | 0.031761 | 0.27101 | -0.23925 |
| STX16 | ENSG00000124222 | 20:58652007-58652087 | + | TS | 0.004862 | 0.032497 | -0.02764 |
| STX16 | ENSG00000124222 | 20:58676187-58676328 | + | TE | 0.000158 | 0.042017 | -0.04186 |
| STX17 | ENSG00000136874 | 9:99968822-99969540 | + | TE | 0.010391 | 0.05802 | -0.04763 |
| STX3 | ENSG00000166900 | 11:59800855-59800962 | + | TE | 0.13982 | 0.000389 | 0.13943 |
| STX4 | ENSG00000103496 | 16:31040013-31040163 | + | TE | 0.033259 | 0.14674 | -0.11348 |
| SUCLG1 | ENSG00000163541 | 2:84423523-84423529 | - | TE | 0.013579 | 0.1674 | -0.15382 |
| SUCO | ENSG00000094975 | 1:172533383-172533497 | + | TS | 0.6917 | 0.11771 | 0.57399 |
| SULT1A1 | ENSG00000196502 | 16:28605628-28605933 | - | TE | 0.47278 | 0.10931 | 0.36347 |
| SUMF2 | ENSG00000129103 | 7:56080639-56080665 | + | TE | 0.011165 | 0.07512 | -0.06396 |
| SUN1 | ENSG00000164828 | 7:873215-874899 | + | TE | 0.27238 | 0.00657 | 0.26581 |
| SUPT7L | ENSG00000119760 | 2:27653052-27653062 | - | TE | 0.11629 | 0.010486 | 0.10581 |
| SUZ12 | ENSG00000178691 | 17:31998658-31999376 | + | TE | 0.003632 | 0.048939 | -0.04531 |
| SWT1 | ENSG00000116668 | 1:185290674-185291007 | + | TE | 0.035745 | 0.003044 | 0.032702 |
| SYNE1 | ENSG00000131018 | 6:152121769-152121779 | - | TE | 0.013672 | 0.001811 | 0.011861 |
| SYNJ2 | ENSG00000078269 | 6:158095618-158096463 | + | TE | 0.053388 | 0.003238 | 0.05015 |
| SYNRG | ENSG00000275066 | 17:37518887-37519009 | - | TE | 0.001333 | 0.029963 | -0.02863 |
| SYTL2 | ENSG00000137501 | 11:85694726-85694741 | - | TE | 0.012159 | 0.075944 | -0.06378 |
| SYTL2 | ENSG00000137501 | 11:85694288-85694290 | - | TE | 0.29392 | 0.038279 | 0.25564 |
| SZT2 | ENSG00000198198 | 1:43445252-43445636 | + | TS | 0.10569 | 0.43417 | -0.32848 |
| SZT2 | ENSG00000198198 | 1:43450728-43450908 | + | TE | 0.032021 | 0.003231 | 0.02879 |
| TADA2B | ENSG00000173011 | 4:7054432-7056641 | + | TE | 0.000559 | 0.010723 | -0.01016 |
| TAF11 | ENSG00000064995 | 6:34877778-34878720 | - | TE | 0.09016 | 0.007436 | 0.082724 |
| TAF1C | ENSG00000103168 | 16:84187050-84187052 | - | TS | 0.31991 | 0.067559 | 0.25235 |
| TANK | ENSG00000136560 | 2:161236173-161236187 | + | TE | 0.06387 | 0.002172 | 0.061698 |
| TAP2 | ENSG00000204267 | 6:32828944-32829034 | - | TE | 0.000191 | 0.077129 | -0.07694 |
| TAS2R14 | ENSG00000212127 | 12:10937408-10938577 | - | TE | 0.15892 | 0.036251 | 0.12267 |
| TAX1BP3 | ENSG00000213977 | 17:3662902-3663885 | - | TE | 0.054674 | 0.37671 | -0.32204 |
| TBC1D10B | ENSG00000169221 | 16:30357102-30357106 | - | TE | 0.004142 | 0.043592 | -0.03945 |
| TBC1D17 | ENSG00000104946 | 19:49877666-49877701 | + | TS | 0.48751 | 0.10522 | 0.38229 |
| TBCK | ENSG00000145348 | 4:106046570-106046680 | - | TE | 0.007856 | 0.065364 | -0.05751 |
| TBRG1 | ENSG00000154144 | 11:124632239-124632405 | + | TE | 0.00888 | 0.002023 | 0.006857 |
| TBRG1 | ENSG00000154144 | 11:124632093-124632238 | + | TE | 0.26497 | 0.064951 | 0.20002 |
| TCAF1 | ENSG00000198420 | 7:143853696-143853827 | - | TE | 0.026633 | 0.18116 | -0.15453 |
| TCEANC2 | ENSG00000116205 | 1:54104295-54106077 | + | TE | 0.14669 | 0.00875 | 0.13794 |
| TCERG1 | ENSG00000113649 | 5:146511506-146511508 | + | TE | 0.15616 | 0.015351 | 0.14081 |
| TCERG1 | ENSG00000113649 | 5:146480095-146480216 | + | TE | 0.025047 | 0.12197 | -0.09692 |
| TCF3 | ENSG00000071564 | 19:1609472-1609599 | - | TE | 0.61724 | 0.13034 | 0.4869 |
| TCF3 | ENSG00000071564 | 19:1609293-1609471 | - | TE | 0.09763 | 0.56522 | -0.46759 |
| TCOF1 | ENSG00000070814 | 5:150400294-150400296 | + | TE | 0.019429 | 0.14076 | -0.12133 |
| TCP11L1 | ENSG00000176148 | 11:33072474-33072845 | + | TE | 0.058362 | 0.012354 | 0.046008 |
| TCTN1 | ENSG00000204852 | 12:110614106-110614106 | + | TS | 0.90043 | 0.13982 | 0.76062 |
| TCTN1 | ENSG00000204852 | 12:110614027-110614101 | + | TS | 0.10635 | 0.85677 | -0.75042 |
| TCTN3 | ENSG00000119977 | 10:95693924-95693926 | - | TS | 0.12342 | 0.49834 | -0.37492 |
| TCTN3 | ENSG00000119977 | 10:95663396-95663400 | - | TE | 0.15181 | 0.017134 | 0.13468 |
| TDRP | ENSG00000180190 | 8:489792-489802 | - | TE | 0.15304 | 0.026428 | 0.12661 |
| TESPA1 | ENSG00000135426 | 12:54950170-54950353 | - | TE | 0.024738 | 0.005456 | 0.019281 |
| TEX264 | ENSG00000164081 | 3:51704019-51704317 | + | TE | 0.70464 | 0.13594 | 0.5687 |
| TFAP2E | ENSG00000116819 | 1:35594394-35595261 | + | TE | 0.013149 | 0.13722 | -0.12407 |
| TFDP2 | ENSG00000114126 | 3:141952496-141952696 | - | TE | 0.001912 | 0.020103 | -0.01819 |
| TFR2 | ENSG00000106327 | 7:100620420-100621126 | - | TE | 0.21763 | 0.97731 | -0.75968 |
| TFR2 | ENSG00000106327 | 7:100620416-100620419 | - | TE | 0.78236 | 0.022277 | 0.76008 |
| TGFBI | ENSG00000120708 | 5:136063804-136063815 | + | TE | 0.007299 | 0.032339 | -0.02504 |
| TGOLN2 | ENSG00000152291 | 2:85328025-85328251 | - | TS | 0.51301 | 0.076586 | 0.43643 |
| THAP5 | ENSG00000177683 | 7:108564993-108565105 | - | TE | 0.021798 | 0.001141 | 0.020657 |
| THAP5 | ENSG00000177683 | 7:108564570-108564992 | - | TE | 0.014494 | 0.072567 | -0.05807 |
| THUMPD3 | ENSG00000134077 | 3:9384871-9385702 | + | TE | 0.002018 | 0.01156 | -0.00954 |
| TIAL1 | ENSG00000151923 | 10:119575412-119575424 | - | TE | 0.019845 | 0.12773 | -0.10788 |
| TIAM2 | ENSG00000146426 | 6:155257191-155257200 | + | TE | 0.015316 | 0.083053 | -0.06774 |
| TIAM2 | ENSG00000146426 | 6:155256484-155257190 | + | TE | 0.015551 | 0.071619 | -0.05607 |
| TIMM17B | ENSG00000126768 | X:48893447-48893448 | - | TE | 0.42206 | 0.072338 | 0.34972 |
| TIMMDC1 | ENSG00000113845 | 3:119498532-119498546 | + | TS | 0.19544 | 0.012963 | 0.18248 |
| TINF2 | ENSG00000092330 | 14:24242664-24242674 | - | TS | 0.17083 | 0.040873 | 0.12996 |
| TIPARP | ENSG00000163659 | 3:156706751-156706766 | + | TE | 0.001584 | 0.014542 | -0.01296 |
| TIPARP | ENSG00000163659 | 3:156704684-156705120 | + | TE | 0.001608 | 0.025844 | -0.02424 |
| TIPRL | ENSG00000143155 | 1:168178933-168179054 | + | TS | 0.28001 | 0.069578 | 0.21043 |
| TK2 | ENSG00000166548 | 16:66511632-66511683 | - | TE | 0.019723 | 0.000999 | 0.018724 |
| TLE3 | ENSG00000140332 | 15:70050097-70050204 | - | TE | 0.000818 | 0.18076 | -0.17995 |
| TLL2 | ENSG00000095587 | 10:96513511-96513911 | - | TS | 0.55433 | 0.11668 | 0.43765 |
| TMA16 | ENSG00000198498 | 4:163494708-163494804 | + | TS | 0.10012 | 0.52789 | -0.42777 |
| TMED9 | ENSG00000184840 | 5:177592158-177592216 | + | TS | 0.065816 | 0.50143 | -0.43562 |
| TMEM11 | ENSG00000178307 | 17:21214091-21214161 | - | TS | 0.12929 | 0.87866 | -0.74937 |
| TMEM11 | ENSG00000178307 | 17:21214162-21214163 | - | TS | 0.87099 | 0.12327 | 0.74771 |
| TMEM123 | ENSG00000152558 | 11:102452553-102452839 | - | TS | 0.011989 | 0.051603 | -0.03962 |
| TMEM126B | ENSG00000171204 | 11:85636503-85636507 | + | TE | 0.31357 | 0.049045 | 0.26452 |
| TMEM134 | ENSG00000172663 | 11:67469244-67469250 | - | TS | 0.15857 | 0.88723 | -0.72866 |
| TMEM134 | ENSG00000172663 | 11:67469251-67469272 | - | TS | 0.83584 | 0.11467 | 0.72117 |
| TMEM138 | ENSG00000149483 | 11:61362361-61362386 | + | TS | 0.1213 | 0.50195 | -0.38064 |
| TMEM138 | ENSG00000149483 | 11:61367999-61368556 | + | TE | 0.2883 | 0.066507 | 0.22179 |
| TMEM144 | ENSG00000164124 | 4:158210485-158210554 | + | TS | 0.49153 | 0.076653 | 0.41488 |
| TMEM144 | ENSG00000164124 | 4:158255205-158255287 | + | TE | 0.32254 | 0.061875 | 0.26066 |
| TMEM147 | ENSG00000105677 | 19:35547523-35547523 | + | TE | 0.10867 | 0.024658 | 0.084008 |
| TMEM169 | ENSG00000163449 | 2:216101066-216102382 | + | TE | 0.048175 | 0.011795 | 0.03638 |
| TMEM175 | ENSG00000127419 | 4:957824-958652 | + | TE | 0.030319 | 0.29875 | -0.26843 |
| TMEM184B | ENSG00000198792 | 22:38219312-38219323 | - | TE | 0.001543 | 0.03876 | -0.03722 |
| TMEM189 | ENSG00000240849 | 20:50123737-50123769 | - | TE | 0.018784 | 0.003327 | 0.015457 |
| TMEM191C | ENSG00000206140 | 22:21469642-21469933 | + | TE | 0.88332 | 0.10214 | 0.78117 |
| TMEM191C | ENSG00000206140 | 22:21469934-21469935 | + | TE | 0.11817 | 0.89786 | -0.7797 |
| TMEM205 | ENSG00000105518 | 19:11342779-11342784 | - | TE | 0.70572 | 0.16775 | 0.53798 |
| TMEM220 | ENSG00000187824 | 17:10715422-10715425 | - | TE | 0.00186 | 0.16447 | -0.16261 |
| TMEM231 | ENSG00000205084 | 16:75538151-75538349 | - | TE | 0.66941 | 0.091776 | 0.57764 |
| TMEM250 | ENSG00000238227 | 9:136114581-136114588 | - | TE | 0.019265 | 0.27309 | -0.25383 |
| TMEM251 | ENSG00000153485 | 14:93186260-93187086 | + | TE | 0.009029 | 0.13096 | -0.12193 |
| TMEM256 | ENSG00000205544 | 17:7402978-7403209 | - | TE | 0.85301 | 0.10364 | 0.74937 |
| TMEM256 | ENSG00000205544 | 17:7402975-7402977 | - | TE | 0.14506 | 0.88663 | -0.74157 |
| TMEM44 | ENSG00000145014 | 3:194588529-194588639 | - | TE | 0.01176 | 0.1099 | -0.09814 |
| TMEM44 | ENSG00000145014 | 3:194587678-194587680 | - | TE | 0.166 | 0.013832 | 0.15217 |
| TMEM45B | ENSG00000151715 | 11:129858574-129859215 | + | TE | 0.007818 | 0.032234 | -0.02442 |
| TMEM50A | ENSG00000183726 | 1:25337917-25338319 | + | TS | 0.026291 | 0.18227 | -0.15598 |
| TMEM50A | ENSG00000183726 | 1:25338320-25338322 | + | TS | 0.025711 | 0.18196 | -0.15625 |
| TMEM59 | ENSG00000116209 | 1:54031663-54031675 | - | TE | 0.002142 | 0.008961 | -0.00682 |
| TMEM63A | ENSG00000196187 | 1:225846576-225846932 | - | TE | 0.012892 | 0.001591 | 0.011301 |
| TMEM86B | ENSG00000180089 | 19:55226748-55226750 | - | TE | 0.006062 | 0.1298 | -0.12374 |
| TMF1 | ENSG00000144747 | 3:69019827-69019841 | - | TE | 0.29614 | 0.071365 | 0.22477 |
| TMUB2 | ENSG00000168591 | 17:44190501-44190721 | + | TE | 0.021779 | 0.13882 | -0.11704 |
| TMX3 | ENSG00000166479 | 18:68715059-68715077 | - | TS | 0.057785 | 0.321 | -0.26322 |
| TMX3 | ENSG00000166479 | 18:68715058-68715058 | - | TS | 0.057524 | 0.31797 | -0.26045 |
| TNC | ENSG00000041982 | 9:115020946-115021156 | - | TE | 0.14883 | 0.77019 | -0.62135 |
| TNFSF13B | ENSG00000102524 | 13:108269629-108269717 | + | TS | 0.000776 | 0.031566 | -0.03079 |
| TNFSF15 | ENSG00000181634 | 9:114789320-114790906 | - | TE | 0.26146 | 0.028483 | 0.23297 |
| TNIK | ENSG00000154310 | 3:171063330-171063880 | - | TE | 0.00511 | 0.02612 | -0.02101 |
| TNK2 | ENSG00000061938 | 3:195868356-195868709 | - | TE | 0.00462 | 0.01995 | -0.01533 |
| TNS2 | ENSG00000111077 | 12:53063744-53064369 | + | TE | 0.17634 | 0.037711 | 0.13863 |
| TOE1 | ENSG00000132773 | 1:45343082-45343964 | + | TE | 0.18944 | 0.033518 | 0.15592 |
| TOMM6 | ENSG00000214736 | 6:41789897-41789898 | + | TE | 0.079905 | 0.00362 | 0.076286 |
| TOP3A | ENSG00000177302 | 17:18273967-18274723 | - | TE | 0.021067 | 0.15724 | -0.13617 |
| TP53BP1 | ENSG00000067369 | 15:43407214-43407215 | - | TE | 0.005958 | 0.079497 | -0.07354 |
| TP53I11 | ENSG00000175274 | 11:44932356-44933035 | - | TE | 0.000575 | 0.003021 | -0.00245 |
| TP53I3 | ENSG00000115129 | 2:24077435-24077633 | - | TE | 0.56313 | 0.07486 | 0.48827 |
| TP53I3 | ENSG00000115129 | 2:24077634-24077761 | - | TE | 0.025761 | 0.4269 | -0.40114 |
| TP53INP1 | ENSG00000164938 | 8:94949367-94949411 | - | TS | 0.02484 | 0.10752 | -0.08268 |
| TP63 | ENSG00000073282 | 3:189894222-189894231 | + | TE | 0.53789 | 0.094563 | 0.44333 |
| TPM2 | ENSG00000198467 | 9:35689914-35689925 | - | TS | 0.14719 | 0.60523 | -0.45804 |
| TPM2 | ENSG00000198467 | 9:35689926-35690056 | - | TS | 0.85776 | 0.099703 | 0.75805 |
| TPP1 | ENSG00000166340 | 11:6612976-6613032 | - | TE | 0.000285 | 0.002469 | -0.00218 |
| TPP1 | ENSG00000166340 | 11:6613033-6613303 | - | TE | 0.004129 | 0.000226 | 0.003903 |
| TPRN | ENSG00000176058 | 9:137191617-137191618 | - | TE | 0.29947 | 0.024013 | 0.27545 |
| TRA2A | ENSG00000164548 | 7:23531981-23531981 | - | TS | 0.025938 | 0.12319 | -0.09725 |
| TRABD | ENSG00000170638 | 22:50198712-50199595 | + | TE | 0.003727 | 0.077043 | -0.07332 |
| TRAF3IP3 | ENSG00000009790 | 1:209756066-209756178 | + | TS | 0.12208 | 0.87349 | -0.75141 |
| TRAF3IP3 | ENSG00000009790 | 1:209756049-209756065 | + | TS | 0.87694 | 0.12321 | 0.75372 |
| TRAF3IP3 | ENSG00000009790 | 1:209782316-209782320 | + | TE | 0.1486 | 0.033926 | 0.11467 |
| TRAK1 | ENSG00000182606 | 3:42212616-42212623 | + | TE | 0.15072 | 0.012559 | 0.13817 |
| TRAPPC9 | ENSG00000167632 | 8:139730343-139730344 | - | TE | 0.078374 | 0.012373 | 0.066001 |
| TRAPPC9 | ENSG00000167632 | 8:139730892-139731228 | - | TE | 0.033199 | 0.007186 | 0.026014 |
| TRIB3 | ENSG00000101255 | 20:396198-396748 | + | TE | 0.1006 | 0.010356 | 0.09024 |
| TRIM16L | ENSG00000108448 | 17:18735491-18735667 | + | TE | 0.81485 | 0.12254 | 0.69231 |
| TRIM23 | ENSG00000113595 | 5:65590176-65590302 | - | TE | 0.27105 | 0.025449 | 0.2456 |
| TRIM36 | ENSG00000152503 | 5:115180038-115180159 | - | TS | 0.10814 | 0.76118 | -0.65303 |
| TRIM6 | ENSG00000121236 | 11:5610777-5611279 | + | TE | 0.23037 | 0.016862 | 0.2135 |
| TRIM6 | ENSG00000121236 | 11:5611588-5611700 | + | TE | 0.008563 | 0.036502 | -0.02794 |
| TRIM62 | ENSG00000116525 | 1:33146702-33147727 | - | TE | 0.004515 | 0.032086 | -0.02757 |
| TRIM7 | ENSG00000146054 | 5:181193925-181193929 | - | TE | 0.12333 | 0.026356 | 0.096975 |
| TRIM73 | ENSG00000178809 | 7:75405606-75405613 | + | TE | 0.88695 | 0.13728 | 0.74967 |
| TRIM73 | ENSG00000178809 | 7:75405205-75405605 | + | TE | 0.12084 | 0.85877 | -0.73793 |
| TRIM8 | ENSG00000171206 | 10:102658192-102658262 | + | TE | 0.010478 | 0.08823 | -0.07775 |
| TRIP6 | ENSG00000087077 | 7:100867387-100867388 | + | TS | 0.78801 | 0.15303 | 0.63498 |
| TRIP6 | ENSG00000087077 | 7:100867389-100867497 | + | TS | 0.094018 | 0.74071 | -0.64669 |
| TRIQK | ENSG00000205133 | 8:92885714-92886232 | - | TE | 0.001688 | 0.017514 | -0.01583 |
| TRMT10A | ENSG00000145331 | 4:99546709-99546726 | - | TE | 0.026718 | 0.23547 | -0.20875 |
| TRPV1 | ENSG00000196689 | 17:3565444-3565458 | - | TE | 0.032542 | 0.26578 | -0.23323 |
| TSC1 | ENSG00000165699 | 9:132906140-132906532 | - | TS | 0.41318 | 0.065539 | 0.34765 |
| TSC2 | ENSG00000103197 | 16:2088693-2088700 | + | TE | 0.085735 | 0.018947 | 0.066788 |
| TSC2 | ENSG00000103197 | 16:2088446-2088577 | + | TE | 0.15196 | 0.018837 | 0.13312 |
| TSC22D2 | ENSG00000196428 | 3:150408335-150411008 | + | TS | 0.001283 | 0.18215 | -0.18087 |
| TSEN15 | ENSG00000198860 | 1:184095690-184095845 | + | TE | 0.19661 | 0.047077 | 0.14953 |
| TSEN15 | ENSG00000198860 | 1:184073952-184074127 | + | TE | 0.004985 | 0.022961 | -0.01798 |
| TSLP | ENSG00000145777 | 5:111076132-111076793 | + | TE | 0.75092 | 0.1421 | 0.60881 |
| TSLP | ENSG00000145777 | 5:111075946-111076131 | + | TE | 0.14318 | 0.79717 | -0.65398 |
| TSPAN10 | ENSG00000182612 | 17:81648295-81648748 | + | TE | 0.11355 | 0.78574 | -0.67219 |
| TSPAN10 | ENSG00000182612 | 17:81647901-81648294 | + | TE | 0.87187 | 0.13295 | 0.73892 |
| TSPAN14 | ENSG00000108219 | 10:80517905-80518216 | + | TE | 0.007537 | 0.030685 | -0.02315 |
| TSPAN14 | ENSG00000108219 | 10:80519629-80519635 | + | TE | 0.004233 | 0.000854 | 0.003379 |
| TSPAN32 | ENSG00000064201 | 11:2302013-2302030 | + | TS | 0.57756 | 0.11049 | 0.46707 |
| TSPAN32 | ENSG00000064201 | 11:2302040-2302146 | + | TS | 0.10229 | 0.71314 | -0.61085 |
| TSPO2 | ENSG00000112212 | 6:41043940-41044212 | + | TE | 0.1878 | 0.87214 | -0.68434 |
| TSPO2 | ENSG00000112212 | 6:41044257-41044337 | + | TE | 0.50564 | 0.11816 | 0.38748 |
| TSSC4 | ENSG00000184281 | 11:2402845-2402964 | + | TE | 0.028133 | 0.22741 | -0.19927 |
| TSTA3 | ENSG00000104522 | 8:143612621-143612965 | - | TE | 0.62881 | 0.14903 | 0.47978 |
| TSTD2 | ENSG00000136925 | 9:97600083-97602767 | - | TE | 0.12416 | 0.003698 | 0.12046 |
| TTC13 | ENSG00000143643 | 1:230978839-230978859 | - | TS | 0.068133 | 0.2781 | -0.20997 |
| TTC26 | ENSG00000105948 | 7:139189666-139189800 | + | TE | 0.013915 | 0.27505 | -0.26113 |
| TTC31 | ENSG00000115282 | 2:74493084-74494559 | + | TE | 0.25972 | 0.060744 | 0.19898 |
| TTC33 | ENSG00000113638 | 5:40716145-40716498 | - | TE | 0.002649 | 0.040403 | -0.03775 |
| TTC39B | ENSG00000155158 | 9:15307210-15307360 | - | TS | 0.082935 | 0.48059 | -0.39765 |
| TTC39C | ENSG00000168234 | 18:24133028-24133028 | + | TE | 0.013161 | 0.090014 | -0.07685 |
| TTC7A | ENSG00000068724 | 2:47075558-47076135 | + | TE | 0.015241 | 0.002073 | 0.013167 |
| TTC7B | ENSG00000165914 | 14:90540588-90540641 | - | TE | 0.029267 | 0.006933 | 0.022334 |
| TTI1 | ENSG00000101407 | 20:37983007-37983021 | - | TE | 0.009858 | 0.091684 | -0.08183 |
| TUBA1A | ENSG00000167552 | 12:49186163-49186309 | - | TE | 0.027965 | 0.12762 | -0.09966 |
| TUBA4A | ENSG00000127824 | 2:219251194-219251323 | - | TE | 0.066057 | 0.000487 | 0.06557 |
| TUBB6 | ENSG00000176014 | 18:12308241-12308243 | + | TS | 0.49114 | 0.10646 | 0.38468 |
| TUBGCP2 | ENSG00000130640 | 10:133279640-133279640 | - | TE | 0.013404 | 0.19935 | -0.18595 |
| TUBGCP6 | ENSG00000128159 | 22:50244565-50244971 | - | TS | 0.006911 | 0.057592 | -0.05068 |
| TUFT1 | ENSG00000143367 | 1:151583577-151583583 | + | TE | 0.16165 | 0.027638 | 0.13401 |
| TXNDC17 | ENSG00000129235 | 17:6642951-6643067 | + | TE | 0.016415 | 0.1171 | -0.10069 |
| TXNRD1 | ENSG00000198431 | 12:104348584-104348644 | + | TE | 0.001295 | 0.008554 | -0.00726 |
| TYMS | ENSG00000176890 | 18:672860-672997 | + | TE | 0.02633 | 0.24878 | -0.22245 |
| U2AF2 | ENSG00000063244 | 19:55655065-55655081 | + | TS | 0.22812 | 0.007573 | 0.22055 |
| U2SURP | ENSG00000163714 | 3:143056312-143056500 | + | TE | 0.001983 | 0.04664 | -0.04466 |
| UACA | ENSG00000137831 | 15:70656935-70656976 | - | TE | 0.23377 | 0.05668 | 0.17709 |
| UBAC2 | ENSG00000134882 | 13:99385228-99385394 | + | TE | 0.003798 | 0.17379 | -0.16999 |
| UBALD1 | ENSG00000153443 | 16:4609368-4609908 | - | TE | 0.0185 | 0.002587 | 0.015913 |
| UBC | ENSG00000150991 | 12:124911604-124911647 | - | TE | 7.26E-05 | 0.000793 | -0.00072 |
| UBC | ENSG00000150991 | 12:124913370-124913403 | - | TE | 0.003057 | 0.028042 | -0.02499 |
| UBE2D1 | ENSG00000072401 | 10:58368720-58369043 | + | TE | 0.000363 | 0.051219 | -0.05086 |
| UBE2D2 | ENSG00000131508 | 5:139626856-139627115 | + | TE | 0.001347 | 0.013745 | -0.0124 |
| UBE2F | ENSG00000184182 | 2:238041887-238041933 | + | TE | 0.005984 | 0.036048 | -0.03006 |
| UBE2H | ENSG00000186591 | 7:129952632-129952949 | - | TS | 0.015854 | 0.29704 | -0.28119 |
| UBE2H | ENSG00000186591 | 7:129834978-129835061 | - | TE | 0.000383 | 0.02226 | -0.02188 |
| UBE2I | ENSG00000103275 | 16:1317126-1317190 | + | TE | 0.23558 | 0.042032 | 0.19355 |
| UBE2T | ENSG00000077152 | 1:202331738-202331960 | - | TE | 0.87727 | 0.14849 | 0.72878 |
| UBE2T | ENSG00000077152 | 1:202331657-202331737 | - | TE | 0.13016 | 0.86224 | -0.73208 |
| UBE2W | ENSG00000104343 | 8:73792363-73794115 | - | TE | 0.000252 | 0.002542 | -0.00229 |
| UBE3A | ENSG00000114062 | 15:25438542-25439024 | - | TS | 0.054616 | 0.013565 | 0.041051 |
| UBE4B | ENSG00000130939 | 1:10179895-10180016 | + | TE | 0.0143 | 0.57977 | -0.56547 |
| UBFD1 | ENSG00000103353 | 16:23570859-23574387 | + | TE | 0.24271 | 0.003505 | 0.23921 |
| UBR4 | ENSG00000127481 | 1:19074506-19074509 | - | TE | 0.051918 | 0.006874 | 0.045044 |
| UBR4 | ENSG00000127481 | 1:19074510-19074513 | - | TE | 0.008936 | 0.037778 | -0.02884 |
| UBR5 | ENSG00000104517 | 8:102253736-102253945 | - | TE | 0.031194 | 0.00251 | 0.028684 |
| UBTF | ENSG00000108312 | 17:44207240-44207367 | - | TE | 0.11766 | 0.001999 | 0.11566 |
| UBXN6 | ENSG00000167671 | 19:4445007-4445009 | - | TE | 0.12876 | 0.001871 | 0.12689 |
| UBXN6 | ENSG00000167671 | 19:4445006-4445006 | - | TE | 0.007392 | 0.001784 | 0.005608 |
| UFD1 | ENSG00000070010 | 22:19449910-19450620 | - | TE | 0.070552 | 0.63166 | -0.56111 |
| UGGT2 | ENSG00000102595 | 13:96053401-96053482 | - | TS | 0.099354 | 0.73638 | -0.63702 |
| UMAD1 | ENSG00000219545 | 7:7877281-7877661 | + | TE | 0.007836 | 0.00173 | 0.006105 |
| UPF3B | ENSG00000125351 | X:119834022-119834025 | - | TE | 0.036146 | 0.27924 | -0.2431 |
| UPP1 | ENSG00000183696 | 7:48108218-48108415 | + | TE | 0.006465 | 0.037276 | -0.03081 |
| UQCC1 | ENSG00000101019 | 20:35346747-35347163 | - | TE | 0.066779 | 0.004945 | 0.061834 |
| URGCP | ENSG00000106608 | 7:43878844-43879047 | - | TE | 0.008393 | 0.07677 | -0.06838 |
| USP13 | ENSG00000058056 | 3:179784048-179784406 | + | TE | 0.011207 | 0.047693 | -0.03649 |
| USP28 | ENSG00000048028 | 11:113798805-113798978 | - | TE | 0.027727 | 0.005904 | 0.021823 |
| USP30 | ENSG00000135093 | 12:109085923-109086377 | + | TE | 0.013817 | 0.061393 | -0.04758 |
| USP33 | ENSG00000077254 | 1:77695987-77695990 | - | TE | 0.21944 | 0.012794 | 0.20665 |
| USP35 | ENSG00000118369 | 11:78214139-78214419 | + | TE | 0.013614 | 0.097839 | -0.08422 |
| USP39 | ENSG00000168883 | 2:85649281-85649282 | + | TE | 0.08133 | 0.008836 | 0.072494 |
| UTP23 | ENSG00000147679 | 8:116766503-116766504 | + | TS | 0.1469 | 0.72413 | -0.57722 |
| UTP6 | ENSG00000108651 | 17:31863174-31863348 | - | TE | 0.17061 | 0.029269 | 0.14134 |
| UTP6 | ENSG00000108651 | 17:31863349-31863516 | - | TE | 0.04268 | 0.24953 | -0.20686 |
| VAV3 | ENSG00000134215 | 1:107572848-107572949 | - | TE | 0.013539 | 0.001352 | 0.012187 |
| VCAN | ENSG00000038427 | 5:83471764-83471776 | + | TS | 0.013109 | 0.001 | 0.012109 |
| VDAC2 | ENSG00000165637 | 10:75231276-75231277 | + | TE | 0.067596 | 0.006641 | 0.060955 |
| VIPAS39 | ENSG00000151445 | 14:77426675-77426681 | - | TE | 0.41418 | 0.088787 | 0.3254 |
| VIPR1 | ENSG00000114812 | 3:42537568-42537573 | + | TE | 0.14955 | 0.007108 | 0.14244 |
| VOPP1 | ENSG00000154978 | 7:55471968-55472202 | - | TE | 0.000594 | 0.00427 | -0.00368 |
| VPS11 | ENSG00000160695 | 11:119081966-119081967 | + | TE | 0.027536 | 0.14168 | -0.11415 |
| VPS53 | ENSG00000141252 | 17:519119-519298 | - | TE | 0.010485 | 0.062204 | -0.05172 |
| VSIG4 | ENSG00000155659 | X:66040108-66040125 | - | TS | 0.13899 | 0.64102 | -0.50203 |
| VTI1A | ENSG00000151532 | 10:112447234-112447257 | + | TS | 0.91449 | 0.2011 | 0.71339 |
| VTI1A | ENSG00000151532 | 10:112447258-112447263 | + | TS | 0.056982 | 0.70568 | -0.6487 |
| VWA3B | ENSG00000168658 | 2:98087116-98087176 | + | TS | 0.74159 | 0.12694 | 0.61465 |
| WARS | ENSG00000140105 | 14:100376260-100376299 | - | TS | 0.14621 | 0.87509 | -0.72888 |
| WDPCP | ENSG00000143951 | 2:63121831-63122056 | - | TE | 0.6797 | 0.09545 | 0.58425 |
| WDR11 | ENSG00000120008 | 10:120909521-120909523 | + | TE | 0.11079 | 0.02109 | 0.089699 |
| WDR19 | ENSG00000157796 | 4:39285781-39285809 | + | TE | 0.52256 | 0.080281 | 0.44228 |
| WDR27 | ENSG00000184465 | 6:169457216-169457354 | - | TE | 0.080552 | 0.40271 | -0.32216 |
| WDR37 | ENSG00000047056 | 10:1132171-1132297 | + | TE | 0.001085 | 0.007386 | -0.0063 |
| WDR45 | ENSG00000196998 | X:49074433-49074639 | - | TE | 0.013608 | 0.085068 | -0.07146 |
| WDR48 | ENSG00000114742 | 3:39096665-39096671 | + | TE | 0.33859 | 0.064096 | 0.27449 |
| WDR86 | ENSG00000187260 | 7:151381128-151381746 | - | TE | 0.096397 | 0.88799 | -0.7916 |
| WDR86 | ENSG00000187260 | 7:151381121-151381127 | - | TE | 0.89908 | 0.11102 | 0.78806 |
| WDR89 | ENSG00000140006 | 14:63599263-63599973 | - | TE | 0.011926 | 0.06278 | -0.05085 |
| WIPF1 | ENSG00000115935 | 2:174597766-174597804 | - | TS | 0.004949 | 0.025044 | -0.0201 |
| WIPI2 | ENSG00000157954 | 7:5231065-5231327 | + | TE | 0.008123 | 0.001059 | 0.007064 |
| WISP3 | ENSG00000112761 | 6:112069339-112069658 | + | TE | 0.29396 | 0.068678 | 0.22528 |
| WLS | ENSG00000116729 | 1:68125492-68125973 | - | TE | 0.002209 | 0.017497 | -0.01529 |
| WTAP | ENSG00000146457 | 6:159756318-159756319 | + | TE | 0.30773 | 0.044119 | 0.26361 |
| WWP1 | ENSG00000123124 | 8:86466794-86467476 | + | TE | 0.003812 | 0.1321 | -0.12829 |
| WWP2 | ENSG00000198373 | 16:69940177-69940178 | + | TE | 0.001189 | 0.006876 | -0.00569 |
| XAF1 | ENSG00000132530 | 17:6756062-6756078 | + | TS | 0.70883 | 0.050514 | 0.65832 |
| XAF1 | ENSG00000132530 | 17:6755838-6755839 | + | TS | 0.11041 | 0.90599 | -0.79559 |
| XAF1 | ENSG00000132530 | 17:6773664-6774108 | + | TE | 0.000227 | 0.001064 | -0.00084 |
| XAF1 | ENSG00000132530 | 17:6773113-6773169 | + | TE | 0.00023 | 0.001084 | -0.00085 |
| XAF1 | ENSG00000132530 | 17:6756079-6756110 | + | TS | 0.30777 | 0.056008 | 0.25176 |
| XBP1 | ENSG00000100219 | 22:28794589-28795732 | - | TE | 0.000318 | 0.025288 | -0.02497 |
| XPO5 | ENSG00000124571 | 6:43522337-43524005 | - | TE | 0.01022 | 0.12932 | -0.1191 |
| YAF2 | ENSG00000015153 | 12:42160266-42160607 | - | TE | 0.001778 | 0.043604 | -0.04183 |
| YBEY | ENSG00000182362 | 21:46286355-46286383 | + | TS | 0.083776 | 0.38366 | -0.29988 |
| YME1L1 | ENSG00000136758 | 10:27154421-27155266 | - | TS | 0.43297 | 0.020817 | 0.41216 |
| YPEL5 | ENSG00000119801 | 2:30159139-30159551 | + | TE | 0.000328 | 0.005596 | -0.00527 |
| YWHAB | ENSG00000166913 | 20:44906382-44906504 | + | TE | 0.061728 | 0.000202 | 0.061526 |
| YWHAE | ENSG00000108953 | 17:1361681-1361901 | - | TE | 0.041727 | 0.19498 | -0.15326 |
| ZBED8 | ENSG00000221886 | 5:160400037-160400097 | - | TS | 0.87705 | 0.14767 | 0.72938 |
| ZBED8 | ENSG00000221886 | 5:160399639-160400036 | - | TS | 0.12072 | 0.85683 | -0.73611 |
| ZBTB17 | ENSG00000116809 | 1:15941871-15942252 | - | TE | 0.1719 | 0.7368 | -0.5649 |
| ZBTB38 | ENSG00000177311 | 3:141444702-141445976 | + | TE | 0.000428 | 0.004267 | -0.00384 |
| ZBTB46 | ENSG00000130584 | 20:63744525-63744681 | - | TE | 0.023206 | 0.21106 | -0.18785 |
| ZC3H10 | ENSG00000135482 | 12:56120699-56120828 | + | TE | 0.017204 | 0.001772 | 0.015432 |
| ZC3H15 | ENSG00000065548 | 2:186508543-186508690 | + | TE | 0.15805 | 0.012516 | 0.14553 |
| ZC3H18 | ENSG00000158545 | 16:88631931-88631934 | + | TE | 0.029962 | 0.14717 | -0.11721 |
| ZCCHC14 | ENSG00000140948 | 16:87406257-87407014 | - | TE | 0.010963 | 0.070187 | -0.05922 |
| ZCCHC17 | ENSG00000121766 | 1:31364032-31364936 | + | TE | 0.018528 | 0.46881 | -0.45028 |
| ZDHHC13 | ENSG00000177054 | 11:19176416-19176422 | + | TE | 0.16163 | 0.031268 | 0.13036 |
| ZDHHC16 | ENSG00000171307 | 10:97457364-97457366 | + | TE | 0.28937 | 0.014127 | 0.27524 |
| ZDHHC23 | ENSG00000184307 | 3:113962979-113962982 | + | TE | 0.15704 | 0.034209 | 0.12284 |
| ZDHHC24 | ENSG00000174165 | 11:66545723-66546235 | - | TS | 0.21797 | 0.041732 | 0.17624 |
| ZDHHC9 | ENSG00000188706 | X:129805329-129806486 | - | TE | 0.089841 | 0.38324 | -0.2934 |
| ZFC3H1 | ENSG00000133858 | 12:71663825-71663858 | - | TS | 0.15292 | 0.011084 | 0.14184 |
| ZFR | ENSG00000056097 | 5:32354852-32355939 | - | TE | 0.00243 | 0.011369 | -0.00894 |
| ZFX | ENSG00000005889 | X:24179183-24179219 | + | TE | 0.15003 | 0.024827 | 0.12521 |
| ZFYVE28 | ENSG00000159733 | 4:2269582-2269597 | - | TE | 0.11584 | 0.028849 | 0.086995 |
| ZMAT3 | ENSG00000172667 | 3:179071595-179072215 | - | TS | 0.12712 | 0.68978 | -0.56266 |
| ZMIZ2 | ENSG00000122515 | 7:44768689-44768694 | + | TE | 0.002653 | 0.086328 | -0.08368 |
| ZMYM6 | ENSG00000163867 | 1:35017250-35018512 | - | TE | 0.031271 | 0.18729 | -0.15602 |
| ZMYND10 | ENSG00000004838 | 3:50345733-50346852 | - | TS | 0.13837 | 0.034245 | 0.10413 |
| ZNF10 | ENSG00000256223 | 12:133156969-133156991 | + | TE | 0.017832 | 0.13058 | -0.11274 |
| ZNF106 | ENSG00000103994 | 15:42457217-42457513 | - | TS | 0.066643 | 0.36498 | -0.29833 |
| ZNF107 | ENSG00000196247 | 7:64706580-64711023 | + | TE | 0.007666 | 0.11627 | -0.1086 |
| ZNF134 | ENSG00000213762 | 19:57620160-57620447 | + | TE | 0.027763 | 0.24854 | -0.22078 |
| ZNF142 | ENSG00000115568 | 2:218638145-218638808 | - | TE | 0.013943 | 0.10045 | -0.08651 |
| ZNF175 | ENSG00000105497 | 19:51589548-51589738 | + | TE | 0.014278 | 0.068999 | -0.05472 |
| ZNF181 | ENSG00000197841 | 19:34740611-34740818 | + | TE | 0.17272 | 0.015894 | 0.15682 |
| ZNF207 | ENSG00000010244 | 17:32369599-32370343 | + | TE | 9.39E-05 | 0.021098 | -0.021 |
| ZNF217 | ENSG00000171940 | 20:53568820-53569264 | - | TE | 0.000367 | 0.004712 | -0.00434 |
| ZNF224 | ENSG00000267680 | 19:44108327-44109886 | + | TE | 0.35257 | 0.066766 | 0.2858 |
| ZNF227 | ENSG00000131115 | 19:44234702-44234835 | + | TE | 0.010109 | 0.20534 | -0.19523 |
| ZNF23 | ENSG00000167377 | 16:71447597-71447635 | - | TE | 0.075934 | 0.013013 | 0.062921 |
| ZNF25 | ENSG00000175395 | 10:37950567-37953195 | - | TE | 0.042952 | 0.004651 | 0.038301 |
| ZNF253 | ENSG00000256771 | 19:19892590-19893130 | + | TE | 0.032148 | 0.14462 | -0.11248 |
| ZNF268 | ENSG00000090612 | 12:133202144-133202348 | + | TE | 0.013455 | 0.09562 | -0.08217 |
| ZNF268 | ENSG00000090612 | 12:133205024-133205121 | + | TE | 0.038105 | 0.005169 | 0.032937 |
| ZNF280D | ENSG00000137871 | 15:56631298-56632122 | - | TE | 0.007945 | 0.051722 | -0.04378 |
| ZNF286A | ENSG00000187607 | 17:15716059-15716490 | + | TE | 0.082188 | 0.010044 | 0.072143 |
| ZNF3 | ENSG00000166526 | 7:100070177-100071929 | - | TE | 0.12845 | 0.004882 | 0.12356 |
| ZNF320 | ENSG00000182986 | 19:52863866-52863921 | - | TE | 0.12211 | 0.56415 | -0.44204 |
| ZNF331 | ENSG00000130844 | 19:53578082-53578386 | + | TE | 0.055309 | 0.012513 | 0.042796 |
| ZNF343 | ENSG00000088876 | 20:2484527-2484656 | - | TE | 0.019053 | 0.25058 | -0.23153 |
| ZNF345 | ENSG00000251247 | 19:36878602-36879569 | + | TE | 0.020561 | 0.12434 | -0.10377 |
| ZNF385C | ENSG00000187595 | 17:42025625-42027133 | - | TE | 0.68736 | 0.15578 | 0.53158 |
| ZNF408 | ENSG00000175213 | 11:46704353-46704522 | + | TE | 0.005987 | 0.068844 | -0.06286 |
| ZNF426 | ENSG00000130818 | 19:9528007-9529296 | - | TE | 0.02827 | 0.004585 | 0.023686 |
| ZNF440 | ENSG00000171295 | 19:11831368-11831678 | + | TE | 0.058747 | 0.00954 | 0.049207 |
| ZNF446 | ENSG00000083838 | 19:58480704-58481224 | + | TE | 0.50689 | 0.096102 | 0.41079 |
| ZNF493 | ENSG00000196268 | 19:21422913-21423040 | + | TE | 0.13003 | 0.002022 | 0.12801 |
| ZNF497 | ENSG00000174586 | 19:58355561-58357649 | - | TE | 0.088031 | 0.01683 | 0.071201 |
| ZNF506 | ENSG00000081665 | 19:19786562-19786854 | - | TE | 0.003902 | 0.068141 | -0.06424 |
| ZNF507 | ENSG00000168813 | 19:32383249-32383254 | + | TE | 0.075598 | 0.004459 | 0.071139 |
| ZNF567 | ENSG00000189042 | 19:36721322-36721326 | + | TE | 0.082723 | 0.39833 | -0.31561 |
| ZNF568 | ENSG00000198453 | 19:36996317-36996518 | + | TE | 0.37644 | 0.088684 | 0.28775 |
| ZNF570 | ENSG00000171827 | 19:37485321-37485358 | + | TE | 0.014419 | 0.0789 | -0.06448 |
| ZNF574 | ENSG00000105732 | 19:42078587-42081405 | + | TE | 0.039243 | 0.19693 | -0.15769 |
| ZNF577 | ENSG00000161551 | 19:51872550-51873529 | - | TE | 0.06592 | 0.26388 | -0.19796 |
| ZNF584 | ENSG00000171574 | 19:58408688-58408688 | + | TS | 0.11899 | 0.51198 | -0.39298 |
| ZNF606 | ENSG00000166704 | 19:57978265-57980279 | - | TE | 0.057925 | 0.29012 | -0.2322 |
| ZNF608 | ENSG00000168916 | 5:124648237-124648262 | - | TE | 0.006954 | 0.049442 | -0.04249 |
| ZNF611 | ENSG00000213020 | 19:52706710-52706864 | - | TE | 0.002171 | 0.040298 | -0.03813 |
| ZNF614 | ENSG00000142556 | 19:52017001-52017116 | - | TE | 0.006352 | 0.11671 | -0.11036 |
| ZNF628 | ENSG00000197483 | 19:55481511-55484373 | + | TE | 0.09722 | 0.004007 | 0.093213 |
| ZNF644 | ENSG00000122482 | 1:90916791-90916990 | - | TE | 0.003967 | 0.071512 | -0.06755 |
| ZNF689 | ENSG00000156853 | 16:30604540-30604617 | - | TE | 0.004777 | 0.07092 | -0.06614 |
| ZNF708 | ENSG00000182141 | 19:21329411-21329425 | - | TS | 0.085834 | 0.35154 | -0.26571 |
| ZNF711 | ENSG00000147180 | X:85270651-85271921 | + | TE | 0.050743 | 0.3873 | -0.33656 |
| ZNF74 | ENSG00000185252 | 22:20407394-20407407 | + | TE | 0.012507 | 0.17504 | -0.16254 |
| ZNF74 | ENSG00000185252 | 22:20408456-20408461 | + | TE | 0.21413 | 0.031957 | 0.18217 |
| ZNF75A | ENSG00000162086 | 16:3309450-3309472 | + | TE | 0.2913 | 0.042864 | 0.24843 |
| ZNF764 | ENSG00000169951 | 16:30555638-30556107 | - | TE | 0.002756 | 0.013758 | -0.011 |
| ZNF780A | ENSG00000197782 | 19:40072993-40073179 | - | TE | 0.021083 | 0.26751 | -0.24643 |
| ZNF816 | ENSG00000180257 | 19:52949379-52949441 | - | TE | 0.26188 | 0.03333 | 0.22855 |
| ZNF823 | ENSG00000197933 | 19:11721993-11723112 | - | TE | 0.028551 | 0.13554 | -0.10699 |
| ZNF823 | ENSG00000197933 | 19:11723113-11723342 | - | TE | 0.028465 | 0.19774 | -0.16928 |
| ZNRD1 | ENSG00000066379 | 6:30061263-30061329 | + | TS | 0.20081 | 0.045181 | 0.15563 |
| ZNRD1 | ENSG00000066379 | 6:30061259-30061262 | + | TS | 0.20176 | 0.045132 | 0.15663 |
| ZNRF1 | ENSG00000186187 | 16:74999345-75000095 | + | TS | 0.013803 | 0.13521 | -0.12141 |
| ZSCAN18 | ENSG00000121413 | 19:58083843-58083858 | - | TE | 0.020539 | 0.14117 | -0.12063 |
| ZSWIM8 | ENSG00000214655 | 10:73785624-73785654 | + | TS | 0.085498 | 0.48674 | -0.40125 |
| ZWINT | ENSG00000122952 | 10:56357438-56357444 | - | TE | 0.16825 | 0.040728 | 0.12753 |

Table S6 GO Term Analysis of all significant gene expression differences between COVID-19 patients who died versus lived.

| GO biological process complete | Human (20851) | Significant Genes (281) | Expected count | over/under | Fold Enrichment | P-value | FDR |
| --- | --- | --- | --- | --- | --- | --- | --- |
| protein homotrimerization (GO:0070207) | 15 | 4 | 0.2 | + | 19.79 | 1.01E-04 | 1.92E-02 |
| negative regulation of interleukin-10 production (GO:0032693) | 18 | 4 | 0.24 | + | 16.49 | 1.85E-04 | 2.81E-02 |
| protein trimerization (GO:0070206) | 18 | 4 | 0.24 | + | 16.49 | 1.85E-04 | 2.78E-02 |
| retinal ganglion cell axon guidance (GO:0031290) | 18 | 4 | 0.24 | + | 16.49 | 1.85E-04 | 2.75E-02 |
| axonal fasciculation (GO:0007413) | 23 | 4 | 0.31 | + | 12.9 | 4.22E-04 | 4.90E-02 |
| neuron projection fasciculation (GO:0106030) | 23 | 4 | 0.31 | + | 12.9 | 4.22E-04 | 4.86E-02 |
| positive regulation of signaling receptor activity (GO:2000273) | 38 | 6 | 0.51 | + | 11.72 | 2.42E-05 | 7.40E-03 |
| regulation of NMDA receptor activity (GO:2000310) | 40 | 6 | 0.54 | + | 11.13 | 3.14E-05 | 8.76E-03 |
| forebrain neuron differentiation (GO:0021879) | 51 | 7 | 0.69 | + | 10.18 | 1.15E-05 | 4.35E-03 |
| regulation of activated T cell proliferation (GO:0046006) | 39 | 5 | 0.53 | + | 9.51 | 2.85E-04 | 3.87E-02 |
| forebrain generation of neurons (GO:0021872) | 63 | 8 | 0.85 | + | 9.42 | 4.56E-06 | 2.34E-03 |
| regulation of glutamate receptor signaling pathway (GO:1900449) | 68 | 7 | 0.92 | + | 7.64 | 6.26E-05 | 1.40E-02 |
| hair follicle development (GO:0001942) | 74 | 7 | 1 | + | 7.02 | 1.02E-04 | 1.92E-02 |
| regulation of neurotransmitter receptor activity (GO:0099601) | 85 | 8 | 1.15 | + | 6.98 | 3.39E-05 | 9.29E-03 |
| skin epidermis development (GO:0098773) | 77 | 7 | 1.04 | + | 6.75 | 1.29E-04 | 2.23E-02 |
| hair cycle process (GO:0022405) | 77 | 7 | 1.04 | + | 6.75 | 1.29E-04 | 2.20E-02 |
| molting cycle process (GO:0022404) | 77 | 7 | 1.04 | + | 6.75 | 1.29E-04 | 2.18E-02 |
| synapse assembly (GO:0007416) | 103 | 9 | 1.39 | + | 6.48 | 1.90E-05 | 6.43E-03 |
| negative regulation of leukocyte proliferation (GO:0070664) | 89 | 7 | 1.2 | + | 5.84 | 2.97E-04 | 3.96E-02 |
| hair cycle (GO:0042633) | 91 | 7 | 1.23 | + | 5.71 | 3.37E-04 | 4.28E-02 |
| molting cycle (GO:0042303) | 91 | 7 | 1.23 | + | 5.71 | 3.37E-04 | 4.25E-02 |
| regulation of signaling receptor activity (GO:0010469) | 182 | 12 | 2.45 | + | 4.89 | 1.19E-05 | 4.28E-03 |
| regulation of cation channel activity (GO:2001257) | 186 | 11 | 2.51 | + | 4.39 | 6.97E-05 | 1.50E-02 |
| cellular response to lipopolysaccharide (GO:0071222) | 187 | 11 | 2.52 | + | 4.36 | 7.29E-05 | 1.53E-02 |
| positive regulation of synaptic transmission (GO:0050806) | 154 | 9 | 2.08 | + | 4.34 | 3.44E-04 | 4.24E-02 |
| cellular response to metal ion (GO:0071248) | 196 | 11 | 2.64 | + | 4.16 | 1.08E-04 | 1.98E-02 |
| cellular response to molecule of bacterial origin (GO:0071219) | 197 | 11 | 2.65 | + | 4.14 | 1.13E-04 | 2.04E-02 |
| synapse organization (GO:0050808) | 294 | 16 | 3.96 | + | 4.04 | 4.44E-06 | 2.35E-03 |
| response to molecule of bacterial origin (GO:0002237) | 336 | 18 | 4.53 | + | 3.98 | 1.38E-06 | 1.04E-03 |
| response to lipopolysaccharide (GO:0032496) | 318 | 17 | 4.29 | + | 3.97 | 2.80E-06 | 1.78E-03 |
| central nervous system neuron differentiation (GO:0021953) | 192 | 10 | 2.59 | + | 3.86 | 3.91E-04 | 4.63E-02 |
| cellular response to biotic stimulus (GO:0071216) | 221 | 11 | 2.98 | + | 3.69 | 2.93E-04 | 3.94E-02 |
| regulation of leukocyte proliferation (GO:0070663) | 242 | 12 | 3.26 | + | 3.68 | 1.62E-04 | 2.50E-02 |
| cell junction assembly (GO:0034329) | 284 | 14 | 3.83 | + | 3.66 | 4.96E-05 | 1.23E-02 |
| cellular response to inorganic substance (GO:0071241) | 224 | 11 | 3.02 | + | 3.64 | 3.26E-04 | 4.25E-02 |
| cellular response to tumor necrosis factor (GO:0071356) | 246 | 12 | 3.32 | + | 3.62 | 1.87E-04 | 2.75E-02 |
| negative regulation of cytokine production (GO:0001818) | 275 | 13 | 3.71 | + | 3.51 | 1.37E-04 | 2.27E-02 |
| regulation of ion transmembrane transporter activity (GO:0032412) | 269 | 12 | 3.63 | + | 3.31 | 4.07E-04 | 4.75E-02 |
| cell junction organization (GO:0034330) | 510 | 22 | 6.87 | + | 3.2 | 2.90E-06 | 1.71E-03 |
| synaptic signaling (GO:0099536) | 465 | 20 | 6.27 | + | 3.19 | 8.63E-06 | 3.61E-03 |
| cellular response to lipid (GO:0071396) | 499 | 21 | 6.72 | + | 3.12 | 7.05E-06 | 3.11E-03 |
| chemotaxis (GO:0006935) | 549 | 23 | 7.4 | + | 3.11 | 2.74E-06 | 1.82E-03 |
| forebrain development (GO:0030900) | 406 | 17 | 5.47 | + | 3.11 | 5.66E-05 | 1.32E-02 |
| taxis (GO:0042330) | 551 | 23 | 7.43 | + | 3.1 | 2.90E-06 | 1.65E-03 |
| modulation of chemical synaptic transmission (GO:0050804) | 460 | 19 | 6.2 | + | 3.06 | 2.48E-05 | 7.45E-03 |
| regulation of trans-synaptic signaling (GO:0099177) | 461 | 19 | 6.21 | + | 3.06 | 2.56E-05 | 7.52E-03 |
| response to metal ion (GO:0010038) | 374 | 15 | 5.04 | + | 2.98 | 2.41E-04 | 3.42E-02 |
| neutrophil degranulation (GO:0043312) | 482 | 19 | 6.5 | + | 2.93 | 4.56E-05 | 1.17E-02 |
| regulation of ion transport (GO:0043269) | 711 | 28 | 9.58 | + | 2.92 | 7.08E-07 | 6.25E-04 |
| neutrophil activation involved in immune response (GO:0002283) | 486 | 19 | 6.55 | + | 2.9 | 5.07E-05 | 1.22E-02 |
| trans-synaptic signaling (GO:0099537) | 436 | 17 | 5.88 | + | 2.89 | 1.30E-04 | 2.18E-02 |
| neutrophil mediated immunity (GO:0002446) | 493 | 19 | 6.64 | + | 2.86 | 6.09E-05 | 1.40E-02 |
| neutrophil activation (GO:0042119) | 495 | 19 | 6.67 | + | 2.85 | 6.42E-05 | 1.42E-02 |
| chemical synaptic transmission (GO:0007268) | 417 | 16 | 5.62 | + | 2.85 | 2.43E-04 | 3.42E-02 |
| anterograde trans-synaptic signaling (GO:0098916) | 417 | 16 | 5.62 | + | 2.85 | 2.43E-04 | 3.39E-02 |
| response to bacterium (GO:0009617) | 730 | 28 | 9.84 | + | 2.85 | 1.17E-06 | 9.26E-04 |
| granulocyte activation (GO:0036230) | 500 | 19 | 6.74 | + | 2.82 | 7.29E-05 | 1.54E-02 |
| leukocyte degranulation (GO:0043299) | 504 | 19 | 6.79 | + | 2.8 | 8.06E-05 | 1.62E-02 |
| regulation of cell morphogenesis (GO:0022604) | 508 | 19 | 6.85 | + | 2.78 | 8.91E-05 | 1.75E-02 |
| positive regulation of cell adhesion (GO:0045785) | 432 | 16 | 5.82 | + | 2.75 | 3.55E-04 | 4.34E-02 |
| myeloid leukocyte mediated immunity (GO:0002444) | 514 | 19 | 6.93 | + | 2.74 | 1.03E-04 | 1.91E-02 |
| myeloid cell activation involved in immune response (GO:0002275) | 520 | 19 | 7.01 | + | 2.71 | 1.19E-04 | 2.09E-02 |
| detection of stimulus involved in sensory perception (GO:0050906) | 549 | 20 | 7.4 | + | 2.7 | 8.23E-05 | 1.63E-02 |
| regulation of cell adhesion (GO:0030155) | 722 | 26 | 9.73 | + | 2.67 | 8.48E-06 | 3.64E-03 |
| myeloid leukocyte activation (GO:0002274) | 584 | 21 | 7.87 | + | 2.67 | 6.53E-05 | 1.42E-02 |
| regulation of transmembrane transport (GO:0034762) | 579 | 20 | 7.8 | + | 2.56 | 1.64E-04 | 2.50E-02 |
| regulated exocytosis (GO:0045055) | 696 | 24 | 9.38 | + | 2.56 | 3.73E-05 | 1.00E-02 |
| positive regulation of cell migration (GO:0030335) | 526 | 18 | 7.09 | + | 2.54 | 3.86E-04 | 4.61E-02 |
| metal ion transport (GO:0030001) | 647 | 22 | 8.72 | + | 2.52 | 9.62E-05 | 1.84E-02 |
| positive regulation of locomotion (GO:0040017) | 566 | 19 | 7.63 | + | 2.49 | 3.36E-04 | 4.31E-02 |
| sensory perception (GO:0007600) | 984 | 33 | 13.26 | + | 2.49 | 3.04E-06 | 1.66E-03 |
| exocytosis (GO:0006887) | 793 | 26 | 10.69 | + | 2.43 | 5.39E-05 | 1.28E-02 |
| nervous system process (GO:0050877) | 1411 | 46 | 19.02 | + | 2.42 | 3.89E-08 | 7.72E-05 |
| secretion by cell (GO:0032940) | 986 | 32 | 13.29 | + | 2.41 | 6.88E-06 | 3.13E-03 |
| response to lipid (GO:0033993) | 842 | 27 | 11.35 | + | 2.38 | 4.78E-05 | 1.21E-02 |
| export from cell (GO:0140352) | 1035 | 33 | 13.95 | + | 2.37 | 6.53E-06 | 3.15E-03 |
| negative regulation of cell population proliferation (GO:0008285) | 700 | 22 | 9.43 | + | 2.33 | 3.35E-04 | 4.33E-02 |
| detection of stimulus (GO:0051606) | 709 | 22 | 9.55 | + | 2.3 | 3.80E-04 | 4.58E-02 |
| system process (GO:0003008) | 2072 | 64 | 27.92 | + | 2.29 | 3.95E-10 | 2.09E-06 |
| secretion (GO:0046903) | 1104 | 34 | 14.88 | + | 2.29 | 1.16E-05 | 4.30E-03 |
| regulation of cell migration (GO:0030334) | 889 | 27 | 11.98 | + | 2.25 | 1.49E-04 | 2.37E-02 |
| cell-cell signaling (GO:0007267) | 1141 | 34 | 15.38 | + | 2.21 | 1.90E-05 | 6.30E-03 |
| cell adhesion (GO:0007155) | 949 | 28 | 12.79 | + | 2.19 | 1.41E-04 | 2.30E-02 |
| biological adhesion (GO:0022610) | 955 | 28 | 12.87 | + | 2.18 | 1.52E-04 | 2.36E-02 |
| response to cytokine (GO:0034097) | 1126 | 33 | 15.17 | + | 2.17 | 5.01E-05 | 1.23E-02 |
| regulation of locomotion (GO:0040012) | 992 | 29 | 13.37 | + | 2.17 | 1.16E-04 | 2.06E-02 |
| cellular response to cytokine stimulus (GO:0071345) | 1038 | 30 | 13.99 | + | 2.14 | 1.51E-04 | 2.37E-02 |
| regulation of anatomical structure morphogenesis (GO:0022603) | 1110 | 32 | 14.96 | + | 2.14 | 8.00E-05 | 1.63E-02 |
| positive regulation of developmental process (GO:0051094) | 1410 | 40 | 19 | + | 2.11 | 1.23E-05 | 4.35E-03 |
| positive regulation of cell differentiation (GO:0045597) | 1010 | 28 | 13.61 | + | 2.06 | 3.98E-04 | 4.69E-02 |
| response to biotic stimulus (GO:0009607) | 1457 | 40 | 19.64 | + | 2.04 | 2.93E-05 | 8.32E-03 |
| response to other organism (GO:0051707) | 1423 | 39 | 19.18 | + | 2.03 | 4.09E-05 | 1.08E-02 |
| positive regulation of phosphorus metabolic process (GO:0010562) | 1169 | 32 | 15.75 | + | 2.03 | 2.11E-04 | 3.05E-02 |
| positive regulation of phosphate metabolic process (GO:0045937) | 1169 | 32 | 15.75 | + | 2.03 | 2.11E-04 | 3.02E-02 |
| response to external biotic stimulus (GO:0043207) | 1425 | 39 | 19.2 | + | 2.03 | 4.13E-05 | 1.08E-02 |
| response to external stimulus (GO:0009605) | 2540 | 68 | 34.23 | + | 1.99 | 3.96E-08 | 7.00E-05 |
| regulation of transport (GO:0051049) | 1839 | 49 | 24.78 | + | 1.98 | 6.73E-06 | 3.15E-03 |
| regulation of localization (GO:0032879) | 2794 | 74 | 37.65 | + | 1.97 | 1.09E-08 | 2.89E-05 |
| regulation of cell population proliferation (GO:0042127) | 1670 | 44 | 22.51 | + | 1.96 | 2.26E-05 | 7.04E-03 |
| regulation of protein phosphorylation (GO:0001932) | 1467 | 38 | 19.77 | + | 1.92 | 1.47E-04 | 2.38E-02 |
| G protein-coupled receptor signaling pathway (GO:0007186) | 1324 | 34 | 17.84 | + | 1.91 | 3.40E-04 | 4.23E-02 |
| response to chemical (GO:0042221) | 4414 | 112 | 59.49 | + | 1.88 | 2.16E-12 | 1.72E-08 |
| positive regulation of multicellular organismal process (GO:0051240) | 1787 | 45 | 24.08 | + | 1.87 | 6.18E-05 | 1.40E-02 |
| defense response (GO:0006952) | 1430 | 36 | 19.27 | + | 1.87 | 3.40E-04 | 4.26E-02 |
| cellular response to chemical stimulus (GO:0070887) | 2927 | 72 | 39.45 | + | 1.83 | 4.09E-07 | 4.07E-04 |
| anatomical structure morphogenesis (GO:0009653) | 2199 | 54 | 29.63 | + | 1.82 | 2.23E-05 | 7.10E-03 |
| regulation of phosphorylation (GO:0042325) | 1639 | 40 | 22.09 | + | 1.81 | 3.22E-04 | 4.23E-02 |
| cellular response to organic substance (GO:0071310) | 2358 | 56 | 31.78 | + | 1.76 | 2.81E-05 | 8.11E-03 |
| regulation of biological quality (GO:0065008) | 4103 | 97 | 55.29 | + | 1.75 | 6.96E-09 | 2.21E-05 |
| regulation of signaling (GO:0023051) | 3638 | 82 | 49.03 | + | 1.67 | 1.73E-06 | 1.25E-03 |
| regulation of cell communication (GO:0010646) | 3599 | 81 | 48.5 | + | 1.67 | 2.33E-06 | 1.61E-03 |
| response to organic substance (GO:0010033) | 3025 | 68 | 40.77 | + | 1.67 | 1.87E-05 | 6.45E-03 |
| regulation of developmental process (GO:0050793) | 2668 | 59 | 35.96 | + | 1.64 | 1.47E-04 | 2.37E-02 |
| multicellular organismal process (GO:0032501) | 7139 | 155 | 96.21 | + | 1.61 | 1.13E-12 | 1.80E-08 |
| signaling (GO:0023052) | 5516 | 118 | 74.34 | + | 1.59 | 2.67E-08 | 6.07E-05 |
| multicellular organism development (GO:0007275) | 5115 | 109 | 68.93 | + | 1.58 | 1.96E-07 | 2.59E-04 |
| regulation of multicellular organismal process (GO:0051239) | 3243 | 69 | 43.7 | + | 1.58 | 9.46E-05 | 1.83E-02 |
| cell communication (GO:0007154) | 5608 | 118 | 75.58 | + | 1.56 | 6.97E-08 | 1.11E-04 |
| regulation of signal transduction (GO:0009966) | 3164 | 66 | 42.64 | + | 1.55 | 3.02E-04 | 4.00E-02 |
| system development (GO:0048731) | 4516 | 94 | 60.86 | + | 1.54 | 5.72E-06 | 2.84E-03 |
| anatomical structure development (GO:0048856) | 5497 | 114 | 74.08 | + | 1.54 | 2.76E-07 | 3.13E-04 |
| animal organ development (GO:0048513) | 3266 | 67 | 44.01 | + | 1.52 | 3.73E-04 | 4.52E-02 |
| developmental process (GO:0032502) | 5952 | 121 | 80.21 | + | 1.51 | 2.63E-07 | 3.22E-04 |
| signal transduction (GO:0007165) | 5154 | 103 | 69.46 | + | 1.48 | 1.06E-05 | 4.11E-03 |
| response to stimulus (GO:0050896) | 8531 | 167 | 114.97 | + | 1.45 | 6.87E-10 | 2.73E-06 |
| cellular response to stimulus (GO:0051716) | 6799 | 133 | 91.63 | + | 1.45 | 3.68E-07 | 3.90E-04 |
| regulation of biological process (GO:0050789) | 11953 | 205 | 161.09 | + | 1.27 | 9.89E-08 | 1.43E-04 |
| regulation of cellular process (GO:0050794) | 11396 | 195 | 153.58 | + | 1.27 | 6.36E-07 | 5.95E-04 |
| biological regulation (GO:0065007) | 12625 | 210 | 170.14 | + | 1.23 | 7.49E-07 | 6.27E-04 |
| cellular process (GO:0009987) | 15625 | 243 | 210.57 | + | 1.15 | 2.86E-06 | 1.75E-03 |
| biological_process (GO:0008150) | 17984 | 266 | 242.36 | + | 1.1 | 9.70E-06 | 3.95E-03 |
| cellular nitrogen compound metabolic process (GO:0034641) | 3400 | 23 | 45.82 | - | 0.5 | 1.14E-04 | 2.03E-02 |
| cellular aromatic compound metabolic process (GO:0006725) | 2984 | 20 | 40.21 | - | 0.5 | 2.80E-04 | 3.84E-02 |
| nucleobase-containing compound metabolic process (GO:0006139) | 2740 | 16 | 36.93 | - | 0.43 | 7.63E-05 | 1.58E-02 |
| Unclassified (UNCLASSIFIED) | 2867 | 15 | 38.64 | - | 0.39 | 9.70E-06 | 3.86E-03 |
| gene expression (GO:0010467) | 2112 | 11 | 28.46 | - | 0.39 | 1.92E-04 | 2.80E-02 |
| nucleic acid metabolic process (GO:0090304) | 2240 | 10 | 30.19 | - | 0.33 | 2.22E-05 | 7.19E-03 |
| RNA metabolic process (GO:0016070) | 1628 | 7 | 21.94 | - | 0.32 | 2.76E-04 | 3.82E-02 |

Table S7 GO Term Analysis of all significant alternative transcription differences between COVID-19 patients who died versus lived

| GO biological process complete | Human (20851) | Significant Genes (281) | Expected count | over/under | Fold Enrichment | P-value | FDR |
| --- | --- | --- | --- | --- | --- | --- | --- |
| RNA polymerase I preinitiation complex assembly (GO:0001188) | 9 | 6 | 0.76 | + | 7.92 | 5.90E-04 | 4.32E-02 |
| regulation of RNA splicing (GO:0043484) | 158 | 34 | 13.31 | + | 2.56 | 7.80E-06 | 9.68E-04 |
| regulation of mRNA splicing, via spliceosome (GO:0048024) | 115 | 23 | 9.68 | + | 2.37 | 5.69E-04 | 4.21E-02 |
| regulation of mRNA processing (GO:0050684) | 154 | 29 | 12.97 | + | 2.24 | 2.32E-04 | 2.01E-02 |
| purine-containing compound biosynthetic process (GO:0072522) | 176 | 32 | 14.82 | + | 2.16 | 2.05E-04 | 1.83E-02 |
| positive regulation of binding (GO:0051099) | 177 | 32 | 14.91 | + | 2.15 | 2.16E-04 | 1.91E-02 |
| positive regulation of protein catabolic process (GO:0045732) | 232 | 41 | 19.54 | + | 2.1 | 5.10E-05 | 5.13E-03 |
| organelle localization by membrane tethering (GO:0140056) | 170 | 30 | 14.32 | + | 2.1 | 4.67E-04 | 3.54E-02 |
| purine nucleotide biosynthetic process (GO:0006164) | 165 | 29 | 13.9 | + | 2.09 | 6.29E-04 | 4.50E-02 |
| chromatin remodeling (GO:0006338) | 181 | 31 | 15.24 | + | 2.03 | 6.62E-04 | 4.70E-02 |
| nucleotide biosynthetic process (GO:0009165) | 233 | 39 | 19.62 | + | 1.99 | 2.16E-04 | 1.91E-02 |
| nucleoside phosphate biosynthetic process (GO:1901293) | 237 | 39 | 19.96 | + | 1.95 | 2.55E-04 | 2.15E-02 |
| regulation of mRNA metabolic process (GO:1903311) | 350 | 57 | 29.48 | + | 1.93 | 1.38E-05 | 1.59E-03 |
| regulation of cysteine-type endopeptidase activity (GO:2000116) | 244 | 39 | 20.55 | + | 1.9 | 4.61E-04 | 3.51E-02 |
| mitochondrion organization (GO:0007005) | 460 | 72 | 38.74 | + | 1.86 | 3.61E-06 | 5.07E-04 |
| organophosphate biosynthetic process (GO:0090407) | 533 | 80 | 44.89 | + | 1.78 | 5.24E-06 | 7.06E-04 |
| regulation of chromosome organization (GO:0033044) | 367 | 55 | 30.91 | + | 1.78 | 1.49E-04 | 1.39E-02 |
| vesicle organization (GO:0016050) | 312 | 46 | 26.28 | + | 1.75 | 6.76E-04 | 4.77E-02 |
| positive regulation of catabolic process (GO:0009896) | 464 | 68 | 39.08 | + | 1.74 | 5.13E-05 | 5.13E-03 |
| nucleotide metabolic process (GO:0009117) | 431 | 63 | 36.3 | + | 1.74 | 1.15E-04 | 1.09E-02 |
| nucleoside phosphate metabolic process (GO:0006753) | 439 | 63 | 36.97 | + | 1.7 | 1.84E-04 | 1.67E-02 |
| MAPK cascade (GO:0000165) | 406 | 57 | 34.19 | + | 1.67 | 5.70E-04 | 4.20E-02 |
| nucleobase-containing compound biosynthetic process (GO:0034654) | 1015 | 141 | 85.48 | + | 1.65 | 6.91E-08 | 1.64E-05 |
| signal transduction by protein phosphorylation (GO:0023014) | 419 | 58 | 35.29 | + | 1.64 | 6.90E-04 | 4.86E-02 |
| lipid biosynthetic process (GO:0008610) | 602 | 83 | 50.7 | + | 1.64 | 6.01E-05 | 5.93E-03 |
| viral process (GO:0016032) | 816 | 112 | 68.72 | + | 1.63 | 3.11E-06 | 4.50E-04 |
| positive regulation of cell migration (GO:0030335) | 526 | 72 | 44.3 | + | 1.63 | 1.89E-04 | 1.71E-02 |
| heterocycle biosynthetic process (GO:0018130) | 1089 | 148 | 91.71 | + | 1.61 | 9.92E-08 | 2.22E-05 |
| nucleobase-containing small molecule metabolic process (GO:0055086) | 524 | 71 | 44.13 | + | 1.61 | 3.28E-04 | 2.59E-02 |
| organelle localization (GO:0051640) | 605 | 81 | 50.95 | + | 1.59 | 1.54E-04 | 1.42E-02 |
| aromatic compound biosynthetic process (GO:0019438) | 1098 | 147 | 92.47 | + | 1.59 | 2.64E-07 | 5.06E-05 |
| regulation of cell cycle process (GO:0010564) | 762 | 102 | 64.17 | + | 1.59 | 2.28E-05 | 2.49E-03 |
| organophosphate metabolic process (GO:0019637) | 894 | 119 | 75.29 | + | 1.58 | 5.94E-06 | 7.87E-04 |
| protein localization to organelle (GO:0033365) | 776 | 103 | 65.35 | + | 1.58 | 2.68E-05 | 2.86E-03 |
| organic cyclic compound biosynthetic process (GO:1901362) | 1236 | 164 | 104.09 | + | 1.58 | 7.31E-08 | 1.71E-05 |
| carbohydrate derivative biosynthetic process (GO:1901137) | 613 | 81 | 51.62 | + | 1.57 | 2.24E-04 | 1.95E-02 |
| regulation of cell cycle (GO:0051726) | 1201 | 158 | 101.14 | + | 1.56 | 2.62E-07 | 5.08E-05 |
| regulation of catabolic process (GO:0009894) | 1043 | 137 | 87.84 | + | 1.56 | 1.83E-06 | 2.85E-04 |
| positive regulation of organelle organization (GO:0010638) | 642 | 84 | 54.07 | + | 1.55 | 2.33E-04 | 2.00E-02 |
| establishment of protein localization (GO:0045184) | 1595 | 208 | 134.33 | + | 1.55 | 3.70E-09 | 1.40E-06 |
| negative regulation of cellular component organization (GO:0051129) | 724 | 94 | 60.97 | + | 1.54 | 1.38E-04 | 1.29E-02 |
| regulation of cellular catabolic process (GO:0031329) | 879 | 114 | 74.03 | + | 1.54 | 2.63E-05 | 2.84E-03 |
| regulation of cell migration (GO:0030334) | 889 | 115 | 74.87 | + | 1.54 | 2.27E-05 | 2.50E-03 |
| positive regulation of hydrolase activity (GO:0051345) | 774 | 100 | 65.18 | + | 1.53 | 1.02E-04 | 9.72E-03 |
| cellular protein localization (GO:0034613) | 1647 | 212 | 138.7 | + | 1.53 | 7.89E-09 | 2.37E-06 |
| cellular macromolecule localization (GO:0070727) | 1656 | 213 | 139.46 | + | 1.53 | 6.63E-09 | 2.11E-06 |
| positive regulation of programmed cell death (GO:0043068) | 661 | 85 | 55.67 | + | 1.53 | 3.67E-04 | 2.83E-02 |
| regulation of mitotic cell cycle (GO:0007346) | 638 | 82 | 53.73 | + | 1.53 | 4.95E-04 | 3.73E-02 |
| regulation of cell motility (GO:2000145) | 952 | 122 | 80.17 | + | 1.52 | 1.88E-05 | 2.12E-03 |
| symbiotic process (GO:0044403) | 908 | 116 | 76.47 | + | 1.52 | 3.51E-05 | 3.65E-03 |
| mRNA metabolic process (GO:0016071) | 698 | 89 | 58.78 | + | 1.51 | 3.15E-04 | 2.52E-02 |
| positive regulation of cell death (GO:0010942) | 715 | 91 | 60.21 | + | 1.51 | 2.85E-04 | 2.31E-02 |
| regulation of locomotion (GO:0040012) | 992 | 126 | 83.54 | + | 1.51 | 2.07E-05 | 2.32E-03 |
| intracellular transport (GO:0046907) | 1529 | 194 | 128.77 | + | 1.51 | 8.46E-08 | 1.95E-05 |
| protein localization (GO:0008104) | 2199 | 279 | 185.19 | + | 1.51 | 5.79E-11 | 3.84E-08 |
| amide transport (GO:0042886) | 1577 | 200 | 132.81 | + | 1.51 | 5.82E-08 | 1.44E-05 |
| positive regulation of apoptotic process (GO:0043065) | 655 | 83 | 55.16 | + | 1.5 | 5.96E-04 | 4.32E-02 |
| protein transport (GO:0015031) | 1510 | 191 | 127.17 | + | 1.5 | 1.52E-07 | 3.17E-05 |
| positive regulation of transcription by RNA polymerase II (GO:0045944) | 1212 | 153 | 102.07 | + | 1.5 | 3.18E-06 | 4.51E-04 |
| regulation of cell adhesion (GO:0030155) | 722 | 91 | 60.8 | + | 1.5 | 3.90E-04 | 2.99E-02 |
| peptide transport (GO:0015833) | 1542 | 193 | 129.86 | + | 1.49 | 2.54E-07 | 4.98E-05 |
| negative regulation of nucleobase-containing compound metabolic process (GO:0045934) | 1559 | 195 | 131.29 | + | 1.49 | 2.27E-07 | 4.56E-05 |
| intracellular protein transport (GO:0006886) | 992 | 124 | 83.54 | + | 1.48 | 4.43E-05 | 4.49E-03 |
| positive regulation of protein metabolic process (GO:0051247) | 1722 | 215 | 145.02 | + | 1.48 | 5.41E-08 | 1.36E-05 |
| phosphorus metabolic process (GO:0006793) | 2192 | 273 | 184.6 | + | 1.48 | 5.61E-10 | 2.79E-07 |
| regulation of cellular component biogenesis (GO:0044087) | 981 | 122 | 82.62 | + | 1.48 | 6.54E-05 | 6.34E-03 |
| regulation of organelle organization (GO:0033043) | 1335 | 166 | 112.43 | + | 1.48 | 2.54E-06 | 3.85E-04 |
| regulation of cellular component movement (GO:0051270) | 1039 | 129 | 87.5 | + | 1.47 | 4.00E-05 | 4.10E-03 |
| negative regulation of RNA metabolic process (GO:0051253) | 1442 | 179 | 121.44 | + | 1.47 | 1.07E-06 | 1.77E-04 |
| positive regulation of protein modification process (GO:0031401) | 1250 | 155 | 105.27 | + | 1.47 | 7.03E-06 | 8.86E-04 |
| nitrogen compound transport (GO:0071705) | 1858 | 230 | 156.47 | + | 1.47 | 2.79E-08 | 7.53E-06 |
| macromolecule localization (GO:0033036) | 2550 | 315 | 214.75 | + | 1.47 | 4.17E-11 | 3.01E-08 |
| cellular response to stress (GO:0033554) | 1749 | 216 | 147.29 | + | 1.47 | 1.08E-07 | 2.32E-05 |
| cellular localization (GO:0051641) | 3014 | 372 | 253.83 | + | 1.47 | 3.62E-13 | 5.23E-10 |
| phosphate-containing compound metabolic process (GO:0006796) | 2165 | 267 | 182.33 | + | 1.46 | 2.56E-09 | 9.92E-07 |
| cellular macromolecule catabolic process (GO:0044265) | 922 | 113 | 77.65 | + | 1.46 | 2.18E-04 | 1.91E-02 |
| regulation of hydrolase activity (GO:0051336) | 1302 | 159 | 109.65 | + | 1.45 | 1.29E-05 | 1.51E-03 |
| apoptotic process (GO:0006915) | 911 | 111 | 76.72 | + | 1.45 | 3.18E-04 | 2.53E-02 |
| protein phosphorylation (GO:0006468) | 971 | 118 | 81.77 | + | 1.44 | 1.98E-04 | 1.78E-02 |
| positive regulation of molecular function (GO:0044093) | 1838 | 223 | 154.79 | + | 1.44 | 2.05E-07 | 4.17E-05 |
| negative regulation of cellular macromolecule biosynthetic process (GO:2000113) | 1495 | 181 | 125.9 | + | 1.44 | 3.98E-06 | 5.55E-04 |
| cellular biosynthetic process (GO:0044249) | 2701 | 327 | 227.47 | + | 1.44 | 1.49E-10 | 8.43E-08 |
| cellular lipid metabolic process (GO:0044255) | 976 | 118 | 82.2 | + | 1.44 | 2.54E-04 | 2.15E-02 |
| positive regulation of cellular component organization (GO:0051130) | 1225 | 148 | 103.17 | + | 1.43 | 3.79E-05 | 3.91E-03 |
| positive regulation of cellular protein metabolic process (GO:0032270) | 1631 | 197 | 137.36 | + | 1.43 | 1.60E-06 | 2.51E-04 |
| organic substance biosynthetic process (GO:1901576) | 2807 | 339 | 236.4 | + | 1.43 | 7.11E-11 | 4.34E-08 |
| negative regulation of macromolecule biosynthetic process (GO:0010558) | 1558 | 188 | 131.21 | + | 1.43 | 3.06E-06 | 4.46E-04 |
| regulation of intracellular signal transduction (GO:1902531) | 1827 | 220 | 153.86 | + | 1.43 | 4.68E-07 | 8.46E-05 |
| establishment of localization in cell (GO:0051649) | 2386 | 287 | 200.94 | + | 1.43 | 5.54E-09 | 1.96E-06 |
| negative regulation of transcription, DNA-templated (GO:0045892) | 1297 | 156 | 109.23 | + | 1.43 | 3.08E-05 | 3.24E-03 |
| macromolecule catabolic process (GO:0009057) | 1057 | 127 | 89.02 | + | 1.43 | 1.80E-04 | 1.65E-02 |
| negative regulation of nucleic acid-templated transcription (GO:1903507) | 1340 | 161 | 112.85 | + | 1.43 | 2.12E-05 | 2.36E-03 |
| negative regulation of RNA biosynthetic process (GO:1902679) | 1342 | 161 | 113.02 | + | 1.42 | 2.64E-05 | 2.83E-03 |
| cellular nitrogen compound biosynthetic process (GO:0044271) | 1577 | 189 | 132.81 | + | 1.42 | 4.31E-06 | 5.90E-04 |
| lipid metabolic process (GO:0006629) | 1219 | 146 | 102.66 | + | 1.42 | 6.99E-05 | 6.73E-03 |
| nucleobase-containing compound metabolic process (GO:0006139) | 2740 | 328 | 230.75 | + | 1.42 | 4.82E-10 | 2.47E-07 |
| positive regulation of catalytic activity (GO:0043085) | 1479 | 177 | 124.56 | + | 1.42 | 1.08E-05 | 1.28E-03 |
| carbohydrate derivative metabolic process (GO:1901135) | 1015 | 121 | 85.48 | + | 1.42 | 3.32E-04 | 2.62E-02 |
| biosynthetic process (GO:0009058) | 2863 | 341 | 241.11 | + | 1.41 | 3.16E-10 | 1.67E-07 |
| positive regulation of RNA metabolic process (GO:0051254) | 1715 | 204 | 144.43 | + | 1.41 | 2.69E-06 | 3.99E-04 |
| negative regulation of gene expression (GO:0010629) | 2043 | 243 | 172.05 | + | 1.41 | 2.39E-07 | 4.74E-05 |
| cell cycle (GO:0007049) | 1380 | 164 | 116.22 | + | 1.41 | 3.33E-05 | 3.49E-03 |
| phosphorylation (GO:0016310) | 1308 | 155 | 110.16 | + | 1.41 | 6.32E-05 | 6.17E-03 |
| cellular macromolecule biosynthetic process (GO:0034645) | 1647 | 195 | 138.7 | + | 1.41 | 6.51E-06 | 8.34E-04 |
| chromosome organization (GO:0051276) | 1065 | 126 | 89.69 | + | 1.4 | 3.59E-04 | 2.78E-02 |
| positive regulation of nucleic acid-templated transcription (GO:1903508) | 1623 | 192 | 136.68 | + | 1.4 | 8.66E-06 | 1.06E-03 |
| positive regulation of RNA biosynthetic process (GO:1902680) | 1624 | 192 | 136.77 | + | 1.4 | 8.71E-06 | 1.06E-03 |
| positive regulation of nitrogen compound metabolic process (GO:0051173) | 3209 | 379 | 270.25 | + | 1.4 | 5.76E-11 | 3.98E-08 |
| positive regulation of macromolecule biosynthetic process (GO:0010557) | 1882 | 222 | 158.5 | + | 1.4 | 1.56E-06 | 2.49E-04 |
| negative regulation of biosynthetic process (GO:0009890) | 1646 | 194 | 138.62 | + | 1.4 | 8.10E-06 | 9.98E-04 |
| positive regulation of transcription, DNA-templated (GO:0045893) | 1536 | 181 | 129.36 | + | 1.4 | 1.87E-05 | 2.13E-03 |
| macromolecule biosynthetic process (GO:0009059) | 1699 | 200 | 143.08 | + | 1.4 | 7.06E-06 | 8.83E-04 |
| positive regulation of signaling (GO:0023056) | 1869 | 220 | 157.4 | + | 1.4 | 2.21E-06 | 3.38E-04 |
| negative regulation of cellular biosynthetic process (GO:0031327) | 1617 | 190 | 136.18 | + | 1.4 | 1.28E-05 | 1.51E-03 |
| positive regulation of phosphorus metabolic process (GO:0010562) | 1169 | 137 | 98.45 | + | 1.39 | 2.83E-04 | 2.32E-02 |
| positive regulation of phosphate metabolic process (GO:0045937) | 1169 | 137 | 98.45 | + | 1.39 | 2.83E-04 | 2.31E-02 |
| regulation of catalytic activity (GO:0050790) | 2395 | 280 | 201.7 | + | 1.39 | 1.02E-07 | 2.25E-05 |
| positive regulation of macromolecule metabolic process (GO:0010604) | 3560 | 416 | 299.81 | + | 1.39 | 1.55E-11 | 1.29E-08 |
| regulation of cellular component organization (GO:0051128) | 2439 | 285 | 205.4 | + | 1.39 | 6.87E-08 | 1.66E-05 |
| positive regulation of phosphorylation (GO:0042327) | 1096 | 128 | 92.3 | + | 1.39 | 5.25E-04 | 3.92E-02 |
| organelle organization (GO:0006996) | 3568 | 416 | 300.48 | + | 1.38 | 2.02E-11 | 1.60E-08 |
| positive regulation of signal transduction (GO:0009967) | 1667 | 194 | 140.39 | + | 1.38 | 1.68E-05 | 1.92E-03 |
| organic substance transport (GO:0071702) | 2219 | 258 | 186.88 | + | 1.38 | 5.07E-07 | 9.06E-05 |
| heterocycle metabolic process (GO:0046483) | 2936 | 341 | 247.26 | + | 1.38 | 4.24E-09 | 1.57E-06 |
| positive regulation of nucleobase-containing compound metabolic process (GO:0045935) | 1886 | 219 | 158.83 | + | 1.38 | 5.52E-06 | 7.37E-04 |
| positive regulation of cellular biosynthetic process (GO:0031328) | 1981 | 230 | 166.83 | + | 1.38 | 2.67E-06 | 4.01E-04 |
| RNA metabolic process (GO:0016070) | 1628 | 189 | 137.1 | + | 1.38 | 2.52E-05 | 2.74E-03 |
| nucleic acid metabolic process (GO:0090304) | 2240 | 260 | 188.65 | + | 1.38 | 5.59E-07 | 9.87E-05 |
| positive regulation of cell communication (GO:0010647) | 1861 | 216 | 156.73 | + | 1.38 | 6.00E-06 | 7.88E-04 |
| negative regulation of nitrogen compound metabolic process (GO:0051172) | 2486 | 288 | 209.36 | + | 1.38 | 1.31E-07 | 2.77E-05 |
| positive regulation of biosynthetic process (GO:0009891) | 2014 | 233 | 169.61 | + | 1.37 | 3.12E-06 | 4.47E-04 |
| regulation of protein modification process (GO:0031399) | 1885 | 218 | 158.75 | + | 1.37 | 6.76E-06 | 8.60E-04 |
| positive regulation of metabolic process (GO:0009893) | 3863 | 446 | 325.33 | + | 1.37 | 1.12E-11 | 9.91E-09 |
| cellular catabolic process (GO:0044248) | 1823 | 210 | 153.53 | + | 1.37 | 1.34E-05 | 1.56E-03 |
| positive regulation of cellular metabolic process (GO:0031325) | 3392 | 390 | 285.66 | + | 1.37 | 6.60E-10 | 3.18E-07 |
| cellular aromatic compound metabolic process (GO:0006725) | 2984 | 342 | 251.3 | + | 1.36 | 1.50E-08 | 4.27E-06 |
| cellular protein modification process (GO:0006464) | 3195 | 365 | 269.07 | + | 1.36 | 6.10E-09 | 2.06E-06 |
| protein modification process (GO:0036211) | 3195 | 365 | 269.07 | + | 1.36 | 6.10E-09 | 2.02E-06 |
| cellular macromolecule metabolic process (GO:0044260) | 5152 | 588 | 433.88 | + | 1.36 | 2.75E-15 | 4.38E-12 |
| regulation of molecular function (GO:0065009) | 3082 | 351 | 259.56 | + | 1.35 | 1.88E-08 | 5.15E-06 |
| organic cyclic compound metabolic process (GO:1901360) | 3216 | 366 | 270.84 | + | 1.35 | 8.19E-09 | 2.41E-06 |
| negative regulation of signal transduction (GO:0009968) | 1293 | 147 | 108.89 | + | 1.35 | 5.32E-04 | 3.95E-02 |
| negative regulation of cellular metabolic process (GO:0031324) | 2685 | 305 | 226.12 | + | 1.35 | 2.66E-07 | 5.03E-05 |
| intracellular signal transduction (GO:0035556) | 1689 | 191 | 142.24 | + | 1.34 | 8.87E-05 | 8.49E-03 |
| negative regulation of macromolecule metabolic process (GO:0010605) | 2928 | 331 | 246.59 | + | 1.34 | 1.05E-07 | 2.29E-05 |
| cellular nitrogen compound metabolic process (GO:0034641) | 3400 | 384 | 286.34 | + | 1.34 | 6.47E-09 | 2.10E-06 |
| macromolecule modification (GO:0043412) | 3420 | 386 | 288.02 | + | 1.34 | 6.97E-09 | 2.13E-06 |
| regulation of apoptotic process (GO:0042981) | 1561 | 176 | 131.46 | + | 1.34 | 2.22E-04 | 1.94E-02 |
| organonitrogen compound biosynthetic process (GO:1901566) | 1405 | 158 | 118.32 | + | 1.34 | 5.91E-04 | 4.31E-02 |
| negative regulation of signaling (GO:0023057) | 1414 | 159 | 119.08 | + | 1.34 | 5.14E-04 | 3.86E-02 |
| regulation of programmed cell death (GO:0043067) | 1583 | 178 | 133.31 | + | 1.34 | 2.42E-04 | 2.06E-02 |
| positive regulation of gene expression (GO:0010628) | 2305 | 259 | 194.12 | + | 1.33 | 6.31E-06 | 8.23E-04 |
| cellular protein metabolic process (GO:0044267) | 3829 | 430 | 322.47 | + | 1.33 | 9.70E-10 | 4.41E-07 |
| regulation of RNA metabolic process (GO:0051252) | 3821 | 429 | 321.79 | + | 1.33 | 1.14E-09 | 5.03E-07 |
| negative regulation of cell communication (GO:0010648) | 1411 | 158 | 118.83 | + | 1.33 | 6.12E-04 | 4.42E-02 |
| small molecule metabolic process (GO:0044281) | 1744 | 195 | 146.87 | + | 1.33 | 1.33E-04 | 1.25E-02 |
| macromolecule metabolic process (GO:0043170) | 6340 | 707 | 533.93 | + | 1.32 | 4.71E-17 | 9.36E-14 |
| cellular metabolic process (GO:0044237) | 7790 | 867 | 656.05 | + | 1.32 | 1.09E-22 | 8.63E-19 |
| regulation of nucleobase-containing compound metabolic process (GO:0019219) | 4090 | 455 | 344.45 | + | 1.32 | 7.75E-10 | 3.62E-07 |
| cellular component assembly (GO:0022607) | 2393 | 266 | 201.53 | + | 1.32 | 8.89E-06 | 1.07E-03 |
| organic substance catabolic process (GO:1901575) | 1782 | 198 | 150.07 | + | 1.32 | 1.82E-04 | 1.66E-02 |
| nitrogen compound metabolic process (GO:0006807) | 7092 | 788 | 597.26 | + | 1.32 | 1.79E-19 | 5.70E-16 |
| catabolic process (GO:0009056) | 2099 | 233 | 176.77 | + | 1.32 | 4.27E-05 | 4.35E-03 |
| regulation of cell death (GO:0010941) | 1714 | 190 | 144.35 | + | 1.32 | 2.83E-04 | 2.33E-02 |
| regulation of protein metabolic process (GO:0051246) | 2842 | 315 | 239.34 | + | 1.32 | 1.25E-06 | 2.03E-04 |
| primary metabolic process (GO:0044238) | 7575 | 839 | 637.94 | + | 1.32 | 5.94E-21 | 2.36E-17 |
| negative regulation of metabolic process (GO:0009892) | 3178 | 351 | 267.64 | + | 1.31 | 3.23E-07 | 6.04E-05 |
| gene expression (GO:0010467) | 2112 | 233 | 177.87 | + | 1.31 | 6.32E-05 | 6.20E-03 |
| cellular component biogenesis (GO:0044085) | 2654 | 292 | 223.51 | + | 1.31 | 6.46E-06 | 8.34E-04 |
| regulation of phosphate metabolic process (GO:0019220) | 1838 | 202 | 154.79 | + | 1.3 | 2.63E-04 | 2.20E-02 |
| regulation of phosphorus metabolic process (GO:0051174) | 1839 | 202 | 154.87 | + | 1.3 | 2.65E-04 | 2.20E-02 |
| negative regulation of response to stimulus (GO:0048585) | 1840 | 202 | 154.96 | + | 1.3 | 2.66E-04 | 2.20E-02 |
| organic substance metabolic process (GO:0071704) | 8009 | 879 | 674.49 | + | 1.3 | 2.73E-21 | 1.45E-17 |
| regulation of macromolecule biosynthetic process (GO:0010556) | 4064 | 446 | 342.26 | + | 1.3 | 6.90E-09 | 2.15E-06 |
| organonitrogen compound metabolic process (GO:1901564) | 5446 | 596 | 458.64 | + | 1.3 | 3.63E-12 | 4.12E-09 |
| metabolic process (GO:0008152) | 8590 | 939 | 723.42 | + | 1.3 | 3.17E-23 | 5.03E-19 |
| regulation of signal transduction (GO:0009966) | 3164 | 345 | 266.46 | + | 1.29 | 1.49E-06 | 2.40E-04 |
| protein metabolic process (GO:0019538) | 4416 | 480 | 371.9 | + | 1.29 | 4.56E-09 | 1.65E-06 |
| regulation of cellular macromolecule biosynthetic process (GO:2000112) | 3939 | 428 | 331.73 | + | 1.29 | 5.85E-08 | 1.43E-05 |
| positive regulation of cellular process (GO:0048522) | 5680 | 617 | 478.35 | + | 1.29 | 3.83E-12 | 4.06E-09 |
| negative regulation of cellular process (GO:0048523) | 4955 | 538 | 417.29 | + | 1.29 | 3.00E-10 | 1.64E-07 |
| regulation of cellular protein metabolic process (GO:0032268) | 2685 | 290 | 226.12 | + | 1.28 | 2.71E-05 | 2.87E-03 |
| regulation of nucleic acid-templated transcription (GO:1903506) | 3549 | 383 | 298.88 | + | 1.28 | 7.18E-07 | 1.25E-04 |
| regulation of gene expression (GO:0010468) | 4907 | 529 | 413.25 | + | 1.28 | 1.37E-09 | 5.89E-07 |
| regulation of RNA biosynthetic process (GO:2001141) | 3554 | 383 | 299.31 | + | 1.28 | 8.57E-07 | 1.45E-04 |
| regulation of biosynthetic process (GO:0009889) | 4297 | 463 | 361.88 | + | 1.28 | 3.28E-08 | 8.68E-06 |
| regulation of transcription by RNA polymerase II (GO:0006357) | 2593 | 279 | 218.37 | + | 1.28 | 5.45E-05 | 5.41E-03 |
| regulation of cell communication (GO:0010646) | 3599 | 387 | 303.1 | + | 1.28 | 9.65E-07 | 1.61E-04 |
| regulation of signaling (GO:0023051) | 3638 | 391 | 306.38 | + | 1.28 | 7.75E-07 | 1.33E-04 |
| regulation of cellular metabolic process (GO:0031323) | 6347 | 681 | 534.52 | + | 1.27 | 1.02E-12 | 1.24E-09 |
| regulation of primary metabolic process (GO:0080090) | 6135 | 658 | 516.67 | + | 1.27 | 4.33E-12 | 4.31E-09 |
| regulation of cellular biosynthetic process (GO:0031326) | 4216 | 452 | 355.06 | + | 1.27 | 8.81E-08 | 2.00E-05 |
| negative regulation of biological process (GO:0048519) | 5618 | 601 | 473.13 | + | 1.27 | 1.43E-10 | 8.44E-08 |
| regulation of nitrogen compound metabolic process (GO:0051171) | 5937 | 632 | 499.99 | + | 1.26 | 6.75E-11 | 4.29E-08 |
| regulation of transcription, DNA-templated (GO:0006355) | 3478 | 370 | 292.91 | + | 1.26 | 4.90E-06 | 6.65E-04 |
| positive regulation of biological process (GO:0048518) | 6272 | 665 | 528.21 | + | 1.26 | 2.59E-11 | 1.96E-08 |
| regulation of macromolecule metabolic process (GO:0060255) | 6508 | 689 | 548.08 | + | 1.26 | 9.43E-12 | 8.82E-09 |
| regulation of metabolic process (GO:0019222) | 7059 | 746 | 594.48 | + | 1.25 | 5.97E-13 | 7.91E-10 |
| establishment of localization (GO:0051234) | 4694 | 490 | 395.31 | + | 1.24 | 4.20E-07 | 7.67E-05 |
| cellular component organization (GO:0016043) | 5695 | 594 | 479.61 | + | 1.24 | 9.06E-09 | 2.62E-06 |
| regulation of localization (GO:0032879) | 2794 | 291 | 235.3 | + | 1.24 | 2.91E-04 | 2.35E-02 |
| cellular component organization or biogenesis (GO:0071840) | 5915 | 612 | 498.14 | + | 1.23 | 1.55E-08 | 4.32E-06 |
| regulation of response to stimulus (GO:0048583) | 4391 | 454 | 369.8 | + | 1.23 | 4.08E-06 | 5.64E-04 |
| localization (GO:0051179) | 5850 | 603 | 492.67 | + | 1.22 | 3.61E-08 | 9.41E-06 |
| transport (GO:0006810) | 4562 | 466 | 384.2 | + | 1.21 | 9.61E-06 | 1.15E-03 |
| response to stress (GO:0006950) | 3644 | 368 | 306.89 | + | 1.2 | 3.43E-04 | 2.67E-02 |
| regulation of biological quality (GO:0065008) | 4103 | 406 | 345.54 | + | 1.17 | 6.23E-04 | 4.48E-02 |
| cellular process (GO:0009987) | 15625 | 1467 | 1315.88 | + | 1.11 | 6.12E-17 | 1.08E-13 |
| regulation of cellular process (GO:0050794) | 11396 | 1067 | 959.73 | + | 1.11 | 7.41E-07 | 1.28E-04 |
| regulation of biological process (GO:0050789) | 11953 | 1109 | 1006.64 | + | 1.1 | 1.95E-06 | 3.01E-04 |
| biological regulation (GO:0065007) | 12625 | 1171 | 1063.23 | + | 1.1 | 3.41E-07 | 6.29E-05 |
| biological_process (GO:0008150) | 17984 | 1632 | 1514.55 | + | 1.08 | 1.56E-17 | 4.13E-14 |
| sensory perception (GO:0007600) | 984 | 52 | 82.87 | - | 0.63 | 4.40E-04 | 3.36E-02 |
| G protein-coupled receptor signaling pathway (GO:0007186) | 1324 | 65 | 111.5 | - | 0.58 | 2.95E-06 | 4.34E-04 |
| Unclassified (UNCLASSIFIED) | 2867 | 124 | 241.45 | - | 0.51 | 1.56E-17 | 3.54E-14 |
| detection of stimulus (GO:0051606) | 709 | 25 | 59.71 | - | 0.42 | 1.08E-06 | 1.77E-04 |
| humoral immune response (GO:0006959) | 386 | 13 | 32.51 | - | 0.4 | 2.96E-04 | 2.37E-02 |
| detection of chemical stimulus (GO:0009593) | 521 | 13 | 43.88 | - | 0.3 | 1.76E-07 | 3.64E-05 |
| sensory perception of chemical stimulus (GO:0007606) | 540 | 13 | 45.48 | - | 0.29 | 4.87E-08 | 1.25E-05 |
| detection of stimulus involved in sensory perception (GO:0050906) | 549 | 11 | 46.23 | - | 0.24 | 1.97E-09 | 8.03E-07 |
| sensory perception of smell (GO:0007608) | 468 | 8 | 39.41 | - | 0.2 | 5.98E-09 | 2.07E-06 |
| complement activation (GO:0006956) | 177 | 3 | 14.91 | - | 0.2 | 6.45E-04 | 4.60E-02 |
| detection of chemical stimulus involved in sensory perception (GO:0050907) | 484 | 8 | 40.76 | - | 0.2 | 2.12E-09 | 8.44E-07 |
| detection of chemical stimulus involved in sensory perception of smell (GO:0050911) | 439 | 6 | 36.97 | - | 0.16 | 1.65E-09 | 6.89E-07 |
| complement activation, classical pathway (GO:0006958) | 161 | 2 | 13.56 | - | 0.15 | 3.41E-04 | 2.67E-02 |
| humoral immune response mediated by circulating immunoglobulin (GO:0002455) | 167 | 2 | 14.06 | - | 0.14 | 2.38E-04 | 2.04E-02 |
